# Benchmarking AWaRe: estimating optimal levels of AWaRe antibiotic use in 186 countries, territories and areas based on clinical infection and resistance burden

**DOI:** 10.64898/2026.01.26.26344900

**Authors:** Aislinn Cook, Ben Cooper, Mike Thorn, Nam Nguyen, Cherry Lim, Myo Maung Maung Swe, Kasim Allel, Gisela Robles Aguilar, Catrin E. Moore, Joseph A. Lewnard, Jennifer Cohn, Marc Mendelson, Ramanan Laxminarayan, Padmini Srikantiah, Koen B. Pouwels, Michael Sharland

## Abstract

**Background:** Ensuring appropriate access to essential antibiotics is a critical global public health goal. The UN General Assembly agreed that 70% of global antibiotic use should be WHO Access group. A standard method to estimate optimal antibiotic use based on burden of disease, resistance and local context is needed to inform national policies.

**Methods:** Using data from multiple global datasets, we clustered 186 countries, territories and areas (CTAs) into ‘peer groups’ based on sociodemographic factors, infection and resistance incidence using a latent class model. Within each cluster, benchmark countries with low antibiotic use and infection mortality were identified. We estimated optimal Total DID given infection burden, Reserve DID based on relevant resistance burdens, Watch DID based on clinical infections requiring Watch antibiotics from the WHO AWaRe Book and Access DID as the residual volume.

**Findings:** Globally 43.0 billion DDDs (95%CI:35.4billion–57.7billion) of antibiotics in 2019 were needed in 186 CTAs, of which 76% would optimally be Access (95%CI:70%-82%). CTAs in lower-income clusters required more Watch and Reserve antibiotics than higher income CTAs. Among CTAs with actual use data available, 72% (48/67) of CTAs used more Total and 99% (66/67) used more Watch antibiotics than estimated optimal levels.

**Implications:** We present the first estimates of optimal AWaRe antibiotic levels for 186 CTAs. After accounting for country-specific needs, the UNGA 70% Access target is globally appropriate. AWaRe benchmarking enables CTAs to estimate under and overuse of AWaRe antibiotics to inform national policies.

**Funding:** The ADILA Project funded by the Wellcome Trust [222051/Z/20/Z].

**Research in context:** *Evidence before this study:* Member states agreed at the 2024 United Nations General Assembly (UNGA) AMR meeting that the WHO AWaRe (Access, Watch, Reserve) system should underpin global antibiotic surveillance and that 70% of global use should be from the AWaRe Access group, while accounting for national contexts but did not agree any method for derived country-level targets. The WHO GLASS antimicrobial use (GLASS-AMU) surveillance system and other studies have reported observed antibiotic use. GLASS reports actual national medicine-level antibiotic use from 2015-2022 for 60 CTAs. The GRAM study (Browne, et al., 2021) modelled estimated total antibiotic use from 2000-2018 for 204 CTAs but were only able to estimate AWaRe antibiotic use for 76 CTAs where data from the commercial IQVIA MIDAS® database were available. Recent estimates from Klein et al. (2024) reported changes in antibiotic use from 2016 – 2023 for 67 CTAs in IQVIA MIDAS®. Despite these studies quantifying estimates of current national medicine-level antibiotic use, there are very few estimates of what levels of antibiotic use are “optimal” both overall and by AWaRe group. We searched PubMed for studies from January 2015 through October 2025 on national-level antibiotic targets and AWaRe antibiotic policy using Boolean search terms. We identified few studies that estimated optimal levels of antibiotic use, but none for all AWaRe groups. As part of the Lancet AMR series, Mendelson et al. (2024) estimated expected total antibiotic use based on infection burden using the WHO AWaRe book guidance. This study developed a framework for estimating required Watch antibiotic use but did not provide comprehensive estimates for Access and Reserve use. Summan et al. (2025) estimated antibiotic needs for chronic obstructive pulmonary disease (COPD) and pneumonia in 20 CTAs using disease burden and bacterial aetiology. These estimates focused only on penicillins and cephalosporins and did not provide population based standardised measures to allow for cross-national comparison among CTAs. Mishra et al. (2025) estimated the gap in Reserve antibiotic treatment courses for use in carbapenem resistant Gram-negative infections in 8 countries using GRAM estimates and IQVIA sales data. While these studies have estimated expected antibiotic use for specific antibiotics or infections, there is no reported method to estimate optimal national levels of total and AWaRe antibiotic use.

*Added value of this study:* We developed a standard method for deriving optimal ranges of total and AWaRe antibiotic use in defined daily doses (DDD) per 1000 inhabitants per day (DID) accounting for national level infection and antibiotic resistance burden, population socio-demographics, national income, health system infrastructure and healthcare access using a benchmarking approach. Our study provides the first comprehensive estimates of optimal levels of total antibiotic use and disaggregated by AWaRe group in DID for a 2019 baseline for 186 CTAs and compares actual levels of AWaRe use to expected optimal use for 67 CTAs using the IQVIA MIDAS® dataset. We used publicly available data from the Global Burden of Disease (GBD), Global Research on Antimicrobial Resistance (GRAM) and the World Bank to provide an open-access, standardised AWaRe-clinical framework that could inform national and global evidence-based antibiotic policy development and implementation.

*Implications of all the available evidence:* Although some recent studies have attempted to derive estimates for optimal levels of antibiotic use, they have been focused on specific infections or antibiotics. The UNGA AMR commitments provide a clear direction for global target setting for antibiotic use. Our study provides the first method to estimate optimal antibiotic levels at a country level for both total and AWaRe, based on local infection burden and antibiotic resistance for 186 CTAs. Estimating optimal antibiotic levels with an agreed standard methodology across CTAs would allow for benchmarking and peer-comparison, assisting with target setting and shared learning across National Action Plans from different policy initiatives and outcomes.

## Introduction

Ensuring equitable, appropriate access to effective essential antibiotics is central to Sustainable Development Goal (SDG) 3, and universal health coverage (UHC)^(1)^. In 2017, the World Health Organization (WHO) introduced the AWaRe (Access, Watch, Reserve) system, which groups more than 250 antibiotics based on efficacy, safety, and selection for resistance.^(2,3)^ The 2023 WHO Model list of Essential Medicines List (EML) only includes 41 antibiotics (Box 1). The 2022 WHO AWaRe Antibiotic book provides treatment guidance for the 35 most common infections in primary care and hospital settings, including recommendation on the choice of EML antibiotic (if indicated), dose, and duration.^(4,5)^

Rising global human antibiotic use is now driven largely by increased use in middle-income countries, particularly of Watch antibiotics.^(6)^ High-income countries generally still have higher antibiotic use rates per capita than lower income groups countries,^(7,8)^ and access to essential antibiotics remains limited in many low-income countries across all AWaRe groups, from oral amoxicillin (Access) to new Reserve antibiotics.^(9,10)^

The 2024 United Nations General Assembly (UNGA) high-level meeting on antimicrobial resistance (AMR), endorsed two key commitments on antibiotic use (ABU): 1) the WHO AWaRe system should underpin global surveillance and 2) by 2030, at least 70% of global human antibiotic use should be from the Access group.^(11)^ While this target provides an important global benchmark, it does not account for large differences in infection burden, antibiotic resistance (ABR), population demographics, and healthcare provision.^(12-14)^

Countries with very different absolute levels of ABU can meet the 70% Access target, obscuring both excessive use and critical gaps in access (Box 2). To address these limitations, complementary metrics are required that estimate optimal antibiotic volumes, expressed as defined daily doses (DDD) per 1000 inhabitants per day (DID), for both total use and across AWaRe groups, while accounting for local epidemiological contexts and prevalence of ABR.

Here, we develop and apply a burden-adjusted framework for estimating expected optimal national levels of AWaRe ABU, based on infection burden, ABR, and sociodemographic characteristics. We provide estimates of the expected volumes of total, Access, Watch, and Reserve antibiotic use for 186 CTAs, that could be compared to levels of actual use, to help inform national antibiotic policies and identify access gaps.

### Box 1.

*Box describing the AWaRe classification and an overview of the split of AWaRe antibiotics (n=257) by Model Essential Medicines List (EML) and non-EML status (EML TA = therapeutic alternative to an EML antibiotic)*.

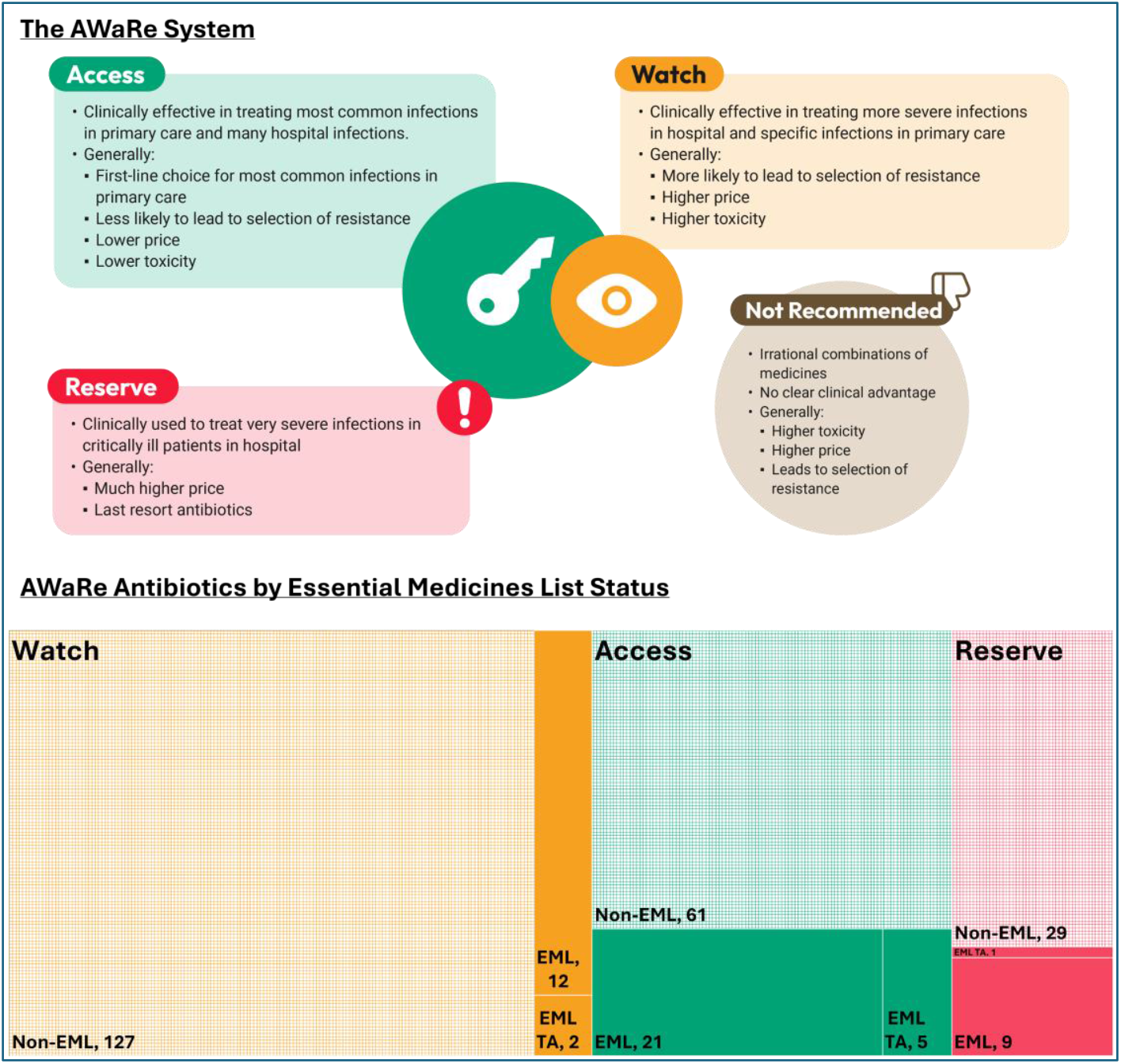

### Box 2.

*An example illustration of how countries can be meeting the 70% Access target with different absolute total antibiotic volumes*.

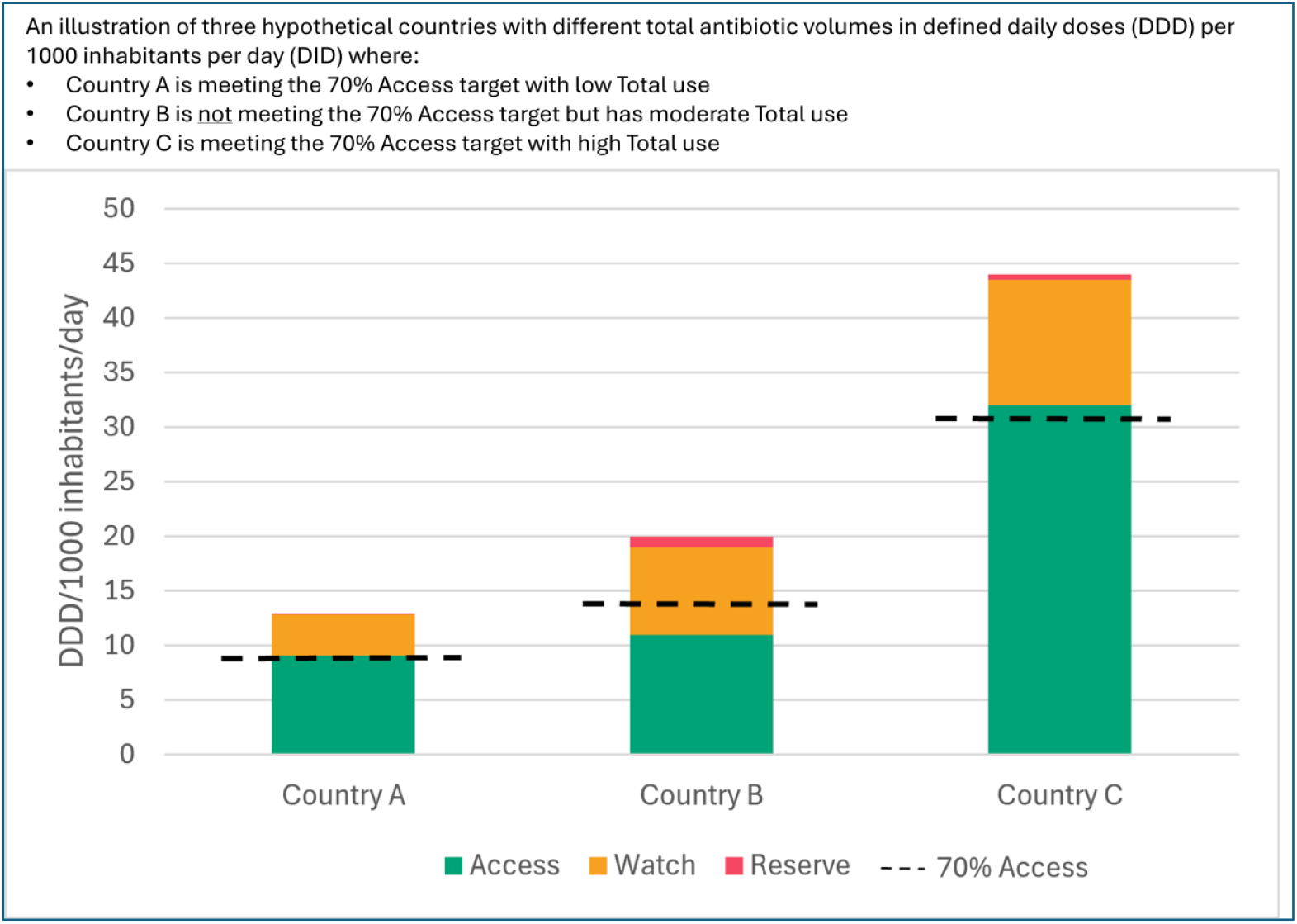

## Methods

### OVERVIEW

We estimated country-level optimal ranges of ABU volumes (DID) for total use and by AWaRe group for 2019 for 186 CTAs based on country-specific infection burden, ABR (Figure 1), and sociodemographic factors such as age groups, national income and health system factors that may influence infection risk, poor infection outcomes, and healthcare access.

**Figure 1.**
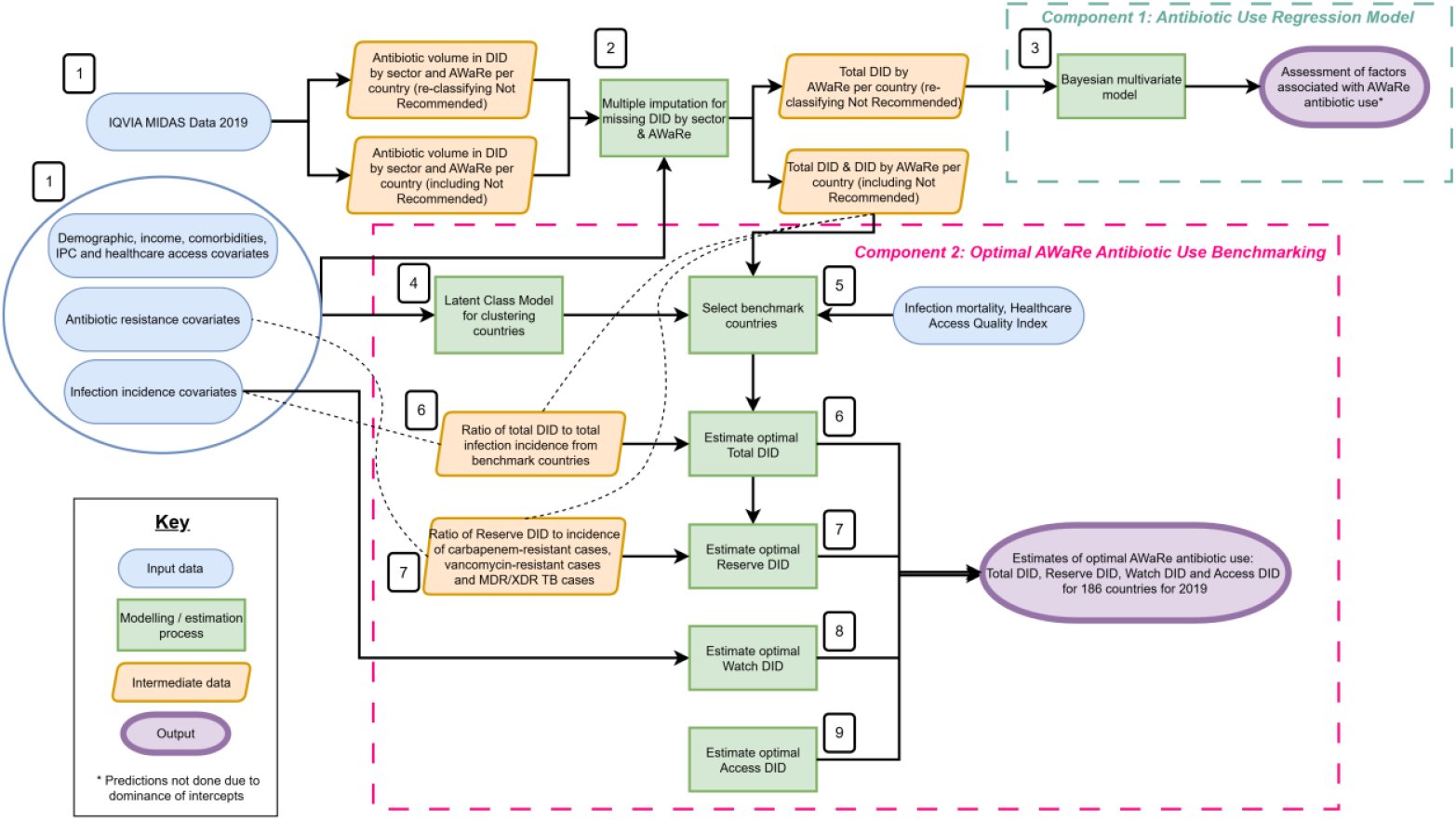
Study schema for deriving estimates of expected AWaRe antibiotic use based on infection burden, antibiotic resistance, socio-demographics, healthcare infrastructure and access using the WHO AWaRe book as a clinical framework. Figure shows the two main components of analysis: Component 1: Antibiotic Use Regression Model, Component 2: Optimal AWaRe Antibiotic Use Benchmarking. The numbers correspond to the step they are next to: 1. Data preparation, 2. Missing data imputation, 3. Antibiotic use regression model, 4. Latent Class Model, 5. Benchmark country selection, 6. Estimate optimal total DID, 7. Estimate optimal Reserve DID, 8. Estimate optimal Watch DID, 9. Estimate optimal Access DID

We used two complementary analytic approaches, described briefly below and in full in the supplementary methods. First, we applied a multivariate ecological regression model to estimate expected levels of AWaRe antibiotic use given country-level infection, ABR burden and population risk-factors. The fitted models were then used to generate predicted (expected) AWaRe-specific antibiotic use volumes for each CTA.

Second, we developed a benchmarking approach to estimate expected levels of AWaRe ABU based on peer-country comparisons. CTAs were clustered into groups with similar characteristics using a Latent Class Model (LCM). Within each cluster, benchmark CTAs were selected based on total DID, infection mortality and percentage of Access volume use. Optimal total antibiotic levels were estimated within each cluster based on infection burden, after which Reserve need was derived from relevant ABR burdens, Watch levels from the burden of infections requiring Watch antibiotics, and Access was estimated as the residual volume after accounting for Watch and Reserve needs (Figure 1).

### DATA SOURCES

We combined data from multiple sources with full details in the supplemental methods. We used data from 2019, the baseline year specified in the UNGA declaration, and to avoid disruption from the Covid-19 pandemic.

Country-level antibiotic sales data were obtained from the IQVIA MIDAS® Quarterly Sales Database^(15)^ for 72 CTAs, covering hospital and retail sectors. Sales volumes were converted from kilograms to DDDs using the 2025 WHO/ATC method^(16)^ and expressed as DID using World Bank population estimates or alternate sources.^(17,18)^

Infection incidence and mortality data for conditions where antibiotics are recommended in the WHO AWaRe book were taken from the Global Burden of Disease (GBD) 2021 study^(19,20)^ where available: cellulitis, upper respiratory infections, otitis media, lower respiratory infections (excluding hospital-acquired pneumonia ‘HAP’), multi-drug-resistant (MDR) and extensively drug-resistant (XDR) tuberculosis (TB), typhoid and paratyphoid, diarrhoea, urinary tract infections, chlamydia, syphilis, gonorrhoea, trichomoniasis, and sepsis. We approximated incidence of cholera, bloody diarrhoea (Watch antibiotics indicated) and hospital infections not included in GBD (HAP and intra-abdominal infections (IAI)) (supplemental methods).

We used data from the GRAM project^(21)^ to estimate incidence of infections caused by ABR pathogens where resistance may require escalating AWaRe group category (e.g. where carbapenem resistance is common, sepsis treatment may more frequently require a Reserve antibiotic such as colistin). We included other covariates that may be associated with antibiotic use at a population level (Figure 1#1, supplemental methods). Missing sector-level data from MIDAS and missing covariates were imputed using multiple imputation by chained equations (MICE) (Figure 1#2, supplemental methods). After imputation, five countries where total DID was below 9.7 DID, the lowest value observed in a high-income country, were excluded from the MIDAS data leaving 67/72 for further analysis.

### STATISTICAL ANALYSES

#### Antibiotic use regression model

We initially applied a Bayesian multivariate ecological regression model with a gamma distribution and a log-link function to country-level data from 67 CTAs in MIDAS® to quantify how national factors potentially associated with antibiotic need, including infectious disease burden, ABR, and other covariates selected *a priori*, relate to ABU volumes across AWaRe categories (Figure 1:Component 1).

We used normal priors for the intercepts (Access, Watch and Reserve) deriving mean and standard deviation from the GRAM estimates of ABU in 2018.^(7)^ For the covariates, we used horseshoe priors with a parameter ratio of 0.8. Models were fitted separately to each of 1000 imputed datasets, and posterior draws were pooled to propagate uncertainty arising from both missing data and model estimation. We used the fitted model to predict expected national volumes of Access, Watch, and Reserve antibiotic use for 186 CTAs with available covariates.

#### Optimal AWaRe Antibiotic Use Benchmarking

We also developed an alternative benchmarking approach to estimate optimal national antibiotic use volumes by AWaRe group (Figure 1:Component 2).

The 186 CTAs with complete covariates were grouped into ‘peer’ clusters using latent class modelling of 40 sociodemographic and clinical covariates (Figure 1#4, supplemental methods).^(22,23)^ Clusters were ordered by median GNI per capita within a cluster from low to high income CTAs. Within each cluster, a benchmark CTA was identified by minimising total ABU and infection mortality, while maximising the percentage of Access use (Figure 1#5). In cases where no suitable benchmark CTA was identified within a cluster due to having low ABU but higher infection mortality than the benchmark CTA in the next higher cluster, we used the benchmark CTA from the next higher cluster to represent a minimum optimal range of ABU required for that cluster.

We used the benchmark CTAs to estimate optimal total DID for other CTAs in the same cluster, assuming their infection burden would be treated according to the prescribing practices of the benchmark country. For each benchmark CTA, we calculated the ratio of total antibiotic use to total infection incidence. This ratio was applied to the infection burden of the remaining CTAs within the same cluster to get expected Total DID (Figure 1#6). For our primary analyses, we adjusted total case counts for UTI and cellulitis to account for potential underdiagnosis of infections requiring antibiotics in GBD (supplemental methods).

Expected Reserve DID was estimated using ratios of Reserve antibiotics to resistant infections derived from the benchmark CTA in the highest-income cluster (cluster 4), assuming this HIC has no antibiotic access issues (Figure 1#7). Ratios were calculated separately by pathogen group and multiplied by the corresponding resistant infection burdens in each CTA, then summed to estimate total Reserve requirements.

Expected Watch antibiotic use was estimated from case counts of infections for which Watch antibiotics are recommended in WHO guidelines (Figure 1#8): typhoid, dysentery, LRI, UTI, MDR TB, sepsis, HAP, IAI and necrotising fasciitis. Given diagnostic uncertainty, we inflated typhoid cases to account for suspected cases that may require antibiotics for CTAs with typhoid burden above 50 per 100,000. We calculated the proportion of UTIs and LRIs for which Watch antibiotics are recommended (upper UTIs and severe LRIs) using estimates from the literature. For each infection, we multiplied the case counts by the maximum DDDs recommended to treat a case then summed these estimates to get total Watch antibiotic needs (supplemental methods).

Expected Access antibiotic use was calculated as the residual volume after subtracting estimated Watch and Reserve requirements from expected total antibiotic use. (Figure 1#9). We assumed optimal levels of Not Recommended antibiotics should be zero and did not estimate these.

To account for uncertainty in the GBD infection and resistance estimates, we sampled 1000 draws from the distribution of each infection (supplemental methods). All estimates for optimal total, Reserve, Watch and Access were repeated on the 1000 draws per country. Final DDD estimates were converted back to DID using population estimates and presented as median and 95% confidence intervals (CI) of the draws. We also summarised the global estimated billions of DDD and global percentage Access antibiotics.

We compared estimates of optimal total, Access, Watch and Reserve DID to actual DID for CTAs where data were available in IQVIA MIDAS®. CTAs were classified as having actual use higher or lower than optimal antibiotic use when actual DID exceeded or remained below ≥80% of draws of optimal DID respectively.

We conducted several sensitivity analyses to explore the impact of our assumptions of adjusted case counts on expected ABU, particularly the impact on Watch ABU and the impact of benchmark country selection on expected ABU (Table S7). We estimate a “high Watch” scenario (scenario 2), an “unadjusted case counts” scenario (scenario 3) and an “alternate benchmark CTA” scenario (scenario 4).

The variables used in each component of data preparation and analysis are in Table S8. All analyses were done in R version 4.4.3. The funder had no involvement in the study.

## Results

### Antibiotic Use Regression Model

We found no clear associations between any of the 22 clinical infection, ABR burden and sociodemographic variables, and national AWaRe DID. Posterior estimates were dominated by the intercept terms, indicating that most variation in Access, Watch, and Reserve DID was unexplained by the included covariates (Figure S16, Table S9). Therefore, we did not use regression-based predictions to estimate expected national AWaRe antibiotic use.

### AWaRe ABU Benchmarking

#### Clustering and benchmark selection

The chosen LCM separated the 186 CTAs into four clusters (Figure 2). The covariates contributing most strongly to cluster separation were the proportion of population living below the international poverty line, dysentery incidence, population age group proportions and sepsis incidence. We ordered clusters by median GNI per capita where Cluster 1 included the lowest-income CTAs and Cluster 4 included the highest income CTAs (Supp.pp:50-53).

**Figure 2.**
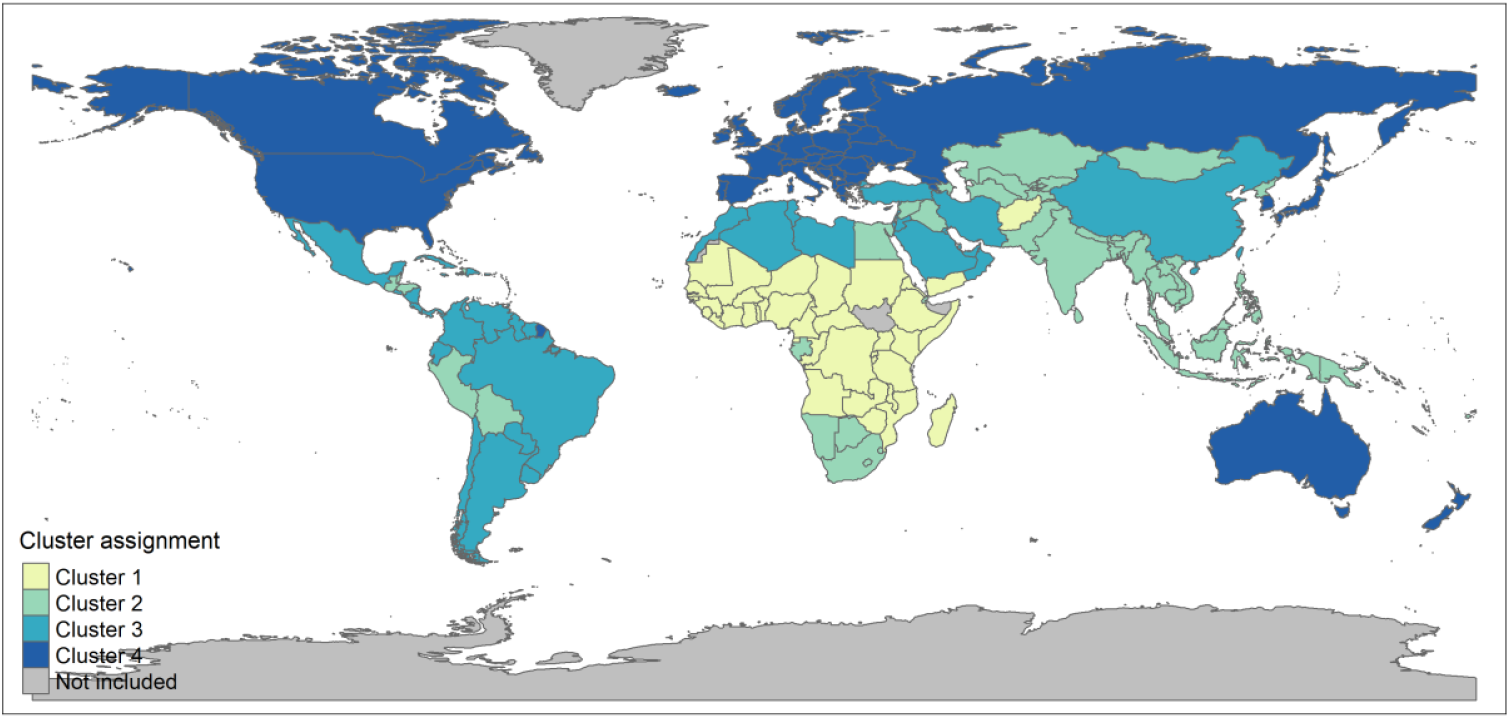
Map showing which of 186 countries, areas, territories (CTAs) clustered together using Latent Class Model (LCM) based on infection burden, antibiotic resistance, sociodemographic factors and healthcare infrastructure and access factors. Cluster 1 countries are generally lowest income countries and Cluster 4 countries are generally highest income countries.

Suitable benchmark CTAs were not identified within Cluster 1 due to lack of data from LICs in IQVIA MIDAS®, and none were identified within Cluster 2 because CTAs with lower antibiotic use had higher infection mortality than those in Cluster 3, indicating potential underuse.

Morocco (Cluster 3) was used as the benchmark country for Clusters 1-3 given its similar healthcare access and quality index (HAQI) to Clusters 1 and 2 CTAs and it provides a minimum estimate for these CTAs. Switzerland was chosen as the benchmark country for Cluster 4. For a sensitivity analysis, we used Colombia, which has a higher HAQI and lower ABU than Morocco, as the benchmark CTA for Clusters 1-3 (Supp.pp:54).

#### Expected ABU Estimate

Across 186 CTAs, we estimated 43.0 billion DDDs (95%CI: 35.4 billion – 57.7 billion) of total antibiotics were needed in 2019 to treat their overall infectious disease burden when also accounting for ABR and population risk factors. By AWaRe group, 32.9 billion DDD would be Access (95%CI:35.4-57.7 billion), 8.9 billion would be Watch (95%CI:8.4-9.5 billion) and 1.2 billion would be Reserve (95%CI:0.6-2.3 billion). We estimated that 76% (95%CI: 71%-83%) of the global antibiotics needed would be Access antibiotics (Table S11).

CTAs in Clusters 1 and 2 were estimated to require higher total antibiotic volumes, particularly Watch and Reserve, compared to CTAs in Clusters 3 and 4 (Figure 3). Median estimated optimal total DID was 18.6 (95%CI:14.7–25.8) for Cluster 1, 16.3 DID (95%CI:12.8–22.6) for Cluster 2, 14 DID (95%CI:11–19.6) for Cluster 3, and 12.5 DID (95%CI:11.3–13.9) for Cluster 4. Median optimal Watch use was highest in Cluster 1 (5.8 DID, 95%CI:5.4-6.4) and lowest in Cluster 4 (1.1 DID, 95%CI:1–1.1). Median estimated optimal Reserve use was also highest in Cluster 1 (0.44 DID, 95%CI:0.23–0.77) and Cluster 2 (0.69 DID, 95%CI:0.34–1.59), and lowest in Cluster 3 (0.10 DID, 95% CI:0.04–0.31) (Table 1). There was marked variation in estimated optimal use between CTAs, even within the same Cluster with some CTAs showing similar estimated optimal total DID but differing substantially in the required AWaRe volumes (Figure 3, Table S12).

**Table 1.** Summary of the median and 95%CI estimated optimal antibiotic use for Total, Access, Watch and Reserve (AWaRe) antibiotic use by benchmark group in defined daily doses (DDD) per 1000 inhabitants per day (DID) and percentage of global DDD (95%CI) of antibiotic use by AWaRe accounted for by each benchmark group for primary analyses.

| Estimated optimal median DID (95% CI) |  |  |  |  |
| --- | --- | --- | --- | --- |
| Clusters* | Total DID | Access DID | Watch DID | Reserve DID |
| Cluster 1 (lowest income)) | 18.6 (14.7, 25.8) | 12.3 (8.4, 19.5) | 5.8 (5.4, 6.4) | 0.44 (0.23, 0.77) |
| Cluster 2 | 16.3 (12.8, 22.6) | 10.9 (7.3, 17.3) | 4.6 (4.2, 5) | 0.69 (0.34, 1.59) |
| Cluster 3 | 14 (11, 19.6) | 12.6 (9.6, 18.1) | 1.3 (1.2, 1.4) | 0.1 (0.04, 0.31) |
| Cluster 4 (highest income)) | 12.5 (11.3, 13.9) | 11.1 (9.9, 12.5) | 1.1 (1, 1.1) | 0.27 (0.14, 0.5) |
| Percentage of estimated optimal global AWaRe DDD use by cluster |  |  |  |  |
| Clusters | % of Total | % of Access | % of Watch | % of Reserve |
| Cluster 1 (lowest income) | 17.6 (16.6, 18.6) | 15.2 (13.3, 17.1) | 26.7 (24.7, 29) | 15.3 (8.8, 22.9) |
| Cluster 2 | 40.3 (38.3, 42.5) | 35.5 (30.7, 39.4) | 54.9 (52.5, 57.5) | 64.4 (49.9, 78.8) |
| Cluster 3 | 28 (26.3, 29.8) | 32.9 (31.1, 35.1) | 12.6 (11.5, 13.8) | 7.2 (3.3, 19.1) |
| Cluster 4 (highest income) | 14.1 (10.4, 17.7) | 16.5 (11.3, 22.2) | 5.8 (5.4, 6.1) | 11.5 (6.9, 17.3) |

**Figure 3.**
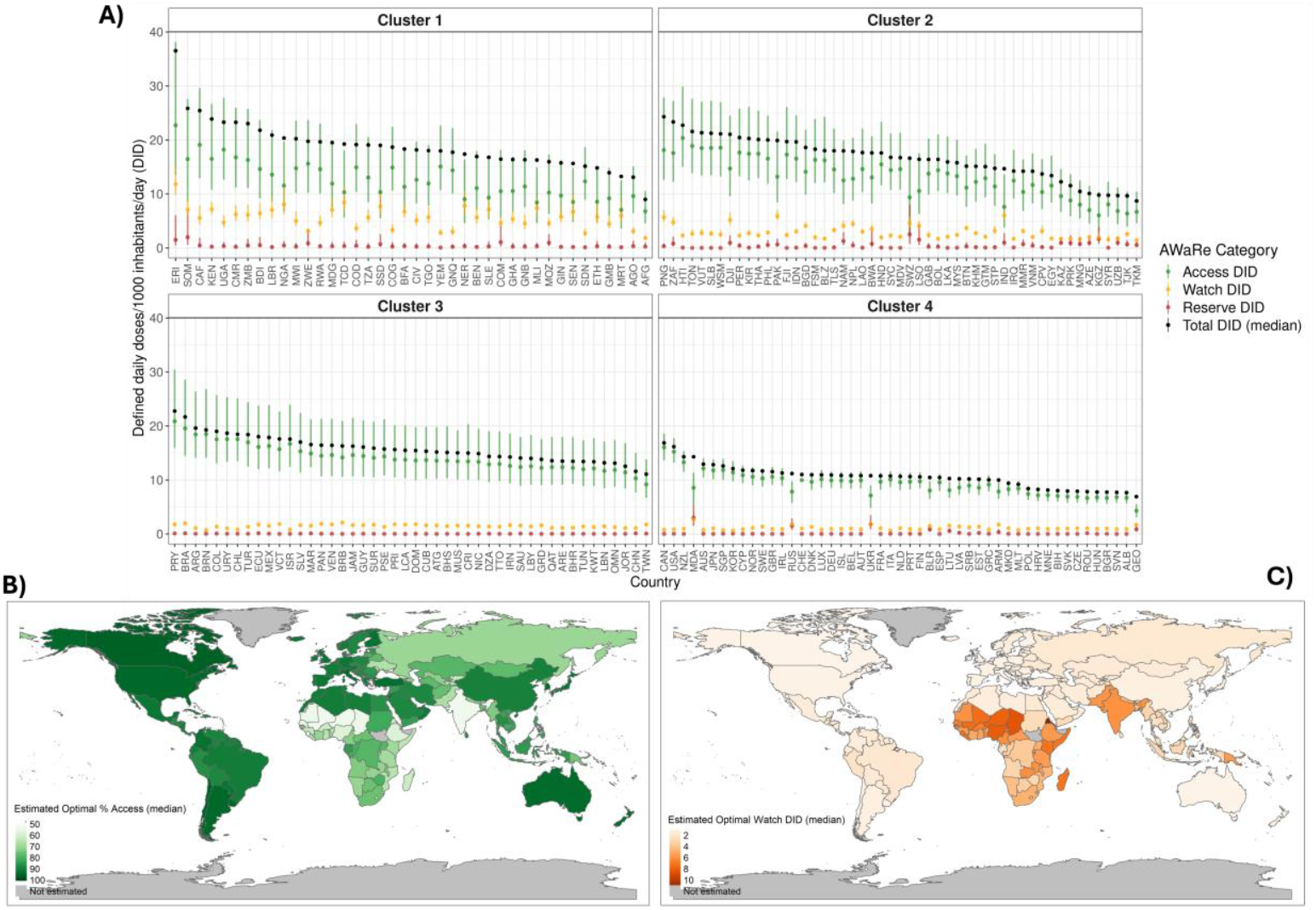
A) Estimates of optimal Total, Access, Watch and Reserve defined daily doses (DDD) per 1000 inhabitants per day (DID) for 186 CTAs using clusters. Access, Watch and Reserve are presented as the median with 80% (thick bar) and 95% (thin bar) CIs and Total DID is presented as the median. CTAs are identified using ISO-3 codes. B) Estimated optimal percent Access (median) for 186 CTAs. C) Estimated optimal DID Watch antibiotics (DID).

Overall, if estimated optimal levels were met, 80.7% (95%CI: 68.5%-88.9%) of global Reserve antibiotics and 81.7% (95%CI:80.3%-82.9%) of global Watch antibiotics are estimated to be needed by CTAs in Clusters 1 and 2. Only 14.1% (95%CI:10.4%-17.7%) of total global antibiotics are expected to be needed in Cluster 4 (highest income) CTAs (Table 1). 67% (124/186) of CTAs are estimated to be able to use at least 70% of Access antibiotics at optimal levels (using a 95% probability) (Figure 3, Table S13).

#### Actual compared to optimal ABU

We compared reported actual ABU to estimated optimal ABU for the 67 CTAs with available 2019 IQVIA MIDAS® data, indicating patterns of both overuse and underuse (Figure 4). Cluster 1 contained no CTAs with available actual antibiotic use data, so comparisons were restricted to Clusters 2, 3 and 4. Across all included CTAs, 72% (48/67) used higher total antibiotic volumes than estimated optimal (at 80% probability level). Almost all CTAs (99%, 66/67) used more Watch antibiotics than optimal, indicating widespread overuse in this category. Overuse of total antibiotics was most frequent in high income settings (Figure S23); 87% (33/38) of CTAs in Cluster 4 exceeded estimated optimal Total DID, compared with 60% (6/10) in Cluster 2 and 47% (9/19) in Cluster 3. In contrast, 69% (46/67) of CTAs overall used less Reserve antibiotics than estimated optimal. This was most pronounced in Cluster 2 (80%, 8/10) and Cluster 3 (95%, 18/18) although potential underuse of Reserve antibiotics was also common in Cluster 4 (53%, 20/38). Overall, 58% (39/67) of CTAs were using less Access antibiotics than estimated optimal.

**Figure 4.**
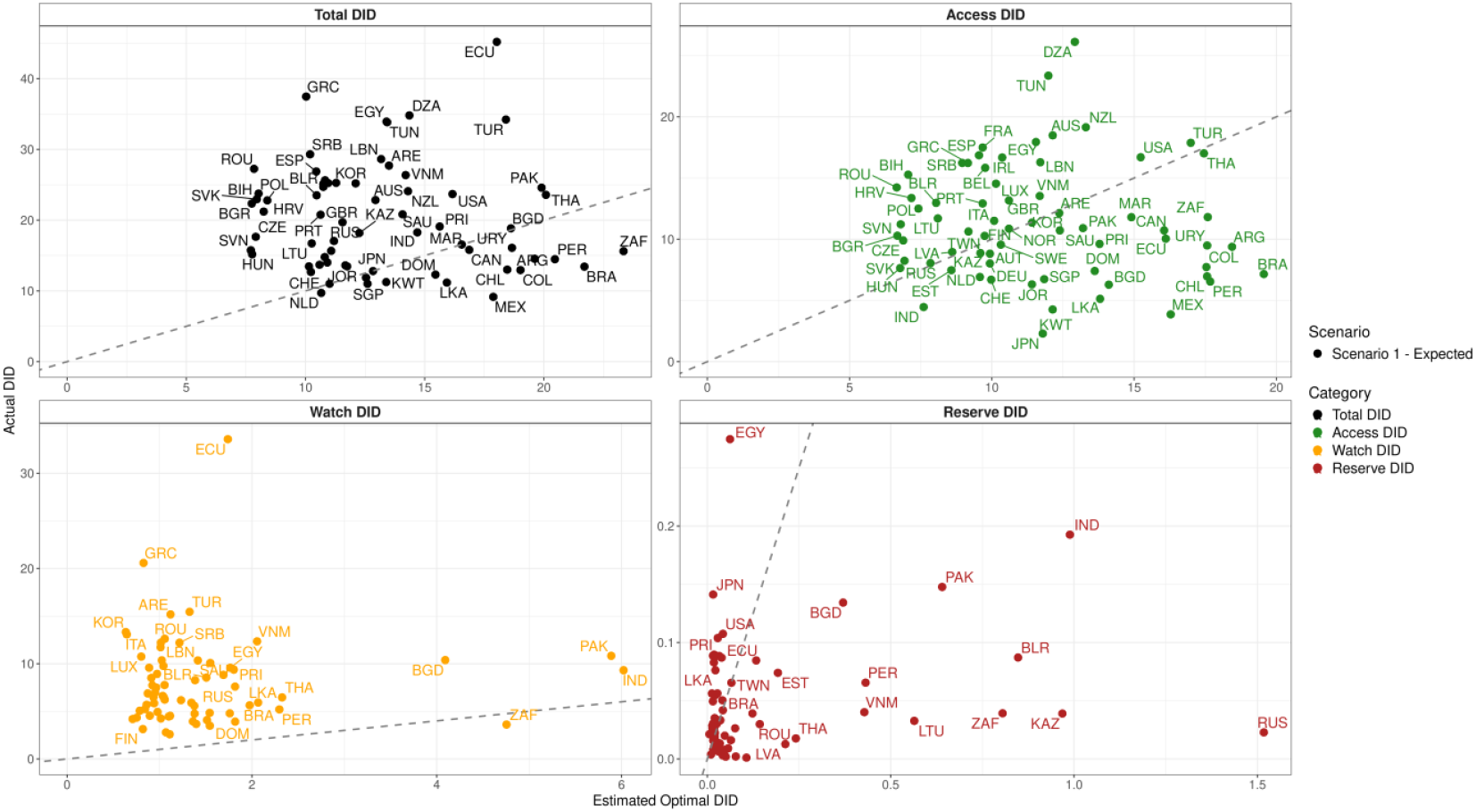
Actual use compared to. median estimated optimal antibiotic use for Total and AWaRe group in defined daily doses (DDD) per 1000 inhabitants per day (DID). Dashed line indicates where observed = expected. Source: Actual DID based on IQVIA MIDAS® data, Jan-Dec 2019, reflecting estimates of real-world activity. Copyright IQVIA. All Rights Reserved

#### Sensitivity analyses

Under a high Watch scenario (scenario 2), we estimated the same total 43.0 billion DDDs (95%CI:35.4 billion–57.7 billion) were required, with Access antibiotics comprising 66% (95%CI:58%-75%). When using unadjusted cases and excluding TB (scenario 3), 42.8 billion DDDs (95%CI:35.2 billion–57.4 billion) would be required, of which 85% (95%CI:81%-89%) would be Access antibiotics. When using a benchmark country with lower total DID (Colombia) for Clusters 1-3 (scenario 4) we estimated an expected 31 billion DDDs (95%CI:25.5 billion– 44.3 billion) globally and 67% Access (95%CI:59%-77%) (Table S11, Table S13). When comparing global optimal Watch antibiotic need, our high Watch scenario (4.7 DID, 95%CI:4.4-5.0 DID) is only expected to be 2.5 DID higher than our low, unadjusted scenario (2.2 DID, 95%CI:2.1–2.4 DID) (Supp.pp:56-103).

## Discussion

We present the first estimates of expected levels of optimal national antibiotic use by AWaRe group for 186 CTAs based on infection burden, antibiotic resistance, population demographics, national income, health system infrastructure and access measures.

In our ecological regression model, we did not identify clear associations between clinical infection burden, resistance or sociodemographic covariates and national antibiotic use by AWaRe group. These findings suggest that the marked differences in antibiotic use seen between CTAs are not well explained by available national-level clinical, resistance and sociodemographic factors, but are likely influenced by difficult to measure country-specific cultural, behavioural, health system, or regulatory factors that shape antibiotic use practices.^(24,25)^

We estimated that globally at least 43.0 billion DDDs (95%CI: 35.4 billion – 57.7 billion) of antibiotics were needed in 2019 to meet clinical infection need, accounting for antibiotic resistance, population and healthcare factors, equivalent to approximately one course of antibiotics per person/year. This estimate is similar to published estimates of global antibiotic use in 2018 of 40.2 billion DDDs (95% CI: 37·2–43·7 billion).^(7)^ However, our findings suggest that HICs clinically require considerably lower total antibiotic volumes compared to low-and-middle-income CTAs (LMICs), the inverse of the current observed use patterns.^(6-8)^

This is the first study to quantify the volume of optimal global antibiotic levels by AWaRe. Our findings support the relevance of the 2024 UNGA target as based on clinical need, we found that 76% (95%CI:71%-83%) of global use should be Access antibiotics, compared to the current GLASS AMU estimate of 52%.^(26)^ However, among 67 CTAs with observed data available in our analysis, 72% used higher total antibiotic volumes than expected and 99% used more Watch antibiotics than expected.

Our analysis highlights that some CTAs with low Access use could potentially reach expected levels of both Total and Access use by replacing part of their excess Watch with Access antibiotics, while also reducing overuse of other Watch antibiotics. Access and Watch antibiotics are 97.4% of total use in GLASS AMU and 94.3% of antibiotics are oral antibiotics.^(26)^ Addressing unnecessary use of total, not recommended and oral Watch antibiotics in primary care which accounts for 80-90% of human antibiotic use, will be a critical part of future antibiotic policy and stewardship interventions, alongside improving access for patients.^(27-29)^ The specific antibiotics within AWaRe groups to be prioritised to improve access will necessarily depend on country-specific infection patterns, resistance profiles, and other priorities.

Our analysis enables estimation of the minimum volume range of AWaRe antibiotics required to treat the given disease burden in a CTA. Previous analyses have focused on specific infections or antibiotics^(30,31)^ while we provide estimates across all AWaRe categories and most infections. To meet any antibiotic use target, particularly where Access antibiotics are underused, robust supply chains and affordable access to key Access antibiotics are essential. This is particularly pertinent for amoxicillin (+/-clavulanate), which is recommended for 10/12 of the most common primary care infections in the WHO AWaRe book.^(4,10)^ Our approach also highlights CTAs facing an access gap to effective antibiotics for resistant serious bacterial infections. This gap is particularly evident in CTAs with lower incomes and higher infection and resistance burdens, where >80% of countries in MIDAS® used less Reserve antibiotics than expected.

Although Reserve antibiotic use accounted for only between 0.005%-1.1% of total observed antibiotic use in MIDAS® CTAs and 0.18% in GLASS,^(26)^ infections requiring Reserve antibiotics have a high mortality^(14)^ and patients may not be getting the treatment they need,^(31)^ highlighting the importance of initiatives such as WHO/GARDP’s SECURE initiative to expand affordable access to antibiotics.^(32)^ Increasing Reserve use where required will need to be accompanied by good antibiotic and diagnostic stewardship.

Our analysis has several limitations. We used modelled estimates of infection burden and from antibiotic resistance. While our analysis accounted for reported uncertainty in these quantities and we conducted sensitivity analyses to explore these areas of uncertainty, sparse data particularly in LMICs may mean that there are important biases in these estimates that have been unaccounted for. For some infections, we assumed proportions of severe infections requiring Watch antibiotics were the same across all CTAs which may not be the case, and some areas may need to treat a higher proportion of certain infections (e.g. LRI) with Watch antibiotics. However, we assume that all LRIs are treated with antibiotics (Access and Watch) which may in converse overestimate the estimated optimal antibiotics needed as a large proportion of these infections are caused by viral pathogens. Antibiotic use data were obtained from IQVIA MIDAS®, a widely used source for estimating antibiotic consumption;^(6,7,31,33-35)^ however, data were only available for 67 countries, for which we imputed missing hospital sector data for 26 countries, and coverage varies by country, with limited representation from low-income settings and variable public and private sector coverage. Availability of IQVIA MIDAS® data limited the countries available for benchmark selection particularly in clusters 1 and 2. By using the benchmark country from cluster 3 for clusters 1-3 we were still able to estimate a minimum range of antibiotics required in these countries given burden of disease estimates. When comparing actual vs. optimal, IQVIA may underestimate current use in some CTAs compared to other sources of national antibiotic use (e.g. GLASS); however, our estimates illustrate the general utility of this method to identify areas of overuse and underuse. This method could be extended to any trusted local data source for a country to compare actual and optimal use. This analysis is focused on the national level, therefore may mask local variation, with both underuse due to lack of access and overuse within a country.

The AWaRe benchmarking framework offers a data-driven method for estimating antibiotic need and assist countries to evaluate their levels of use compared to optimal patterns. As data availability and quality improve – through reduced reliance on spatiotemporal smoothing in GRAM/GBD estimates and improving national data collection on infection burden and antibiotic use – the accuracy, precision and policy utility of these estimates will strengthen.

Benchmarking is an established quality-improvement tool in healthcare^(36)^ and could similarly be used to monitor progress towards national antibiotic use targets, inform procurement strategies, and enhance shared learning of policy interventions across countries.

## Data Availability

The underlying IQVIA MIDAS® antibiotic use dataset is proprietary and cannot be publicly shared. Analysis code, underlying datasets for all other variables and derived estimates will be made available on GitHub with publication.

## Acknowledgments

The ADILA Project funded by the Wellcome Trust [222051/Z/20/Z].

## Notes

### Competing Interest Statement

The authors have declared no competing interest.

### Summary of Updates

Re-uploading the supplementary material that was lost in the previous version.

