## Supplemental material for "Benchmarking AWaRe: estimating optimal levels of AWaRe antibiotic use in 186 countries, territories and areas based on clinical infection and resistance burden"

Cook, Aislinn et al.

Figure 4. Study schema for deriving estimates of expected AWaRe antibiotic use based on infection burden, antibiotic resistance, socio-demographics, healthcare infrastructure and access using the WHO AWaRe book as a clinical framework. This figure is also presented in the main manuscript but is included here for ease. 1. Data preparation, 2. Missing data imputation, 3. Antibiotic use regression model, 4. Latent Class Model, 5. Benchmark CTA selection, 6. Estimate optimal total DID, 7. Estimate optimal Reserve DID, 8, Estimate optimal Watch DID, 9. Estimate optimal Access DID ........................................................................................................................ 19

Figure 5. Distribution of covariates by World Bank Income Group. GNI = gross national income, UHC = universal health coverage, LRI = lower respiratory tract infection, MDR & XDR TB = multi-drug resistant and extremely-drug resistant tuberculosis, URTI = upper respiratory tract infection, UTI = urinary tract infection, CR = carbapenem resistant, ESBL = extended-spectrum beta-lactamase, MRSA = methicillin-resistant Staphylococcus aureus, VR. E faecium = vancomycin-resistant Enterococcus faecium, VR-S.aureus = vancomycin-resistant Staphylococcus aureus, Prop = proportion, pop = population, PCV = pneumococcal conjugate vaccine ................................................................................................................................ 22

Figure 6. Distribution of covariates by WHO Region. GNI = gross national income, UHC = universal health coverage, LRI = lower respiratory tract infection, MDR & XDR TB = multi-drug resistant and extremely-drug resistant tuberculosis, URTI = upper respiratory tract infection, UTI = urinary tract infection, CR = carbapenem resistant, ESBL = extended-spectrum beta-lactamase, MRSA = methicillin-resistant Staphylococcus aureus, VR. E faecium = vancomycin-resistant Enterococcus faecium, VR-S.aureus = vancomycin-resistant Staphylococcus aureus, Prop = proportion, pop = population, PCV = pneumococcal conjugate vaccine..................................... 23

Figure 7. Bayesian information criterion (BIC) of latent class models considering 1 to 6 clusters using available covariates (n=40) for 186 analysis CTAs. ............................................................................................. 24

Figure 8. Point estimates of infection incidence vs. infection mortality by benchmark group and coloured by Healthcare Access and Quality Index (HAQI) showing the MIDAS® CTAs with available antibiotic use data available.......................................................................................................................................... 26

Figure 9. MIDAS® CTAs with available antibiotic use data by benchmark group (note: no benchmark group 1 CTAs had antibiotic use data available in MIDAS®) showing infection mortality per 100,000 people against total DID sized by % Access antibiotics and coloured by Healthcare Access and Quality Index (HAQI). Uncertainty bars are for infection mortality. Source: Total DID based on IQVIA MIDAS® data for 2019, reflecting estimates of real-world activity. Copyright IQVIA. All Rights Reserved.......................................... 27

Figure 14. A figure describing the process of estimating what percentage of urinary tract infections (UTIs) could be upper UTIs and thus requiring Watch antibiotics per the AWaRe Book. The figure outlines the classification strategy for assumed lower UTI requiring Access antibiotics and upper UTI requiring Watch antibiotics from cases in Clinical Practice Research Datalink (CPRD) between 2015 and 2021. The figure shows three scenarios and we use scenario 2 for our analysis which gives the highest estimate of upper UTI (21%). .......... 40

Figure 15. Illustration of the subset of cases from GBD considered A) Upper UTI and B) severe LRI used for estimating Watch antibiotic use out of total cases. For UTI, in the primary analysis, the subset of cases was taken after adjusting the case counts (described in supplemental methods 1.2.2.5) .......... 41

Figure 16. Results of antibiotic use regression model showing the distribution of the posterior draws and 95%CrI for Access, Watch and Reserve antibiotic use. DID = defined daily doses (DDD)/1000 inhabitants/year. GNI = gross national income; UHC = universal health coverage; PCV = pneumococcal vaccine; LRI = lower respiratory infection; URI = upper respiratory infection; UTI = urinary tract infection; STI = sexually transmitted infection; MDR & XDR TB = multidrug resistant and extensively drug resistant tuberculosis; EBSL = extended spectrum beta-lactamase producing Enterobacterales; MRSA = methicillin-resistant staphylococcus aureus; CRO = carbapenem resistant organism; VRO = vancomycin-resistant organism.......... 47

Figure 17. Discriminative power of the variables included in the four groups cluster model. Negative bars indicate the variable was not discriminative and was not used by the model. .......... 49

Figure 18. Distribution of covariates by cluster for the four-group cluster model. GNI = gross national income, UHC = universal health coverage, LRI = lower respiratory tract infection, MDR & XDR TB = multi-drug resistant and extremely-drug resistant tuberculosis, URTI = upper respiratory tract infection, UTI = urinary tract infection, CR = carbapenem resistant, ESBL = extended-spectrum beta-lactamase, MRSA = methicillin-resistant Staphylococcus aureus, VR. E faecium = vancomycin-resistant Enterococcus faecium, VR-S.aureus = vancomycin-resistant Staphylococcus aureus, Prop = proportion, pop = population, PCV = pneumococcal conjugate vaccine .......... 50

Figure 19. Comparison of cluster assignments to World Bank Income Groups. .......... 51

Figure 20. Comparison of cluster assignments to WHO regions .......... 52

Figure 21. Infection deaths per 100,000 people and Total DID by cluster for CTAs with data available in IQVIA MIDAS® showing percentage Access antibiotics (size) and Healthcare Access and Quality Index (HAQI) (colour) with potential benchmark CTA codes highlighted in red (MAR (Morocco), COL (Colombia) in benchmark group 3 and CHE (Switzerland) in benchmark group 4). Source: Total DID based on IQVIA MIDAS® data for 2019, reflecting estimates of real-world activity. Copyright IQVIA. All Rights Reserved... 53

Figure 22. Distribution of the 1000 draws of estimated expected Total, Access, Watch and Reserve (AWaRe) antibiotic use by cluster for primary analyses. Note: x-axis scales are different between the AWaRe categories. .......... 54

Figure 23. Median observed (IQVIA MIDAS®) vs. median estimated expected antibiotic use by AWaRe category and benchmark group for primary analyses for CTAs in IQVIA MIDAS® with observed antibiotic use data available. Dashed line indicates where observed = expected. Source: Actual DID based on IQVIA MIDAS® data for 2019, reflecting estimates of real-world activity. Copyright IQVIA. All Rights Reserved... 54

Figure 28. Comparison of actual (IQVIA MIDAS) vs. median estimated optimal by analysis scenario (rows) and AWaRe category (columns) for CTAs in IQVIA MIDAS® with observed antibiotic use data available Dashed line indicates where observed = expected. Scenario 1 = primary analysis, Scenario 2 = high watch scenario, Scenario 3= unadjusted case counts & excl. TB, Scenario 4 = alternate benchmark CTA. Source: Actual DID based on IQVIA MIDAS® data for 2019, reflecting estimates of real-world activity. Copyright IQVIA. All Rights Reserved ........ 93

Table 6. Highest DDDs per treatment course of the infections included in analysis for estimating Watch antibiotic use. Where the AWaRe Book recommends several Watch antibiotics, we used the Watch antibiotic with the highest DDD per treatment course. There are also several infections for which the first choice of treatment is recommended as either an Access or Watch antibiotic and for these we use the Watch DDD estimate unless otherwise stated in the case adjustments. Note that some oral antibiotics have higher DDDs than parenteral treatment recommended for the same infection. ........ 37

### 1 SUPPLEMENTAL METHODS

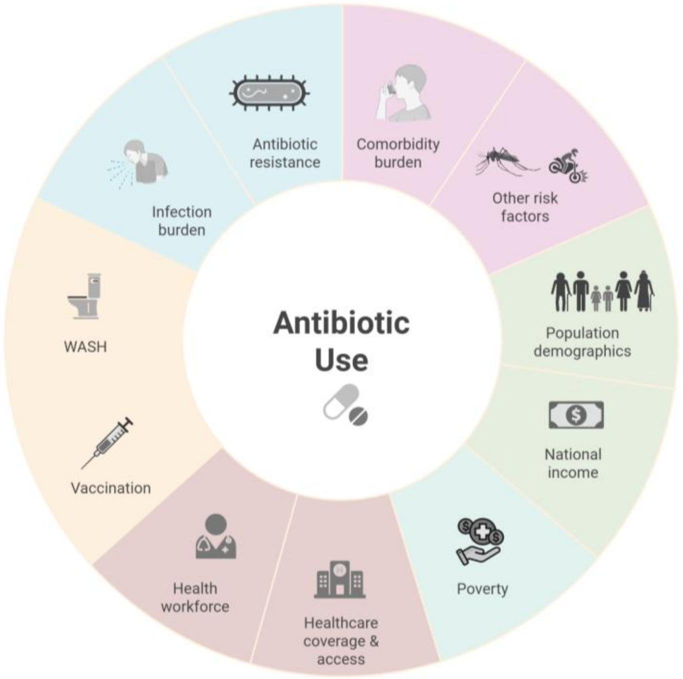

*Figure 1. Factors that would be expected to impact on necessary antibiotic use for both total use and AWaRe antibiotic use. Created in BioRender. Cook, A. (2025) https://BioRender.com/d94em8e*

#### 1.1 DATA SOURCES AND PROCESSING

##### 1.1.1 Antibiotic Use Data

IQIVA MIDAS® quarterly sales data for 2019 for antibiotics were obtained under license from IQVIA and reflect estimates of real-world activity (Copyright IQVIA. All rights reserved). Antibiotic data in 2019 covers 75 countries, territories and areas (CTAs); we excluded the aggregate areas of French West Africa and Central America and Hong Kong given unavailability of covariates leaving 72 CTAs. Antibiotic data is reported by kilograms of individual product and separately by healthcare sector (hospital or retail). We removed any antibiotics and combinations that were exclusively for tuberculosis and any combinations of methenamine (a urinary antiseptic). Kilograms were converted to defined daily doses (DDD) using the WHO/ATC methodology.[1] Where DDDs were unavailable for a given product on the WHO/ATC site, best efforts were made to find a relevant DDD to ensure as much data as possible was retained for analysis. Antibiotics that were from the "Not Recommended" AWaRe category or were in MIDAS® but unclassified by the AWaRe system were grouped together into a single "Not Recommended" category.[2]

Data were reported from the retail sector for all 72 CTAs and from the hospital sector for 46 CTAs. DDDs were summed separately by sector and AWaRe category and converted to DDD/1000 inhabitants/day (DID) using World Bank population estimates.

##### 1.1.2 Infection data

We extracted 2019 incidence and mortality for infections where antibiotics are recommended in the WHO AWaRe Antibiotic book from the Global Burden of Disease 2021 release (GBD)[3-5] where available, including cellulitis, upper respiratory infections, otitis media, lower respiratory infections (excludes hospital-acquired pneumonia (HAP)), , multi-drug resistant (MDR) and extensively drug-resistant (XDR) tuberculosis (for which non-TB-exclusive antibiotics are recommended), typhoid and paratyphoid, all-cause diarrhoea, urinary tract infections, chlamydia, syphilis, gonorrhoea, and trichomoniasis. Sepsis was not available in the GBD 2021, so incidence and mortality were extracted from a previous GBD publication for the year 2017; we assumed sepsis rates were similar between 2017 and 2019.[6] We include MDR and XDR TB cases because treatment recommendations include Watch (levofloxacin or moxifloxacin) and Reserve (linezolid) antibiotics in addition to exclusively TB medications.

The AWaRe Book recommends antibiotics for bloody diarrhoea/dysentery, not for acute watery diarrhoea, therefore to estimate dysentery-specific cases of diarrhoea, we approximated using diarrhoeal bacterial aetiologies commonly associated with dysentery per the AWaRe Book[7]: *Shigella* spp., enterotoxigenic *E. coli*, non-typhoidal *Salmonella spp.* and *Campylobacter spp.*. Aetiology-specific and all-cause diarrhoea years lived with disability (YLD) were used from GBD.[8] We estimated the incidence of each aetiology by estimating the non-fatal attributable fraction of all-cause diarrhoea cases attributed to each pathogen and multiplying this by the incident all-cause diarrhoea cases. Non-fatal attributable fraction was estimated using the YLDs for each aetiology divided by the YLDs for all-cause diarrhoea. That fraction was then multiplied by the total diarrhoea cases to get the aetiology-specific case counts. We used the same process to estimate incident cases of cholera. Diarrhoea aetiology-specific mortality is provided by GBD so this was used directly.

**<u>Estimates of incidence for non-GBD infections</u>**

For hospital infections in the AWaRe Book but not included in GBD, such as hospital-acquired pneumonia (HAP)[9] and intra-abdominal infections (IAI), these were estimated using data from alternate sources.

For HAP, we used surveillance data from the European Centres for Disease Prevention and Control (ECDC) from point prevalence surveys (PPS) on HAI for 2022-2023.[10] ECDC estimates the total number of HAP cases per year in European hospitals therefore we used the estimate of the number of HAP cases per year in the participating CTAs (953,971 cases (95%CI: 626,128-1,449,959) to estimate the number of HAP cases in other CTAs. We took the ratio of the total HAP cases in the ECDC HAI PPS CTAs to the sum of the LRI cases from GBD for the same CTAs. We then multiplied this ratio of HAP:LRI by the number of LRI cases in all other CTAs to estimate total number of HAP cases per CTA assuming the ratio between HAP:LRI is the same in all CTAs. This calculation was done draw-wise on 1000 draws from the distribution of ECDC HAP cases and 1000 draws from the distribution of LRI cases from GBD to account for uncertainty from both case estimates.

For IAI, we used data extracted from Versporten et al[11] (extracted from eTable2 in the publication) of the distribution of patients receiving antibiotics with each type of diagnosis included in the study.

We assume all IAI cases require antibiotics, thus took the ratio of the total IAI cases in the study to the total sepsis cases reported (ratio = 2.6 IAI cases to sepsis cases), assuming the case mix is similar across CTAs given limited data by region or CTA. Using this ratio, we multiply the GBD sepsis case estimates for each CTA by this ratio to get a national estimate of total IAI cases. This operation was repeated on 1000 draws from the distribution of sepsis cases to populate uncertainty around IAI cases.

##### 1.1.3 Antibiotic resistance data

We used interim data from the GRAM project (see GRAM methods for complete details[12,13]) to estimate incidence of infections caused by resistant pathogens, where resistance may require shifting AWaRe category (e.g. where carbapenem resistance is common, sepsis treatment may more frequently be a Reserve antibiotic (e.g. colistin) than a Watch antibiotic . We focused on extended-spectrum beta-lactamase (ESBL) *Enterobacterales,* carbapenem-resistant *Acinterobacter baumanii* (CRAB)*, Pseudomonas aeruginosa* (CRPA)*, Enterobacterales* (CRE)*,* methicillin-resistant *Staphylococcus aureus* (MRSA), vancomycin-resistant *Staphylococcus aureus* (VRSA) and *Enterococcus faecium* (VR-*E.faecium*).

First, we estimate of the relative prevalence of each bug-drug specific resistance pattern and relative prevalence of that pathogen within the total number of pathogens for each CTA. To estimate incidence of resistant cases of severe infections, we multiplied the proportions of resistance and proportions of pathogens by the case counts for sepsis from GBD (Equation 1).

$$Cases\ of\ bug - drug\ XY = Proportion\ of\ total\ caused\ by\ bug\ X * Proportion\ of\ bug\ X\ resistant\ to\ drug\ Y * sepsis\ cases$$

*Equation 1. Formula for calculating cases of specific pathogen (bug) and antibiotic (drug) combinations where X represents pathogen of interest and Y represents antibiotic of interest.*

##### 1.1.4 Other covariate data

We included other covariates that may be associated with antibiotic use at a population level including proportion of population in different age groups (0-4, 5-14, 15-64, 65-74, 75+ years),(14,15) proportion of population living in rural areas,(16,17) proportion of population with comorbidities(18),universal health coverage (UHC) index,(19,20) vaccination coverage for pneumococcal vaccine(21-23) and rotavirus vaccine(22,24,25), time to travel to health facilities(26), pharmacists(27,28), doctors(27-29) and nurses(27,28,30) per 1,000 people, gross national income (GNI) per capita(31-37), proportion of population living beneath the $2.15 per day poverty line(38), proportion of the population with access to safe sanitation(39). We also included malaria incidence estimates from GBD(5) as febrile illnesses in malaria endemic areas are commonly treated with antibiotics even if unnecessary,(40) protein energy malnutrition prevalence estimates(5) which can put people at risk of increased susceptibility to infection and worse infection outcomes and incidence of burns and wounds from road traffic accidents and fires as these injuries may be at risk of infection. Data sources for covariates are summarised in Table 1.

Where data was not in the primary data source, alternate data sources were sought to limit missing data as much as possible. Generally, we sourced as many years as possible of data between 1998 and 2023 for our covariates for processing even where there was missing data for the year of interest (2019). Linear interpolation using the *zoo* package in R was used in some cases to fill in missing years of data where data for 2019 was not available but at least two other years were available. Data processing for covariates is described below and in Table 1..

For healthcare workforce estimates of doctors, nurses and pharmacists per 1000 people, we used a mix of World Bank World Development Indicators,(29,30) WHO Global Data Observatory(27) and Global Burden of Disease estimates.(28) For doctors and nurses per 1000 people, our primary data source was World Bank. Where data was missing for any CTA-years (1998-2023) from World Bank, we used estimates from WHO Global Data Observatory to fill in those years. If CTA-years were still missing after combining World Bank and WHO data, we used linear interpolation where at least two years of data were available to estimate 2019 work force. Data for pharmacists per 1000 people was only available in the WHO Global Data Observatory; linear interpolation was used to fill in missing years where at least two years of data were available. Where data was missing for CTAs for all years (i.e. no available data in World Bank or WHO), we used GBD estimates of healthcare workforce for doctors, nurses and pharmacists for 2019(28) for these CTAs.

Universal health coverage (UHC) service coverage index data was sourced from the World Bank World Development Indicators database(19) which is a combined index measuring access to affordable essential healthcare using tracer indicators across the health spectrum. Generally, UHC was reported on evenly numbered years (e.g. 2016, 2018, etc) so linear interpolation was used to estimate coverage in the missing years. Where a CTA did not have any data available or only one year available in World Bank, alternate UHC index data were used from Global Burden of Disease estimates of progress towards Sustainable Development Goals.(20)

Travel time to healthcare facilities was estimated using spatial raster files from Weiss et al.(26) which provide pixel-level estimates of motorised travel time to the nearest healthcare facility(41). To derive CTA-level estimates, we combined these data with gridded population counts from WorldPop(42) and calculated population-weighted average travel time to healthcare for each CTA.

Vaccination coverage for pneumococcal conjugate vaccine (PCV) and rotavirus vaccine were downloaded from the WHO Immunization Data portal (https://immunizationdata.who.int/global?topic=&location=). For PCV, we used the coverage estimate of the percentage of children receiving the final dose as estimated by WHO/UNICEF estimates of national immunization coverage (WUENIC).(23) For rotavirus we used the estimates of percentage of children receiving the final dose (either 2 or 3 doses depending on which vaccine is used) as estimated by WUENIC(25). Where a coverage estimate wasn’t available for a year, we checked whether the vaccine had been introduced in the CTA using data from VIEW-hub from the International Vaccine Access Center (IVAC).(21,24) Where the year of universal introduction was after 2019, we assumed that vaccination coverage prior to this was zero; where the introduction status was indicated as "No Decision", "Gavi approved/approved with clarification", "Gavi plan to apply", "Program Suspended", "Non-Gavi planning introduction", or "Private Market Use", we also assumed coverage was 0 in the year of interest (this may have underestimated true coverage if there had been large private market use prior to universal introduction). Where vaccination coverage estimates were still missing for CTAs after checking introduction year (either no data in WUENIC or the vaccine had been introduced but no estimates), we used vaccination coverage estimates from the Global Burden of Disease study to fill in missing data.(22)

Data on proportion of the population living below $2.15 (2017 PPP) poverty line and proportion of the population with access to safe sanitation were sourced from World Bank.[38,39] After interpolation, 40 CTAs were missing poverty estimates and 66 CTAs were missing sanitation estimates for 2019.CTAs missing data for 2019 were imputed using multiple imputation by chained equations (MICE) (see supplemental methods section 1.1.5 for details). Imputed values for safe sanitation were restricted to be not more than the proportion of population with access to basic sanitation[43] where this data was available in the same CTA (n=58/66).

Data for the proportion of population living with 1 or more comorbidity that contributes to higher risk of infection or worse infection outcomes was sourced from estimates of the population at increased risk of severe COVID-19 due to underlying health conditions[18] with the assumption that these similar comorbidities also put people at risk of worse outcomes due to bacterial infections. Estimates were downloaded from the available tool for the proportion of the population with 1+ underlying condition (https://www.dropbox.com/scl/fi/2tvsjo8w58cz50htjekk7/Covid-19-analysis-v1.50-release.xlsb?rlkey=3rygfvghipue809slzuah8jlw&e=1&dl=0; accessed 05 September 2023).

*Table 1. Summary of covariates included across the analysis workflow, data sources and process for dealing with missing data from the primary data source*

| Covariate type | Variable | Primary Source | Dealing with missing data | References | Notes |
|---|---|---|---|---|---|
| **Population demographics** | **Population age group proportions (0-4, 5-14, 15-64, 65-74, 75+)** | World Bank | Taiwan is not in World Bank | World Bank Data Bank: Population estimates and projections; accessed 08/July/2024 | |
| | | | Taiwan | Downloaded from the Taiwan Dept. of Household Registration<br>Date accessed: 25/09/2023<br>https://www.ris.gov.tw/app/en/3910 | |
| | **Rural population proportion** | World Bank | Taiwan is not in World Bank | World Bank Data Bank: World Development Indicators; Rural population (% of total pop); accessed 08/July/2024 | |
| | | | Taiwan (TWN) | UN Department of Economic and Social Affairs Population Division: World Urbanisation Prospects 2018; https://population.un.org/wup/downloads; accessed on 13 February 2025 | Used the estimated urban population percentage between 2018 and 2020 to calculate rural percentage as 100- urban % |
| **Income** | **GNI per capita** | World Bank | Several CTAs did not have GNI per capita (Atlas method, USD) in World Bank so alternate sources for these were sought | World Bank Data Bank: World Development Indicators; GNI per capita, Atlas method (current US$); accessed 08/July/2024<br><br>World Bank Country and Lending Groups; https://datahelpdesk.worldbank.org/knowledgebase/articles/906519-world-bank-country-and-lending-groups; accessed 08/July/2024 | |
| | | | Taiwan (TWN) | National Statistics Republic of China (Taiwan); 09. National Economics and Business Activities: Per capita income and consumption; https://eng.stat.gov.tw/News_Content.aspx?n=4302&s=232171; accessed on 25/09/2023 | |
| | | | Venezuela (VEN) | Maldonado, L., & Olivo, V. (2022). Is Venezuela Still an Upper-Middle-Income Country? Estimating the GNI per Capita for 2015–2021. https://doi.org/10.18235/0004612 | |
| | | | American Samoa (ASM) | https://www.doc.as.gov/post/press-release-american-samoa-statistical-yearbook-2022 | gross domestic income 2019: 647 million dollars; 8425$ estimated per capita income 2020 census |
| | | | Eritrea (ERI) | https://data.un.org/Data.aspx?q=GNI+per+capita&d=SNAAMA&f=grID%3a103%3bcurrID%3aUSD%3bpcFlag%3a1; accessed 15/Feb/2025 | Per capita GNI at current prices - USD; year = 2019 |
| | | | Greenland (GRL) | https://data.un.org/Data.aspx?q=GNI+per+capita&d=SNAAMA&f=grID%3a103%3bcurrID%3aUSD%3bpcFlag%3a1; accessed 15/Feb/2025 | Per capita GNI at current prices - USD; year = 2019 |
| | | | Guam (GUM) | https://bsp.guam.gov/guam-statistical-yearbook-2/ | Gross Domestic Income 2019: 6355 million dollars |
| | | | Northern Mariana Island (MNP) | Not found - excluded | |
| | | | North Korea (PRK) | National accounts statistics from here: https://kosis.kr/bukhan/statisticsList/statisticsListIndex.do?menuId=M_01_01_01&vwcd=MT_BUKHAN&rootId=101_001&parmTabId=M_01_01_01 | per capita GNI in 2019 = 1408000 SK won, converted to USD on 15 Feb 2025; |
| | | | South Sudan (SSD) | https://data.un.org/Data.aspx?q=GNI+per+capita&d=SNAAMA&f=grID%3a103%3bcurrID%3aUSD%3bpcFlag%3a1; accessed 15/Feb/2025 | Per capita GNI at current prices - USD; year = 2019 |
| | | | Yemen (YEM) | https://data.un.org/Data.aspx?q=GNI+per+capita&d=SNAAMA&f=grID%3a103%3bcurrID%3aUSD%3bpcFlag%3a1; accessed 15/Feb/2025 | Per capita GNI at current prices - USD; year = 2019 |

| Covariate type | Variable | Primary Source | Dealing with missing data | References | Notes |
| --- | --- | --- | --- | --- | --- |
| | **Proportion of population living below $2.15 poverty line** | World Bank | 1. Where missing in World Bank, but available for at least two years between 1998 and 2023, used linear interpolation to fill in gaps (*zoo* package in R)<br>2. Where still missing for 2019 after linear interpolation (40 CTAs), did multiple imputation with chained equations | World Bank Data Bank: World Development Indicators; Poverty headcount ratio at $2.15 a day (2017 PPP); accessed 05/July/2024 | Missing in Afghanistan, Andorra, American Samoa, Antiqua and Barbuda, Bahrain, The Bahamas, Bermuda, Barbados, Brunei Darussalam, Cuba, Dominica, Algeria, Eritrea, Equatorial Guinea, Grenada, Greenland, Guam, Guyana, Haiti, Cambodia, Kuwait, Lebanon, Libya, St. Lucia, Marshall Islands, Northern Mariana Islands, New Zealand, Oman, Papua New Guinea, Puerto Rico, North Korea, Qatar, Saudi Arabia, Singapore, SOmalia, Suriname, Turkmenistan, Trinidada and Tobago, Taiwan, St. Vincent and the Grenadines; Note: WB has updated its poverty line to $3.00 (2021 PPP) in June 2025 |
| **Healthcare workforce** | **Doctors per 1000 people** | World Bank | Not all CTA-year combinations available in World Bank | World Bank Data Bank: World Development Indicators; Physicians (per 1000 people); accessed 08/July/2024 | |
| | | WHO Global Data Observatory | 1. Where data was missing on number of doctors, used WHO Global Data Observatory estimate of number of medical doctors per 10,000 for all years 1998-2023<br>2. Where both WHO and World Bank were available but missing for 2019, used linear interpolation to fill in gaps given there were at least two data points available | The National health Workforce Accounts database, World Health Organization, Geneva; accessed 05/July/2024 (https://apps.who.int/nhwaportal, https://www.who.int/activities/improving-health-workforce-data-and-evidence) | |
| | | Global Burden of Disease | 3. Data was further missing in seven CTAs - sought alternate sources from GBD | Global Burden of Disease Collaborative Network. Global Burden of Disease Study 2019 (GBD 2019) Human Resources for Health 1990-2019. Seattle, United States of America: Institute for Health Metrics and Evaluation (IHME), 2022. | |
| | **Nurses & midwives per 1000 people** | World Bank | Not all CTA-year combinations available in World Bank | World Bank Data Bank: World Development Indicators; Nurses and midwives (per 1000 people); accessed 08/July/2024 | |
| | | WHO Global Data Observatory | 1. Where data was missing on number of doctors, used WHO Global Data Observatory estimate of number of nursing and midwifery personnel per 10,000 for all years 1998-2023 | The National health Workforce Accounts database, World Health Organization, Geneva; accessed 05/July/2024 (https://apps.who.int/nhwaportal, https://www.who.int/activities/improving-health-workforce-data-and-evidence) | |
| | | Global Burden of Disease | 2. Where both WHO and World Bank were available but missing for 2019, used linear interpolation (zoo) to fill in gaps given there were at least two data points available<br>3. Data was missing in World Bank and WHO in seven CTAs - sought alternate sources from GBD | Global Burden of Disease Collaborative Network. Global Burden of Disease Study 2019 (GBD 2019) Human Resources for Health 1990-2019. Seattle, United States of America: Institute for Health Metrics and Evaluation (IHME), 2022. | |
| | **Pharmacists per 1000 people** | WHO Global Data Observatory | 1. Linear interpolation to fill in missing years where data is available for at least two years in WHO data (*zoo* package) | The National health Workforce Accounts database, World Health Organization, Geneva; accessed 05/July/2024 (https://apps.who.int/nhwaportal, https://www.who.int/activities/improving-health-workforce-data-and-evidence) | |

| Covariate type | Variable | Primary Source | Dealing with missing data | References | Notes |
|---|---|---|---|---|---|
| | | Global Burden of Disease | 2. Missing in World Bank and WHO for 17 CTAs, sought alternate sources from GBD | Global Burden of Disease Collaborative Network. Global Burden of Disease Study 2019 (GBD 2019) Human Resources for Health 1990-2019. Seattle, United States of America: Institute for Health Metrics and Evaluation (IHME), 2022. | |
| Healthcare access | UHC Index | World Bank | 1. Data was available for 1998-2023 - used linear interpolation using *zoo* package in R where there were at least 2 years of data in the dataset | World Bank Data Bank: World Development Indicators; UHC Service Coverage Index; accessed 08/July/2024 | |
| | | Global Burden of Disease | 2. GBD UHC coverage index used where the CTA didn't have any data available in World Bank/still missing after interpolation | Global Burden of Disease Collaborative Network. Global Burden of Disease Study 2017 (GBD 2017) Health-related Sustainable Development Goals (SDG) Indicators 1990-2030. Seattle, United States of America: Institute for Health Metrics and Evaluation (IHME), 2018. | |
| | Population weighted time to travel to healthcare facilities | Literature - Malaria Atlas Project - Accessibility to Healthcare | None | Weiss, D. J., et al. (2020). "Global maps of travel time to healthcare facilities." Nature Medicine 26(12): 1835-1838.<br><br>Raster file for motorized travel time to healthcare downloaded on 05 July 2024: https://malariaatlas.org/project-resources/accessibility-to-healthcare/ | |
| | Healthcare Access & Quality Index (HAQI) | Global Burden of Disease | | Haakenstad A, Yearwood JA, Fullman N, Bintz C, Bienhoff K, Weaver MR, et al.. Assessing performance of the Healthcare Access and Quality Index, overall and by select age groups, for 204 countries and territories, 1990–2019: a systematic analysis from the Global Burden of Disease Study 2019. The Lancet Global Health. 2022;10(12):e1715–43; Data accessed on 25 September 2025 from https://doi.org/10.6069/97EM-P280 | |
| Infection prevention | PCV vaccination coverage proportion | WHO Immunization data | Where coverage wasn't available, checked whether the vaccine had been universally introduced (https://view-hub.org/vaccine/pcv?set=current-vaccine-intro-status&group=vaccine-introduction&category=pcv) (accessed 06 july 2024); if it had not been introduced by 2019 or no decision made, assumed coverage was 0 | https://immunizationdata.who.int/global/wiise-detail-page/pneumococcal-vaccination-coverage; accessed 30 January 2024 | Used the coverage of the third dose percentage using WUENIC estimates; assume coverage as 0 prior to universal introduction however this may underestimate true coverage if there is a large private market use |
| | | Global Burden of Disease | Where introduction indicated it had been introduced but there was no estimate for coverage or no data at all from WUENIC, sought alternate data for coverage estimate in GBD | Global Burden of Disease Collaborative Network. Global Burden of Disease Study 2020, Release 1 (GBD 2020 R1) Routine Childhood Vaccination Coverage 1980-2019. Seattle, United States of America: Institute for Health Metrics and Evaluation (IHME), 2021.; accessed 13 August 2024 | |
| | Rotavirus vaccination coverage proportion | WHO Immunization data | Where coverage wasn't available, checked whether the vaccine had been universally introduced on VIEW-hub (https://view-hub.org/vaccine/rota?set=vaccine-introduction-over-time&group=vaccine-introduction&category=rv) (accessed 06 july 2024); if it had not been introduced by 2019 or no decision made, assumed coverage was 0 | https://immunizationdata.who.int/global/wiise-detail-page/rotavirus-vaccination-coverage; accessed 30 January 2024 | Used the coverage of the final dose percentage using WUENIC estimates; assume coverage as 0 prior to universal introduction however this may underestimate true coverage if there is a large private market use |
| | | Global Burden of Disease | Where introduction indicated it had been introduced but there was no estimate for coverage or no data at all from WUENIC, sought alternate data for coverage estimate in GBD | Global Burden of Disease Collaborative Network. Global Burden of Disease Study 2020, Release 1 (GBD 2020 R1) Routine Childhood Vaccination Coverage 1980-2019. Seattle, United States of America: Institute for Health Metrics and Evaluation (IHME), 2021.; accessed 13 August 2024 | |

| Covariate type | Variable | Primary Source | Dealing with missing data | References | Notes |
|---|---|---|---|---|---|
| | **Proportion of population with access to safe sanitation** | World Bank | Data missing for 66 CTAs and no option to interpolate for the missing years. We use multiple imputation using chained equations (mice) to impute the data, restricting the imputations to be below the percentage of population with access to basic sanitation services (World Bank) where that data is available | World Bank Data Bank: World Development Indicators; People Using safely managed sanitation services (% of population); accessed on 08 July 2024 | |
| **Comorbidities and other clinical risk factors** | **Proportion of population living with 1 or more comorbdities** | CMMID modelled estimates | Missing in 7 CTAs | Clark, A., et al. (2020). "Global, regional, and national estimates of the population at increased risk of severe COVID-19 due to underlying health conditions in 2020: a modelling study." The Lancet Global Health 8(8): e1003-e1017. | Missing for Andorra (AND), American Samoa (ASM), Bermuda (BMU), Dominica (DMA), Greenland (GRL), Marshall Islands (MHL), Northern Mariana Islands (MNP) |
| | **Malaria incidence** | Global Burden of Disease | None | Institute for Health Metrics and Evaluation (IHME). GBD Results 2021. Seattle, WA: IHME, University of Washington, 2024. Available from https://vizhub.healthdata.org/gbd-results/. (Accessed 05 July 2024) | |
| | **Protein energy malnutrition prevalence** | Global Burden of Disease | None | Institute for Health Metrics and Evaluation (IHME). GBD Results 2021. Seattle, WA: IHME, University of Washington, 2024. Available from https://vizhub.healthdata.org/gbd-results/. (Accessed 05 July 2024) | |
| **Infection incidence** | **Lower respiratory infection incidence** | Global Burden of Disease | None | Institute for Health Metrics and Evaluation (IHME). GBD Results 2021. Seattle, WA: IHME, University of Washington, 2024. Available from https://vizhub.healthdata.org/gbd-results/. (Accessed 05 July 2024) | |
| | **Upper respiratory infection incidence** | | None | | |
| | **Otitis media incidence** | | None | | |
| | **Gonorrhoea incidence** | | None | | |
| | **Chlamydia incidence** | | None | | |
| | **Syphilis incidence** | | None | | |
| | **Trichomoniasis incidence** | | None | | |
| | **Cellulitis incidence** | | None | | |
| | **Burns and wounds from road traffic accidents and fires incidence** | | None | | |
| | **Dysentery incidence (*Shigella* spp., non-typhoidal *Salmonella* spp., Enterotoxigenic *E. coli*, *Campylobacter* spp.)** | | None; derived using aetiology-specific data as incidence by aetiology is not available | | |

| Covariate type | Variable | Primary Source | Dealing with missing data | References | Notes |
|---|---|---|---|---|---|
| | **Diarrhoea incidence** | | None | | |
| | **Typhoid incidence** | | None | | |
| | **Cholera incidence** | | None | | |
| | **UTI incidence** | | None | | |
| | **MDR and XDR TB incidence** | | None | | |
| | **Sepsis incidence** | | Data available for 194 CTAs; used the estimates for 2017 and assumed incidence similar for 2019 | Rudd, K. E., et al. (2020). "Global, regional, and national sepsis incidence and mortality, 1990–2017: analysis for the Global Burden of Disease Study." The Lancet 395(10219): 200-211. | |
| | **HAP incidence** | Global PPS & Global Burden of Disease | Data available for 194 CTAs where sepsis estimates are available | Estimates of HAP cases in European countries (Table 22, page 103) of: ECDC (2024). Point prevalence survey of healthcare-associated infections and antimicrobial use in European acute care hospitals. Stockholm. | |
| | **IAI incidence** | Global PPS & Global Burden of Disease | Data available for 194 CTAs where sepsis estimates are available | Case mix of hospital infections (supplemental table 2): Versporten, A., et al. (2018). "Antimicrobial consumption and resistance in adult hospital inpatients in 53 countries: results of an internet-based global point prevalence survey." The Lancet Global Health 6(6): e619-e629. Sepsis estimates: Rudd, K. E., et al. (2020). "Global, regional, and national sepsis incidence and mortality, 1990–2017: analysis for the Global Burden of Disease Study." The Lancet 395(10219): 200-211. | |

| Covariate type | Variable | Primary Source | Dealing with missing data | References | Notes |
|---|---|---|---|---|---|
| | **Necrotising fasciitis incidence** | Global PPS & Global Burden of Disease | Data available for 194 CTAs where sepsis estimates are available | Case mix of hospital infections (supplemental table 2): Versporten, A., et al. (2018). "Antimicrobial consumption and resistance in adult hospital inpatients in 53 countries: results of an internet-based global point prevalence survey." The Lancet Global Health 6(6): e619-e629.<br>Estimates of necrotising fasciitis out of hospital SSTI: Gomes Siqueira, G. L., et al. (2020). "Non-necrotizing and necrotizing soft tissue infections in South America: A retrospective cohort study." Annals of Medicine and Surgery 59: 24-30. AND Thean, L. J., et al. (2020). "Hospital admissions for skin and soft tissue infections in a population with endemic scabies: A prospective study in Fiji, 2018–2019." PLOS Neglected Tropical Diseases 14(12): e0008887.<br>Sepsis estimates: Rudd, K. E., et al. (2020). "Global, regional, and national sepsis incidence and mortality, 1990–2017: analysis for the Global Burden of Disease Study." The Lancet 395(10219): 200-211. | |
| **Antibiotic resistant infection incidence** | **ESBL sepsis incidence**<br>**Carbapenem-resistant Acinetobacter baumannii sepsis incidence**<br>**CR-Pseudomonas aeruginosa sepsis incidence**<br>**CR-Enterobacterales sepsis incidence**<br>**MRSA sepsis incidence**<br>**Vancomycin resistant S. aureus sepsis incidence**<br>**Vancomycin resistant Enterococcus faecium sepsis incidence** | Global Research on Antimicrobial Resistance (GRAM) and Global Burden of Disease | Data available for 194 CTAs where sepsis estimates are available | Sepsis estimates from: Rudd, K. E., et al. (2020). "Global, regional, and national sepsis incidence and mortality, 1990–2017: analysis for the Global Burden of Disease Study." The Lancet 395(10219): 200-211.<br>Resistance estimates: Murray, C. J. L., et al. (2022). "Global burden of bacterial antimicrobial resistance in 2019: a systematic analysis." The Lancet 399(10325): 629-655. (data from unpublished GRAM modelling) | |

##### 1.1.5 Missing data imputation

Given our analysis focuses on national-level antibiotic use, we imputed missing data for the hospital sector for 26/72 CTAs in IQVIA MIDAS and for CTAs missing key covariates in 2019. We used *mice* package in R for multiple imputation using predictive mean matching method for hospital AWaRe antibiotic DID. We used passive imputation for the CTA-level totals (summed values of hospital and retail sectors) for total Access, total Watch and total Reserve use and overall total DID. We also imputed proportion of the population with access to safe sanitation for 66 CTAs and proportion of the population living below the $2.15 poverty line where missing for 40 CTAs. Predictive mean matching is a robust imputation method that uses a regression model to predict values for the missing data; it uses the observed values in the data to find "donor" values from the data to use to replace the missing value. This retains the distribution of the data and draws from realistic values. Passive imputation imputes the totals as sums of the separately imputed sector data to retain the relationships between the variables rather than imputing each variable separately. Variables selected for use in the imputation model focused on those used in the main analysis; where covariates were correlated with each other above 0.8 (absolute value), we retained only one of the covariate pair by priority of use in the main analysis with top priority given to infection and resistance covariates. We imputed data for all 186 CTAs to maintain the relationship between covariates and antibiotic use in the imputation model, however, only retained the antibiotic use estimates for CTAs in IQVIA MIDAS dataset and retained imputed poverty and sanitation for all CTAs.

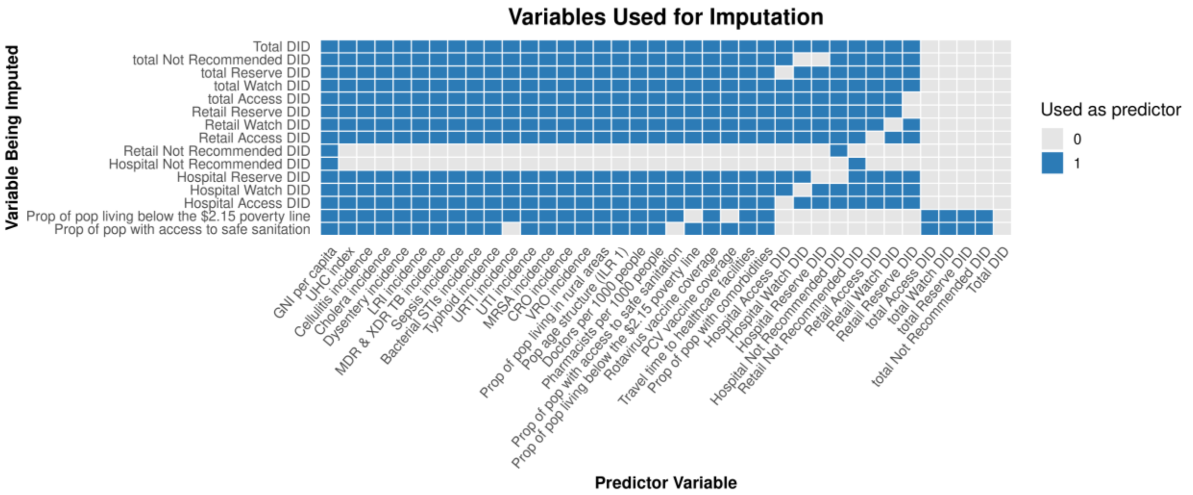

*Figure 2. Imputation matrix showing the variables used as predictors for imputation of each antibiotic and covariate of interest for the full MIDAS® dataset which includes Not Recommended antibiotics*

##### 1.1.6 Data processing post imputation

Where an imputed value for proportion of population with access to safe sanitation was greater than the proportion with access to basic sanitation (available in 58/66 CTAs from World Bank Development Indicators), we replaced the imputed value with the proportion with access to basic sanitation; for Taiwan we assumed access to safe sanitation was comparable to other HICs and all imputed values were replaced with 0.99. For computational reasons in downstream analyses, we also took the median of the imputations for safe sanitation and proportion living below the $2.15 poverty line.

##### 1.1.7 Re-classifying Not Recommended antibiotics to AWaRe Category

The WHO AWaRe system includes a list of "Not Recommended" antibiotics which are predominantly fixed dose combinations of two or more antibiotics or other products. We assume that expected "Not recommended" antibiotic use should be zero in all CTAs, therefore we did not include that as a category in our model. However, we assume that this total antibiotic use in CTAs where Not Recommended antibiotics are reported in MIDAS® is still needed, therefore we re-classified these antibiotics into a relevant AWaRe category as if they were replaced with a similar antibiotic from the Access, Watch or Reserve categories.

To re-classify Not Recommended antibiotics into their relevant AWaRe category (Access, Watch or Reserve), we mapped them to the highest AWaRe category of the antibiotics in the antibiotic combination (where Access < Watch < Reserve). For example, the Not Recommended combination "Ampicillin+Cloxacillin" maps to Access since both antibiotics are Access, or "Amoxicillin+Clarithromycin+Omeprazole" maps to a derived category of Watch given the Clarithromycin is a Watch antibiotic. After re-classifying these antibiotics, we recalculated the total DID by AWaRe and repeated the missing data imputation by AWaRe and hospital and retail sector when using these updated volumes through the same process described in supplemental methods section 1.1.5 and supplemental Figure 3.

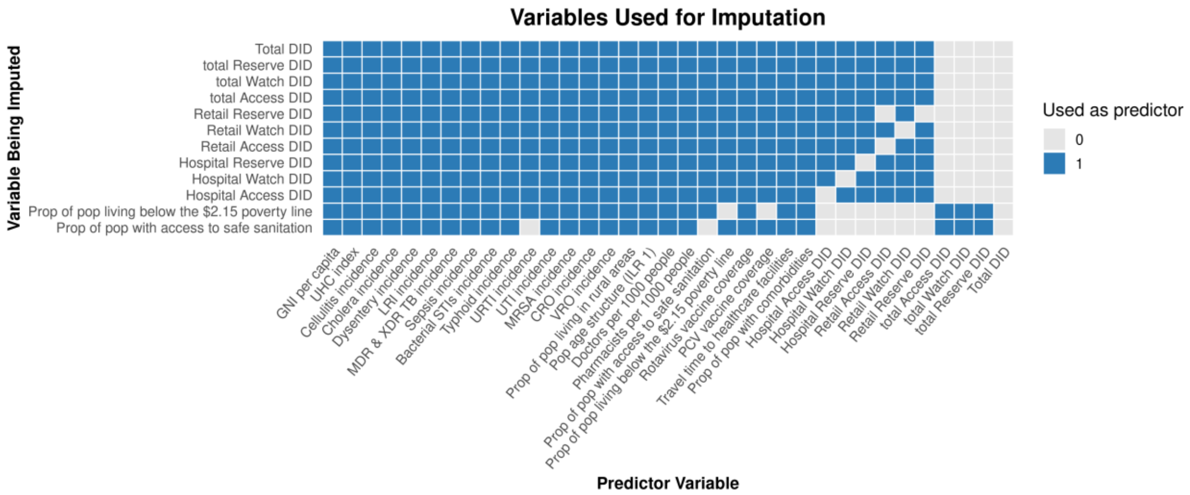

*Figure 3. Imputation matrix showing the variables used as predictors for imputation of each antibiotic and covariate of interest for the MIDAS® dataset with the re-classified Not Recommended antibiotics*

#### 1.2 Statistical Analysis

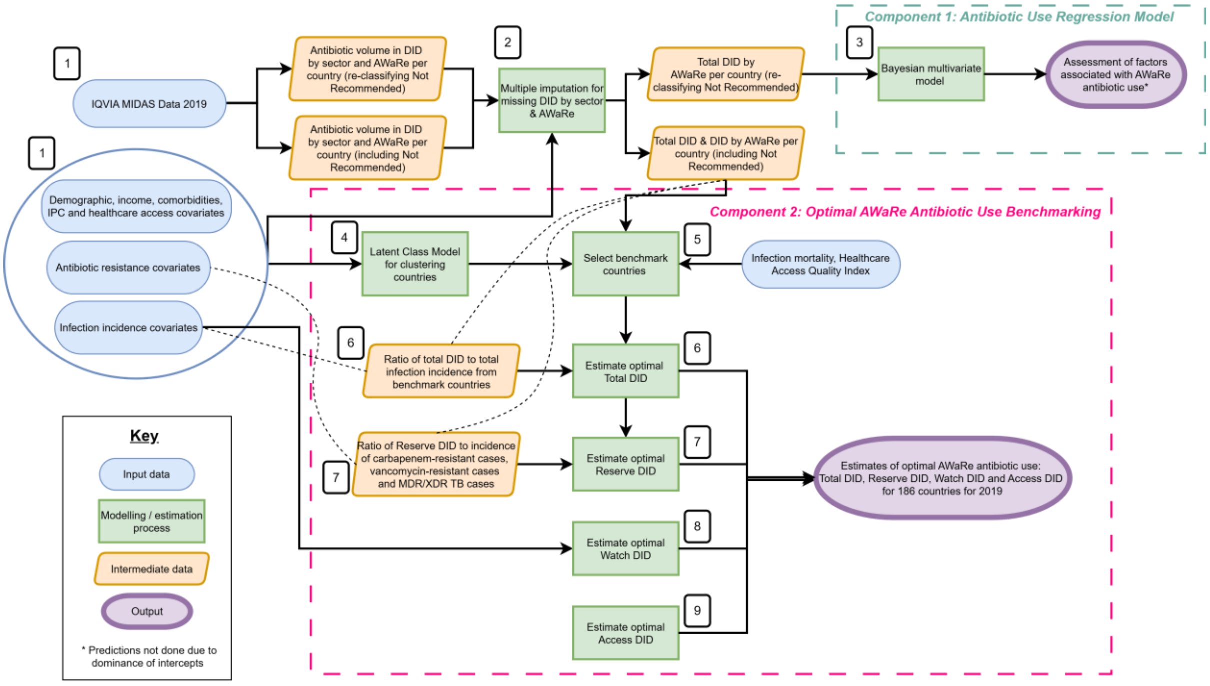

*Figure 4. Study schema for deriving estimates of expected AWaRe antibiotic use based on infection burden, antibiotic resistance, socio-demographics, healthcare infrastructure and access using the WHO AWaRe book as a clinical framework. This figure is also presented in the main manuscript but is included here for ease. 1. Data preparation, 2. Missing data imputation, 3. Antibiotic use regression model, 4. Latent Class Model, 5. Benchmark CTA selection, 6. Estimate optimal total DID, 7. Estimate optimal Reserve DID, 8, Estimate optimal Watch DID, 9. Estimate optimal Access DID*

##### 1.2.1 Antibiotic use Regression model

We applied a Bayesian multivariate ecological regression model (Figure 4 Component 1) using CTA-level data to quantify how factors plausibly associated with antibiotic need such as infectious disease burden, ABR and other covariates () relate to antibiotic use volume across AWaRe categories.

We fit the model separately to 1000 imputed datasets, imputed after re-classifying "Not recommended" antibiotics to their relevant AWaRe category (see section 1.1.7). We used DDD/1000 inhabitants/year as the outcome instead of DID to avoid having to model values very close to zero for Reserve DID (<0.5 DID in most CTAs). To avoid that the model was fit on data from CTAs where antibiotic use may have unsafe low levels of antibiotics or implausibly low reported antibiotic use levels, we excluded CTAs where the majority (>50%) of imputed total DIDs were below 9.7 DID, the lowest value observed in a high-income CTA (the Netherlands). Five CTAs (China, Indonesia, Malaysia, Philippines, and Venezuela) were excluded on this basis leaving 67/72 CTAs for model fitting.

Covariates were defined a priori based on a review of the literature with model covariates for Access, Watch and Reserve use based on factors that have been documented in literature and clinical / structural plausibility to affect AWaRe antibiotic use. Covariates for each AWaRe antibiotic volume were specified separately to account for specific clinical and resistance variables that may be relevant to that AWaRe category specifically based on the literature and the AWaRe Book (Table 2). For example, incidence of carbapenem resistant infections were only included in the Reserve model, while sepsis incidence was included in both the Watch and Reserve models based on AWaRe Book & other treatment recommendations. Proportion of population of different age groups were treated as compositional variables for modelling as they contain relative information and the different parts sum to a whole. These types of variables need to be transformed for inclusion in most models because they violate the assumptions of independence (e.g. when the proportion of one age group decreases, another age group proportion increases). We consider these to be Aitchison compositions and use an isometric log-ratio transformation (R package: *compositions*[44]) to transform the variables to be used in modelling.[45-47] We excluded any variables in the regression model that were correlated with each other higher than 0.8 and prioritised retaining infection and resistance data with higher disease burdens or antibiotic prescribing requirements. We used the point estimates of incidence per 100,000 people for all included GBD infection and resistance estimates.

We fit a Bayesian multivariate model with a gamma distribution using a log-link function. We used normal priors for the intercepts (Access, Watch and Reserve) deriving mean and standard deviation from the Browne et al. estimates of antibiotic use in 2018;[48] these were then log transformed given the model uses a log-link function. For the coefficients, we used horseshoe priors with a parameter ratio of 0.8 for the covariates. Horseshoe priors allow for variable selection and shrinkage of less relevant parameters. A parameter ratio of 0.8 assumes 80% of the predictors are relevant, which puts less shrinkage in the model than a lower parameter ratio.

*Table 2. Covariates included per AWaRe antibiotic category for the multivariate model and priors on intercepts and covariates.*

| Model outcome | Covariates included | Priors on intercept | Priors on covariates |
|---|---|---|---|
| Access DI | GNI per capita, population age group ratio 1*, UHC coverage, population weighted travel time to healthcare facilities, PCV vaccine coverage, rotavirus vaccine coverage, rural population proportion, proportion of population with 1+ comorbidity, URI incidence, LRI incidence, typhoid incidence, dysentery incidence, UTI incidence, burns & wounds incidences, cellulitis incidence, bacterial STI incidence, MRSA incidence and ESBL incidence | Normal(8.21, 1.8) | Horseshoe priors with parameter ratio of 0.8 |
| Watch DI | GNI per capita, population age group ratio 1*, UHC coverage, population weighted travel time to healthcare facilities, PCV vaccine coverage, rotavirus vaccine coverage, rural population proportion, proportion of population with 1+ comorbidity, LRI incidence, typhoid incidence, dysentery incidence, burns & wounds incidence, cellulitis incidence, sepsis incidence, UTI incidence, MDR/XDR TB incidence, MRSA incidence, ESBL incidence | Normal(7.78, 1.61) | Horseshoe priors with parameter ratio of 0.8 |
| Reserve DI | GNI per capita, population age group ratio 1*, UHC coverage, rural population proportion, proportion of population with 1+ comorbidity, LRI incidence, burns & wounds incidence, cellulitis incidence, typhoid incidence, sepsis incidence, MDR/XDR TB incidence, ESBL incidence, CRO incidence, VRO incidence | Normal(2.49, 0.95) | Horseshoe priors with parameter ratio of 0.8 |

*DI = DDD/1000 Inhabitants/Year; converted from DID (DDD/1000 inhabitants/day) to shift small values away from zero; *population age group ratio 1 is an isometric log ratio of the relationship between the population age group proportions after transformation; only on of the four resulting ratios were retained for the model due to correlation.*

We initially planned to use the fitted model to predict expected optimal national volumes of Access, Watch, and Reserve antibiotic use for all 186 CTAs with available covariate data given each CTA's covariate profile. In this context, optimal antibiotic use refers to model-derived estimates based on the observed associations across all CTAs in the dataset. However, after reviewing the estimated model coefficients, we found that the posterior distributions were dominated by the intercepts, suggesting that the model captured overall antibiotic volume rather than variation explained by CTA-level covariates. We therefore did not use this model to generate predicted value of AWaRe antibiotic use (see Results section 2.1).

##### 1.2.2 Optimal AWaRe Antibiotic Use Benchmarking

###### *1.2.2.1 Overview*

We developed an alternate benchmarking methodology for estimating expected optimal AWaRe antibiotic need. In brief, 186 CTAs with complete covariates were clustered into comparable "peer" clusters using latent class modelling, then within each cluster, a benchmark CTA was identified based on minimising total antibiotic use and infection mortality and high Access use percentage. These benchmark CTAs were then used to estimate optimal total DID antibiotic need for other CTAs in the same cluster. We consider the benchmark CTA as an "optimal" antibiotic prescriber and this approach estimates optimal antibiotic need as though the infection burden for each CTA within a cluster is treated according to the prescribing practices of the benchmark CTA in that cluster (Figure 4:Component 2). Watch and Reserve DID are estimated given relevant infection and ABR burdens and Access is considered residual antibiotics after accounting for Watch and Reserve need.

###### *1.2.2.2 Defining peer groups – clustering analysis*

Commonly World Bank income group or WHO region are used to group CTAs to describe patterns of antibiotic use; [(49-51)] however, there may still be differences in other factors such as infection incidence, antibiotic resistance or healthcare infrastructure (Figure 5 and Figure 6) within an income group or WHO region that may make these sub-optimal grouping for benchmarking.

To appropriately identify peer groups for comparison of antibiotic use, we used a Latent Class Model (LCM) and all considered covariates (n=40) across the factors that may influence antibiotic use to group the 186 analysis CTAs into clusters. Covariates were normalised prior to inclusion in LCM. LCM was performed using the R package *VarSelLCM.*[(52,53)] This implementation of LCM was used because it allows for variable selection using the integrated complete-data likelihood, computes discriminative power of the variables and allows model comparison of different number of groups. We trialled different number of clusters between 1 and 6 groups, noting that current common methods to group CTAs include World Bank income group (4 clusters) and WHO regions (6 clusters). We used BIC for variable selection and for selection of optimal number of groups to avoid overfitting and prioritise a less complex model. See below for model fitting statistics (Table 3,Figure 7). We ordered clusters by mean GNI per capita within a cluster with cluster 1 being lowest income CTAs and cluster 4 being highest income CTAs.

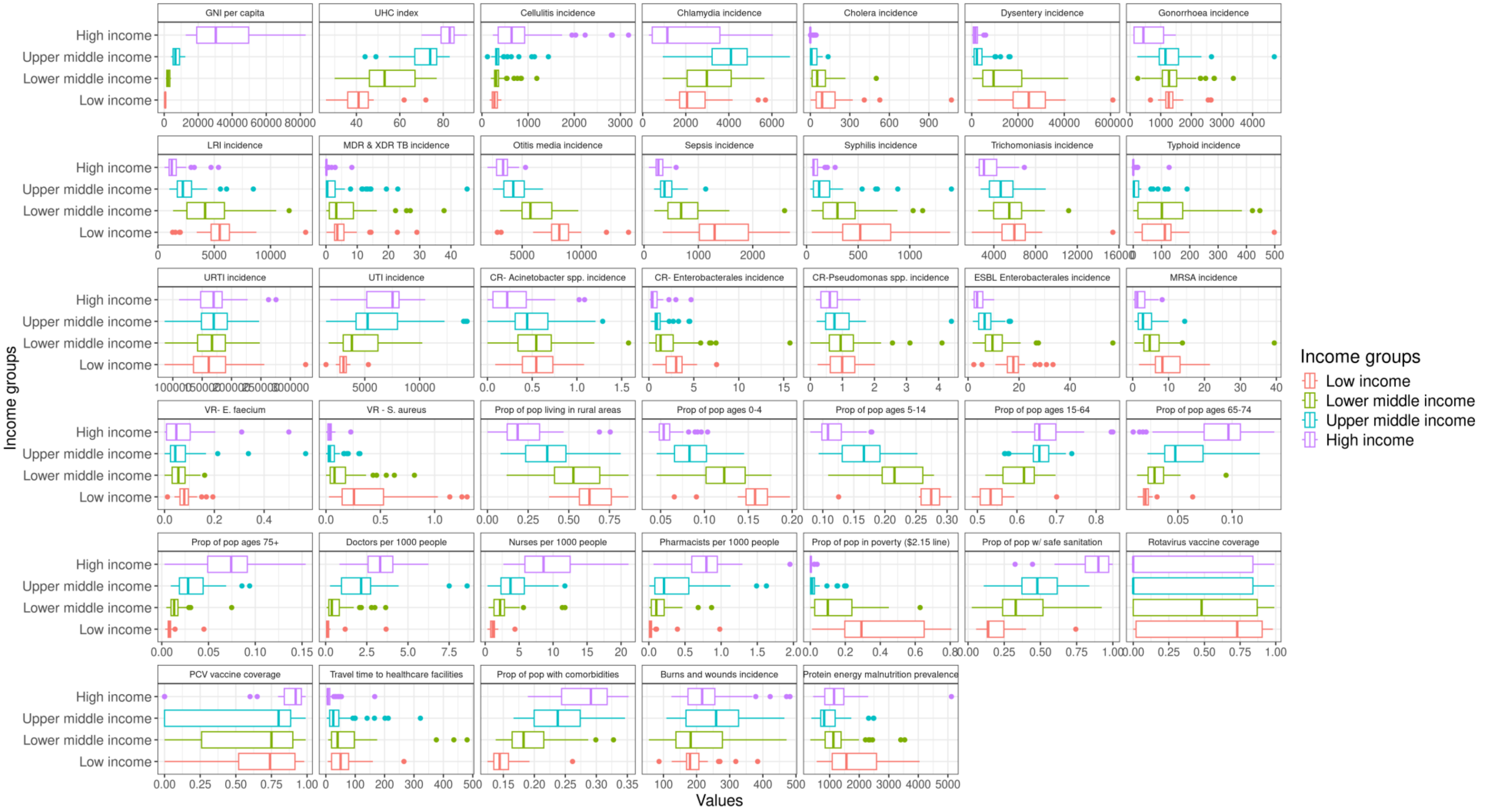

*Figure 5. Distribution of covariates by World Bank Income Group. GNI = gross national income, UHC = universal health coverage, LRI = lower respiratory tract infection, MDR & XDR TB = multi-drug resistant and extremely-drug resistant tuberculosis, URTI = upper respiratory tract infection, UTI = urinary tract infection, CR = carbapenem resistant, ESBL = extended-spectrum beta-lactamase, MRSA = methicillin-resistant Staphylococcus aureus, VR. E faecium = vancomycin-resistant Enterococcus faecium, VR-S.aureus = vancomycin-resistant Staphylococcus aureus, Prop = proportion, pop = population, PCV = pneumococcal conjugate vaccine*

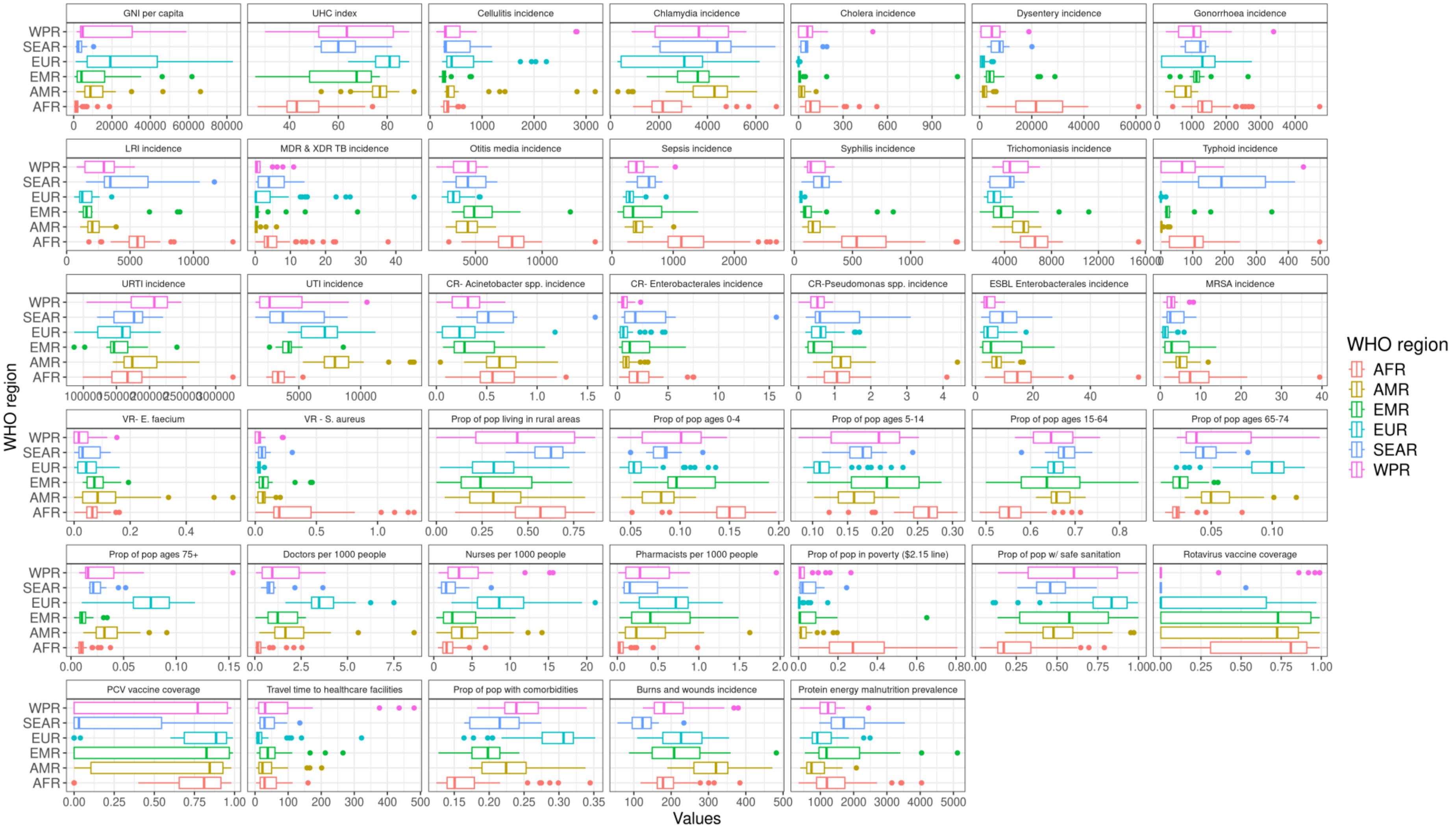

*Figure 6. Distribution of covariates by WHO Region. GNI = gross national income, UHC = universal health coverage, LRI = lower respiratory tract infection, MDR & XDR TB = multi-drug resistant and extremely-drug resistant tuberculosis, URTI = upper respiratory tract infection, UTI = urinary tract infection, CR = carbapenem resistant, ESBL = extended-spectrum beta-lactamase, MRSA = methicillin-resistant Staphylococcus aureus, VR. E faecium = vancomycin-resistant Enterococcus faecium, VR-S.aureus = vancomycin-resistant Staphylococcus aureus, Prop = proportion, pop = population, PCV = pneumococcal conjugate vaccine*

*Table 3. Latent class model fitting showing variables not selected by the model comparing 1 to 6 clusters.*

| | | Latent Class Model fitting summary |
|---|---|---|
| | **Number of variables included in model** | 40 |
| **1 cluster** | **BIC** | -10745.9 |
| | **Variables not used by the model** | 0 |
| **2 clusters** | **BIC** | -7857.8 |
| | **Variables not used by the model** | Rotavirus vaccination, PCV vaccination, URI incidence, MDR&XDR TB incidence |
| **3 clusters** | **BIC** | -6953.5 |
| | **Variables not used by the model** | Rotavirus vaccination, PCV vaccination, URI incidence, MDR&XDR TB incidence |
| **4 clusters** | **BIC** | -6465.4 |
| | **Variables not used by the model** | Rotavirus vaccination, PCV vaccination, URI incidence |
| **5 clusters** | **BIC** | -6265.0 |
| | **Variables not used by the model** | Rotavirus vaccination, URI incidence |
| **6 clusters** | **BIC** | -6095.7 |
| | **Variables not used by the model** | Rotavirus vaccination, PCV vaccination, URI incidence |

*BIC = Bayesian Information Criterion, MDR & XDR TB = multi-drug resistant and extremely-drug resistant tuberculosis, URTI = upper respiratory tract infection, PCV = pneumococcal conjugate vaccine*

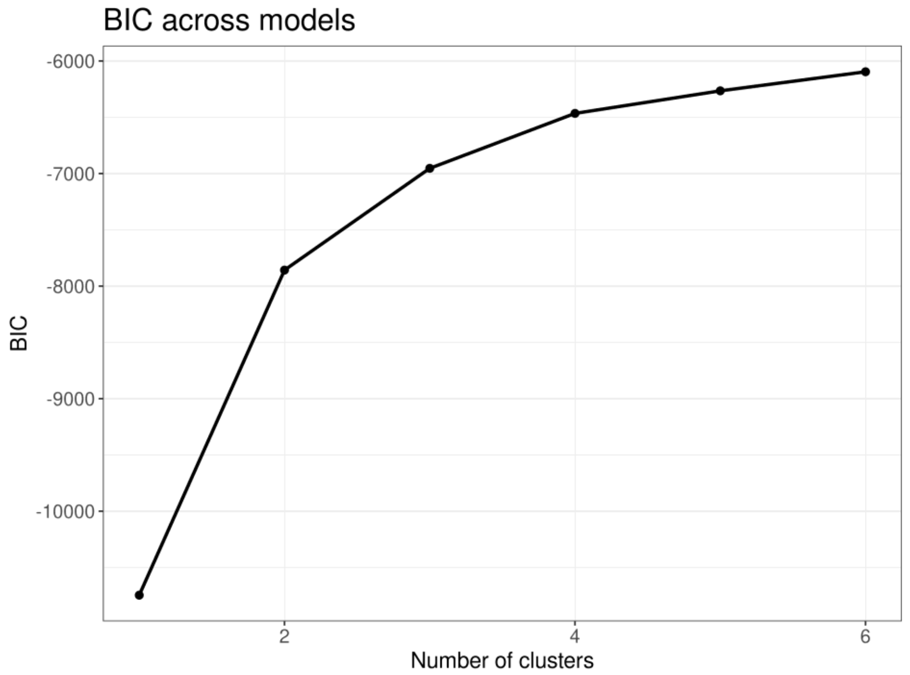

*Figure 7. Bayesian information criterion (BIC) of latent class models considering 1 to 6 clusters using available covariates (n=40) for 186 analysis CTAs.*

***Benchmark CTA selection***

Within each cluster, CTAs with total and AWaRe antibiotic use data in IQVIA MIDAS® available after imputation were identified; we excluded the 5 CTAs with total DID less than 9.7 DID as for the regression analysis. We compared total DID of antibiotics to infection mortality per 100,000 people and percentage of Access antibiotic use within each cluster. Supplemental Figure 8 shows infection incidence against infection mortality for all 186 CTAs with infection burden data with the IQVIA MIDAS CTAs highlighted.

In selecting benchmark CTAs, we aimed to minimise total antibiotic use and infection mortality while maximising percentage of Access use and aiming to select a benchmark CTA that used minimal DID of Not Recommended antibiotics (Figure 9). We first identified countries with low DID and low infection mortality then considered their percentage of Access. Given the previous WHO CTA-level target of Access antibiotic use was 60% of total use,[54] a benchmark CTA needed to meet a minimum of 60% Access to be selected, though many CTAs met a higher target. Where possible we then tried to select a CTA with low or zero Not Recommended antibiotic use. One benchmark CTA was selected for each cluster. Where there were several potential candidates based on the above criteria, we aimed to select a benchmark CTA in each cluster that was a similar or lower range of Healthcare Access and Quality Index[55] (HAQI) to the other CTAs in the cluster even if that meant antibiotic use was not the lowest in the cluster. This was done to ensure that the benchmark CTA more closely represented the CTAs it was going to be a benchmark for.

Where potential benchmark CTAs in a cluster had low antibiotic use but had higher infection mortality rates than the CTAs in the next cluster with similar antibiotic use levels (i.e. from Cluster 1 to 2, 2 to 3, 3 to 4, moving from lowest income CTAs to highest income CTAs), we considered that those CTAs would be inappropriate benchmarks as they may have possible antibiotic access issues (e.g. low antibiotic use but still higher mortality which could illustrate an access issue). Where there is no suitable benchmark CTA within a cluster, we benchmark to the CTA from the next cluster. However, we note that this represents a minimum estimated range of antibiotic use required for CTAs in that cluster given the higher infection mortality and lower HAQI in the lower clusters where more antibiotics than estimated are likely needed to treat the infection burden.

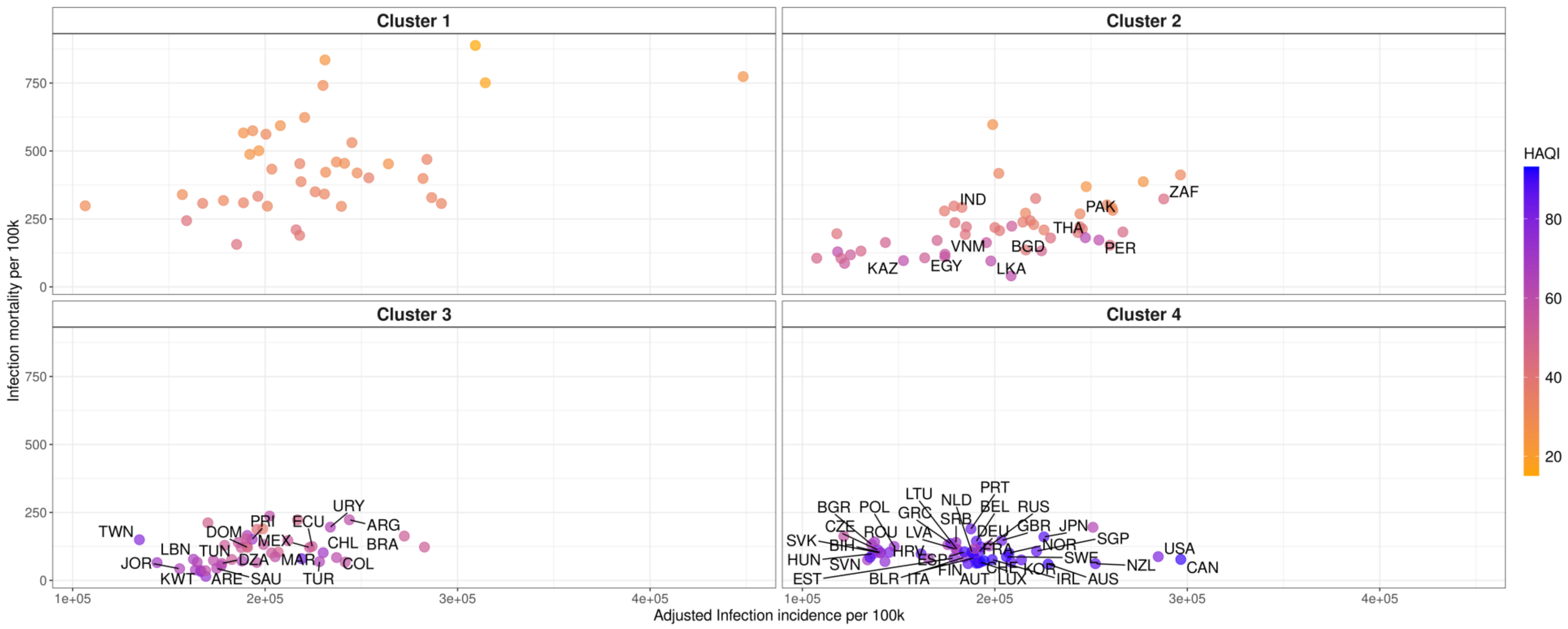

*Figure 8. Point estimates of infection incidence vs. infection mortality by benchmark group and coloured by Healthcare Access and Quality Index (HAQI) showing the MIDAS® CTAs with available antibiotic use data available.*

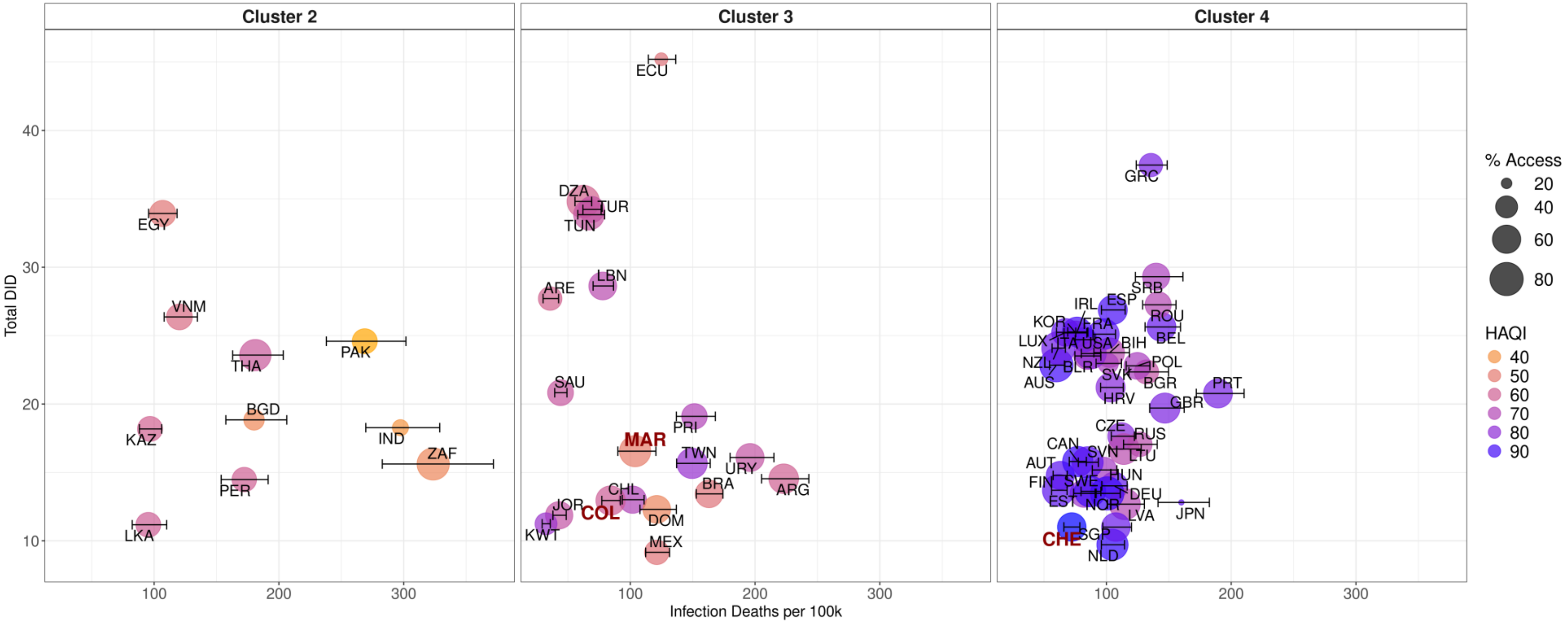

*Figure 9. MIDAS® CTAs with available antibiotic use data by benchmark group (note: no benchmark group 1 CTAs had antibiotic use data available in MIDAS®) showing infection mortality per 100,000 people against total DID sized by % Access antibiotics and coloured by Healthcare Access and Quality Index (HAQI). Uncertainty bars are for infection mortality. Source: Total DID based on IQVIA MIDAS® data for 2019, reflecting estimates of real-world activity. Copyright IQVIA. All Rights Reserved*

*Table 4. Criteria used to select benchmark CTAs within each cluster for which IQVIA MIDAS data is available. For CTAs in IQVIA MIDAS where only retail sector was available, data were imputed for the missing sector and therefore Total DID, percent Access and Not Recommended DID are shown with their 95% CI. Infection deaths and infection incidence are the sum of infections of interest available in GBD.*

| CTA Name | ISO3 Code | Cluster | Income group | Sectors available in IQVIA MIDAS* | Total DID (95%CI)* | Infection deaths per 100K (95%CI) | Percent Access use* | Not Recommended DID (95%CI)* | Infection incidence per 100k (95%CI) |
|---|---|---|---|---|---|---|---|---|---|
| **Sri Lanka** | LKA | Cluster 2 | Upper middle income | Retail Only | 11.2 (10.1, 12.1) | 95.4 (82.5, 110.1) | 48 (43.3, 52.2) | 0 (0, 0) | 197939.2 (176202.8, 219449.9) |
| **Peru** | PER | Cluster 2 | Upper middle income | Retail Only | 14.5 (13.2, 16) | 172.2 (153.7, 191.4) | 47.1 (44.2, 50) | 2.2 (2.1, 2.2) | 254064.1 (227135.6, 283122) |
| **South Africa** | ZAF | Cluster 2 | Upper middle income | Hospital and Retail | 15.6 (15.6, 15.6) | 323.6 (282.8, 371.9) | 75.6 (75.6, 75.6) | 0.2 (0.2, 0.2) | 287747.6 (258798.2, 318324.1) |
| **Kazakhstan** | KAZ | Cluster 2 | Upper middle income | Hospital and Retail | 18.2 (18.2, 18.2) | 96.7 (88.2, 106.1) | 48.8 (48.8, 48.8) | 0.4 (0.4, 0.4) | 152525.3 (136912.2, 170662.7) |
| **India** | IND | Cluster 2 | Lower middle income | Retail Only | 18.3 (16, 21) | 297.4 (269.6, 328.9) | 27.1 (23.8, 37.8) | 3.4 (3.3, 3.4) | 178812.4 (161216.6, 196578.9) |
| **Bangladesh** | BGD | Cluster 2 | Lower middle income | Retail Only | 18.8 (16.8, 19.9) | 180.1 (157.5, 206.3) | 36.2 (32.8, 38.5) | 1.7 (1.7, 1.8) | 228939.2 (206622.5, 254310.9) |
| **Thailand** | THA | Cluster 2 | Upper middle income | Hospital and Retail | 23.6 (23.6, 23.6) | 181.1 (163, 203.5) | 72.2 (72.2, 72.2) | 0.1 (0.1, 0.1) | 247037.4 (223428.6, 275012.1) |
| **Pakistan** | PAK | Cluster 2 | Lower middle income | Retail Only | 24.6 (22, 27.4) | 268.8 (238, 301.8) | 48.9 (45.9, 53.7) | 0.9 (0.9, 0.9) | 244241.9 (221108.6, 272332) |
| **Viet Nam** | VNM | Cluster 2 | Lower middle income | Hospital and Retail | 26.4 (26.4, 26.4) | 120.3 (108.1, 134.7) | 51.3 (51.3, 51.3) | 0.4 (0.4, 0.4) | 174083.2 (154333, 198498.1) |
| **Egypt, Arab Rep.** | EGY | Cluster 2 | Lower middle income | Hospital and Retail | 33.9 (33.9, 33.9) | 106.9 (95.6, 118.4) | 52.9 (52.9, 52.9) | 6.1 (6.1, 6.1) | 163564.4 (146832.7, 182845) |
| **Mexico** | MEX | Cluster 3 | Upper middle income | Retail Only | 9.2 (7.9, 10.5) | 121.3 (112.3, 131.5) | 47.1 (40.2, 51.1) | 0.7 (0.6, 0.7) | 223039.8 (198026.4, 251299) |
| **Kuwait** | KWT | Cluster 3 | High income | Retail Only | 11.2 (10.1, 12.3) | 32.5 (29.4, 35.9) | 39.9 (36.1, 43.1) | 0.4 (0.4, 0.4) | 166596.6 (145822.8, 188933.3) |
| **Jordan** | JOR | Cluster 3 | Upper middle income | Retail Only | 11.9 (10.6, 13.7) | 43 (38, 48.7) | 56.6 (53.5, 61.2) | 0.3 (0.3, 0.3) | 155591.4 (136957.3, 176590.3) |
| **Dominican Republic** | DOM | Cluster 3 | Upper middle income | Retail Only | 12.3 (10.3, 14.7) | 121.2 (107.7, 136.9) | 58 (50.2, 63.5) | 0.9 (0.9, 1) | 190903 (170997.3, 214575.5) |
| **Colombia** | COL | Cluster 3 | Upper middle income | Retail Only | 12.9 (11.4, 14.5) | 84.2 (77.2, 92) | 63.8 (57.6, 67.4) | 0 (0, 0) | 237091.6 (212575.5, 266926.3) |
| **Chile** | CHL | Cluster 3 | High income | Retail Only | 13 (11.8, 14.5) | 101.9 (93.6, 111.1) | 56.5 (51.5, 59.2) | 0.3 (0.3, 0.3) | 230155.3 (205971.3, 257610.9) |
| **Brazil** | BRA | Cluster 3 | Upper middle income | Retail Only | 13.4 (11.8, 14.5) | 163.1 (152.9, 174.2) | 57.5 (50.7, 61.4) | 0 (0, 0) | 272313.9 (244863.2, 300855.9) |

| CTA Name | ISO3 Code | Cluster | Income group | Sectors available in IQVIA MIDAS* | Total DID (95%CI)* | Infection deaths per 100K (95%CI) | Percent Access use* | Not Recommended DID (95%CI)* | Infection incidence per 100k (95%CI) |
|---|---|---|---|---|---|---|---|---|---|
| **Argentina** | ARG | Cluster 3 | Upper middle income | Retail Only | 14.5 (13.9, 16.1) | 223 (205.2, 243.1) | 65.2 (63.4, 67.6) | 0.6 (0.6, 0.6) | 243767.1 (218516.7, 273409.6) |
| **Taiwan** | TWN | Cluster 3 | High income | Hospital and Retail | 15.7 (15.7, 15.7) | 149.5 (137.1, 164.1) | 67.9 (67.9, 67.9) | 0.2 (0.2, 0.2) | 134683.7 (121930.3, 149721.8) |
| **Uruguay** | URY | Cluster 3 | High income | Retail Only | 16.1 (15.7, 16.7) | 196 (180, 215.1) | 59.1 (58, 60.4) | 0.3 (0.3, 0.3) | 233880.6 (209173.6, 261594.9) |
| **Morocco** | MAR | Cluster 3 | Lower middle income | Retail Only | 16.6 (15.2, 18.4) | 103.9 (90.1, 120.5) | 71.3 (69, 73.3) | 1.1 (1.1, 1.2) | 206806.6 (184309.3, 232226.5) |
| **Puerto Rico** | PRI | Cluster 3 | High income | Hospital and Retail | 19.1 (19.1, 19.1) | 151.4 (136.9, 168.2) | 50.3 (50.3, 50.3) | 0 (0, 0) | 193291.2 (175469.8, 212595.9) |
| **Saudi Arabia** | SAU | Cluster 3 | High income | Hospital and Retail | 20.8 (20.8, 20.8) | 44.2 (39.4, 49.3) | 51.5 (51.5, 51.5) | 0 (0, 0) | 174821.8 (155065.9, 198814.1) |
| **United Arab Emirates** | ARE | Cluster 3 | High income | Retail Only | 27.7 (26.7, 28.6) | 35.8 (30.1, 42.6) | 44.4 (42.8, 46.1) | 0.3 (0.3, 0.3) | 169052.2 (146501.6, 193646.3) |
| **Lebanon** | LBN | Cluster 3 | Upper middle income | Retail Only | 28.6 (27.3, 30.4) | 77.9 (70.2, 86.6) | 58.2 (56.5, 59.6) | 1.4 (1.4, 1.4) | 162777.5 (144910.6, 183833.2) |
| **Tunisia** | TUN | Cluster 3 | Lower middle income | Hospital and Retail | 33.8 (33.8, 33.8) | 66.9 (58, 79.5) | 69 (69, 69) | 2.2 (2.2, 2.2) | 164720.9 (147306.9, 184867) |
| **Türkiye** | TUR | Cluster 3 | Upper middle income | Hospital and Retail | 34.2 (34.2, 34.2) | 69.1 (62.1, 76.7) | 52.2 (52.2, 52.2) | 0.8 (0.8, 0.8) | 228252.4 (203959.3, 256747.8) |
| **Algeria** | DZA | Cluster 3 | Upper middle income | Retail Only | 34.8 (33.7, 36.4) | 62.3 (55.7, 69.1) | 76.5 (75.7, 77.5) | 2.6 (2.6, 2.6) | 177522.7 (157500.6, 198704.8) |
| **Ecuador** | ECU | Cluster 3 | Upper middle income | Retail Only | 45.2 (43.4, 45.9) | 124.9 (114.5, 136.4) | 22.4 (21.8, 23.2) | 1.5 (1.5, 1.6) | 224433.5 (199522.2, 250329.2) |
| **Netherlands** | NLD | Cluster 4 | High income | Hospital and Retail | 9.7 (9.7, 9.7) | 104.9 (96, 114.5) | 71.2 (71.2, 71.2) | 0 (0, 0) | 188066.9 (171222.3, 208304.4) |
| **Singapore** | SGP | Cluster 4 | High income | Hospital and Retail | 11 (11, 11) | 107.6 (97.2, 120.1) | 61.2 (61.2, 61.2) | 0 (0, 0) | 221668.2 (196817.3, 248219.1) |
| **Switzerland** | CHE | Cluster 4 | High income | Hospital and Retail | 11 (11, 11) | 72.3 (65.9, 78.7) | 60.8 (60.8, 60.8) | 0 (0, 0) | 194355.2 (176293.3, 215214.8) |
| **Latvia** | LVA | Cluster 4 | High income | Hospital and Retail | 12.7 (12.7, 12.7) | 114.7 (100.9, 130.4) | 70.7 (70.7, 70.7) | 0 (0, 0) | 179845.2 (162656.9, 199002.6) |
| **Japan** | JPN | Cluster 4 | High income | Hospital and Retail | 12.8 (12.8, 12.8) | 160.1 (141.4, 182.5) | 17.9 (17.9, 17.9) | 0 (0, 0) | 225512.6 (204863.7, 253226.4) |
| **Estonia** | EST | Cluster 4 | High income | Retail Only | 13.5 (12.1, 14.9) | 82 (73.7, 91.8) | 58.1 (54.8, 60.7) | 0 (0, 0) | 178629.7 (160318.6, 197221) |
| **Norway** | NOR | Cluster 4 | High income | Hospital and Retail | 13.5 (13.5, 13.5) | 100.7 (91, 111.1) | 80.7 (80.7, 80.7) | 0 (0, 0) | 207336.2 (186603.4, 228786.7) |

| CTA Name | ISO3 Code | Cluster | Income group | Sectors available in IQVIA MIDAS* | Total DID (95%CI)* | Infection deaths per 100K (95%CI) | Percent Access use* | Not Recommended DID (95%CI)* | Infection incidence per 100k (95%CI) |
|---|---|---|---|---|---|---|---|---|---|
| **Sweden** | SWE | Cluster 4 | High income | Retail Only | 13.6 (13.1, 14.5) | 86.9 (79.7, 95.6) | 70.7 (68.2, 72.4) | 0 (0, 0) | 206009.3 (186138.6, 226924.2) |
| **Finland** | FIN | Cluster 4 | High income | Hospital and Retail | 13.7 (13.7, 13.7) | 61.6 (56.1, 68.5) | 75 (75, 75) | 0.3 (0.3, 0.3) | 186109.7 (168888.4, 207787.3) |
| **Germany** | DEU | Cluster 4 | High income | Hospital and Retail | 14 (14, 14) | 106.1 (96.3, 116.7) | 57.2 (57.2, 57.2) | 0.1 (0.1, 0.1) | 191886.2 (174329.8, 210963.2) |
| **Austria** | AUT | Cluster 4 | High income | Hospital and Retail | 14.8 (14.8, 14.8) | 63.1 (57.3, 68.6) | 59.7 (59.7, 59.7) | 0.2 (0.2, 0.2) | 190386.4 (173339.8, 211020.9) |
| **Hungary** | HUN | Cluster 4 | High income | Hospital and Retail | 15.2 (15.2, 15.2) | 98 (88.7, 108.4) | 50.3 (50.3, 50.3) | 0 (0, 0) | 136699.5 (123952.8, 150929.3) |
| **Slovenia** | SVN | Cluster 4 | High income | Retail Only | 15.7 (15.2, 16.3) | 84.9 (77.5, 93.6) | 71.1 (69.5, 72.5) | 0 (0, 0) | 135177.5 (122272.2, 150758.1) |
| **Canada** | CAN | Cluster 4 | High income | Hospital and Retail | 15.8 (15.8, 15.8) | 77.1 (70.3, 83.9) | 67.9 (67.9, 67.9) | 0 (0, 0) | 296605.9 (266332.8, 328329.7) |
| **Lithuania** | LTU | Cluster 4 | High income | Hospital and Retail | 16.7 (16.7, 16.7) | 113.9 (102, 127.9) | 70 (70, 70) | 0.1 (0.1, 0.1) | 180409.6 (162101.1, 198836.2) |
| **Russian Federation** | RUS | Cluster 4 | Upper middle income | Hospital and Retail | 17.1 (17.1, 17.1) | 126.8 (113.7, 140.7) | 47.2 (47.2, 47.2) | 1.4 (1.4, 1.4) | 197108.4 (177994.9, 220286.7) |
| **Czechia** | CZE | Cluster 4 | High income | Hospital and Retail | 17.6 (17.6, 17.6) | 112.1 (103.9, 122.5) | 56.1 (56.1, 56.1) | 1.1 (1.1, 1.1) | 139234.8 (126348.8, 153889.1) |
| **United Kingdom** | GBR | Cluster 4 | High income | Hospital and Retail | 19.7 (19.7, 19.7) | 146.9 (134.7, 162.3) | 66.8 (66.8, 66.8) | 0 (0, 0) | 203521.9 (184385.8, 225771.7) |
| **Portugal** | PRT | Cluster 4 | High income | Hospital and Retail | 20.8 (20.8, 20.8) | 189.4 (172.1, 210.3) | 62.2 (62.2, 62.2) | 0.1 (0.1, 0.1) | 187600.2 (169661.8, 207084.8) |
| **Croatia** | HRV | Cluster 4 | High income | Hospital and Retail | 21.2 (21.2, 21.2) | 103.4 (95.3, 113.5) | 63 (63, 63) | 0 (0, 0) | 145251.8 (130997.4, 162232.4) |
| **Bulgaria** | BGR | Cluster 4 | Upper middle income | Hospital and Retail | 22.3 (22.3, 22.3) | 132.1 (118, 149.8) | 46 (46, 46) | 0.3 (0.3, 0.3) | 135930.5 (122806.2, 149547.3) |
| **Poland** | POL | Cluster 4 | High income | Hospital and Retail | 22.8 (22.8, 22.8) | 124.8 (115.9, 134.8) | 54.9 (54.9, 54.9) | 1.3 (1.3, 1.3) | 147707.2 (133726.2, 164581.5) |
| **Australia** | AUS | Cluster 4 | High income | Hospital and Retail | 22.8 (22.8, 22.8) | 60 (54.4, 66.4) | 80.9 (80.9, 80.9) | 0 (0, 0) | 227663.1 (205682.4, 251802.9) |
| **Slovak Republic** | SVK | Cluster 4 | High income | Hospital and Retail | 23 (23, 23) | 101.4 (91.8, 112.1) | 35.9 (35.9, 35.9) | 2.5 (2.5, 2.5) | 140303.1 (125935.8, 156595) |
| **Belarus** | BLR | Cluster 4 | Upper middle income | Hospital and Retail | 23.5 (23.5, 23.5) | 84.7 (74.7, 95.6) | 55.2 (55.2, 55.2) | 1.9 (1.9, 1.9) | 184081.6 (165903.6, 202843.5) |
| **United States** | USA | Cluster 4 | High income | Hospital and Retail | 23.7 (23.7, 23.7) | 87.4 (79.9, 95.4) | 70.5 (70.5, 70.5) | 0 (0, 0) | 284895.5 (257427.6, 312333.6) |

| CTA Name | ISO3 Code | Cluster | Income group | Sectors available in IQVIA MIDAS* | Total DID (95%CI)* | Infection deaths per 100K (95%CI) | Percent Access use* | Not Recommended DID (95%CI)* | Infection incidence per 100k (95%CI) |
|---|---|---|---|---|---|---|---|---|---|
| **Bosnia and Herzegovina** | BIH | Cluster 4 | Upper middle income | Retail Only | 23.8 (22.1, 26.2) | 102.6 (90, 118.5) | 64.9 (63.2, 66.7) | 1 (1, 1.1) | 140733.7 (126776, 155850.8) |
| **New Zealand** | NZL | Cluster 4 | High income | Hospital and Retail | 24.1 (24.1, 24.1) | 61.5 (56.5, 67.2) | 79.5 (79.5, 79.5) | 0 (0, 0) | 252252.9 (226792.5, 281215.5) |
| **Italy** | ITA | Cluster 4 | High income | Hospital and Retail | 24.7 (24.7, 24.7) | 82.6 (75, 90.8) | 46.6 (46.6, 46.6) | 0 (0, 0) | 189354.8 (171742, 209623.2) |
| **France** | FRA | Cluster 4 | High income | Hospital and Retail | 25.1 (25.1, 25.1) | 97.8 (89.3, 107.3) | 69.7 (69.7, 69.7) | 1.1 (1.1, 1.1) | 188792.7 (171673.6, 209189.2) |
| **Korea, Rep.** | KOR | Cluster 4 | High income | Hospital and Retail | 25.2 (25.2, 25.2) | 75.4 (66, 84.3) | 45.1 (45.1, 45.1) | 0.5 (0.5, 0.5) | 213820.4 (188409.3, 240782.7) |
| **Luxembourg** | LUX | Cluster 4 | High income | Retail Only | 25.2 (23.8, 26.9) | 67.3 (60, 75.4) | 57.5 (55.5, 59.5) | 0.2 (0.2, 0.2) | 192543.2 (173067.8, 213175.5) |
| **Ireland** | IRL | Cluster 4 | High income | Hospital and Retail | 25.3 (25.3, 25.3) | 76.5 (69, 85.2) | 66 (66, 66) | 0 (0, 0) | 198651.1 (179133.4, 221366.6) |
| **Belgium** | BEL | Cluster 4 | High income | Hospital and Retail | 25.6 (25.6, 25.6) | 144.1 (131, 159.4) | 61.8 (61.8, 61.8) | 0 (0, 0) | 190406.2 (172714.3, 210182.8) |
| **Spain** | ESP | Cluster 4 | High income | Hospital and Retail | 26.9 (26.9, 26.9) | 105.1 (96, 115.1) | 62.7 (62.7, 62.7) | 0.4 (0.4, 0.4) | 184253.9 (163627.3, 204942.2) |
| **Romania** | ROU | Cluster 4 | High income | Hospital and Retail | 27.3 (27.3, 27.3) | 141.5 (129.1, 155.8) | 52.2 (52.2, 52.2) | 0.4 (0.4, 0.4) | 137499.9 (123642.9, 153304.5) |
| **Serbia** | SRB | Cluster 4 | Upper middle income | Retail Only | 29.3 (28.9, 30) | 139.9 (123.3, 161.3) | 55.7 (54.9, 56.4) | 0.6 (0.6, 0.7) | 179842.4 (159325.6, 200563.4) |
| **Greece** | GRC | Cluster 4 | High income | Retail Only | 37.5 (36.7, 38.8) | 135.7 (123.9, 148.7) | 43.8 (42.6, 44.5) | 0.3 (0.3, 0.3) | 177029.2 (160878.2, 197280) |

*Source: Total DID based on IQVIA MIDAS® data for 2019, reflecting estimates of real-world activity. Copyright IQVIA. All Rights Reserved*

##### 1.2.2.3 Infection draws for uncertainty

To account for uncertainty from GBD infection case estimates throughout the analytical pipeline for estimating optimal Total and AWaRe antibiotic need, we sampled 1000 draws from the distribution of each infection in GBD using the point estimate, upper and lower bounds. Distributions were selected by finding the best fit distribution for each CTA for each infection by minimising the sum of the squared errors (differences between fitted data and observed mean, upper and lower CIs); the most common best fit distribution across CTAs for each infection type was used (i.e. log-normal).

Matrices of case counts per CTA are used in the final estimation of Watch and Reserve antibiotics using the benchmarking approach; for other stages of analysis matrices of incident cases per 100,000 people is used. We drew from the distribution of the case counts for each infection then summed the matrices of infection cases to have a matrix of total infection cases; this was then converted to incidence per 100,000 people using population estimates.

Resistant sepsis cases were converted to incidence as cases per 100,000 people. For estimating resistant cases as outlined in section 1.1.3, for each of the pathogen, resistance and sepsis datasets, we used point estimates and uncertainty intervals to draw 1000 draws from a gamma distribution for each pathogen and bug-drug combination and sepsis cases for each CTA to then calculate the number of resistant sepsis cases for each bug-drug combination of interest which resulted in 1000 draws of resistant cases for each bug-drug combination for each CTA.

##### 1.2.2.4 Adjusted case counts

To assess the infections that may need adjusting due to potential underestimates in GBD, we plotted number of doctors per 1000 people against the infection incidence (per 100,000 people) of all infections from GBD (Figure 10A). A positive relationship exists between doctors per capita and UTI and cellulitis (Figure 10B and Figure 10C). We considered this trend a potential sign of underdiagnosis of these infections where there were fewer doctors per 1000 people and therefore wanted to adjust case counts in these lower doctor areas for estimating expected antibiotic use.

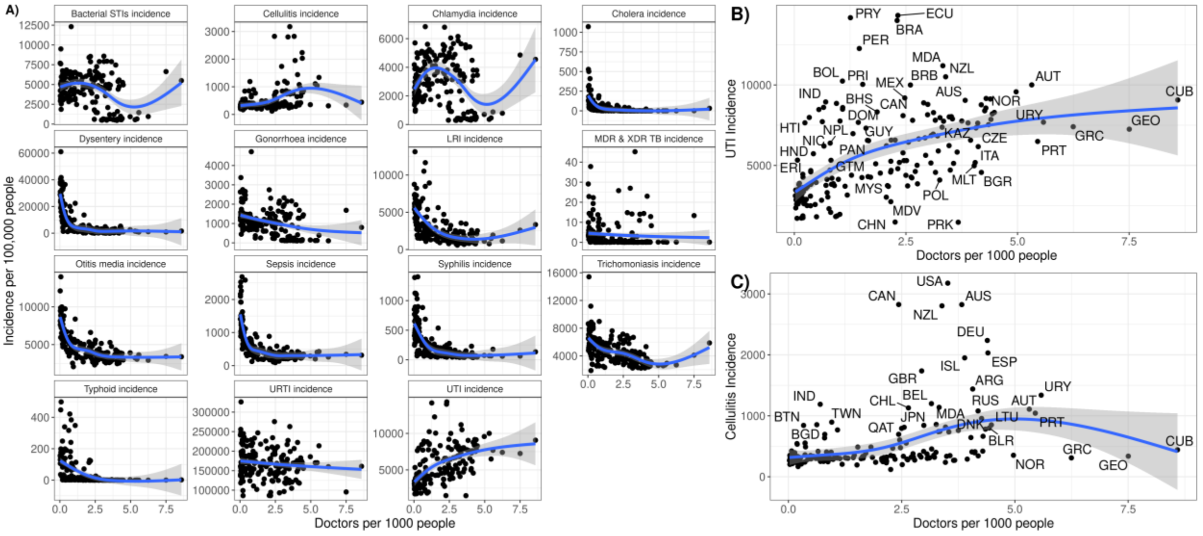

*Figure 10. A) Plot of doctors per 1000 people against infection incidence for infections included in GBD. B) Zoomed plot of doctors per 1000 people against urinary tract infection (UTI) incidence and C) Zoomed plot of doctors per 1000 people against cellulitis incidence where we see an increasing relationship*

To adjust for this, we fit a logistic growth curve model separately for the relationship between doctors per 1000 people and incidence to datasets containing each one of the 1000 draws per CTA from the incidence distribution for UTI and cellulitis respectively and then predicted cases and adjusted for residuals. The logistic growth curve model was defined as (Equation 2):

$$f(x) = \frac{L}{(1 + e^{-k(x-x0)})}$$

*Equation 2. Logistic growth curve model. L = the asymptote, the maximum value as the curve approaches carrying capacity, k = the growth rate, and $x_0$ = the inflection point*

where $L$ = the asymptote, the maximum value as the curve approaches carrying capacity, $k$ = the growth rate, and $x_0$ = the inflection point. The model was fit to each dataset where y = infection incidence and x = doctors per 1000 people using nonlinear least squares with Levenberg-Marquardt algorithm.[56,57] Using the fitted parameters, we then define the threshold $xt$ which finds the x value where the logistic curve is within a tolerance of $L$ with tolerance defined as 0.01 for our purpose (Equation 3)Equation 3. Threshold definition.:

$$\text{Threshold } xt = x0 + \frac{\ln\left(\frac{1}{tolerance} - 1\right)}{k}$$

*Equation 3. Threshold definition. $x_0$ = inflection point*

We adjust the UTI and cellulitis incidence where *number of doctors (x)* $< xt$ and use observed incidence values where number of doctors (x) $\geq xt$. We adjust incidence using the predicted value from the logistic curve + an adjustment defined as the difference between incidence at the threshold ($y_{xt}$) and the predicted value. A sample of the predicted adjusted values vs original observed values are in Figure 11A (UTI) and Figure 11B (cellulitis)

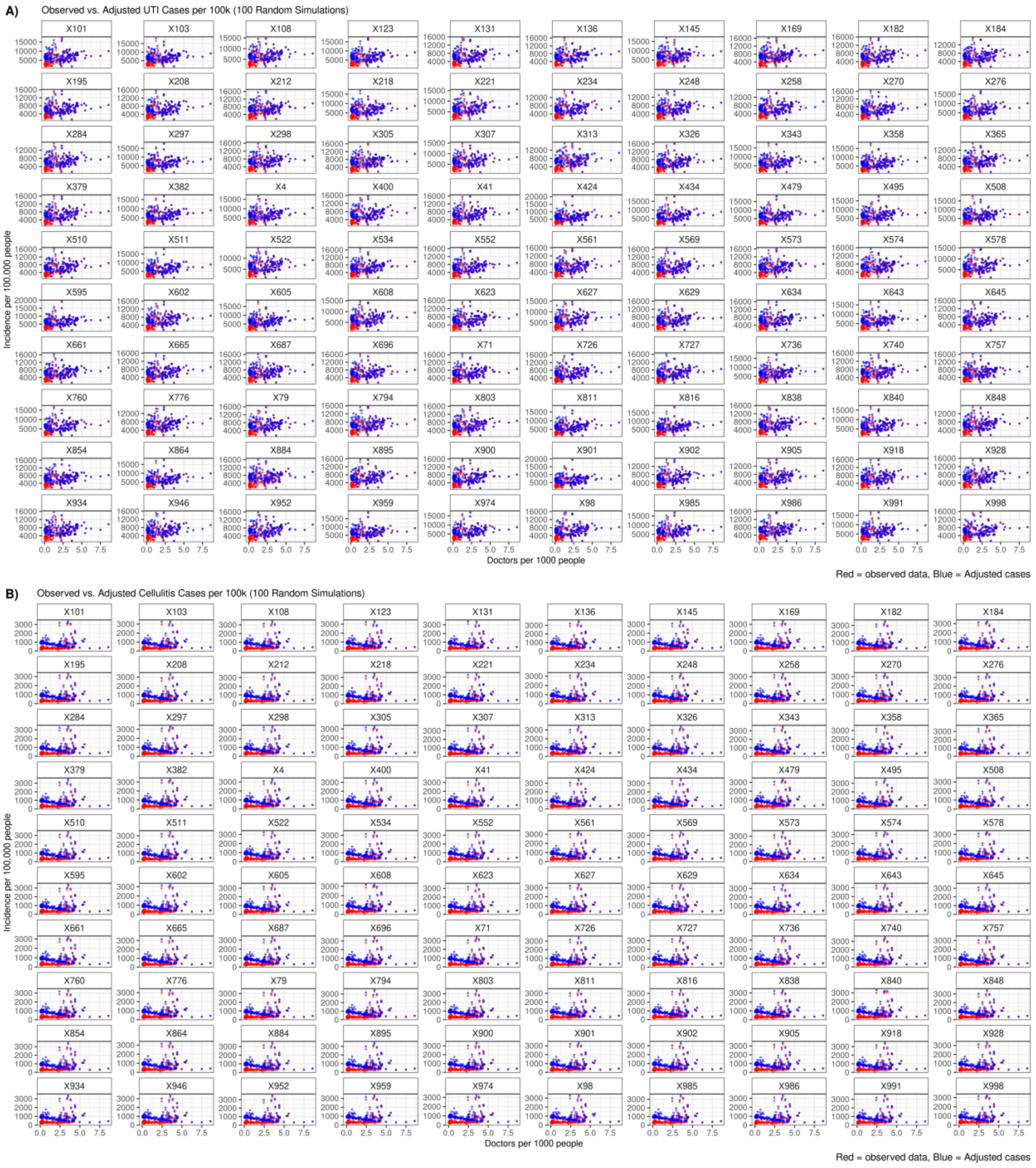

*Figure 11. Scatterplot of the adjusted incidence (blue) compared to observed incidence (red) on a subset of 100 draws for A) UTI and B) cellulitis.*

###### 1.2.2.5 *Optimal antibiotic use benchmarking*

An overview of the process for estimating optimal AWaRe antibiotic use is in supplemental Figure 4 and supplemental Figure 12.

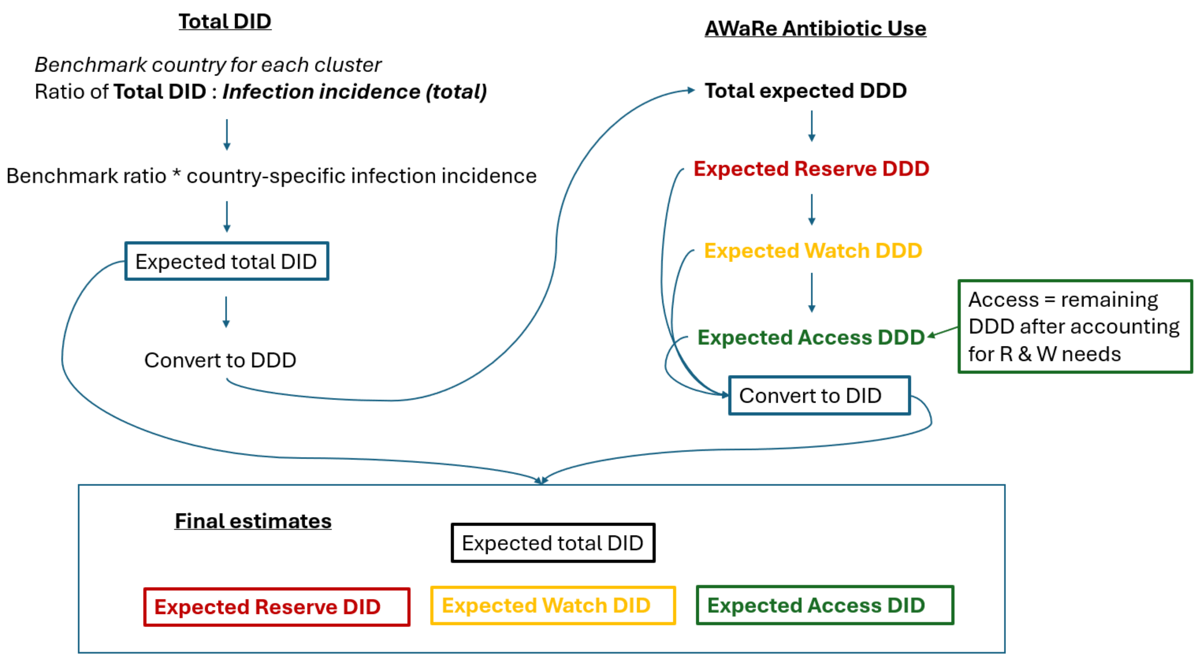

*Figure 12. Overview of the process to estimate expected AWaRe antibiotic use in defined daily doses (DDD) per 1000 inhabitants per day (DID).*

###### 1.2.2.5.1 Expected Total DID

For each benchmark CTA in each cluster, the ratio of total DID to total infection incidence was calculated taken using the reported total DID from MIDAS® and total infection incidence (Equation 4). For each non-benchmark CTA in the cluster, the respective benchmark ratio was multiplied by the total infection incidence in each CTA to estimate the total DID required to treat that infection burden (Equation 5). This was repeated at the draw-level for 1000 imputations of antibiotic use data where the benchmark CTA had imputed data and the 1000 draws of total infection incidence for each CTA in the cluster. For subsequent operations, these estimates of expected total DID were converted to DDD using World Bank population estimates.

**Benchmark ratio**:

$$Benchmark\ ratio = \frac{Total\ DID\ in\ benchmark\ CTA}{Total\ infection\ incidence\ in\ benchmark\ CTA}$$

*Equation 4. Ratio from benchmark CTAs used to estimate Total DID for CTAs within each cluster. CTA = countries, territories and areas, DID = defined daily doses per 1000 inhabitants per day*

**Total DID estimate:**

$$Total\ DID\ (nonbenchmark\ CTA) \\ = (Benchmark\ ratio\ for\ cluster) * (Total\ infection\ incidence\ in\ respective\ CTA)$$

*Equation 5. Equation for estimating Total DID for each CTA in a cluster. CTA = countries, territories and areas, DID = defined daily doses per 1000 inhabitants per day*

###### 1.2.2.5.2 Expected Reserve antibiotic use estimation

We use a similar approach for estimating Reserve antibiotic need as Total antibiotic need. We used the ratio of Reserve antibiotics to resistant infections from the cluster 4 benchmark CTA in each dataset to estimate the total Reserve antibiotics each CTA would require. We use only the cluster 4 benchmark CTA as it is a high-income CTA and we assume there are no access issues to Reserve antibiotics in this CTA. In MIDAS®, antibiotic use is reported separately by molecule, so we summed the separate volumes (DDDs) of Reserve antibiotics covering Gram-negative infections (carbapenem-resistant infections) and Gram-positive (vancomycin-resistant infections) and MDR/XDR TB infections separately for cluster 4 benchmark CTA. This allowed us to account for differing burdens in Gram-negative and Gram-positive and TB resistant infections from the benchmark CTA to non-benchmark CTAs. Table 5 presents the antibiotics, median DDD, median infection incidence (95% CI) used for benchmarking. The respective ratios from the benchmark CTA were multiplied by the respective infection burden of CR-infections and VR-infections and MDR/XDR TB for all non-benchmark CTAs. Resistance-specific Reserve needs were summed to estimate total Reserve needs.

To account for resistant infection incidence uncertainty, the ratio operation was repeated draw-wise across the 1000 draws for each respective resistance pattern. Each ratio from the benchmark CTA was multiplied draw-wise by the 1000 draws of respective resistance patterns for the non-benchmark CTAs. Estimates of optimal Reserve are presented as median and 95%CI.

*Table 5. Values from Cluster 4 benchmark CTA (Switzerland) of DID of Reserve antibiotics covering Gram-positive/MDR&XDR TB and Gram-negative resistance pathogens.*

| **Resistance pattern** | **Antibiotic(s) in dataset** | **Resistance incidence (per 100,000 people)** | **Antibiotic DID*** | **Antibiotic DDD*** |
|---|---|---|---|---|
| VRO (VRSA, VR-E.faecium, MDR/XDR TB | Dalbavancin, Daptomycin, Linezolid, Tedizolid | VRO: 0.047<br>MDR/XDR TB: 0.0225 | 0.018 | 56378.0 |
| CRO (CRE, CRAB, CRPA) | Aztreonam, Ceftaroline-fosamil, Ceftazidime+Avibactam, Ceftolozane+Tazobactam, Colistin, Fosfomycin, Tigecycline | CRO: 0.53 | 0.002 | 6420.7 |

*Note: Switzerland is the cluster 4 (predominantly high income) benchmark CTA. DDD = defined daily doses, DID = DDD/1000 inhabitants/day, VRO = vancomycin resistant organism, VRSA = vancomycin-resistant Staphylococcus aureus, VR-E.faecium = vancomycin-resistant Enterococcus faecium, MDR/XDR TB = multi-drug-resistant/extremely-drug-resistant tuberculosis, CRO = carbapenem resistant organism, CRE = carbapenem resistant Enterobacterales, CRAB = carbapenem resistant Acinetobacter baumannii, CRPA = carbapenem-resistant Pseudomonas aeruginosa. *Source: Total DID based on IQVIA MIDAS® data for 2019, reflecting estimates of real-world activity. Copyright IQVIA. All Rights Reserved*

###### 1.2.2.5.3 Expected Watch antibiotic use estimation

We estimated Watch antibiotic needs based on the case counts of conditions where the AWaRe Book recommends Watch antibiotics and the DDDs of the recommended treatment courses for these infections. From GBD, we include sepsis, typhoid, dysentery, lower respiratory infections and urinary tract infections and MDR TB cases. We include MDR TB cases in Watch as current treatment guidelines include levofloxacin or moxifloxacin (Watch antibiotics). We estimated the burden of hospital infections covered in the AWaRe Book that require Watch antibiotics which do not have estimates in GBD such as severe skin and soft tissue infections (i.e. necrotising fasciitis), hospital-acquired pneumonia (HAP) and intra-abdominal infections using alternate sources (described in section 1.1.2 ).

The AWaRe Book provides risk-stratified guidance for antibiotic selection and for some infections such as LRIs and UTIs Watch antibiotics are only recommended for more severe infections. Given that GBD does not differentiate between upper and lower UTIs and doesn't estimate severity of LRIs, we assumed only a proportion of the cases for UTI and LRIs may require a Watch antibiotic (see below). We also adjust typhoid case counts to account for treatment needs of suspected typhoid cases in endemic areas (see below).

After adjusting the case counts for each infection, we use the maximum DDDs per treatment course for each infection as recommended in the AWaRe Book or other relevant guideline (Table 6) and multiply the case counts by DDDs for a total DDDs required for each infection. For typhoid where the antibiotic choice can differ due to fluoroquinolone resistance, we use the ciprofloxacin DDD for primary analysis and conduct sensitivity analyses using DDD of alternate treatment choices. This operation is performed on 1000 draws from the distribution of cases for each infection. These estimates are summed to a total Watch need per CTA and converted to DID using population estimates and presented as median and 95% CI.

**<u>Watch DDDs per Infection</u>**

*Table 6. Highest DDDs per treatment course of the infections included in analysis for estimating Watch antibiotic use. Where the AWaRe Book recommends several Watch antibiotics, we used the Watch antibiotic with the highest DDD per treatment course. There are also several infections for which the first choice of treatment is recommended as either an Access or Watch antibiotic and for these we use the Watch DDD estimate unless otherwise stated in the case adjustments. Note that some oral antibiotics have higher DDDs than parenteral treatment recommended for the same infection.*

| Infection | Antibiotic | Treatment course from AwaRe book | WHO DDD for administration | Total DDDs for a treatment course |
|---|---|---|---|---|
| **Severe diarrhoea / Dysentery** | Azithromycin | 500mg on day 1 + 250mg once daily for 3 days | 0.3g | 4.2 DDDs |
| **Cholera** | Azithromycin | 1g oral single dose | 0.3g | 3.3 DDDs |
| **Clinical sepsis of unknown origin** | Cefotaxime | 2g every 8 hours for 7 days | 4g | 10.5 DDDs |
| **Lower respiratory tract infections (community acquired pneumonia) (severe CAP)** | Cefotaxime + Clarithromycin (if CURB-65>=2) | 2g every 8 hours for 5 days (Cefotaxime) + 500mg every 12 hours for 5 days (Clarithromycin) | 4g, 1g (parenteral Clarithromycin) | 12.5 DDD (7.5 DDD Cefotaxime, 5 DDD Clarithromycin) |
| **Lower respiratory tract infections (Hospital acquired pneumonia) (severe)** | Cefotaxime | 2g every 8 hours for 7 days | 4g | 10.5 DDDs |

| Infection | Antibiotic | Treatment course from AwaRe book | WHO DDD for administration | Total DDDs for a treatment course |
|---|---|---|---|---|
| **Typhoid**[1] | Ciprofloxacin (recommended for mild and severe cases where low risk of fluoroquinolone resistance) | 500mg twice daily for 10 days (severe) | 1g for oral | 10 DDDs |
| | Azithromycin (recommended for mild cases where there is high risk of fluoroquinolone resistance) | 1g on day 1, 500mg every 24 hours for 7 days | 0.3g for oral | 15 DDD |
| **Upper UTI** (severe cases)[2] | Ciprofloxacin | 500mg twice daily for 7 days | 1g for oral | 7 DDDs |
| **Chlamydia** (uncomplicated urogenital infection) | Azithromycin | 1g single dose (oral) | 0.3g for oral | 3.3 DDD |
| **Gonorrhoea** (genital / anorectal infections) | Ceftriaxone + Azithromycin | Ceftriaxone 250mg IM + Azithromycin 1g oral single dose | 2g parenteral ceftriaxone 0.3g oral azithromycin | 3.425 DDD (0.125 DDD ceftriaxone; 3.3 DDD azithromycin) |
| **Multi-drug-resistant Tuberculosis**[3] | Levofloxacin (* weight-based dosing so have used max for adult) | 2x500mg tablets per day of Levofloxacin for 26 weeks (26 x 7 x 1000mg = 182g) | 0.5g (oral) | 364 DDDs Levofloxacin |
| **Extensively drug-resistant Tuberculosis**[4] | Levofloxacin (where there is fluoroquinolone susceptibility) | 2x500mg tablets per day of Levofloxacin for 87 weeks (87 x 7 x 1000mg = 609g) | 0.5g (oral) | 1218 DDDs Levofloxacin |
| **Intra-abdominal infections**[5] | Cefotaxime (for mild and severe) | 2g every 8 hours (3 x per day) for 5 days = 30g | 4g | 7.5 DDD |
| **SSTIs (necrotising fasciitis)** | Piperacillin/ tazobactam | 4g + 500mg every 6 hours for 2-3 weeks (14-21 days) = 4.5 x 4 x 21 | 14g | 27 DDD |

*[1] Per the AWaRe Book, for typhoid where there is high-risk of fluoroquinolone resistance, recommended to use azithromycin for mild cases (7 days) and ceftriaxone for severe cases (10 days) (10 days of IV ceftriaxone or ciprofloxacin – recommended in low risk of fluoroquinolone resistance) have the same DDDs).*
*[2] Per the AWaRe Book, the maximum DDDs of Watch antibiotics to treat upper UTI is when using Ciprofloxacin (7 DDDs); severe cases may require IV antibiotics (Cefotaxime or Ceftriaxone) but Watch DDD per treatment course is lower than ciprofloxacin.*
*[3] A rapid communication in June 2024 on updates to TB treatment guidelines for MDR-TB(58,59) indicated that a 6-month treatment with bedaquiline, delamanid, linezolid (600mg), levofloxacin and clofazimine could be used instead of the 9-month oral regimen. Where there is fluoroquinolone resistance, recommended to drop levofloxacin, however we have included it for all cases in our estimates.*
*[4] For XDR-TB, longer treatment regimens of 18-months recommended(59); levofloxacin and moxifloxacin are not to be used where there is fluoroquinolone resistance in XDR TB however, we have assumed all XDR-TB cases receive a fluoroquinolone for 18 months for our estimates.*
*[5] For most IAIs, the AWaRe Book also recommends Access antibiotics (oral amoxicillin/clavulanic acid) as a first-line treatment option and recommends adding metronidazole to either amoxicillin/clavulanic acid or third-generation cephalosporin first choice.*

**Adjustments for suspected typhoid:**

We consider that there is diagnostic uncertainty for typhoid in endemic areas given poor diagnostics available(60,61) and nonspecific symptoms of typhoid such as fever, fatigue, and diarrhoea(62) where it may be reasonable for suspected typhoid cases to be treated with Watch antibiotics to prevent the adverse outcomes of untreated typhoid. While diagnostic uncertainty exists even in non-endemic settings, suspected typhoid treatment in typhoid endemic CTAs may have a higher influence on required Watch antibiotics at a national level than in non-endemic settings. In some surveillance studies between 3 to 33 suspected typhoid cases are diagnosed for every confirmed typhoid case.[63,64] We assume that GBD estimates of typhoid are confirmed cases, thus would need to be adjusted to estimate suspected typhoid cases that may require antibiotic treatment with Watch antibiotics. Therefore, for CTAs where median typhoid incidence per 100,000 in GBD is above 50 cases per 100,000 people (Figure 13), we multiplied the case counts by 10 (Surveillance of Enteric Fever in Asia Project estimate[63]) to estimate a higher burden of suspected typhoid that may require antibiotic treatment in these CTAs. In low typhoid CTAs, we have used the typhoid case count as in GBD.

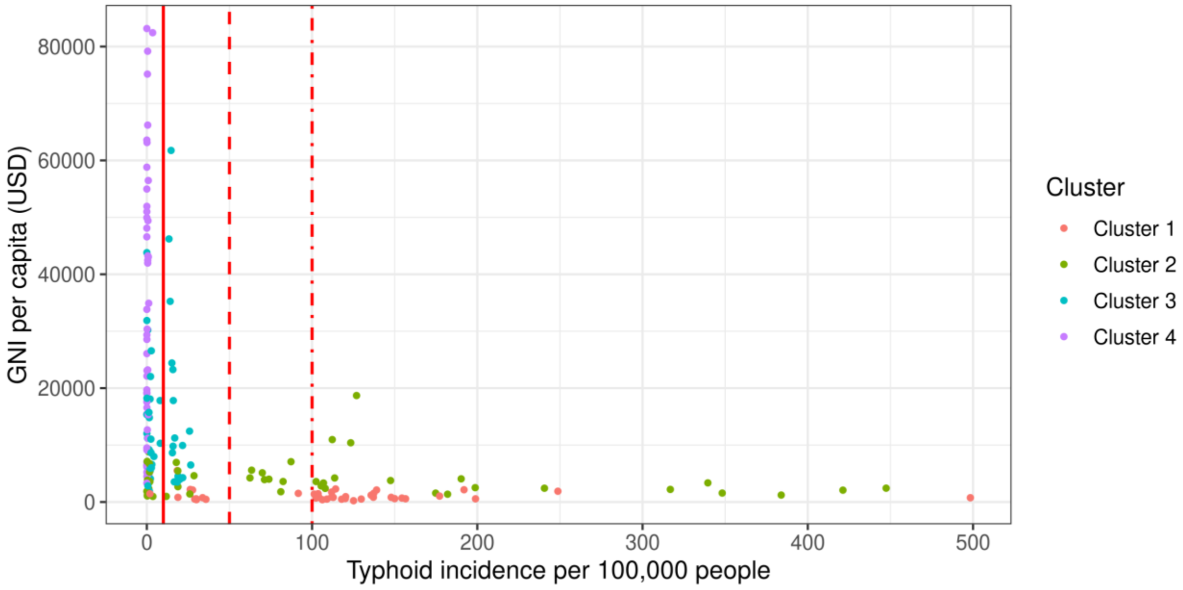

*Figure 13. The median typhoid incidence per 100,000 people from GBD for 186 CTAs, territories and areas (CTAs) in the analysis with cut-offs at 10 cases per 100,000, 50 cases per 100,000 and 100 cases per 100,000. CTAs are coloured by cluster assignment.*

**Upper urinary tract infections requiring Watch antibiotics**

The proportion of UTIs cases that may be upper UTIs and thus requiring a Watch antibiotic per the AWaRe Book was estimated using the Clinical Practice Research Datalink (CPRD) Aurum database[65] (protocol number: 23_003072) a UK database of representative general practices (GP) and has been used extensively as it provides anonymised data for public health research. The database contains information on patients' GP practice, demographic characteristics, diagnoses and symptoms, prescriptions, vaccination history, laboratory tests, and specialist referrals. We estimated the split of cases between upper and lower UTI in CPRD among patients with any UTI diagnosis who received any antibiotic prescription. We first used explicit diagnosis codes for lower and upper UTI, and then for those without an explicit diagnosis code for differentiation, we used the antibiotic treatment choice per NICE guidelines[66,67] as a proxy for differentiating between upper and lower UTIs. Sensitivity analyses for these estimates were performed using different scenarios for treatment length. The largest estimated percentage (20.7%) from the sensitivity analyses were used to estimate the upper UTI from the total estimated UTI cases for all CTAs (Figure 14). We assumed this proportion does not vary by CTA for the purpose of this analysis. We multiply our adjusted UTI cases counts (see section 1.2.2.4) by this percentage to get the number of UTI cases assumed to upper UTI cases that would require Watch antibiotics (Figure 15A).

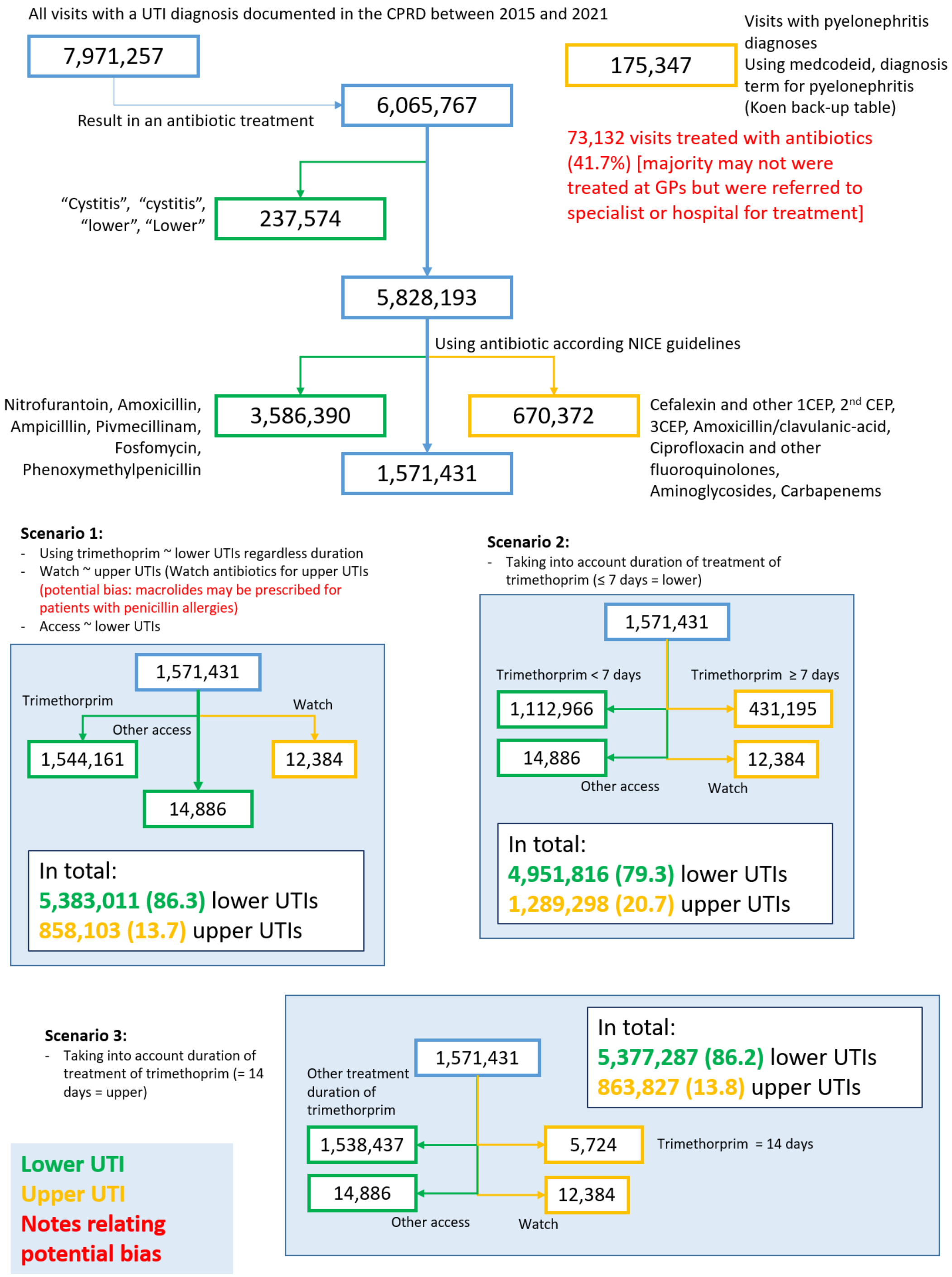

*Figure 14. A figure describing the process of estimating what percentage of urinary tract infections (UTIs) could be upper UTIs and thus requiring Watch antibiotics per the AWaRe Book. The figure outlines the classification strategy for assumed lower UTI requiring Access antibiotics and upper UTI requiring Watch antibiotics from cases in Clinical Practice Research Datalink (CPRD) between 2015 and 2021. The figure shows three scenarios and we use scenario 2 for our analysis which gives the highest estimate of upper UTI (21%).*

#### Lower respiratory infections

The AWaRe Book recommends Watch antibiotics for severe LRIs such as severe community-acquired pneumonia or hospital-acquired pneumonia and Access antibiotics for mild to moderate community-acquired pneumonia. GBD LRI case count includes pneumonia and bronchiolitis but excludes HAP.[(9)] For estimating the cases of severe CAP that would require Watch antibiotics, we assume 15% of LRI cases from GBD are severe CAP which would require Watch antibiotics[(9)] (Figure 15B). We assume this proportion did not vary by CTA for primary analyses. We use separate estimates for HAP that require Watch antibiotics (see section 1.1.2) and assume that all HAP cases are treated with Watch antibiotics.

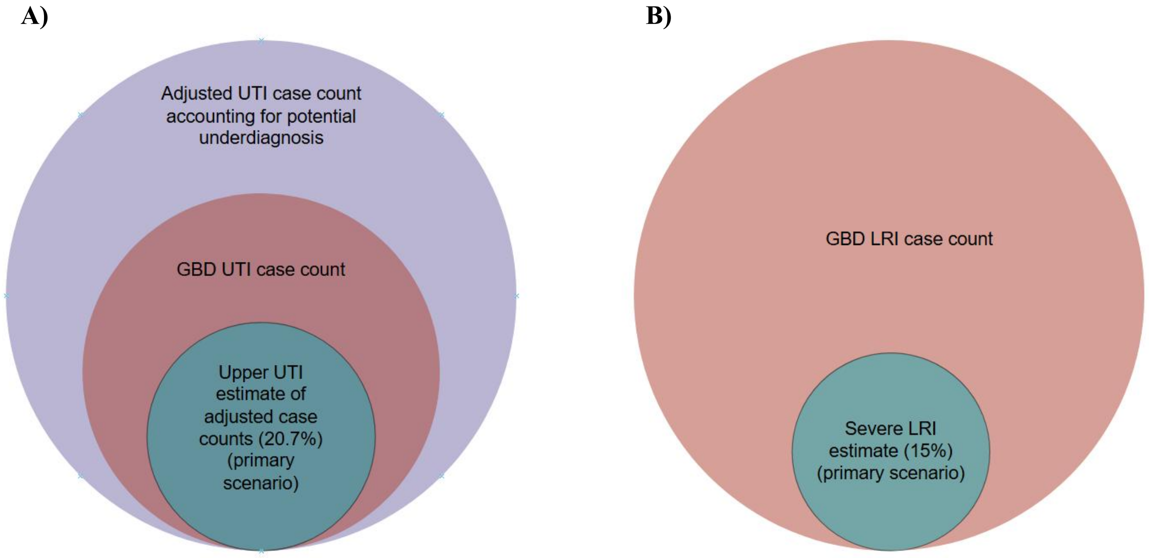

*Figure 15. Illustration of the subset of cases from GBD considered A) Upper UTI and B) severe LRI used for estimating Watch antibiotic use out of total cases. For UTI, in the primary analysis, the subset of cases was taken after adjusting the case counts (described in supplemental methods 1.2.2.4)*

**Other hospital infections requiring Watch antibiotics**

For severe infections such as intra-abdominal infections (IAI), and necrotising fasciitis (NF), which are included in the AWaRe Book as significant infections that may require Watch antibiotics but which do not have estimates in Global Burden of Disease (GBD), we approximated the burden of these infections using Global PPS data (see section 1.1.2 for IAI and below for NF). Using these approximated incidence estimates for total infections, we then adjust the cases to estimate the number of cases that would require a Watch antibiotic for IAI, and NF.

For IAI that could receive Watch antibiotics, for primary analyses, we assumed 80% of patients received one of the first-choice antibiotics in the AWaRe Book – either amoxicillin/clavulanic acid (Access) or a third-generation cephalosporin (cefotaxime or ceftriaxone) (Watch) and that 75% of those received a third-generation cephalosporin (Watch). We assume that the remaining 20% of patients also received a Watch antibiotic giving a percentage of total IAI that received Watch antibiotics of 80% (0.8 * 0.75 + 0.2).

For severe skin and soft tissue infections (SSTIs) such as necrotising fasciitis (NF) where the AWaRe Book recommends a Watch antibiotic (all other SSTI are recommended to receive Access), GBD estimates of cellulitis include the ICD-10 codes for NF thus these infections are already included in the total infection count for our analysis. There are no national estimates of necrotising fasciitis so we assume all NF cases would be hospitalised and derive estimates of NF cases using hospital data from global PPS using the same method as for IAI described in section 1.1.2. For estimating the subset of SSTIs that require Watch antibiotics, we assume that NF accounts for 5% of hospitalised SSTIs[68,69]. Using this percentage, we scale the total SSTIs reported in Versporten et al[11] to estimate the number of cases that could be NF and then take the ratio of these cases to reported sepsis cases in gPPS (ratio = 0.2 NF cases to sepsis). This ratio was multiplied by the GBD estimate of total sepsis cases in each CTA to get an estimate of NF cases in each CTA. We did not adjust for length of stay in PPS as this information was unavailable. We use hospitalised SSTI data for estimating the cases of NF based on reported data in gPPS rather than adjusting 5% of all cellulitis SSTIs as this would over-inflate the number of NF cases due to the high number of mild SSTIs included in the overall estimate from GBD.

##### 1.2.3 Sensitivity analyses

Our primary analysis (scenario 1) used the adjusted case counts as described in the methods above to estimate optimal antibiotic levels required. We conducted three sensitivity analyses to explore the impact of our assumptions of case counts on expected antibiotic use and the impact of benchmark CTA selection. The four analysis scenarios are summarised in Table 7.

For scenario 2, we estimate an upper level of optimal Watch DID antibiotics. Given our case adjustments for suspected typhoid and estimates of proportion of cases of LRI, UTI and hospital infections that may require antibiotics primarily affect the estimates of Watch antibiotics, we look at higher estimates of suspected typhoid and higher estimates of upper UTI and severe LRI cases and HAIs. We use the same total infection incidence with inflated case counts as in the main scenario. For optimal Watch DID, we inflate suspected typhoid cases to be 25 times higher than the GBD estimated typhoid burden in CTAs where incidence was greater than 10 per 100,000 (Figure 13). We also use a higher DDD for typhoid of 15 DDD which is a treatment course of 7 days of azithromycin (instead of 10 DDD for ciprofloxacin or ceftriaxone) (Table 6). We assume that 30% of UTI cases are upper UTI cases in all clusters.(70) For LRI, in clusters 1, 2 and 3 we assume that 30% of LRI are severe(71,72) while for cluster 4 assume 20% are severe.(73) We assumed 100% of IAI cases received a Watch antibiotic. For NF, we assume that 15% of hospitalised SSTIs(74) are NF and follow the same process to take the ratio (ratio = 0.5 NF to sepsis cases) and then multiply by the GBD sepsis cases to get the total cases of NF that would require Watch antibiotics.

For scenario 3, we estimate a lower bound of total and AWaRe antibiotic use by using the case counts as reported directly from GBD without any adjustments for suspected typhoid, or underestimates of UTI and cellulitis. We also exclude MDR/XDR TB from these estimates as TB treatments can be procured under different programs in some CTAs. For typhoid, we use 10 DDD which is the DDD for 10 days of ciprofloxacin or ceftriaxone. Total infection incidence is the sum of infection incidence as reported in GBD. We assume that 20% of UTI cases are upper UTI and that 15% of LRI cases are severe across all benchmark groups.

For scenario 4, we select a different benchmark CTA in benchmark group 3 by using Colombia which has a lower total DID than Morocco (as used in primary analyses) to compare the impact of benchmark CTA selection on estimates of total and AWaRe antibiotic use. We use the same infection parameters as in scenario 1 (primary analysis) and only change the benchmark CTA.

*Table 7. Table of the analysis scenarios used for estimating expected total and AWaRe antibiotic use.*

| | **Scenario 1** | **Scenario 2** | **Scenario 3** | **Scenario 4** |
|---|---|---|---|---|
| **Scenario description** | ***Primary analysis*** | ***Sensitivity analysis 1 - Upper estimate of Watch ("high Watch" scenario)*** | ***Sensitivity analysis 2 - lower estimate of Total, Watch and Access ("unadjusted case counts, excl. TB" scenario)*** | ***Sensitivity analysis 3 – impact of benchmark CTA selection ("alternate benchmark CTA" scenario)*** |
| **Antibiotic use data** | MIDAS® dataset post imputation | MIDAS® dataset post imputation | MIDAS® dataset post imputation | MIDAS® dataset post imputation |
| **Benchmark CTA** | Clusters1, 2, 3: Morocco<br>Cluster4: Switzerland | Clusters 1, 2, 3: Morocco<br>Cluster 4: Switzerland | Clusters 1, 2, 3: Morocco<br>Cluster 4: Switzerland | Clusters 1, 2, 3: Colombia<br>Cluster 4: Switzerland |
| **Total infection incidence** | • GBD infection incidence per 100,000 people as reported (no adjustment) for LRI, URI, OM, syphilis, gonorrhoea, chlamydia, trichomoniasis, sepsis, MDR+XDR TB<br>• Adjusted case counts for UTI and cellulitis for potential underestimation<br>• HAP estimated from ECDC<br>• IAI estimated using gPPS data | • GBD infection incidence per 100,000 people as reported for LRI, URI, OM, syphilis, gonorrhoea, chlamydia, trichomoniasis, sepsis, MDR+XDR TB<br>• Adjusted case counts for UTI and cellulitis for potential underdiagnosis<br>• HAP estimated from ECDC<br>• IAI estimated using gPPS data | • GBD infection incidence per 100,000 people as reported (no adjustment) for LRI, URI, OM, syphilis, gonorrhoea, chlamydia, trichomoniasis, sepsis, UTI and cellulitis (excludes TB)<br>• No additional hospital infections estimated from gPPS | • GBD infection incidence per 100,000 people as reported (no adjustment) for LRI, URI, OM, syphilis, gonorrhoea, chlamydia, trichomoniasis, sepsis, MDR+XDR TB<br>• Adjusted case counts for UTI and cellulitis for potential underestimation<br>• HAP estimated from ECDC<br>• IAI estimated using gPPS data |
| **Watch scenario** | • Adjusted case counts for UTI and cellulitis for potential underestimation<br>• Inflated typhoid cases 10x GBD where incidence was >50 per 100,000 cases<br>• 20% of adjusted UTI cases assumed to be upper UTI across all CTAs<br>• 15% LRI cases considered severe CAP<br>• 80% of IAI cases get Watch<br>• 100% of HAP cases<br>• 5% of hospitalised SSTI in gPPS are necrotising fasciitis | • Adjusted case counts for UTI and cellulitis for potential underestimation<br>• Inflated typhoid 25x GBD where incidence was >10 per 100,000; used higher DDD of 15 DDD per treatment course (azithromycin course)<br>• 30% of adjusted UTI cases assumed to be upper UTI<br>• 30% of LRI cases considered severe CAP in clusters 1, 2, 3 (predominantly LMIC), 20% in cluster 4 (predominantly HIC)<br>• 100% of IAI cases<br>• 100% of HAP cases<br>• 15% of hospitalised SSTI in gPPS are necrotising fasciitis | • Case counts as reported in GBD for all infections (no adjustments for potential underestimation);<br>• No typhoid inflation for suspected typhoid<br>• 20% of UTI assumed to be upper UTI<br>• 15% of LRI cases assumed to be severe<br>• No additional hospital Watch infections from gPPS or ECDC<br>• Excludes MDR/XDR TB | • Adjusted case counts for UTI and cellulitis for potential underestimation<br>• Inflated typhoid cases 10x GBD where incidence was >50 per 100,000 cases<br>• 20% of adjusted UTI cases assumed to be upper UTI across all CTAs<br>• 15% LRI cases considered severe CAP<br>• 80% of IAI cases get Watch<br>• 100% of HAP cases<br>• 5% of hospitalised SSTI in gPPS are necrotising fasciitis |
| **Reserve scenario** | Ratio of cases GNB and GPB/MDR&XDR TB to Reserve DDD by coverage category | Ratio of cases GNB and GPB/MDR&XDR TB to Reserve DDD by coverage category | Ratio of cases GNB and GPB/MDR&XDR TB to Reserve DDD by coverage category | Ratio of cases GNB and GPB/MDR&XDR TB to Reserve DDD by coverage category |
| **Access scenario** | Difference between estimated total DDD and estimated Watch + Reserve | Difference between estimated total DDD and estimated Watch + Reserve | Difference between estimated total DDD and estimated Watch + Reserve | Difference between estimated total DDD and estimated Watch + Reserve |

*CAP = community acquired pneumonia; CTA = countries, territories and areas; DDD = defined daily dose; ECDC = European Centre for Disease Prevention and Control; GBD = Global Burden of Disease; GNB = Gram-negative bacteria; GPB = Gram-positive bacteria; gPPS = Global Point Prevalence Survey; HAP = Hospital acquired pneumonia; HIC = high income CTAs; IAI = intra-abdominal infection; LMIC = lower and middle income CTAs; LRI = lower respiratory infection; MDR TB = multidrug resistant tuberculosis; OM = Otitis media; URI = upper respiratory infection; UTI = urinary tract infection; XDR TB = extensively drug resistant tuberculosis*

*Table 8. Summary table of the variables included in the different stages of the analysis workflow from imputation through estimates of optimal antibiotic levels.*

| | Modelling stage | Imputation | | Bayesian model | | | Clustering analysis | Benchmark CTA selection | Estimating optimal antibiotic use | | | |
|---|---|---|---|---|---|---|---|---|---|---|---|---|
| | **Antibiotic data** | **MIDAS® use (with Not recommended)** | **MIDAS® antibiotic use (with re-classified Not recommended)** | **MIDAS® antibiotic use (with re-classified Not recommended)** | | | **N/A** | **IQVIA MIDAS® use (with Not recommended)** | **Total DID** | **Reserve** | **Watch** | **Access[1]** |
| | | | | **Access DID** | **Watch DID** | **Reserve DID** | | Total DID, percent Access, DID Not recommended | | | | |
| Category of Covariate | **Covariate** | | | | | | | | | | | |
| Population age | Proportion of population 0-4 | | | | | | | | | | | |
| | Proportion of population 5-14 | | | | | | | | | | | |
| | Proportion of population 15-64 | | | | | | | | | | | |
| | Proportion of population 65-74 | | | | | | | | | | | |
| | Proportion of population 75+ | | | | | | | | | | | |
| | Proportion of population living in rural areas | | | | | | | | | | | |
| | *Isometric log-ratio population age group 1*[2] | | | | | | | | | | | |
| | *Isometric log-ratio population age group 2* | | | | | | | | | | | |
| | *Isometric log-ratio population age group 3* | | | | | | | | | | | |
| | *Isometric log-ratio population age group 4* | | | | | | | | | | | |
| Income | GNI per capita | | | | | | | | | | | |
| | Proportion of population living below the $2.15 poverty line | | | | | | | | | | | |
| Healthcare workforce | Doctors per 1000 people | | | | | | | | | | | |
| | Nurses & midwives per 1000 people | | | | | | | | | | | |
| | Pharmacists per 1000 people | | | | | | | | | | | |
| Healthcare access | UHC coverage index | | | | | | | | | | | |
| | Population-weighted travel time to healthcare facilities | | | | | | | | | | | |
| Infection prevention | Rotavirus vaccination coverage proportion | | | | | | | | | | | |
| | PCV vaccination coverage proportion | | | | | | | | | | | |
| | Proportion of population with access to safe sanitation | | | | | | | | | | | |

| | Modelling stage | Imputation | | Bayesian model | | | Clustering analysis | Benchmark CTA selection | Estimating optimal antibiotic use | | | |
|---|---|---|---|---|---|---|---|---|---|---|---|---|
| | **Antibiotic data** | **MIDAS® use (with Not recommended)** | **MIDAS® antibiotic use (with re-classified Not recommended)** | **MIDAS® antibiotic use (with re-classified Not recommended)** | | | **N/A** | **IQVIA MIDAS® use (with Not recommended)** | **Total DID** | **Reserve** | **Watch** | **Access[1]** |
| | | | | **Access DID** | **Watch DID** | **Reserve DID** | | Total DID, percent Access, DID Not recommended | | | | |
| Comorbidities & other clinical risk factors | Proportion of population with 1+ relevant comorbidity | | | | | | | | | | | |
| | Malaria incidence[3] | | | | | | | | | | | |
| | Burns and wounds from road traffic accidents and fires incidence | | | | | | | | | | | |
| | Protein energy malnutrition prevalence | | | | | | | | | | | |
| Infection incidence (used as per 100,000 people; except used as cases for estimating expected Watch antibiotics)[4] | Lower respiratory infection incidence | | | | | | | | | | | |
| | Upper respiratory infection incidence | | | | | | | | | | | |
| | Otitis media incidence | | | | | | | | | | | |
| | Gonorrhoea incidence | | | | | | | | | | | |
| | Chlamydia incidence | | | | | | | | | | | |
| | Syphilis incidence | | | | | | | | | | | |
| | *Bacterial STI incidence (sum of gonorrhoea, chlamydia, syphilis)* | | | | | | | | | | | |
| | Trichomoniasis incidence | | | | | | | | | | | |
| | Cellulitis incidence | | | | | | | | | | | |
| | Dysentery incidence (*Shigella* spp., non-typhoidal *Salmonella* spp., Enterotoxigenic *E. coli*, *Campylobacter* spp.) | | | | | | | | | | | |
| | Diarrhoea incidence | | | | | | | | | | | |
| | Typhoid incidence | | | | | | | | | | | |
| | Cholera incidence | | | | | | | | | | | |
| | UTI incidence | | | | | | | | | | | |
| | Sepsis incidence | Correlated with UHC, MRSA, dysentery | Correlated with UHC, MRSA, dysentery | | | | | | | | | |
| | MDR and XDR TB incidence | | | | | | | | | | | |

| | Modelling stage | Imputation | | Bayesian model | | | Clustering analysis | Benchmark CTA selection | Estimating optimal antibiotic use | | | |
|---|---|---|---|---|---|---|---|---|---|---|---|---|
| | **Antibiotic data** | **MIDAS® use (with Not recommended)** | **MIDAS® antibiotic use (with re-classified Not recommended)** | **MIDAS® antibiotic use (with re-classified Not recommended)** | | | **N/A** | **IQVIA MIDAS® use (with Not recommended)** | **Total DID** | **Reserve** | **Watch** | **Access[1]** |
| | | | | **Access DID** | **Watch DID** | **Reserve DID** | | Total DID, percent Access, DID Not recommended | | | | |
| | HAP incidence[5] | | | | | | | | | | | |
| | IAI incidence[5] | | | | | | | | | | | |
| | Necrotising fasciitis incidence[5] | | | | | | | | | | | |
| | **Total infection incidence** | | | | | | | | | | | |
| Antibiotic Resistance | ESBL sepsis incidence | | | | | | | | | | | |
| | Carbapenem-resistant Acinetobacter baumannii sepsis incidence | | | | | | | | | | | |
| | CR-Pseudomonas aeruginosa sepsis incidence | | | | | | | | | | | |
| | CR-Enterobacterales sepsis incidence | | | | | | | | | | | |
| | *CR-organisms sepsis incidence (sum of CRAB, CRPA, CRE)* | | | | | | | | | | | |
| | MRSA sepsis incidence | | | | | | | | | | | |
| | Vancomycin resistant S. aureus sepsis incidence | | | | | | | | | | | |
| | Vancomycin resistant Enterococcus faecium sepsis incidence | | | | | | | | | | | |
| | *Vancomycin-resistant organism sepsis incidence (sum of VRSA, VR-E.faecium)* | | | | | | | | | | | |
| Aggregate infection estimates | **Infection mortality rate (excludes TB)** | Correlated with dysentery incidence | Correlated with dysentery incidence | | | | | | | | | |
| | **Total infection incidence** | | | | | | | | | | | |

[1]Note: Access is the difference between estimated Total DID and Reserve & Watch therefore not explicitly calculated with variables
[2]Note highly correlated with other population age group ratios so only used one
[3]Note: Malaria excluded from clustering because incidence was heavily zero skewed and in trials this was a very discriminative variable likely due to few CTAs having high incidence
[4] Note where infections are not listed individually as used in a step, they are all included in the total infection incidence which is used for benchmarking & total DID estimation
[5] HAP/IAI/Necrotising fasciitis incidences are derived using ECDC, Global PPS data and GBD data therefore are not included in sensitivity analysis scenario 3 which uses GBD data as is

### 2 SUPPLEMENTAL RESULTS

#### 2.1 ANTIBIOTIC USE REGRESSION MODEL

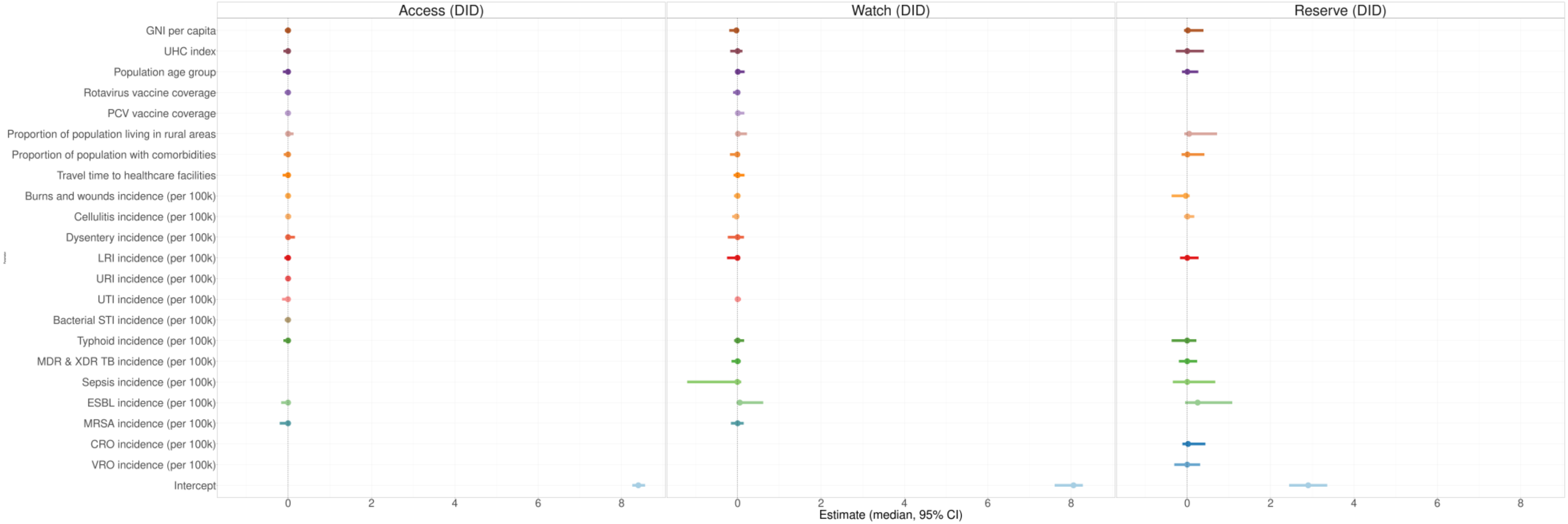

*Figure 16. Results of antibiotic use regression model showing the distribution of the posterior draws and 95%CrI for Access, Watch and Reserve antibiotic use. DID = defined daily doses (DDD)/1000 inhabitants/year. GNI = gross national income; UHC = universal health coverage; PCV = pneumococcal vaccine; LRI = lower respiratory infection; URI = upper respiratory infection; UTI = urinary tract infection; STI = sexually transmitted infection; MDR & XDR TB = multidrug resistant and extensively drug resistant tuberculosis; EBSL = extended spectrum beta-lactamase producing Enterobacterales; MRSA = methicillin-resistant staphylococcus aureus; CRO = carbapenem resistant organism; VRO = vancomycin-resistant organism*

*Table 9. Model estimates from the Bayesian multivariate model for AWaRe antibiotic use modelled using IQVIA MIDAS® data for 67 CTAs.*

| AWaRe Antibiotic Use | Variable | Estimate (95% CI) |
|---|---|---|
| Access | Intercept | 8.4 (8.26, 8.57) |
| | GNI per capita | -0.01 (-0.08, 0.03) |
| | Population age group | -0.01 (-0.12, 0.03) |
| | UHC index | -0.01 (-0.11, 0.06) |
| | Travel time to healthcare facilities | -0.01 (-0.13, 0.05) |
| | PCV vaccine coverage | -0.01 (-0.07, 0.03) |
| | Rotavirus vaccine coverage | -0.01 (-0.08, 0.03) |
| | Proportion of population living in rural areas | 0.01 (-0.03, 0.13) |
| | Proportion of population with comorbidities | -0.01 (-0.1, 0.04) |
| | URI incidence (per 100k) | 0 (-0.05, 0.05) |
| | LRI incidence (per 100k) | 0 (-0.09, 0.05) |
| | Typhoid incidence (per 100k) | -0.01 (-0.11, 0.04) |
| | Dysentery incidence (per 100k) | 0.01 (-0.06, 0.16) |
| | UTI incidence (per 100k) | -0.02 (-0.14, 0.02) |
| | Burns and wounds incidence (per 100k) | 0 (-0.07, 0.03) |
| | Cellulitis incidence (per 100k) | 0.01 (-0.02, 0.07) |
| | Bacterial STI incidence (per 100k) | -0.01 (-0.08, 0.03) |
| | MRSA incidence (per 100k) | -0.02 (-0.2, 0.04) |
| | ESBL incidence (per 100k) | -0.02 (-0.17, 0.04) |
| Watch | Intercept | 8.03 (7.61, 8.28) |
| | GNI per capita | -0.05 (-0.2, 0.02) |
| | Population age group | 0.02 (-0.04, 0.17) |
| | UHC index | -0.01 (-0.18, 0.11) |
| | Travel time to healthcare facilities | 0.01 (-0.1, 0.16) |
| | PCV vaccine coverage | 0.03 (-0.04, 0.16) |
| | Rotavirus vaccine coverage | -0.01 (-0.11, 0.05) |
| | Proportion of population living in rural areas | 0.04 (-0.04, 0.22) |
| | Proportion of population with comorbidities | -0.03 (-0.18, 0.04) |
| | LRI incidence (per 100k) | -0.03 (-0.25, 0.06) |
| | Typhoid incidence (per 100k) | 0.01 (-0.08, 0.16) |
| | Dysentery incidence (per 100k) | -0.01 (-0.24, 0.15) |
| | Burns and wounds incidence (per 100k) | -0.01 (-0.09, 0.06) |
| | Cellulitis incidence (per 100k) | -0.04 (-0.13, 0.02) |
| | Sepsis incidence (per 100k) | -0.16 (-1.21, 0.09) |
| | UTI incidence (per 100k) | 0 (-0.06, 0.08) |
| | MDR & XDR TB incidence (per 100k) | -0.01 (-0.15, 0.07) |
| | MRSA incidence (per 100k) | 0 (-0.16, 0.15) |
| | ESBL incidence (per 100k) | 0.14 (-0.03, 0.61) |
| Reserve | Intercept | 2.9 (2.45, 3.36) |
| | GNI per capita | 0.06 (-0.08, 0.39) |
| | Population age group | 0.02 (-0.13, 0.27) |
| | UHC index | 0.01 (-0.27, 0.4) |
| | Proportion of population living in rural areas | 0.15 (-0.07, 0.72) |
| | Proportion of population with comorbidities | 0.05 (-0.14, 0.41) |
| | LRI incidence (per 100k) | 0.02 (-0.17, 0.27) |
| | Burns and wounds incidence (per 100k) | -0.08 (-0.37, 0.06) |
| | Cellulitis incidence (per 100k) | 0.02 (-0.07, 0.17) |
| | Typhoid incidence (per 100k) | -0.02 (-0.37, 0.22) |
| | Sepsis incidence (per 100k) | 0.03 (-0.35, 0.67) |
| | MDR & XDR TB incidence (per 100k) | 0 (-0.2, 0.24) |
| | ESBL incidence (per 100k) | 0.33 (-0.05, 1.08) |
| | CRO incidence (per 100k) | 0.07 (-0.12, 0.44) |
| | VRO incidence (per 100k) | 0 (-0.31, 0.31) |

*Note: Estimates are on the DDD/1000 inhabitants/year scale not the DDD/1000 inhabitants/day (DID) scale. GNI = gross national income; UHC = universal health coverage; PCV = pneumococcal conjugate vaccine; URI = upper respiratory infection; LRI = lower respiratory infection; STI = sexually transmitted infection; UTI = urinary tract infection; MDR & XDR TB = multi-drug resistant and extremely-drug resistant tuberculosis; MRSA = methicillin-resistant *Staphylococcus aureus*; ESBL = extended-spectrum beta-lactamase; CRO = carbapenem resistant organism; VRO = vancomycin resistant organism

#### 2.2 Optimal AWaRe Antibiotic Use Benchmarking

##### 2.2.1 Clustering analysis

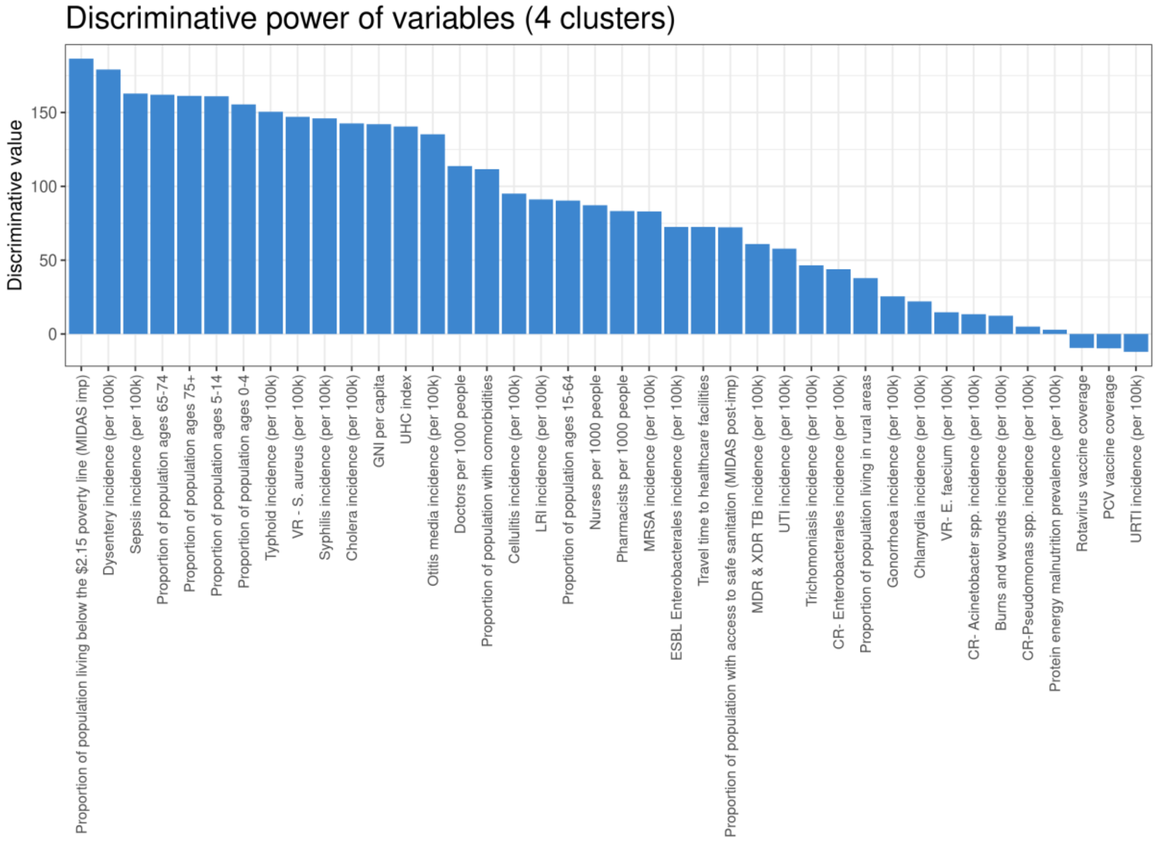

*Figure 17. Discriminative power of the variables included in the four groups cluster model. Negative bars indicate the variable was not discriminative and was not used by the model.*

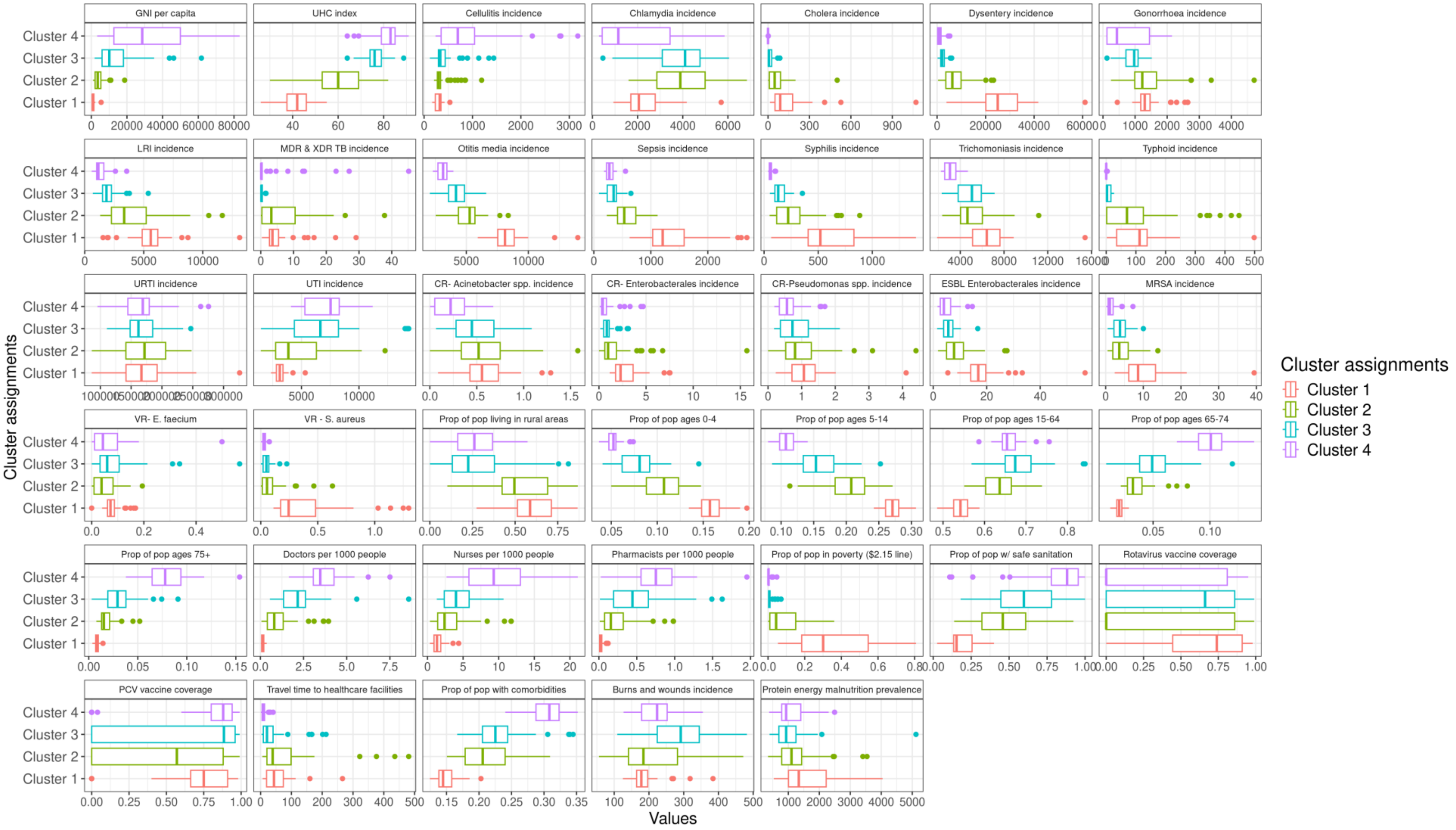

*Figure 18. Distribution of covariates by cluster for the four-group cluster model. GNI = gross national income, UHC = universal health coverage, LRI = lower respiratory tract infection, MDR & XDR TB = multi-drug resistant and extremely-drug resistant tuberculosis, URTI = upper respiratory tract infection, UTI = urinary tract infection, CR = carbapenem resistant, ESBL = extended-spectrum beta-lactamase, MRSA = methicillin-resistant Staphylococcus aureus, VR. E faecium = vancomycin-resistant Enterococcus faecium, VR-S.aureus = vancomycin-resistant Staphylococcus aureus, Prop = proportion, pop = population, PCV = pneumococcal conjugate vaccine*

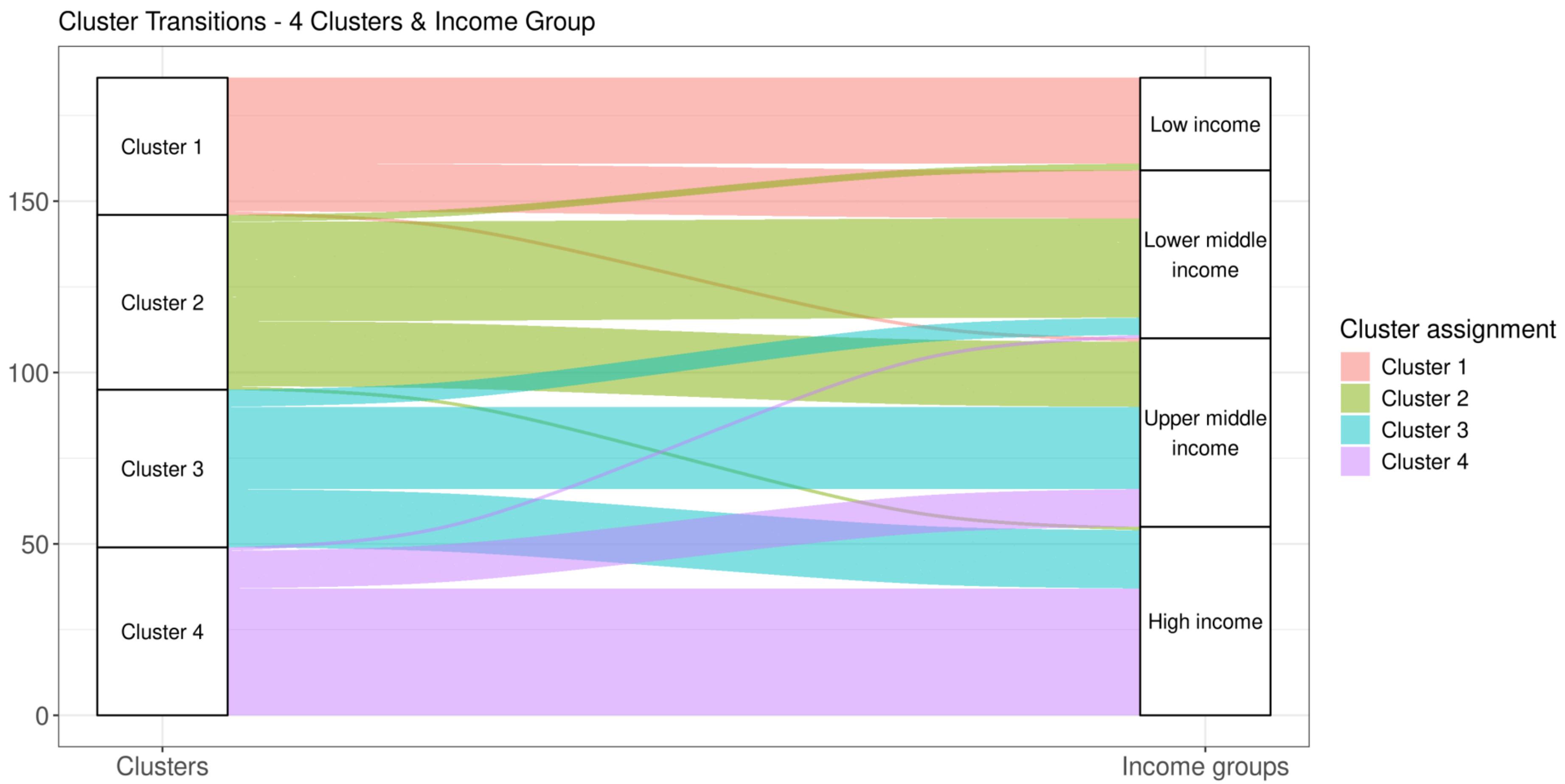

*Figure 19. Comparison of cluster assignments to World Bank Income Groups.*

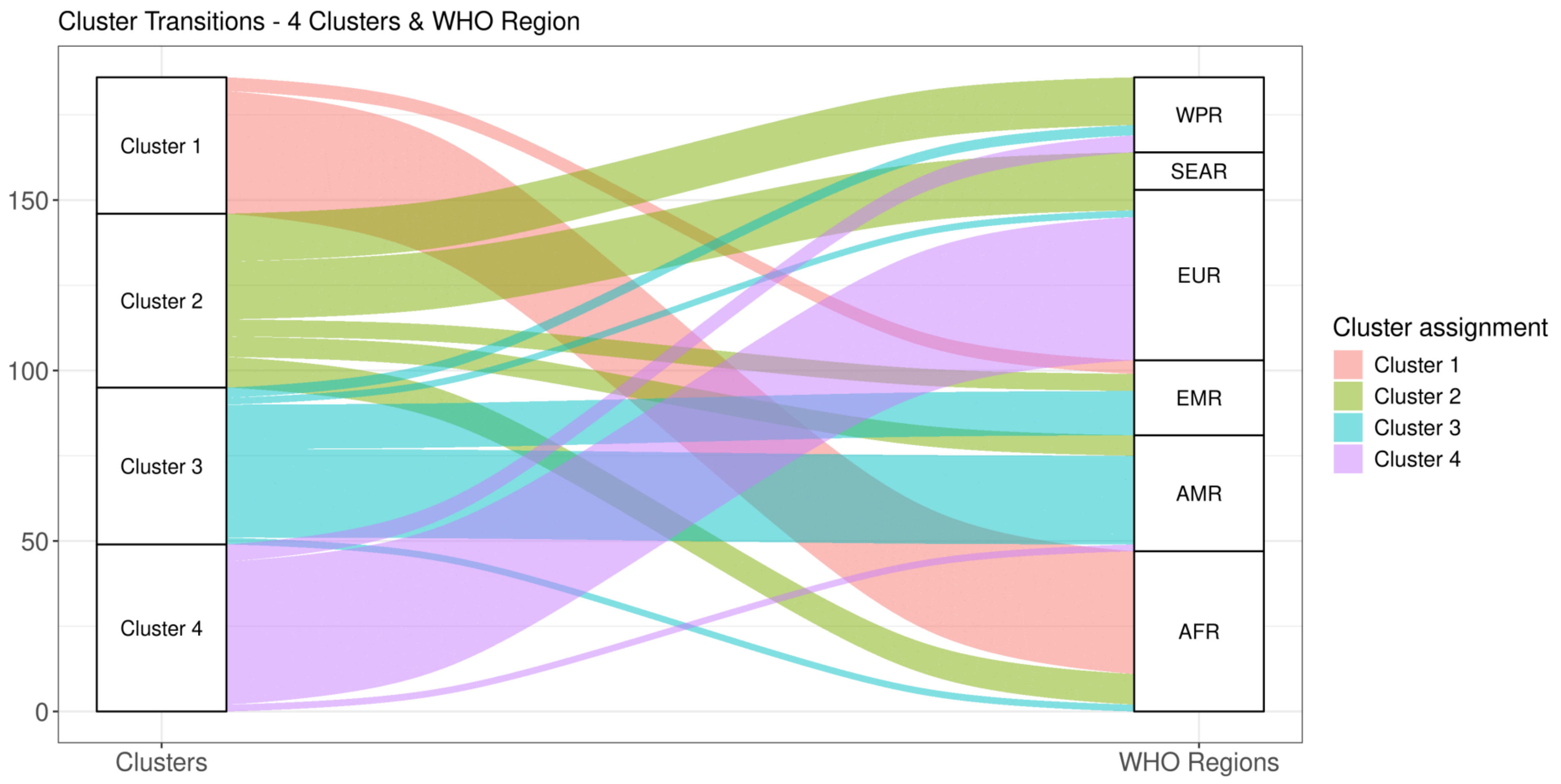

*Figure 20. Comparison of cluster assignments to WHO regions .*

##### 2.2.2 Benchmark CTA selection

Based on our criteria for selecting benchmark CTAs of low total DID with low infection mortality and at least 60% Access antibiotic use (the highest percentage Access where possible) and with low use of "Not recommended" antibiotics, ideally 0, we identified a clear benchmark CTA in cluster 4 (Switzerland). There were no suitable benchmark CTAs in clusters 1 and 2 as no CTAs in cluster 1 had antibiotic data in MIDAS and cluster 2 CTAs had higher infection mortality than CTAs in cluster 3 and low use of Access antibiotics. There were two possible CTAs that could be benchmark CTAs in cluster 3 (which would also be minimum benchmark CTAs for clusters 1 and 2): Morocco and Colombia with slight differences between them (Figure 21). We use Morocco as the benchmark CTA in primary analyses and scenarios 2 and 3 and we used Colombia in scenario 4 to demonstrate the influence of the chosen benchmark CTA on the estimates of expected optimal antibiotic use. We do not use Colombia as the benchmark CTA in primary analyses because they also report data to GLASS AMU which shows higher DID than in MIDAS (19.8 vs. 12.9(95%CI: 11.4-14.5)) and an unusually high use of parenteral antibiotics (23.8% vs. median 5.6%).[75] Table 10 summarises the characteristics of the chosen benchmark CTAs.

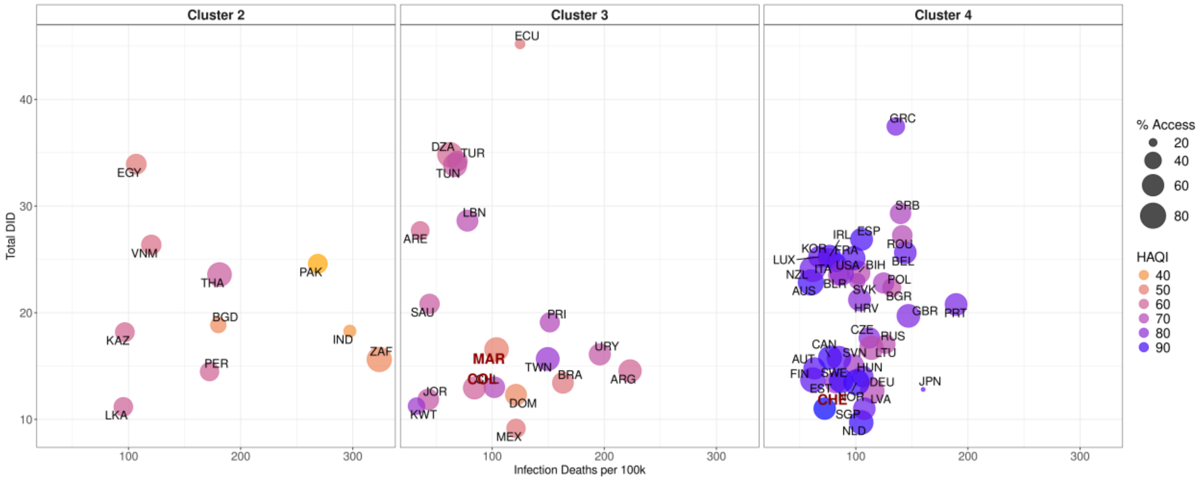

*Figure 21. Infection deaths per 100,000 people and Total DID by cluster for CTAs with data available in IQVIA MIDAS® showing percentage Access antibiotics (size) and Healthcare Access and Quality Index (HAQI) (colour) with potential benchmark CTA codes highlighted in red (MAR (Morocco), COL (Colombia) in benchmark group 3 and CHE (Switzerland) in benchmark group 4). Source: Total DID based on IQVIA MIDAS® data for 2019, reflecting estimates of real-world activity. Copyright IQVIA. All Rights Reserved*

*Table 10. Table of characteristics for considered benchmark CTAs. Benchmark CTA selected from cluster 3 is used as the benchmark CTA to estimate minimum antibiotic need in clusters 1 and 2. Colombia is used only in a sensitivity analysis (scenario 4).*

| Clusters | CTA | Total DID (median)* | Infection deaths per 100,000 (median) | Access percent (median)* | Not Recommended DID (median)* | HAQI (mean) |
|---|---|---|---|---|---|---|
| Cluster 3 | Morocco | 16.6 | 103.9 | 71% | 1.1 | 48.5 |
| Cluster 4 | Switzerland | 11.0 | 72.3 | 61% | 0.05 | 92.6 |
| Cluster 3 (sensitivity analyses – scenario 4) | Colombia | 12.9 | 84.2 | 64% | 0.03 | 61.1 |
| *Source: Total DID based on IQVIA MIDAS® data for 2019, reflecting estimates of real-world activity. Copyright IQVIA. All Rights Reserved | | | | | | |

##### 2.2.3 Estimated expected antibiotic use

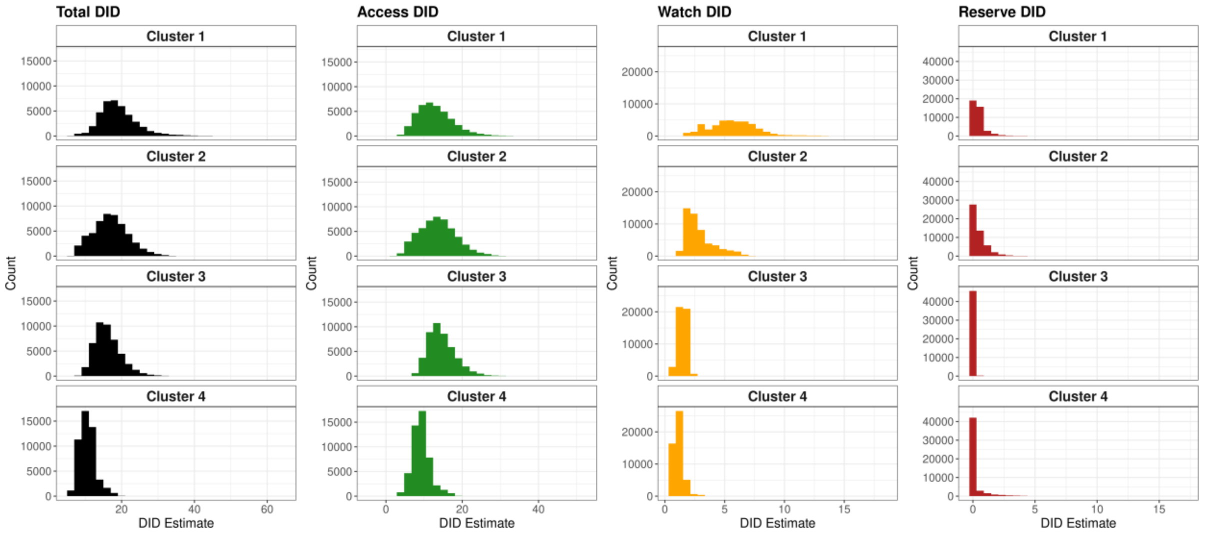

*Figure 22. Distribution of the 1000 draws of estimated expected Total, Access, Watch and Reserve (AWaRe) antibiotic use by cluster for primary analyses. Note: x-axis scales are different between the AWaRe categories.*

##### 2.2.4 Actual vs. estimated optimal antibiotic use

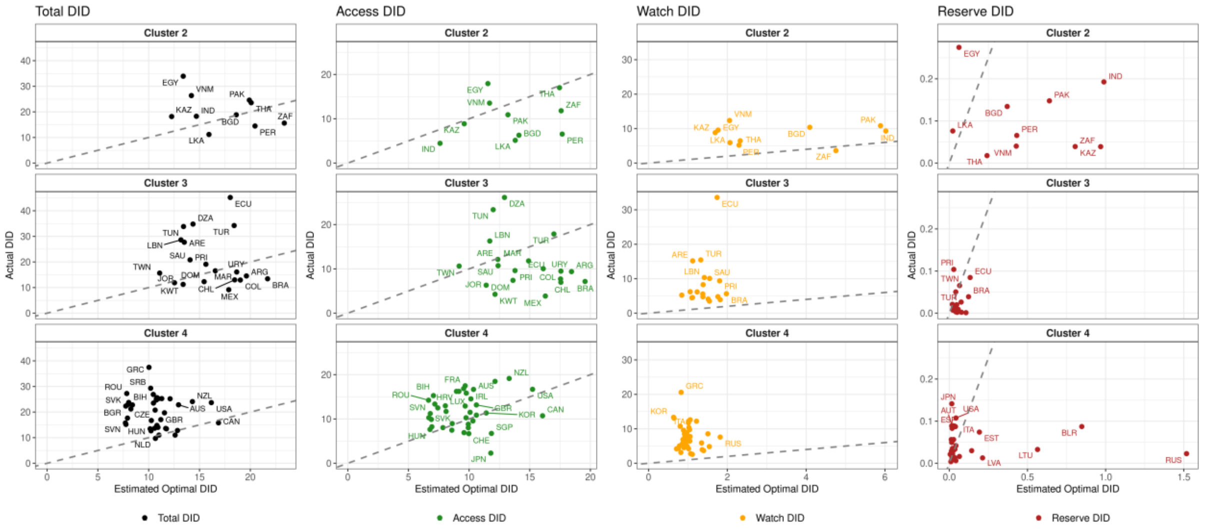

*Figure 23. Median observed (IQVIA MIDAS®) vs. median estimated expected antibiotic use by AWaRe category and benchmark group for primary analyses for CTAs in IQVIA MIDAS® with observed antibiotic use data available. Dashed line indicates where observed = expected. Source: Actual DID based on IQVIA MIDAS® data for 2019, reflecting estimates of real-world activity. Copyright IQVIA. All Rights Reserved*

#### 2.3 Sensitivity analyses

##### 2.3.1 Global antibiotic use

*Table 11. Estimated expected global antibiotic use for Total, Access, Watch and Reserve in billions of defined daily doses (DDD) and DDD/1000 inhabitants/day (DID), and estimated expected global percentage of Access antibiotic use for each analysis scenario.*

| Expected Antibiotic Use | Scenario 1 (Primary analysis) | Scenario 2 (High Watch) | Scenario 3 (Unadjusted cases & excl. TB) | Scenario 4 (Alternate benchmark CTA) |
|---|---|---|---|---|
| Total DDD (billions) | 43 (35.4, 57.7) | 43 (35.4, 57.7) | 42.8 (35.2, 57.4) | 31 (25.5, 44.3) |
| Access DDD (billions) | 32.9 (25.2, 47.3) | 28.5 (20.7, 43.1) | 36.3 (28.6, 50.7) | 20.9 (15.1, 34.1) |
| Watch DDD (billions) | 8.9 (8.4, 9.5) | 13.3 (12.5, 14.1) | 6.2 (5.8, 6.7) | 8.9 (8.4, 9.5) |
| Reserve DDD (billions) | 1.2 (0.6, 2.3) | 1.2 (0.6, 2.3) | 0.3 (0.2, 0.6) | 1.2 (0.6, 2.3) |
| Total DID | 15.2 (12.5, 20) | 15.3 (12.5, 20) | 15.2 (12.5, 20) | 11 (9, 16) |
| Access DID | 11.7 (8.9, 17) | 10.1 (7.3, 15) | 12.9 (10.1, 18) | 7.4 (5.3, 12) |
| Watch DID | 3.2 (3, 3) | 4.7 (4.4, 5) | 2.2 (2.1, 2) | 3.2 (3, 3) |
| Reserve DID | 0.4 (0.2, 1) | 0.4 (0.2, 1) | 0.1 (0.1, 0) | 0.4 (0.2, 1) |
| Access % | 76 (71, 83) | 66 (58, 75) | 85 (81, 89) | 67 (59, 77) |

##### 2.3.2 CTA-level scenario comparisons

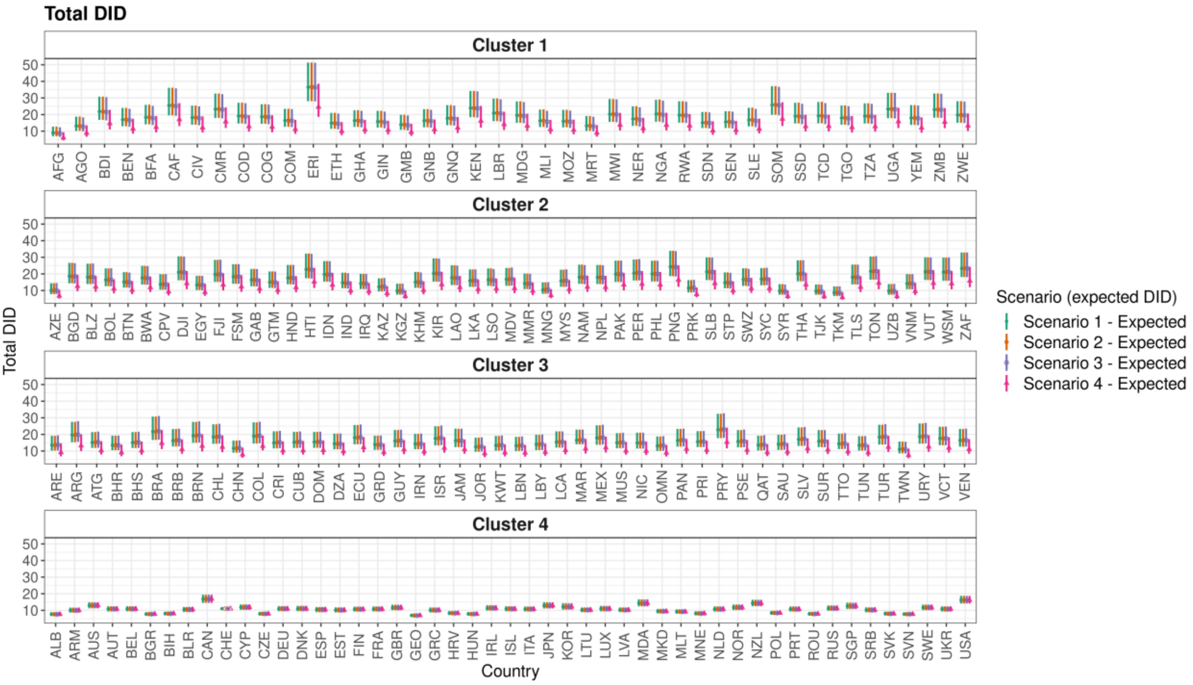

*Figure 24. Estimated optimal Total defined daily doses/1000 inhabitants/year (DID) by CTA and cluster assignment for each analysis scenario. Scenario 1 = primary analysis, Scenario 2 = high watch scenario, Scenario 3 = unadjusted cases & excl. TB, Scenario 4 = alternate benchmark CTA*

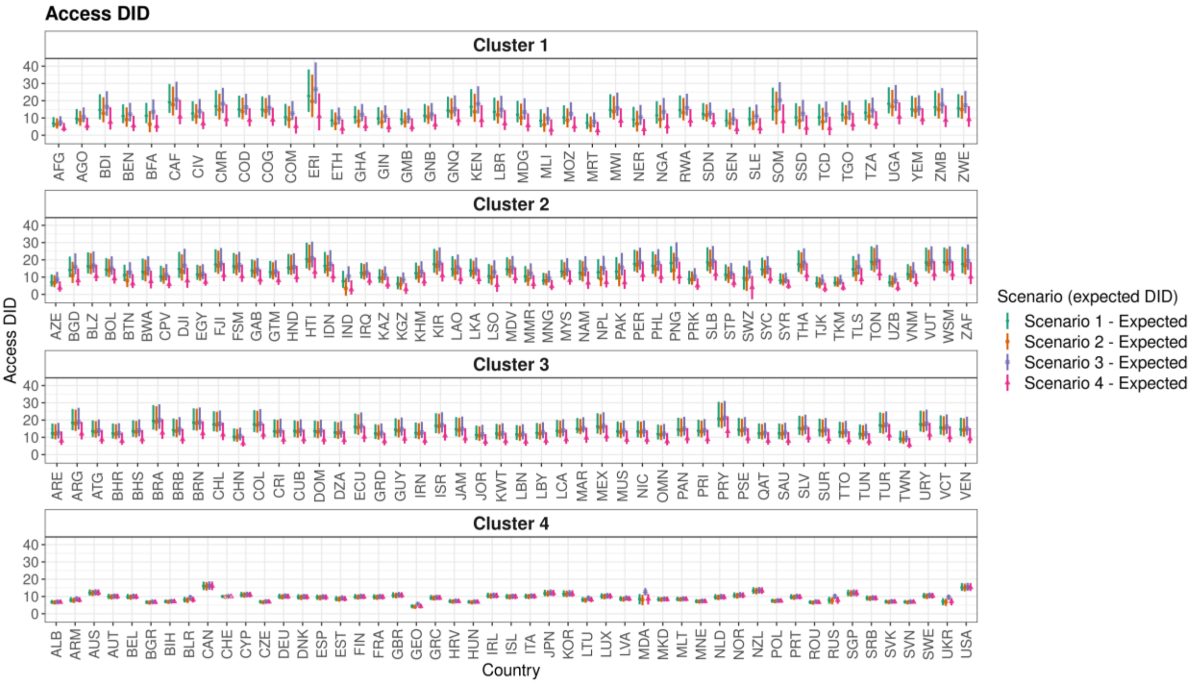

*Figure 25. Estimated optimal Access defined daily doses/1000 inhabitants/year (DID) by CTA and cluster assignment for each analysis scenario. Scenario 1 = primary analysis, Scenario 2 = high watch scenario, Scenario 3 = unadjusted cases & excl. TB, Scenario 4 = alternate benchmark CTA*

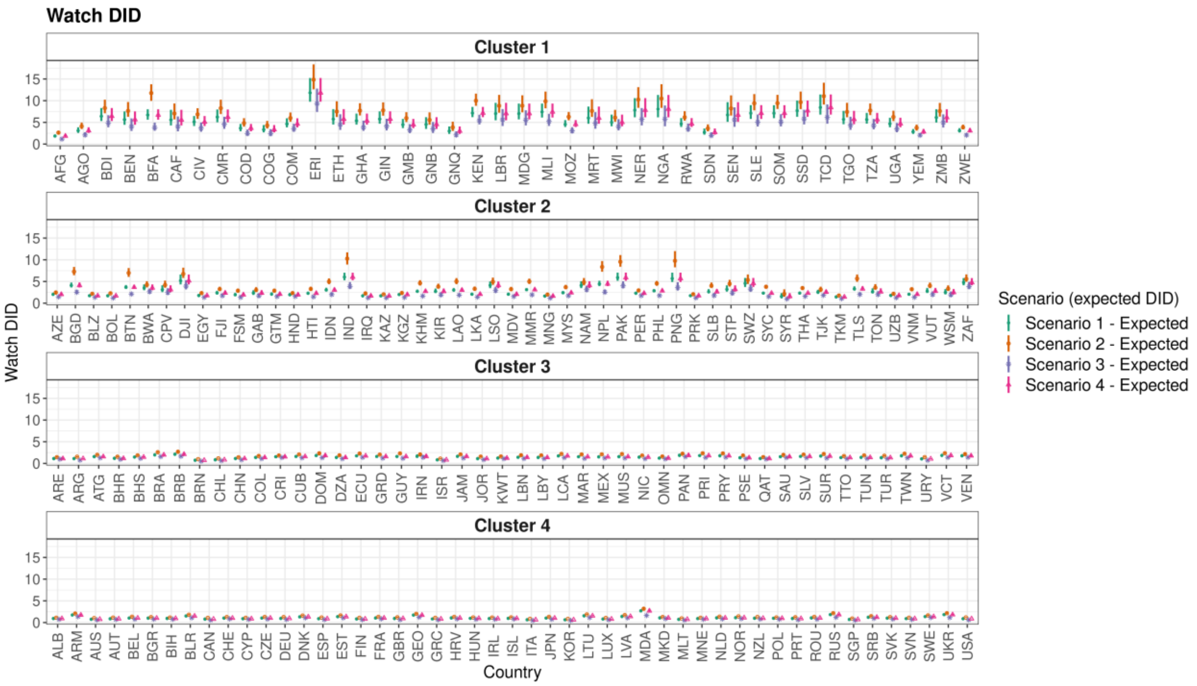

*Figure 26. Estimated optimal Watch defined daily doses/1000 inhabitants/year (DID) by CTA and cluster assignment for each analysis scenario. Scenario 1 = primary analysis, Scenario 2 = high watch scenario, Scenario 3 = unadjusted cases & excl. TB, Scenario 4 = alternate benchmark CTA*

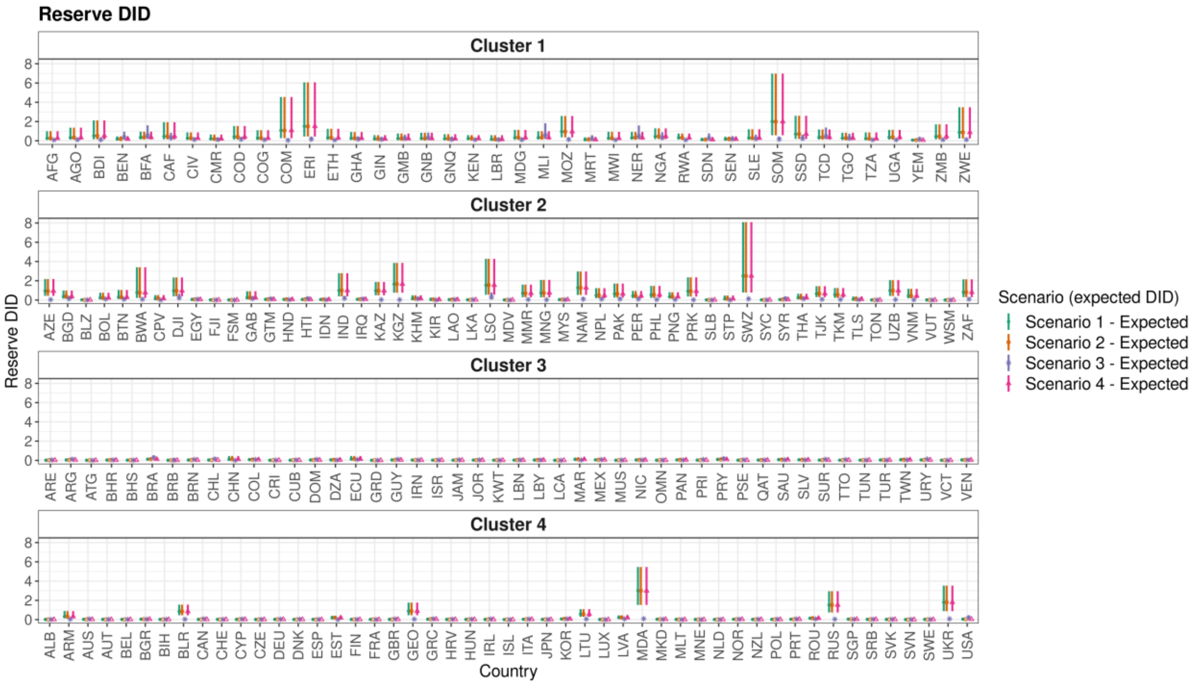

*Figure 27 Estimated optimal Reserve defined daily doses/1000 inhabitants/year (DID) by CTA and cluster assignment for each analysis scenario. Scenario 1 = primary analysis, Scenario 2 = high watch scenario, Scenario 3 = unadjusted cases & excl. TB, Scenario 4 = alternate benchmark CTA*

*Table 12. Estimated optimal total defined daily doses/1000 inhabitants/year (DID) and Access, Watch and Reserve DID for all four analysis scenarios. Scenario 1 = primary analysis, Scenario 2 = high watch scenario, Scenario 3 = unadjusted cases & excl. TB, Scenario 4 = alternate benchmark CTA*

| CTA Name | ISO3 Code | Cluster | WHO Region | Income Group | AWaRe Category | Scenario 1 | Scenario 2 | Scenario 3 | Scenario 4 |
|---|---|---|---|---|---|---|---|---|---|
| **Afghanistan** | AFG | Cluster 1 | EMR | Low income | Total DID | 9 (6.9, 12.8) | 9 (6.9, 12.8) | 8.7 (6.6, 12.4) | 6.1 (4.5, 9.4) |
| **Afghanistan** | AFG | Cluster 1 | EMR | Low income | Access DID | 6.8 (4.6, 10.6) | 6 (3.8, 9.8) | 7.4 (5.3, 11) | 3.9 (2.2, 7.1) |
| **Afghanistan** | AFG | Cluster 1 | EMR | Low income | Watch DID | 1.9 (1.6, 2.2) | 2.6 (2.3, 3.1) | 1.2 (1, 1.5) | 1.9 (1.6, 2.2) |
| **Afghanistan** | AFG | Cluster 1 | EMR | Low income | Reserve DID | 0.3 (0.1, 1) | 0.3 (0.1, 1) | 0.1 (0, 0.2) | 0.3 (0.1, 1) |
| **Albania** | ALB | Cluster 4 | EUR | Upper middle income | Total DID | 7.7 (6.5, 8.9) | 7.7 (6.5, 8.9) | 7.6 (6.5, 8.9) | 7.7 (6.5, 8.9) |
| **Albania** | ALB | Cluster 4 | EUR | Upper middle income | Access DID | 6.7 (5.7, 8) | 6.5 (5.5, 7.8) | 6.8 (5.7, 8.1) | 6.7 (5.7, 8) |
| **Albania** | ALB | Cluster 4 | EUR | Upper middle income | Watch DID | 0.9 (0.8, 1.1) | 1.1 (1, 1.3) | 0.7 (0.6, 0.9) | 0.9 (0.8, 1.1) |
| **Albania** | ALB | Cluster 4 | EUR | Upper middle income | Reserve DID | 0 (0, 0) | 0 (0, 0) | 0 (0, 0.1) | 0 (0, 0) |
| **Algeria** | DZA | Cluster 3 | AFR | Upper middle income | Total DID | 14.4 (11, 20.5) | 14.4 (11, 20.5) | 14.4 (11, 20.6) | 9.8 (7.3, 15) |
| **Algeria** | DZA | Cluster 3 | AFR | Upper middle income | Access DID | 12.9 (9.6, 19.2) | 12.4 (9.1, 18.7) | 13.2 (9.9, 19.5) | 8.3 (5.9, 13.6) |
| **Algeria** | DZA | Cluster 3 | AFR | Upper middle income | Watch DID | 1.4 (1.2, 1.7) | 1.9 (1.6, 2.2) | 1.1 (0.9, 1.4) | 1.4 (1.2, 1.7) |
| **Algeria** | DZA | Cluster 3 | AFR | Upper middle income | Reserve DID | 0.1 (0, 0.2) | 0.1 (0, 0.2) | 0 (0, 0.1) | 0.1 (0, 0.2) |
| **Angola** | AGO | Cluster 1 | AFR | Lower middle income | Total DID | 13.1 (10.1, 18.8) | 13.2 (10.2, 18.8) | 12.8 (9.9, 18.3) | 8.8 (6.7, 13.6) |
| **Angola** | AGO | Cluster 1 | AFR | Lower middle income | Access DID | 9.6 (6.4, 15.1) | 8.6 (5.5, 14.2) | 10.6 (7.5, 16.1) | 5.4 (2.8, 10.2) |
| **Angola** | AGO | Cluster 1 | AFR | Lower middle income | Watch DID | 3.1 (2.6, 3.9) | 4.1 (3.5, 5) | 2.1 (1.7, 2.7) | 3.1 (2.6, 3.9) |
| **Angola** | AGO | Cluster 1 | AFR | Lower middle income | Reserve DID | 0.3 (0.1, 1.4) | 0.3 (0.1, 1.4) | 0.1 (0, 0.3) | 0.3 (0.1, 1.4) |
| **Antigua and Barbuda** | ATG | Cluster 3 | AMR | High income | Total DID | 15.2 (11.9, 21.4) | 15.2 (11.9, 21.4) | 15.4 (12, 21.7) | 10.3 (7.9, 15.9) |
| **Antigua and Barbuda** | ATG | Cluster 3 | AMR | High income | Access DID | 13.6 (10.2, 19.8) | 13.2 (9.8, 19.4) | 14 (10.7, 20.4) | 8.7 (6.3, 14.3) |
| **Antigua and Barbuda** | ATG | Cluster 3 | AMR | High income | Watch DID | 1.6 (1.4, 1.8) | 2 (1.8, 2.2) | 1.3 (1.1, 1.5) | 1.6 (1.4, 1.8) |
| **Antigua and Barbuda** | ATG | Cluster 3 | AMR | High income | Reserve DID | 0 (0, 0) | 0 (0, 0) | 0 (0, 0) | 0 (0, 0) |
| **Argentina** | ARG | Cluster 3 | AMR | Upper middle income | Total DID | 19.6 (15.3, 27.6) | 19.6 (15.3, 27.6) | 19.9 (15.5, 28) | 13.2 (9.9, 20.1) |
| **Argentina** | ARG | Cluster 3 | AMR | Upper middle income | Access DID | 18.4 (14.1, 26.5) | 18 (13.7, 26.1) | 18.9 (14.5, 27.1) | 12 (8.8, 18.9) |

| | | | | | | | | | |
|---|---|---|---|---|---|---|---|---|---|
| **Argentina** | ARG | Cluster 3 | AMR | Upper middle income | Watch DID | 1.1 (1, 1.2) | 1.5 (1.4, 1.7) | 0.8 (0.7, 0.9) | 1.1 (1, 1.2) |
| **Argentina** | ARG | Cluster 3 | AMR | Upper middle income | Reserve DID | 0 (0, 0.1) | 0 (0, 0.1) | 0.1 (0.1, 0.3) | 0 (0, 0.1) |
| **Armenia** | ARM | Cluster 4 | EUR | Upper middle income | Total DID | 10 (8.7, 11.7) | 10 (8.7, 11.7) | 9.9 (8.6, 11.6) | 10 (8.7, 11.7) |
| **Armenia** | ARM | Cluster 4 | EUR | Upper middle income | Access DID | 7.9 (6.5, 9.6) | 7.6 (6.2, 9.3) | 8.5 (7.2, 10.1) | 7.9 (6.5, 9.6) |
| **Armenia** | ARM | Cluster 4 | EUR | Upper middle income | Watch DID | 1.7 (1.5, 2) | 2.1 (1.9, 2.3) | 1.4 (1.2, 1.6) | 1.7 (1.5, 2) |
| **Armenia** | ARM | Cluster 4 | EUR | Upper middle income | Reserve DID | 0.3 (0.1, 0.9) | 0.3 (0.1, 0.9) | 0 (0, 0.1) | 0.3 (0.1, 0.9) |
| **Australia** | AUS | Cluster 4 | WPR | High income | Total DID | 12.9 (11.2, 14.9) | 12.9 (11.2, 14.9) | 12.9 (11.2, 14.9) | 12.9 (11.2, 14.9) |
| **Australia** | AUS | Cluster 4 | WPR | High income | Access DID | 12.1 (10.4, 14.1) | 11.9 (10.2, 13.8) | 12.3 (10.6, 14.2) | 12.1 (10.4, 14.1) |
| **Australia** | AUS | Cluster 4 | WPR | High income | Watch DID | 0.8 (0.7, 0.8) | 1 (0.9, 1.1) | 0.6 (0.5, 0.7) | 0.8 (0.7, 0.8) |
| **Australia** | AUS | Cluster 4 | WPR | High income | Reserve DID | 0 (0, 0) | 0 (0, 0) | 0.1 (0, 0.1) | 0 (0, 0) |
| **Austria** | AUT | Cluster 4 | EUR | High income | Total DID | 10.8 (9.3, 12.4) | 10.8 (9.3, 12.4) | 10.8 (9.3, 12.5) | 10.8 (9.3, 12.4) |
| **Austria** | AUT | Cluster 4 | EUR | High income | Access DID | 9.9 (8.5, 11.6) | 9.7 (8.2, 11.3) | 10.1 (8.6, 11.7) | 9.9 (8.5, 11.6) |
| **Austria** | AUT | Cluster 4 | EUR | High income | Watch DID | 0.9 (0.8, 1) | 1.1 (1, 1.3) | 0.7 (0.6, 0.8) | 0.9 (0.8, 1) |
| **Austria** | AUT | Cluster 4 | EUR | High income | Reserve DID | 0 (0, 0) | 0 (0, 0) | 0 (0, 0) | 0 (0, 0) |
| **Azerbaijan** | AZE | Cluster 2 | EUR | Upper middle income | Total DID | 10.1 (7.8, 14.4) | 10.1 (7.8, 14.4) | 10.1 (7.8, 14.5) | 6.8 (5.2, 10.6) |
| **Azerbaijan** | AZE | Cluster 2 | EUR | Upper middle income | Access DID | 7 (4.5, 11.5) | 6.6 (4.1, 11.1) | 8.6 (6.3, 13) | 3.8 (1.5, 7.5) |
| **Azerbaijan** | AZE | Cluster 2 | EUR | Upper middle income | Watch DID | 2 (1.8, 2.4) | 2.5 (2.2, 2.8) | 1.5 (1.3, 1.7) | 2 (1.8, 2.4) |
| **Azerbaijan** | AZE | Cluster 2 | EUR | Upper middle income | Reserve DID | 0.9 (0.4, 2.2) | 0.9 (0.4, 2.2) | 0 (0, 0.1) | 0.9 (0.4, 2.2) |
| **Bahamas, The** | BHS | Cluster 3 | AMR | High income | Total DID | 15.1 (11.8, 21.4) | 15.1 (11.8, 21.4) | 15.2 (11.9, 21.6) | 10.2 (7.7, 15.7) |
| **Bahamas, The** | BHS | Cluster 3 | AMR | High income | Access DID | 13.6 (10.3, 19.9) | 13.2 (9.9, 19.5) | 14 (10.6, 20.4) | 8.7 (6.2, 14.3) |
| **Bahamas, The** | BHS | Cluster 3 | AMR | High income | Watch DID | 1.5 (1.3, 1.7) | 1.9 (1.7, 2.1) | 1.2 (1, 1.4) | 1.5 (1.3, 1.7) |
| **Bahamas, The** | BHS | Cluster 3 | AMR | High income | Reserve DID | 0 (0, 0.1) | 0 (0, 0.1) | 0 (0, 0.2) | 0 (0, 0.1) |
| **Bahrain** | BHR | Cluster 3 | EMR | High income | Total DID | 13.5 (10.5, 19.2) | 13.5 (10.5, 19.2) | 13.5 (10.5, 19.3) | 9.1 (6.9, 13.9) |
| **Bahrain** | BHR | Cluster 3 | EMR | High income | Access DID | 12.2 (9.3, 18) | 11.9 (8.9, 17.6) | 12.4 (9.5, 18.2) | 7.9 (5.7, 12.8) |
| **Bahrain** | BHR | Cluster 3 | EMR | High income | Watch DID | 1.2 (1, 1.4) | 1.5 (1.4, 1.8) | 1 (0.8, 1.2) | 1.2 (1, 1.4) |

| | | | | | | | | | |
|---|---|---|---|---|---|---|---|---|---|
| **Bahrain** | BHR | Cluster 3 | EMR | High income | Reserve DID | 0 (0, 0.1) | 0 (0, 0.1) | 0.1 (0, 0.1) | 0 (0, 0.1) |
| **Bangladesh** | BGD | Cluster 2 | SEAR | Lower middle income | Total DID | 18.6 (14.4, 26.7) | 18.6 (14.4, 26.7) | 18.5 (14.2, 26.5) | 12.6 (9.6, 19.4) |
| **Bangladesh** | BGD | Cluster 2 | SEAR | Lower middle income | Access DID | 14.1 (9.9, 21.9) | 10.9 (6.6, 18.9) | 15.8 (11.6, 23.8) | 8.1 (5.1, 15) |
| **Bangladesh** | BGD | Cluster 2 | SEAR | Lower middle income | Watch DID | 4.1 (3.6, 4.8) | 7.3 (6.4, 8.3) | 2.5 (2.2, 3.1) | 4.1 (3.6, 4.8) |
| **Bangladesh** | BGD | Cluster 2 | SEAR | Lower middle income | Reserve DID | 0.4 (0.1, 1) | 0.4 (0.1, 1) | 0.1 (0, 0.4) | 0.4 (0.1, 1) |
| **Barbados** | BRB | Cluster 3 | AMR | High income | Total DID | 16.3 (12.7, 23.2) | 16.3 (12.7, 23.3) | 16.4 (12.8, 23.4) | 11 (8.3, 17) |
| **Barbados** | BRB | Cluster 3 | AMR | High income | Access DID | 14.2 (10.6, 21) | 13.6 (10, 20.4) | 14.7 (11.1, 21.7) | 8.8 (6.3, 15) |
| **Barbados** | BRB | Cluster 3 | AMR | High income | Watch DID | 2.1 (1.9, 2.4) | 2.7 (2.4, 3) | 1.7 (1.5, 1.9) | 2.1 (1.9, 2.4) |
| **Barbados** | BRB | Cluster 3 | AMR | High income | Reserve DID | 0 (0, 0.1) | 0 (0, 0.1) | 0 (0, 0.3) | 0 (0, 0.1) |
| **Belarus** | BLR | Cluster 4 | EUR | Upper middle income | Total DID | 10.5 (9, 12.1) | 10.5 (9, 12.1) | 10.5 (9, 12) | 10.5 (9, 12.1) |
| **Belarus** | BLR | Cluster 4 | EUR | Upper middle income | Access DID | 8 (6.5, 9.8) | 7.7 (6.2, 9.5) | 9.3 (7.9, 10.9) | 8 (6.5, 9.8) |
| **Belarus** | BLR | Cluster 4 | EUR | Upper middle income | Watch DID | 1.5 (1.4, 1.7) | 1.8 (1.6, 2) | 1.1 (1, 1.2) | 1.5 (1.4, 1.7) |
| **Belarus** | BLR | Cluster 4 | EUR | Upper middle income | Reserve DID | 0.8 (0.5, 1.6) | 0.8 (0.5, 1.6) | 0 (0, 0.1) | 0.8 (0.5, 1.6) |
| **Belgium** | BEL | Cluster 4 | EUR | High income | Total DID | 10.8 (9.4, 12.5) | 10.8 (9.4, 12.5) | 10.8 (9.4, 12.5) | 10.8 (9.4, 12.5) |
| **Belgium** | BEL | Cluster 4 | EUR | High income | Access DID | 9.8 (8.4, 11.5) | 9.5 (8.1, 11.2) | 10 (8.6, 11.7) | 9.8 (8.4, 11.5) |
| **Belgium** | BEL | Cluster 4 | EUR | High income | Watch DID | 1 (0.9, 1.2) | 1.3 (1.2, 1.5) | 0.8 (0.7, 0.9) | 1 (0.9, 1.2) |
| **Belgium** | BEL | Cluster 4 | EUR | High income | Reserve DID | 0 (0, 0) | 0 (0, 0) | 0 (0, 0.1) | 0 (0, 0) |
| **Belize** | BLZ | Cluster 2 | AMR | Upper middle income | Total DID | 18 (13.9, 26.2) | 18 (13.9, 26.2) | 18.1 (13.9, 26.3) | 12.2 (9.3, 18.9) |
| **Belize** | BLZ | Cluster 2 | AMR | Upper middle income | Access DID | 16.3 (12.1, 24.3) | 15.8 (11.7, 23.9) | 16.7 (12.5, 24.9) | 10.5 (7.6, 17.2) |
| **Belize** | BLZ | Cluster 2 | AMR | Upper middle income | Watch DID | 1.7 (1.5, 2) | 2.2 (1.9, 2.4) | 1.4 (1.2, 1.6) | 1.7 (1.5, 2) |
| **Belize** | BLZ | Cluster 2 | AMR | Upper middle income | Reserve DID | 0 (0, 0.1) | 0 (0, 0.1) | 0 (0, 0.1) | 0 (0, 0.1) |
| **Benin** | BEN | Cluster 1 | AFR | Lower middle income | Total DID | 16.9 (13, 24) | 17 (13, 24.1) | 16.6 (12.6, 23.5) | 11.4 (8.5, 17.5) |
| **Benin** | BEN | Cluster 1 | AFR | Lower middle income | Access DID | 11.1 (7, 18) | 9.1 (4.9, 16.1) | 12.1 (8, 18.8) | 5.5 (2.4, 11.3) |
| **Benin** | BEN | Cluster 1 | AFR | Lower middle income | Watch DID | 5.6 (4.4, 7.5) | 7.6 (6.3, 9.7) | 4 (3, 5.9) | 5.6 (4.4, 7.5) |

| | | | | | | | | | |
|---|---|---|---|---|---|---|---|---|---|
| **Benin** | BEN | Cluster 1 | AFR | Lower middle income | Reserve DID | 0.2 (0.1, 0.5) | 0.2 (0.1, 0.5) | 0.4 (0.1, 0.9) | 0.2 (0.1, 0.5) |
| **Bhutan** | BTN | Cluster 2 | SEAR | Lower middle income | Total DID | 15.1 (11.8, 21.1) | 15.2 (11.8, 21.1) | 14.9 (11.6, 20.8) | 10.3 (7.8, 15.7) |
| **Bhutan** | BTN | Cluster 2 | SEAR | Lower middle income | Access DID | 11.2 (7.6, 17.2) | 8 (4.2, 13.8) | 12.8 (9.3, 18.7) | 6.2 (3.6, 11.8) |
| **Bhutan** | BTN | Cluster 2 | SEAR | Lower middle income | Watch DID | 3.7 (3.3, 4.2) | 7 (6.1, 8.1) | 2.1 (1.9, 2.5) | 3.7 (3.3, 4.2) |
| **Bhutan** | BTN | Cluster 2 | SEAR | Lower middle income | Reserve DID | 0.3 (0.1, 1) | 0.3 (0.1, 1) | 0.1 (0, 0.3) | 0.3 (0.1, 1) |
| **Bolivia** | BOL | Cluster 2 | AMR | Lower middle income | Total DID | 16.4 (12.6, 23.3) | 16.4 (12.6, 23.3) | 16.4 (12.6, 23.4) | 11.1 (8.3, 16.8) |
| **Bolivia** | BOL | Cluster 2 | AMR | Lower middle income | Access DID | 14.4 (10.5, 21) | 13.9 (10, 20.5) | 15.1 (11.3, 22) | 9.1 (6.2, 14.9) |
| **Bolivia** | BOL | Cluster 2 | AMR | Lower middle income | Watch DID | 1.7 (1.5, 1.9) | 2.3 (2, 2.6) | 1.2 (1.1, 1.4) | 1.7 (1.5, 1.9) |
| **Bolivia** | BOL | Cluster 2 | AMR | Lower middle income | Reserve DID | 0.2 (0.1, 0.8) | 0.2 (0.1, 0.8) | 0.1 (0, 0.2) | 0.2 (0.1, 0.8) |
| **Bosnia and Herzegovina** | BIH | Cluster 4 | EUR | Upper middle income | Total DID | 8 (7, 9.2) | 8 (7, 9.2) | 8 (6.9, 9.1) | 8 (7, 9.2) |
| **Bosnia and Herzegovina** | BIH | Cluster 4 | EUR | Upper middle income | Access DID | 7.1 (6, 8.2) | 6.9 (5.8, 8) | 7.2 (6.1, 8.4) | 7.1 (6, 8.2) |
| **Bosnia and Herzegovina** | BIH | Cluster 4 | EUR | Upper middle income | Watch DID | 1 (0.8, 1.1) | 1.1 (1, 1.3) | 0.8 (0.7, 0.9) | 1 (0.8, 1.1) |
| **Bosnia and Herzegovina** | BIH | Cluster 4 | EUR | Upper middle income | Reserve DID | 0 (0, 0) | 0 (0, 0) | 0 (0, 0.1) | 0 (0, 0) |
| **Botswana** | BWA | Cluster 2 | AFR | Upper middle income | Total DID | 17.6 (13.6, 24.9) | 17.6 (13.6, 24.9) | 17.5 (13.4, 24.7) | 11.9 (9.1, 18.4) |
| **Botswana** | BWA | Cluster 2 | AFR | Upper middle income | Access DID | 13.1 (8.2, 20.8) | 12.4 (7.5, 20.1) | 14.7 (10.7, 22.1) | 7.5 (3.4, 13.8) |
| **Botswana** | BWA | Cluster 2 | AFR | Upper middle income | Watch DID | 3.5 (3, 4.3) | 4.2 (3.6, 5.1) | 2.6 (2.2, 3.4) | 3.5 (3, 4.3) |
| **Botswana** | BWA | Cluster 2 | AFR | Upper middle income | Reserve DID | 0.8 (0.2, 3.4) | 0.8 (0.2, 3.4) | 0.1 (0, 0.2) | 0.8 (0.2, 3.4) |
| **Brazil** | BRA | Cluster 3 | AMR | Upper middle income | Total DID | 21.7 (16.6, 30.7) | 21.7 (16.6, 30.8) | 21.9 (16.8, 31.1) | 14.7 (11.2, 22.6) |
| **Brazil** | BRA | Cluster 3 | AMR | Upper middle income | Access DID | 19.6 (14.5, 28.6) | 19 (13.8, 28.1) | 20.1 (14.9, 29.1) | 12.6 (9.1, 20.6) |
| **Brazil** | BRA | Cluster 3 | AMR | Upper middle income | Watch DID | 2 (1.8, 2.2) | 2.6 (2.4, 2.8) | 1.6 (1.4, 1.8) | 2 (1.8, 2.2) |
| **Brazil** | BRA | Cluster 3 | AMR | Upper middle income | Reserve DID | 0.1 (0.1, 0.3) | 0.1 (0.1, 0.3) | 0.3 (0.1, 0.5) | 0.1 (0.1, 0.3) |
| **Brunei Darussalam** | BRN | Cluster 3 | WPR | High income | Total DID | 19.3 (14.9, 27.6) | 19.3 (14.9, 27.5) | 19.5 (15.1, 27.9) | 13 (9.7, 20.1) |

| | | | | | | | | | |
|---|---|---|---|---|---|---|---|---|---|
| **Brunei Darussalam** | BRN | Cluster 3 | WPR | High income | Access DID | 18.5 (14.1, 26.8) | 18.3 (13.9, 26.6) | 18.9 (14.5, 27.3) | 12.2 (9, 19.3) |
| **Brunei Darussalam** | BRN | Cluster 3 | WPR | High income | Watch DID | 0.7 (0.7, 0.8) | 1 (0.9, 1.1) | 0.5 (0.5, 0.6) | 0.7 (0.7, 0.8) |
| **Brunei Darussalam** | BRN | Cluster 3 | WPR | High income | Reserve DID | 0 (0, 0.1) | 0 (0, 0.1) | 0.1 (0.1, 0.2) | 0 (0, 0.1) |
| **Bulgaria** | BGR | Cluster 4 | EUR | Upper middle income | Total DID | 7.7 (6.7, 8.9) | 7.8 (6.7, 8.9) | 7.7 (6.6, 8.9) | 7.7 (6.7, 8.9) |
| **Bulgaria** | BGR | Cluster 4 | EUR | Upper middle income | Access DID | 6.7 (5.6, 7.8) | 6.5 (5.4, 7.6) | 6.9 (5.8, 8) | 6.7 (5.6, 7.8) |
| **Bulgaria** | BGR | Cluster 4 | EUR | Upper middle income | Watch DID | 1 (0.9, 1.2) | 1.2 (1.1, 1.4) | 0.8 (0.7, 0.9) | 1 (0.9, 1.2) |
| **Bulgaria** | BGR | Cluster 4 | EUR | Upper middle income | Reserve DID | 0 (0, 0.1) | 0 (0, 0.1) | 0 (0, 0.1) | 0 (0, 0.1) |
| **Burkina Faso** | BFA | Cluster 1 | AFR | Low income | Total DID | 18.3 (14.1, 25.9) | 18.4 (14.2, 26) | 17.8 (13.7, 25.3) | 12.4 (9.3, 19.1) |
| **Burkina Faso** | BFA | Cluster 1 | AFR | Low income | Access DID | 11.3 (7, 18.8) | 6.4 (1.8, 14.1) | 13.5 (9, 20.8) | 5.3 (1.9, 11.8) |
| **Burkina Faso** | BFA | Cluster 1 | AFR | Low income | Watch DID | 6.7 (5.6, 8) | 11.8 (10, 13.8) | 3.8 (3.1, 4.9) | 6.7 (5.6, 8) |
| **Burkina Faso** | BFA | Cluster 1 | AFR | Low income | Reserve DID | 0.4 (0.1, 0.9) | 0.4 (0.1, 0.9) | 0.5 (0.2, 1.6) | 0.4 (0.1, 0.9) |
| **Burundi** | BDI | Cluster 1 | AFR | Low income | Total DID | 21.8 (16.8, 30.8) | 21.8 (16.8, 30.8) | 21.5 (16.5, 30.5) | 14.7 (11.1, 22.9) |
| **Burundi** | BDI | Cluster 1 | AFR | Low income | Access DID | 14.6 (9.2, 23.8) | 12.8 (7.5, 22) | 16.4 (11.3, 25.5) | 7.6 (3.3, 16.2) |
| **Burundi** | BDI | Cluster 1 | AFR | Low income | Watch DID | 6.4 (5.3, 8.3) | 8.3 (7.1, 10.2) | 4.9 (3.9, 6.7) | 6.4 (5.3, 8.3) |
| **Burundi** | BDI | Cluster 1 | AFR | Low income | Reserve DID | 0.5 (0.2, 2.1) | 0.5 (0.2, 2.1) | 0.1 (0, 0.3) | 0.5 (0.2, 2.1) |
| **Cabo Verde** | CPV | Cluster 2 | AFR | Lower middle income | Total DID | 13.7 (10.5, 19.8) | 13.7 (10.5, 19.9) | 13.8 (10.5, 19.9) | 9.3 (7.1, 14.5) |
| **Cabo Verde** | CPV | Cluster 2 | AFR | Lower middle income | Access DID | 10.4 (7.2, 16.7) | 9.3 (6.1, 15.5) | 11.2 (8, 17.5) | 5.9 (3.6, 11) |
| **Cabo Verde** | CPV | Cluster 2 | AFR | Lower middle income | Watch DID | 3.1 (2.5, 4.1) | 4.2 (3.6, 5.2) | 2.5 (1.9, 3.4) | 3.1 (2.5, 4.1) |
| **Cabo Verde** | CPV | Cluster 2 | AFR | Lower middle income | Reserve DID | 0.1 (0, 0.5) | 0.1 (0, 0.5) | 0 (0, 0.1) | 0.1 (0, 0.5) |
| **Cambodia** | KHM | Cluster 2 | WPR | Lower middle income | Total DID | 15.1 (11.6, 21.2) | 15.1 (11.6, 21.2) | 14.9 (11.4, 20.9) | 10.2 (7.7, 15.5) |
| **Cambodia** | KHM | Cluster 2 | WPR | Lower middle income | Access DID | 12.2 (8.5, 18.4) | 10.3 (6.7, 16.6) | 13.1 (9.6, 19.2) | 7.3 (4.8, 12.8) |
| **Cambodia** | KHM | Cluster 2 | WPR | Lower middle income | Watch DID | 2.7 (2.4, 3.1) | 4.6 (4.1, 5.3) | 1.6 (1.4, 2) | 2.7 (2.4, 3.1) |
| **Cambodia** | KHM | Cluster 2 | WPR | Lower middle income | Reserve DID | 0.1 (0.1, 0.5) | 0.1 (0.1, 0.5) | 0.1 (0.1, 0.3) | 0.1 (0.1, 0.5) |
| **Cameroon** | CMR | Cluster 1 | AFR | Lower middle income | Total DID | 23.3 (18, 32.6) | 23.3 (18, 32.7) | 23 (17.7, 32.3) | 15.7 (11.9, 24.3) |

| | | | | | | | | | |
|---|---|---|---|---|---|---|---|---|---|
| **Cameroon** | CMR | Cluster 1 | AFR | Lower middle income | Access DID | 16.8 (11.2, 26.1) | 14.9 (9.1, 24.1) | 18.3 (12.8, 27.5) | 9.2 (5.1, 17.9) |
| **Cameroon** | CMR | Cluster 1 | AFR | Lower middle income | Watch DID | 6.2 (5, 8) | 8.3 (6.9, 10.2) | 4.6 (3.5, 6.3) | 6.2 (5, 8) |
| **Cameroon** | CMR | Cluster 1 | AFR | Lower middle income | Reserve DID | 0.2 (0.1, 0.6) | 0.2 (0.1, 0.6) | 0.1 (0, 0.3) | 0.2 (0.1, 0.6) |
| **Canada** | CAN | Cluster 4 | AMR | High income | Total DID | 16.9 (14.5, 19.4) | 16.9 (14.5, 19.4) | 16.8 (14.5, 19.4) | 16.9 (14.5, 19.4) |
| **Canada** | CAN | Cluster 4 | AMR | High income | Access DID | 16.1 (13.7, 18.6) | 15.8 (13.4, 18.3) | 16.2 (13.8, 18.7) | 16.1 (13.7, 18.6) |
| **Canada** | CAN | Cluster 4 | AMR | High income | Watch DID | 0.8 (0.7, 0.9) | 1.1 (1, 1.2) | 0.6 (0.5, 0.6) | 0.8 (0.7, 0.9) |
| **Canada** | CAN | Cluster 4 | AMR | High income | Reserve DID | 0 (0, 0) | 0 (0, 0) | 0.1 (0, 0.2) | 0 (0, 0) |
| **Central African Republic** | CAF | Cluster 1 | AFR | Low income | Total DID | 25.5 (19.5, 36.2) | 25.5 (19.5, 36.2) | 25.1 (19.2, 35.8) | 17.1 (13.1, 26.9) |
| **Central African Republic** | CAF | Cluster 1 | AFR | Low income | Access DID | 19.1 (12.9, 29.7) | 17.7 (11.4, 28.2) | 20.6 (14.7, 31) | 10.8 (6.4, 20.5) |
| **Central African Republic** | CAF | Cluster 1 | AFR | Low income | Watch DID | 5.6 (4.3, 7.9) | 7 (5.7, 9.4) | 4 (2.9, 6.4) | 5.6 (4.3, 7.9) |
| **Central African Republic** | CAF | Cluster 1 | AFR | Low income | Reserve DID | 0.5 (0.2, 1.9) | 0.5 (0.2, 1.9) | 0.3 (0.1, 0.8) | 0.5 (0.2, 1.9) |
| **Chad** | TCD | Cluster 1 | AFR | Low income | Total DID | 19.2 (14.7, 27.6) | 19.3 (14.8, 27.7) | 18.7 (14.3, 26.9) | 13 (9.8, 19.9) |
| **Chad** | TCD | Cluster 1 | AFR | Low income | Access DID | 10.3 (5.6, 18.1) | 7.8 (3.1, 15.9) | 12 (7.3, 19.6) | 4.1 (0.1, 10.7) |
| **Chad** | TCD | Cluster 1 | AFR | Low income | Watch DID | 8.4 (6.8, 11.4) | 11 (9.1, 14.1) | 6.2 (4.7, 9.1) | 8.4 (6.8, 11.4) |
| **Chad** | TCD | Cluster 1 | AFR | Low income | Reserve DID | 0.4 (0.1, 1.2) | 0.4 (0.1, 1.2) | 0.5 (0.2, 1.4) | 0.4 (0.1, 1.2) |
| **Chile** | CHL | Cluster 3 | AMR | High income | Total DID | 18.5 (14.3, 26.1) | 18.5 (14.3, 26.1) | 18.7 (14.5, 26.4) | 12.4 (9.3, 19.4) |
| **Chile** | CHL | Cluster 3 | AMR | High income | Access DID | 17.6 (13.5, 25.1) | 17.2 (13.2, 24.8) | 17.9 (13.7, 25.6) | 11.5 (8.4, 18.5) |
| **Chile** | CHL | Cluster 3 | AMR | High income | Watch DID | 0.8 (0.8, 0.9) | 1.2 (1.1, 1.3) | 0.6 (0.6, 0.7) | 0.8 (0.8, 0.9) |
| **Chile** | CHL | Cluster 3 | AMR | High income | Reserve DID | 0 (0, 0.1) | 0 (0, 0.1) | 0.1 (0.1, 0.3) | 0 (0, 0.1) |
| **China** | CHN | Cluster 3 | WPR | Upper middle income | Total DID | 11.6 (9, 16.3) | 11.6 (9, 16.3) | 11.7 (9, 16.4) | 7.8 (6, 12.1) |
| **China** | CHN | Cluster 3 | WPR | Upper middle income | Access DID | 10.3 (7.7, 15.2) | 10 (7.4, 14.9) | 10.8 (8.2, 15.6) | 6.5 (4.7, 10.9) |
| **China** | CHN | Cluster 3 | WPR | Upper middle income | Watch DID | 1.1 (1, 1.3) | 1.4 (1.3, 1.6) | 0.8 (0.7, 1) | 1.1 (1, 1.3) |
| **China** | CHN | Cluster 3 | WPR | Upper middle income | Reserve DID | 0.1 (0, 0.5) | 0.1 (0, 0.5) | 0 (0, 0) | 0.1 (0, 0.5) |
| **Colombia** | COL | Cluster 3 | AMR | Upper middle income | Total DID | 19 (14.5, 27.3) | 19 (14.5, 27.3) | 19.2 (14.6, 27.6) | 12.9 (10.4, 19.3) |

| | | | | | | | | | |
|---|---|---|---|---|---|---|---|---|---|
| **Colombia** | COL | Cluster 3 | AMR | Upper middle income | Access DID | 17.5 (13.1, 25.8) | 17.2 (12.7, 25.4) | 18 (13.5, 26.4) | 11.5 (8.9, 17.9) |
| **Colombia** | COL | Cluster 3 | AMR | Upper middle income | Watch DID | 1.4 (1.2, 1.6) | 1.7 (1.5, 2) | 1.1 (1, 1.4) | 1.4 (1.2, 1.6) |
| **Colombia** | COL | Cluster 3 | AMR | Upper middle income | Reserve DID | 0.1 (0, 0.2) | 0.1 (0, 0.2) | 0.1 (0, 0.2) | 0.1 (0, 0.2) |
| **Comoros** | COM | Cluster 1 | AFR | Lower middle income | Total DID | 16.5 (12.6, 23.5) | 16.5 (12.6, 23.5) | 16.3 (12.5, 23.3) | 11.1 (8.4, 16.8) |
| **Comoros** | COM | Cluster 1 | AFR | Lower middle income | Access DID | 10.5 (5.6, 18.2) | 9.2 (4.1, 16.9) | 12.7 (8.8, 19.8) | 5.2 (1, 10.9) |
| **Comoros** | COM | Cluster 1 | AFR | Lower middle income | Watch DID | 4.6 (3.9, 5.9) | 6 (5.2, 7.3) | 3.5 (2.8, 4.7) | 4.6 (3.9, 5.9) |
| **Comoros** | COM | Cluster 1 | AFR | Lower middle income | Reserve DID | 1.1 (0.3, 4.6) | 1.1 (0.3, 4.6) | 0.1 (0, 0.2) | 1.1 (0.3, 4.6) |
| **Congo, Dem. Rep.** | COD | Cluster 1 | AFR | Low income | Total DID | 19.1 (14.6, 27.1) | 19.2 (14.6, 27.2) | 18.9 (14.3, 26.8) | 12.9 (9.7, 19.6) |
| **Congo, Dem. Rep.** | COD | Cluster 1 | AFR | Low income | Access DID | 14.9 (10.4, 23) | 13.8 (9.2, 21.8) | 16.2 (11.8, 24) | 8.8 (5.2, 15.7) |
| **Congo, Dem. Rep.** | COD | Cluster 1 | AFR | Low income | Watch DID | 3.7 (2.9, 4.6) | 4.8 (4, 6) | 2.4 (1.9, 3.3) | 3.7 (2.9, 4.6) |
| **Congo, Dem. Rep.** | COD | Cluster 1 | AFR | Low income | Reserve DID | 0.4 (0.1, 1.5) | 0.4 (0.1, 1.5) | 0.1 (0.1, 0.4) | 0.4 (0.1, 1.5) |
| **Congo, Rep.** | COG | Cluster 1 | AFR | Lower middle income | Total DID | 18.7 (14.4, 26.2) | 18.7 (14.5, 26.3) | 18.4 (14.2, 26) | 12.5 (9.3, 19.6) |
| **Congo, Rep.** | COG | Cluster 1 | AFR | Lower middle income | Access DID | 14.9 (10.6, 22.5) | 14 (9.7, 21.6) | 15.8 (11.5, 23.3) | 8.9 (5.5, 16) |
| **Congo, Rep.** | COG | Cluster 1 | AFR | Lower middle income | Watch DID | 3.3 (2.7, 4.4) | 4.3 (3.6, 5.3) | 2.4 (1.8, 3.4) | 3.3 (2.7, 4.4) |
| **Congo, Rep.** | COG | Cluster 1 | AFR | Lower middle income | Reserve DID | 0.3 (0.1, 1.1) | 0.3 (0.1, 1.1) | 0.1 (0, 0.3) | 0.3 (0.1, 1.1) |
| **Costa Rica** | CRI | Cluster 3 | AMR | Upper middle income | Total DID | 15 (11.6, 21.9) | 15 (11.6, 21.9) | 15.2 (11.7, 22.1) | 10.2 (7.7, 15.8) |
| **Costa Rica** | CRI | Cluster 3 | AMR | Upper middle income | Access DID | 13.5 (10, 20.4) | 13.2 (9.8, 20.2) | 13.8 (10.3, 20.9) | 8.6 (6.1, 14.2) |
| **Costa Rica** | CRI | Cluster 3 | AMR | Upper middle income | Watch DID | 1.5 (1.3, 1.8) | 1.8 (1.6, 2.1) | 1.4 (1.2, 1.7) | 1.5 (1.3, 1.8) |
| **Costa Rica** | CRI | Cluster 3 | AMR | Upper middle income | Reserve DID | 0 (0, 0) | 0 (0, 0) | 0 (0, 0.1) | 0 (0, 0) |
| **Cote d'Ivoire** | CIV | Cluster 1 | AFR | Lower middle income | Total DID | 18.2 (13.9, 25.4) | 18.2 (13.9, 25.5) | 17.9 (13.6, 25) | 12.1 (9.2, 18.9) |
| **Cote d'Ivoire** | CIV | Cluster 1 | AFR | Lower middle income | Access DID | 12.6 (8.3, 19.7) | 10.9 (6.6, 18) | 13.9 (9.7, 21) | 6.8 (3.6, 13.7) |
| **Cote d'Ivoire** | CIV | Cluster 1 | AFR | Lower middle income | Watch DID | 5.1 (4.2, 6.5) | 6.9 (5.8, 8.3) | 3.7 (2.8, 5) | 5.1 (4.2, 6.5) |
| **Cote d'Ivoire** | CIV | Cluster 1 | AFR | Lower middle income | Reserve DID | 0.3 (0.1, 0.9) | 0.3 (0.1, 0.9) | 0.1 (0, 0.4) | 0.3 (0.1, 0.9) |

| | | | | | | | | | |
|---|---|---|---|---|---|---|---|---|---|
| **Croatia** | HRV | Cluster 4 | EUR | High income | Total DID | 8.2 (7.1, 9.6) | 8.2 (7.1, 9.6) | 8.2 (7.1, 9.6) | 8.2 (7.1, 9.6) |
| **Croatia** | HRV | Cluster 4 | EUR | High income | Access DID | 7.2 (6.1, 8.5) | 6.9 (5.8, 8.3) | 7.3 (6.2, 8.7) | 7.2 (6.1, 8.5) |
| **Croatia** | HRV | Cluster 4 | EUR | High income | Watch DID | 1.1 (0.9, 1.2) | 1.3 (1.2, 1.5) | 0.9 (0.7, 1) | 1.1 (0.9, 1.2) |
| **Croatia** | HRV | Cluster 4 | EUR | High income | Reserve DID | 0 (0, 0) | 0 (0, 0) | 0 (0, 0.1) | 0 (0, 0) |
| **Cuba** | CUB | Cluster 3 | AMR | Upper middle income | Total DID | 15.3 (11.8, 21.5) | 15.3 (11.8, 21.5) | 15.5 (11.9, 21.8) | 10.3 (7.9, 16.1) |
| **Cuba** | CUB | Cluster 3 | AMR | Upper middle income | Access DID | 13.7 (10.2, 19.9) | 13.2 (9.7, 19.4) | 14.2 (10.6, 20.5) | 8.7 (6.2, 14.5) |
| **Cuba** | CUB | Cluster 3 | AMR | Upper middle income | Watch DID | 1.6 (1.4, 1.8) | 2.1 (1.9, 2.3) | 1.3 (1.2, 1.5) | 1.6 (1.4, 1.8) |
| **Cuba** | CUB | Cluster 3 | AMR | Upper middle income | Reserve DID | 0 (0, 0) | 0 (0, 0) | 0 (0, 0.1) | 0 (0, 0) |
| **Cyprus** | CYP | Cluster 4 | EUR | High income | Total DID | 11.8 (10.3, 13.6) | 11.8 (10.3, 13.6) | 11.8 (10.3, 13.6) | 11.8 (10.3, 13.6) |
| **Cyprus** | CYP | Cluster 4 | EUR | High income | Access DID | 10.9 (9.4, 12.7) | 10.7 (9.2, 12.5) | 11 (9.5, 12.8) | 10.9 (9.4, 12.7) |
| **Cyprus** | CYP | Cluster 4 | EUR | High income | Watch DID | 0.9 (0.8, 1) | 1.1 (1, 1.2) | 0.7 (0.7, 0.8) | 0.9 (0.8, 1) |
| **Cyprus** | CYP | Cluster 4 | EUR | High income | Reserve DID | 0 (0, 0) | 0 (0, 0) | 0.1 (0, 0.2) | 0 (0, 0) |
| **Czechia** | CZE | Cluster 4 | EUR | High income | Total DID | 7.9 (6.9, 9.1) | 7.9 (6.9, 9.1) | 7.9 (6.8, 9.1) | 7.9 (6.9, 9.1) |
| **Czechia** | CZE | Cluster 4 | EUR | High income | Access DID | 6.9 (5.8, 8) | 6.6 (5.6, 7.8) | 7.1 (6, 8.2) | 6.9 (5.8, 8) |
| **Czechia** | CZE | Cluster 4 | EUR | High income | Watch DID | 1 (0.9, 1.2) | 1.3 (1.1, 1.4) | 0.8 (0.7, 0.9) | 1 (0.9, 1.2) |
| **Czechia** | CZE | Cluster 4 | EUR | High income | Reserve DID | 0 (0, 0) | 0 (0, 0) | 0 (0, 0.1) | 0 (0, 0) |
| **Denmark** | DNK | Cluster 4 | EUR | High income | Total DID | 11 (9.5, 12.7) | 11 (9.5, 12.7) | 11 (9.5, 12.7) | 11 (9.5, 12.7) |
| **Denmark** | DNK | Cluster 4 | EUR | High income | Access DID | 9.6 (8.2, 11.3) | 9.4 (7.9, 11.1) | 9.8 (8.3, 11.5) | 9.6 (8.2, 11.3) |
| **Denmark** | DNK | Cluster 4 | EUR | High income | Watch DID | 1.3 (1.2, 1.4) | 1.6 (1.5, 1.8) | 1.1 (1, 1.2) | 1.3 (1.2, 1.4) |
| **Denmark** | DNK | Cluster 4 | EUR | High income | Reserve DID | 0 (0, 0) | 0 (0, 0) | 0 (0, 0.1) | 0 (0, 0) |
| **Djibouti** | DJI | Cluster 2 | EMR | Lower middle income | Total DID | 21.1 (16.2, 30.6) | 21.1 (16.2, 30.6) | 20.9 (16.1, 30.4) | 14.3 (10.8, 21.8) |
| **Djibouti** | DJI | Cluster 2 | EMR | Lower middle income | Access DID | 14.7 (9.5, 24.6) | 13.2 (8, 23.2) | 16.6 (11.7, 26.4) | 7.9 (3.8, 15.5) |
| **Djibouti** | DJI | Cluster 2 | EMR | Lower middle income | Watch DID | 5.2 (4.3, 6.6) | 6.8 (5.8, 8.2) | 3.9 (3.2, 5.3) | 5.2 (4.3, 6.6) |
| **Djibouti** | DJI | Cluster 2 | EMR | Lower middle income | Reserve DID | 1 (0.4, 2.3) | 1 (0.4, 2.3) | 0.2 (0.1, 0.7) | 1 (0.4, 2.3) |
| **Dominican Republic** | DOM | Cluster 3 | AMR | Upper middle income | Total DID | 15.5 (11.9, 21.5) | 15.5 (11.9, 21.5) | 15.5 (11.9, 21.6) | 10.5 (7.8, 16.3) |
| **Dominican Republic** | DOM | Cluster 3 | AMR | Upper middle income | Access DID | 13.6 (10, 19.8) | 13.1 (9.5, 19.4) | 14.1 (10.5, 20.3) | 8.6 (5.9, 14.5) |
| **Dominican Republic** | DOM | Cluster 3 | AMR | Upper middle income | Watch DID | 1.8 (1.5, 2.1) | 2.3 (2, 2.7) | 1.3 (1.2, 1.6) | 1.8 (1.5, 2.1) |

| | | | | | | | | | |
|---|---|---|---|---|---|---|---|---|---|
| **Dominican Republic** | DOM | Cluster 3 | AMR | Upper middle income | Reserve DID | 0 (0, 0.1) | 0 (0, 0.1) | 0.1 (0, 0.3) | 0 (0, 0.1) |
| **Ecuador** | ECU | Cluster 3 | AMR | Upper middle income | Total DID | 18 (14.1, 25.7) | 18 (14.1, 25.7) | 18.2 (14.2, 26) | 12.2 (9.2, 18.4) |
| **Ecuador** | ECU | Cluster 3 | AMR | Upper middle income | Access DID | 16.1 (12, 23.8) | 15.6 (11.4, 23.3) | 16.7 (12.6, 24.4) | 10.3 (7.2, 16.5) |
| **Ecuador** | ECU | Cluster 3 | AMR | Upper middle income | Watch DID | 1.7 (1.5, 1.9) | 2.3 (2, 2.5) | 1.4 (1.2, 1.6) | 1.7 (1.5, 1.9) |
| **Ecuador** | ECU | Cluster 3 | AMR | Upper middle income | Reserve DID | 0.1 (0.1, 0.5) | 0.1 (0.1, 0.5) | 0.1 (0, 0.2) | 0.1 (0.1, 0.5) |
| **Egypt, Arab Rep.** | EGY | Cluster 2 | EMR | Lower middle income | Total DID | 13.4 (10.3, 18.9) | 13.4 (10.3, 18.9) | 13.3 (10.2, 18.8) | 9 (6.8, 14.1) |
| **Egypt, Arab Rep.** | EGY | Cluster 2 | EMR | Lower middle income | Access DID | 11.5 (8.4, 17.2) | 11 (7.8, 16.6) | 11.9 (8.7, 17.5) | 7.2 (5, 12.2) |
| **Egypt, Arab Rep.** | EGY | Cluster 2 | EMR | Lower middle income | Watch DID | 1.8 (1.6, 2) | 2.3 (2.1, 2.6) | 1.4 (1.2, 1.5) | 1.8 (1.6, 2) |
| **Egypt, Arab Rep.** | EGY | Cluster 2 | EMR | Lower middle income | Reserve DID | 0.1 (0, 0.1) | 0.1 (0, 0.1) | 0.1 (0, 0.2) | 0.1 (0, 0.1) |
| **El Salvador** | SLV | Cluster 3 | AMR | Upper middle income | Total DID | 17 (13, 24.2) | 17 (13.1, 24.2) | 17.2 (13.2, 24.5) | 11.5 (8.7, 17.6) |
| **El Salvador** | SLV | Cluster 3 | AMR | Upper middle income | Access DID | 15.3 (11.4, 22.5) | 14.9 (11, 22.2) | 15.8 (11.8, 23.1) | 9.8 (7, 16) |
| **El Salvador** | SLV | Cluster 3 | AMR | Upper middle income | Watch DID | 1.6 (1.4, 1.9) | 2 (1.8, 2.3) | 1.3 (1.2, 1.6) | 1.6 (1.4, 1.9) |
| **El Salvador** | SLV | Cluster 3 | AMR | Upper middle income | Reserve DID | 0.1 (0, 0.2) | 0.1 (0, 0.2) | 0.1 (0, 0.1) | 0.1 (0, 0.2) |
| **Equatorial Guinea** | GNQ | Cluster 1 | AFR | Upper middle income | Total DID | 17.7 (13.6, 25.7) | 17.8 (13.6, 25.7) | 17.5 (13.3, 25.4) | 12 (8.9, 18.5) |
| **Equatorial Guinea** | GNQ | Cluster 1 | AFR | Upper middle income | Access DID | 14.4 (10.1, 22.4) | 13.6 (9.1, 21.5) | 15.2 (10.9, 23.1) | 8.7 (5.5, 15.2) |
| **Equatorial Guinea** | GNQ | Cluster 1 | AFR | Upper middle income | Watch DID | 3 (2.3, 4.1) | 3.9 (3, 5.2) | 2.1 (1.6, 2.9) | 3 (2.3, 4.1) |
| **Equatorial Guinea** | GNQ | Cluster 1 | AFR | Upper middle income | Reserve DID | 0.2 (0.1, 0.7) | 0.2 (0.1, 0.7) | 0.1 (0, 0.5) | 0.2 (0.1, 0.7) |
| **Eritrea** | ERI | Cluster 1 | AFR | Low income | Total DID | 36.5 (28, 51.3) | 36.6 (28.1, 51.4) | 36.4 (27.9, 51.2) | 24.6 (18.6, 38.6) |
| **Eritrea** | ERI | Cluster 1 | AFR | Low income | Access DID | 22.7 (13.5, 38.1) | 19.7 (10.5, 35.2) | 26.6 (18.3, 42.2) | 10.9 (2.8, 24.3) |
| **Eritrea** | ERI | Cluster 1 | AFR | Low income | Watch DID | 11.8 (9.8, 15.2) | 14.8 (12.6, 18.4) | 9.3 (7.4, 12.8) | 11.8 (9.8, 15.2) |
| **Eritrea** | ERI | Cluster 1 | AFR | Low income | Reserve DID | 1.5 (0.4, 6.1) | 1.5 (0.4, 6.1) | 0.2 (0.1, 0.4) | 1.5 (0.4, 6.1) |
| **Estonia** | EST | Cluster 4 | EUR | High income | Total DID | 10.1 (8.7, 11.8) | 10.1 (8.8, 11.8) | 10.1 (8.7, 11.8) | 10.1 (8.7, 11.8) |
| **Estonia** | EST | Cluster 4 | EUR | High income | Access DID | 8.6 (7.2, 10.3) | 8.3 (6.9, 10) | 9 (7.6, 10.7) | 8.6 (7.2, 10.3) |
| **Estonia** | EST | Cluster 4 | EUR | High income | Watch DID | 1.3 (1.2, 1.5) | 1.6 (1.5, 1.8) | 1.1 (1, 1.2) | 1.3 (1.2, 1.5) |

| | | | | | | | | | |
|---|---|---|---|---|---|---|---|---|---|
| **Estonia** | EST | Cluster 4 | EUR | High income | Reserve DID | 0.2 (0.1, 0.4) | 0.2 (0.1, 0.4) | 0 (0, 0) | 0.2 (0.1, 0.4) |
| **Eswatini** | SWZ | Cluster 2 | AFR | Lower middle income | Total DID | 16.6 (12.8, 23.4) | 16.6 (12.8, 23.4) | 16.4 (12.5, 23) | 11.2 (8.6, 17.6) |
| **Eswatini** | SWZ | Cluster 2 | AFR | Lower middle income | Access DID | 9.4 (2.7, 16.6) | 8.6 (1.9, 15.9) | 13 (9.1, 19.6) | 4.1 (-2.9, 10.7) |
| **Eswatini** | SWZ | Cluster 2 | AFR | Lower middle income | Watch DID | 4.6 (3.7, 5.8) | 5.3 (4.4, 6.6) | 3.2 (2.6, 4.3) | 4.6 (3.7, 5.8) |
| **Eswatini** | SWZ | Cluster 2 | AFR | Lower middle income | Reserve DID | 2.5 (0.8, 8.1) | 2.5 (0.8, 8.1) | 0.1 (0, 0.3) | 2.5 (0.8, 8.1) |
| **Ethiopia** | ETH | Cluster 1 | AFR | Low income | Total DID | 14.8 (11.4, 21) | 14.9 (11.4, 21.1) | 14.5 (11.1, 20.7) | 10 (7.6, 15.5) |
| **Ethiopia** | ETH | Cluster 1 | AFR | Low income | Access DID | 8.6 (4.8, 15) | 6.8 (3, 13.2) | 9.8 (6.3, 16) | 3.7 (0.6, 9.2) |
| **Ethiopia** | ETH | Cluster 1 | AFR | Low income | Watch DID | 5.7 (4.5, 8) | 7.5 (6.2, 9.9) | 4.5 (3.3, 6.9) | 5.7 (4.5, 8) |
| **Ethiopia** | ETH | Cluster 1 | AFR | Low income | Reserve DID | 0.3 (0.1, 1.2) | 0.3 (0.1, 1.2) | 0.1 (0, 0.2) | 0.3 (0.1, 1.2) |
| **Fiji** | FJI | Cluster 2 | WPR | Upper middle income | Total DID | 19.7 (15.2, 28.5) | 19.7 (15.2, 28.5) | 19.7 (15.2, 28.6) | 13.4 (10.1, 20.4) |
| **Fiji** | FJI | Cluster 2 | WPR | Upper middle income | Access DID | 17.3 (12.8, 26.1) | 16.4 (11.8, 25.3) | 17.9 (13.4, 26.8) | 11 (7.6, 18.2) |
| **Fiji** | FJI | Cluster 2 | WPR | Upper middle income | Watch DID | 2.4 (2, 2.8) | 3.2 (2.8, 3.8) | 1.7 (1.4, 2.2) | 2.4 (2, 2.8) |
| **Fiji** | FJI | Cluster 2 | WPR | Upper middle income | Reserve DID | 0 (0, 0) | 0 (0, 0) | 0 (0, 0.1) | 0 (0, 0) |
| **Finland** | FIN | Cluster 4 | EUR | High income | Total DID | 10.6 (9.3, 12.3) | 10.6 (9.3, 12.2) | 10.6 (9.3, 12.3) | 10.6 (9.3, 12.3) |
| **Finland** | FIN | Cluster 4 | EUR | High income | Access DID | 9.7 (8.4, 11.5) | 9.5 (8.2, 11.2) | 9.9 (8.6, 11.6) | 9.7 (8.4, 11.5) |
| **Finland** | FIN | Cluster 4 | EUR | High income | Watch DID | 0.8 (0.8, 0.9) | 1.1 (1, 1.2) | 0.7 (0.6, 0.7) | 0.8 (0.8, 0.9) |
| **Finland** | FIN | Cluster 4 | EUR | High income | Reserve DID | 0 (0, 0) | 0 (0, 0) | 0 (0, 0) | 0 (0, 0) |
| **France** | FRA | Cluster 4 | EUR | High income | Total DID | 10.7 (9.3, 12.4) | 10.7 (9.3, 12.4) | 10.7 (9.3, 12.3) | 10.7 (9.3, 12.4) |
| **France** | FRA | Cluster 4 | EUR | High income | Access DID | 9.7 (8.2, 11.3) | 9.4 (8, 11.1) | 9.8 (8.4, 11.5) | 9.7 (8.2, 11.3) |
| **France** | FRA | Cluster 4 | EUR | High income | Watch DID | 1 (0.9, 1.2) | 1.3 (1.2, 1.5) | 0.9 (0.8, 1) | 1 (0.9, 1.2) |
| **France** | FRA | Cluster 4 | EUR | High income | Reserve DID | 0 (0, 0) | 0 (0, 0) | 0 (0, 0) | 0 (0, 0) |
| **Gabon** | GAB | Cluster 2 | AFR | Upper middle income | Total DID | 16.4 (12.5, 23) | 16.4 (12.6, 23) | 16.3 (12.4, 22.9) | 11.1 (8.3, 17.2) |
| **Gabon** | GAB | Cluster 2 | AFR | Upper middle income | Access DID | 13.8 (9.9, 20.3) | 13.1 (9.2, 19.6) | 14.4 (10.6, 21) | 8.4 (5.6, 14.5) |
| **Gabon** | GAB | Cluster 2 | AFR | Upper middle income | Watch DID | 2.3 (1.9, 3) | 3 (2.6, 3.7) | 1.7 (1.3, 2.3) | 2.3 (1.9, 3) |
| **Gabon** | GAB | Cluster 2 | AFR | Upper middle income | Reserve DID | 0.2 (0.1, 0.9) | 0.2 (0.1, 0.9) | 0.1 (0, 0.4) | 0.2 (0.1, 0.9) |
| **Gambia, The** | GMB | Cluster 1 | AFR | Low income | Total DID | 13.9 (10.8, 19.9) | 14 (10.8, 19.9) | 13.6 (10.5, 19.4) | 9.4 (7.2, 14.4) |

| | | | | | | | | | |
|---|---|---|---|---|---|---|---|---|---|
| **Gambia, The** | GMB | Cluster 1 | AFR | Low income | Access DID | 9.2 (5.9, 14.8) | 7.7 (4.4, 13.3) | 10 (6.9, 15.6) | 4.6 (2.2, 9.8) |
| **Gambia, The** | GMB | Cluster 1 | AFR | Low income | Watch DID | 4.5 (3.7, 5.8) | 6 (5.1, 7.4) | 3.3 (2.5, 4.6) | 4.5 (3.7, 5.8) |
| **Gambia, The** | GMB | Cluster 1 | AFR | Low income | Reserve DID | 0.2 (0.1, 0.8) | 0.2 (0.1, 0.8) | 0.2 (0.1, 0.6) | 0.2 (0.1, 0.8) |
| **Georgia** | GEO | Cluster 4 | EUR | Upper middle income | Total DID | 6.9 (6, 8) | 6.9 (6, 8) | 6.9 (6, 7.9) | 6.9 (6, 8) |
| **Georgia** | GEO | Cluster 4 | EUR | Upper middle income | Access DID | 4.3 (3.1, 5.5) | 4 (2.8, 5.2) | 5.6 (4.7, 6.7) | 4.3 (3.1, 5.5) |
| **Georgia** | GEO | Cluster 4 | EUR | Upper middle income | Watch DID | 1.7 (1.5, 1.9) | 2 (1.8, 2.2) | 1.2 (1.1, 1.4) | 1.7 (1.5, 1.9) |
| **Georgia** | GEO | Cluster 4 | EUR | Upper middle income | Reserve DID | 0.9 (0.5, 1.8) | 0.9 (0.5, 1.8) | 0 (0, 0.1) | 0.9 (0.5, 1.8) |
| **Germany** | DEU | Cluster 4 | EUR | High income | Total DID | 10.9 (9.5, 12.6) | 10.9 (9.5, 12.6) | 10.9 (9.5, 12.6) | 10.9 (9.5, 12.6) |
| **Germany** | DEU | Cluster 4 | EUR | High income | Access DID | 9.9 (8.5, 11.6) | 9.6 (8.2, 11.3) | 10.1 (8.7, 11.8) | 9.9 (8.5, 11.6) |
| **Germany** | DEU | Cluster 4 | EUR | High income | Watch DID | 0.9 (0.9, 1) | 1.3 (1.1, 1.4) | 0.7 (0.7, 0.8) | 0.9 (0.9, 1) |
| **Germany** | DEU | Cluster 4 | EUR | High income | Reserve DID | 0 (0, 0.1) | 0 (0, 0.1) | 0 (0, 0.1) | 0 (0, 0.1) |
| **Ghana** | GHA | Cluster 1 | AFR | Lower middle income | Total DID | 16.4 (12.6, 22.6) | 16.4 (12.6, 22.7) | 16.1 (12.3, 22.3) | 10.9 (8.4, 16.9) |
| **Ghana** | GHA | Cluster 1 | AFR | Lower middle income | Access DID | 10.6 (6.7, 16.9) | 8.3 (4.4, 14.8) | 11.9 (8.1, 18.1) | 5.2 (2.4, 11.1) |
| **Ghana** | GHA | Cluster 1 | AFR | Lower middle income | Watch DID | 5.4 (4.5, 7) | 7.7 (6.6, 9.4) | 3.8 (3, 5.4) | 5.4 (4.5, 7) |
| **Ghana** | GHA | Cluster 1 | AFR | Lower middle income | Reserve DID | 0.3 (0.1, 0.9) | 0.3 (0.1, 0.9) | 0.2 (0.1, 0.5) | 0.3 (0.1, 0.9) |
| **Greece** | GRC | Cluster 4 | EUR | High income | Total DID | 10 (8.7, 11.7) | 10 (8.7, 11.7) | 10 (8.7, 11.7) | 10 (8.7, 11.7) |
| **Greece** | GRC | Cluster 4 | EUR | High income | Access DID | 9.2 (7.9, 10.9) | 8.9 (7.6, 10.7) | 9.3 (8, 11) | 9.2 (7.9, 10.9) |
| **Greece** | GRC | Cluster 4 | EUR | High income | Watch DID | 0.8 (0.8, 0.9) | 1.1 (1, 1.2) | 0.6 (0.6, 0.7) | 0.8 (0.8, 0.9) |
| **Greece** | GRC | Cluster 4 | EUR | High income | Reserve DID | 0 (0, 0.1) | 0 (0, 0.1) | 0.1 (0, 0.2) | 0 (0, 0.1) |
| **Grenada** | GRD | Cluster 3 | AMR | Upper middle income | Total DID | 13.8 (10.7, 19.2) | 13.8 (10.7, 19.3) | 13.8 (10.7, 19.3) | 9.3 (7.1, 14.5) |
| **Grenada** | GRD | Cluster 3 | AMR | Upper middle income | Access DID | 12.2 (9, 17.7) | 11.8 (8.6, 17.2) | 12.6 (9.4, 18) | 7.6 (5.4, 12.6) |
| **Grenada** | GRD | Cluster 3 | AMR | Upper middle income | Watch DID | 1.6 (1.4, 1.8) | 2 (1.8, 2.3) | 1.2 (1.1, 1.5) | 1.6 (1.4, 1.8) |
| **Grenada** | GRD | Cluster 3 | AMR | Upper middle income | Reserve DID | 0 (0, 0) | 0 (0, 0) | 0 (0, 0) | 0 (0, 0) |
| **Guatemala** | GTM | Cluster 2 | AMR | Upper middle income | Total DID | 15.1 (11.6, 21.5) | 15.1 (11.6, 21.5) | 15 (11.5, 21.4) | 10.2 (7.7, 15.6) |
| **Guatemala** | GTM | Cluster 2 | AMR | Upper middle income | Access DID | 12.9 (9.4, 19.4) | 12.2 (8.6, 18.6) | 13.3 (9.8, 19.7) | 8 (5.5, 13.5) |

| | | | | | | | | | |
|---|---|---|---|---|---|---|---|---|---|
| **Guatemala** | GTM | Cluster 2 | AMR | Upper middle income | Watch DID | 2.1 (1.8, 2.5) | 2.9 (2.5, 3.4) | 1.5 (1.3, 1.9) | 2.1 (1.8, 2.5) |
| **Guatemala** | GTM | Cluster 2 | AMR | Upper middle income | Reserve DID | 0.1 (0, 0.2) | 0.1 (0, 0.2) | 0.1 (0.1, 0.3) | 0.1 (0, 0.2) |
| **Guinea** | GIN | Cluster 1 | AFR | Low income | Total DID | 15.8 (12.2, 22.3) | 15.8 (12.2, 22.3) | 15.4 (11.8, 21.8) | 10.6 (8, 16.3) |
| **Guinea** | GIN | Cluster 1 | AFR | Low income | Access DID | 9.7 (6, 16.3) | 7.7 (4, 14.4) | 11 (7.3, 17.3) | 4.6 (1.7, 10.2) |
| **Guinea** | GIN | Cluster 1 | AFR | Low income | Watch DID | 5.8 (4.7, 7.5) | 7.8 (6.5, 9.6) | 4.1 (3.1, 5.8) | 5.8 (4.7, 7.5) |
| **Guinea** | GIN | Cluster 1 | AFR | Low income | Reserve DID | 0.2 (0.1, 0.6) | 0.2 (0.1, 0.6) | 0.1 (0.1, 0.4) | 0.2 (0.1, 0.6) |
| **Guinea-Bissau** | GNB | Cluster 1 | AFR | Low income | Total DID | 16.3 (12.3, 23.3) | 16.4 (12.4, 23.3) | 16 (12.1, 22.9) | 11 (8.3, 17.1) |
| **Guinea-Bissau** | GNB | Cluster 1 | AFR | Low income | Access DID | 11.4 (7.5, 18.1) | 10.4 (6.5, 17.1) | 12.1 (8.3, 18.7) | 6.1 (3.2, 12.2) |
| **Guinea-Bissau** | GNB | Cluster 1 | AFR | Low income | Watch DID | 4.5 (3.5, 6.2) | 5.6 (4.5, 7.3) | 3.5 (2.6, 5.1) | 4.5 (3.5, 6.2) |
| **Guinea-Bissau** | GNB | Cluster 1 | AFR | Low income | Reserve DID | 0.2 (0.1, 0.8) | 0.2 (0.1, 0.8) | 0.3 (0.1, 0.9) | 0.2 (0.1, 0.8) |
| **Guyana** | GUY | Cluster 3 | AMR | Upper middle income | Total DID | 16.1 (12.2, 22.7) | 16.1 (12.2, 22.7) | 16.2 (12.3, 22.8) | 10.8 (8.1, 16.6) |
| **Guyana** | GUY | Cluster 3 | AMR | Upper middle income | Access DID | 14.4 (10.6, 21.1) | 13.8 (9.9, 20.3) | 14.8 (11, 21.5) | 9.2 (6.5, 15) |
| **Guyana** | GUY | Cluster 3 | AMR | Upper middle income | Watch DID | 1.6 (1.4, 1.8) | 2.3 (2.1, 2.5) | 1.2 (1.1, 1.4) | 1.6 (1.4, 1.8) |
| **Guyana** | GUY | Cluster 3 | AMR | Upper middle income | Reserve DID | 0 (0, 0.1) | 0 (0, 0.1) | 0.1 (0, 0.2) | 0 (0, 0.1) |
| **Haiti** | HTI | Cluster 2 | AMR | Lower middle income | Total DID | 22.7 (17.4, 32.3) | 22.7 (17.4, 32.3) | 22.6 (17.2, 32.1) | 15.3 (11.5, 23.6) |
| **Haiti** | HTI | Cluster 2 | AMR | Lower middle income | Access DID | 20.4 (15, 29.9) | 19.4 (14, 28.8) | 21 (15.6, 30.5) | 12.9 (9.2, 21.4) |
| **Haiti** | HTI | Cluster 2 | AMR | Lower middle income | Watch DID | 2.3 (2, 2.7) | 3.3 (2.8, 3.8) | 1.5 (1.3, 1.7) | 2.3 (2, 2.7) |
| **Haiti** | HTI | Cluster 2 | AMR | Lower middle income | Reserve DID | 0.1 (0, 0.1) | 0.1 (0, 0.1) | 0.1 (0, 0.4) | 0.1 (0, 0.1) |
| **Honduras** | HND | Cluster 2 | AMR | Lower middle income | Total DID | 17.6 (13.8, 25.4) | 17.6 (13.8, 25.4) | 17.6 (13.7, 25.4) | 11.8 (9, 18.6) |
| **Honduras** | HND | Cluster 2 | AMR | Lower middle income | Access DID | 15.5 (11.6, 23.4) | 15.1 (11.2, 23) | 15.9 (11.9, 23.8) | 9.9 (6.9, 16.6) |
| **Honduras** | HND | Cluster 2 | AMR | Lower middle income | Watch DID | 1.9 (1.7, 2.4) | 2.3 (2, 2.7) | 1.6 (1.3, 2) | 1.9 (1.7, 2.4) |
| **Honduras** | HND | Cluster 2 | AMR | Lower middle income | Reserve DID | 0.1 (0, 0.2) | 0.1 (0, 0.2) | 0 (0, 0.1) | 0.1 (0, 0.2) |
| **Hungary** | HUN | Cluster 4 | EUR | High income | Total DID | 7.8 (6.8, 9) | 7.8 (6.8, 9) | 7.7 (6.7, 9) | 7.8 (6.8, 9) |
| **Hungary** | HUN | Cluster 4 | EUR | High income | Access DID | 6.8 (5.8, 8) | 6.6 (5.6, 7.8) | 6.9 (6, 8.1) | 6.8 (5.8, 8) |
| **Hungary** | HUN | Cluster 4 | EUR | High income | Watch DID | 1 (0.8, 1.1) | 1.2 (1, 1.3) | 0.8 (0.7, 0.9) | 1 (0.8, 1.1) |

| | | | | | | | | | |
|---|---|---|---|---|---|---|---|---|---|
| **Hungary** | HUN | Cluster 4 | EUR | High income | Reserve DID | 0 (0, 0) | 0 (0, 0) | 0 (0, 0.1) | 0 (0, 0) |
| **Iceland** | ISL | Cluster 4 | EUR | High income | Total DID | 10.8 (9.4, 12.5) | 10.8 (9.4, 12.5) | 10.8 (9.4, 12.5) | 10.8 (9.4, 12.5) |
| **Iceland** | ISL | Cluster 4 | EUR | High income | Access DID | 9.9 (8.4, 11.5) | 9.7 (8.2, 11.3) | 10 (8.6, 11.6) | 9.9 (8.4, 11.5) |
| **Iceland** | ISL | Cluster 4 | EUR | High income | Watch DID | 1 (0.8, 1.1) | 1.2 (1, 1.4) | 0.8 (0.7, 1) | 1 (0.8, 1.1) |
| **Iceland** | ISL | Cluster 4 | EUR | High income | Reserve DID | 0 (0, 0) | 0 (0, 0) | 0 (0, 0) | 0 (0, 0) |
| **India** | IND | Cluster 2 | SEAR | Lower middle income | Total DID | 14.7 (11.4, 20.7) | 14.7 (11.4, 20.8) | 14.5 (11.2, 20.5) | 9.9 (7.5, 15.5) |
| **India** | IND | Cluster 2 | SEAR | Lower middle income | Access DID | 7.6 (3.8, 13.7) | 3.4 (-0.8, 9.5) | 10.5 (7, 16.3) | 2.8 (-0.1, 8.2) |
| **India** | IND | Cluster 2 | SEAR | Lower middle income | Watch DID | 6 (5.3, 7) | 10.3 (8.9, 11.7) | 3.9 (3.3, 4.9) | 6 (5.3, 7) |
| **India** | IND | Cluster 2 | SEAR | Lower middle income | Reserve DID | 1 (0.4, 2.8) | 1 (0.4, 2.8) | 0.2 (0.1, 0.4) | 1 (0.4, 2.8) |
| **Indonesia** | IDN | Cluster 2 | SEAR | Upper middle income | Total DID | 19.7 (15.4, 27.7) | 19.7 (15.4, 27.7) | 19.6 (15.3, 27.7) | 13.4 (10.1, 20.6) |
| **Indonesia** | IDN | Cluster 2 | SEAR | Upper middle income | Access DID | 16.5 (12.2, 24.6) | 14.6 (10.3, 22.8) | 17.6 (13.2, 25.6) | 10.2 (7, 17.5) |
| **Indonesia** | IDN | Cluster 2 | SEAR | Upper middle income | Watch DID | 3 (2.7, 3.6) | 5 (4.4, 5.7) | 2 (1.7, 2.5) | 3 (2.7, 3.6) |
| **Indonesia** | IDN | Cluster 2 | SEAR | Upper middle income | Reserve DID | 0.1 (0, 0.2) | 0.1 (0, 0.2) | 0 (0, 0.1) | 0.1 (0, 0.2) |
| **Iran, Islamic Rep.** | IRN | Cluster 3 | EMR | Lower middle income | Total DID | 14.3 (11, 20.4) | 14.3 (11, 20.3) | 14.4 (11.1, 20.5) | 9.7 (7.3, 15) |
| **Iran, Islamic Rep.** | IRN | Cluster 3 | EMR | Lower middle income | Access DID | 12.6 (9.4, 18.6) | 12.2 (8.9, 18.2) | 12.9 (9.6, 19) | 8 (5.7, 13.2) |
| **Iran, Islamic Rep.** | IRN | Cluster 3 | EMR | Lower middle income | Watch DID | 1.6 (1.3, 2.1) | 2.1 (1.7, 2.5) | 1.4 (1.1, 1.9) | 1.6 (1.3, 2.1) |
| **Iran, Islamic Rep.** | IRN | Cluster 3 | EMR | Lower middle income | Reserve DID | 0 (0, 0.1) | 0 (0, 0.1) | 0 (0, 0.1) | 0 (0, 0.1) |
| **Iraq** | IRQ | Cluster 2 | EMR | Upper middle income | Total DID | 14.2 (10.9, 20.1) | 14.3 (10.9, 20.1) | 14.2 (10.8, 20.1) | 9.6 (7.2, 14.6) |
| **Iraq** | IRQ | Cluster 2 | EMR | Upper middle income | Access DID | 12.5 (9.1, 18.2) | 11.9 (8.6, 17.6) | 12.8 (9.5, 18.6) | 7.8 (5.4, 12.8) |
| **Iraq** | IRQ | Cluster 2 | EMR | Upper middle income | Watch DID | 1.7 (1.4, 2.1) | 2.2 (1.9, 2.7) | 1.2 (1, 1.6) | 1.7 (1.4, 2.1) |
| **Iraq** | IRQ | Cluster 2 | EMR | Upper middle income | Reserve DID | 0.1 (0, 0.2) | 0.1 (0, 0.2) | 0.1 (0, 0.3) | 0.1 (0, 0.2) |
| **Ireland** | IRL | Cluster 4 | EUR | High income | Total DID | 11.3 (9.9, 13.1) | 11.3 (9.9, 13.1) | 11.3 (9.8, 13.1) | 11.3 (9.9, 13.1) |
| **Ireland** | IRL | Cluster 4 | EUR | High income | Access DID | 10.4 (8.9, 12.2) | 10.1 (8.7, 12) | 10.4 (9, 12.3) | 10.4 (8.9, 12.2) |
| **Ireland** | IRL | Cluster 4 | EUR | High income | Watch DID | 0.9 (0.8, 1) | 1.1 (1, 1.3) | 0.8 (0.7, 0.9) | 0.9 (0.8, 1) |
| **Ireland** | IRL | Cluster 4 | EUR | High income | Reserve DID | 0 (0, 0) | 0 (0, 0) | 0.1 (0, 0.1) | 0 (0, 0) |

| | | | | | | | | | |
|---|---|---|---|---|---|---|---|---|---|
| **Israel** | ISR | Cluster 3 | EUR | High income | Total DID | 17.6 (13.4, 24.9) | 17.6 (13.4, 24.9) | 17.8 (13.5, 25.4) | 11.8 (9, 18.4) |
| **Israel** | ISR | Cluster 3 | EUR | High income | Access DID | 16.7 (12.4, 24) | 16.4 (12.1, 23.7) | 17.1 (12.8, 24.6) | 11 (8.1, 17.6) |
| **Israel** | ISR | Cluster 3 | EUR | High income | Watch DID | 0.8 (0.7, 0.9) | 1.1 (1, 1.2) | 0.7 (0.6, 0.8) | 0.8 (0.7, 0.9) |
| **Israel** | ISR | Cluster 3 | EUR | High income | Reserve DID | 0 (0, 0.1) | 0 (0, 0.1) | 0 (0, 0.1) | 0 (0, 0.1) |
| **Italy** | ITA | Cluster 4 | EUR | High income | Total DID | 10.7 (9.3, 12.3) | 10.7 (9.3, 12.3) | 10.7 (9.3, 12.3) | 10.7 (9.3, 12.3) |
| **Italy** | ITA | Cluster 4 | EUR | High income | Access DID | 10.1 (8.7, 11.6) | 9.9 (8.4, 11.4) | 10.2 (8.8, 11.8) | 10.1 (8.7, 11.6) |
| **Italy** | ITA | Cluster 4 | EUR | High income | Watch DID | 0.6 (0.6, 0.7) | 0.9 (0.8, 0.9) | 0.5 (0.4, 0.5) | 0.6 (0.6, 0.7) |
| **Italy** | ITA | Cluster 4 | EUR | High income | Reserve DID | 0 (0, 0.1) | 0 (0, 0.1) | 0.1 (0, 0.1) | 0 (0, 0.1) |
| **Jamaica** | JAM | Cluster 3 | AMR | Upper middle income | Total DID | 16.3 (12.3, 23.4) | 16.3 (12.3, 23.4) | 16.2 (12.3, 23.4) | 10.9 (8.3, 17) |
| **Jamaica** | JAM | Cluster 3 | AMR | Upper middle income | Access DID | 14.6 (10.7, 21.7) | 14.2 (10.2, 21.3) | 14.9 (10.9, 22.1) | 9.3 (6.7, 15.3) |
| **Jamaica** | JAM | Cluster 3 | AMR | Upper middle income | Watch DID | 1.6 (1.5, 1.9) | 2.1 (1.9, 2.3) | 1.3 (1.1, 1.5) | 1.6 (1.5, 1.9) |
| **Jamaica** | JAM | Cluster 3 | AMR | Upper middle income | Reserve DID | 0 (0, 0) | 0 (0, 0) | 0.1 (0, 0.2) | 0 (0, 0) |
| **Japan** | JPN | Cluster 4 | WPR | High income | Total DID | 12.8 (11.2, 14.8) | 12.8 (11.2, 14.9) | 12.8 (11.1, 14.8) | 12.8 (11.2, 14.8) |
| **Japan** | JPN | Cluster 4 | WPR | High income | Access DID | 11.8 (10.1, 13.8) | 11.5 (9.8, 13.6) | 12 (10.3, 14.1) | 11.8 (10.1, 13.8) |
| **Japan** | JPN | Cluster 4 | WPR | High income | Watch DID | 1 (0.9, 1.2) | 1.3 (1.2, 1.5) | 0.8 (0.7, 0.9) | 1 (0.9, 1.2) |
| **Japan** | JPN | Cluster 4 | WPR | High income | Reserve DID | 0 (0, 0) | 0 (0, 0) | 0 (0, 0) | 0 (0, 0) |
| **Jordan** | JOR | Cluster 3 | EMR | Upper middle income | Total DID | 12.5 (9.5, 18.1) | 12.5 (9.5, 18.1) | 12.7 (9.6, 18.3) | 8.5 (6.3, 13.1) |
| **Jordan** | JOR | Cluster 3 | EMR | Upper middle income | Access DID | 11.4 (8.4, 16.9) | 10.9 (8, 16.4) | 11.7 (8.7, 17.3) | 7.4 (5.1, 11.9) |
| **Jordan** | JOR | Cluster 3 | EMR | Upper middle income | Watch DID | 1.1 (1, 1.3) | 1.6 (1.4, 1.8) | 0.9 (0.8, 1.1) | 1.1 (1, 1.3) |
| **Jordan** | JOR | Cluster 3 | EMR | Upper middle income | Reserve DID | 0 (0, 0.1) | 0 (0, 0.1) | 0.1 (0, 0.1) | 0 (0, 0.1) |
| **Kazakhstan** | KAZ | Cluster 2 | EUR | Upper middle income | Total DID | 12.3 (9.4, 17.3) | 12.3 (9.4, 17.3) | 12.4 (9.5, 17.6) | 8.3 (6.2, 12.7) |
| **Kazakhstan** | KAZ | Cluster 2 | EUR | Upper middle income | Access DID | 9.6 (6.7, 14.7) | 9.3 (6.3, 14.3) | 11.1 (8.2, 16.3) | 5.6 (3.2, 10.2) |
| **Kazakhstan** | KAZ | Cluster 2 | EUR | Upper middle income | Watch DID | 1.7 (1.5, 1.9) | 2 (1.8, 2.3) | 1.2 (1.1, 1.5) | 1.7 (1.5, 1.9) |
| **Kazakhstan** | KAZ | Cluster 2 | EUR | Upper middle income | Reserve DID | 1 (0.5, 1.9) | 1 (0.5, 1.9) | 0 (0, 0.1) | 1 (0.5, 1.9) |
| **Kenya** | KEN | Cluster 1 | AFR | Lower middle income | Total DID | 23.9 (18.4, 34.2) | 23.9 (18.4, 34.2) | 23.7 (18.2, 34) | 16.1 (12.1, 25.2) |

| | | | | | | | | | |
|---|---|---|---|---|---|---|---|---|---|
| **Kenya** | KEN | Cluster 1 | AFR | Lower middle income | Access DID | 16.5 (10.8, 26.7) | 13.6 (7.7, 24) | 18.2 (12.5, 28.4) | 8.7 (4.6, 17.4) |
| **Kenya** | KEN | Cluster 1 | AFR | Lower middle income | Watch DID | 7.1 (6.2, 8.6) | 10 (8.8, 11.6) | 5.4 (4.5, 6.8) | 7.1 (6.2, 8.6) |
| **Kenya** | KEN | Cluster 1 | AFR | Lower middle income | Reserve DID | 0.2 (0.1, 0.6) | 0.2 (0.1, 0.6) | 0.1 (0, 0.3) | 0.2 (0.1, 0.6) |
| **Kiribati** | KIR | Cluster 2 | WPR | Lower middle income | Total DID | 20.3 (15.4, 29.3) | 20.3 (15.4, 29.3) | 20.2 (15.3, 29.2) | 13.6 (10.2, 21.3) |
| **Kiribati** | KIR | Cluster 2 | WPR | Lower middle income | Access DID | 17.4 (12.5, 26.4) | 16.4 (11.4, 25.3) | 18.2 (13.4, 27.2) | 10.8 (7.4, 18.4) |
| **Kiribati** | KIR | Cluster 2 | WPR | Lower middle income | Watch DID | 2.7 (2.3, 3.4) | 3.8 (3.3, 4.5) | 1.9 (1.5, 2.6) | 2.7 (2.3, 3.4) |
| **Kiribati** | KIR | Cluster 2 | WPR | Lower middle income | Reserve DID | 0.1 (0, 0.3) | 0.1 (0, 0.3) | 0 (0, 0) | 0.1 (0, 0.3) |
| **Korea, Dem. People's Rep.** | PRK | Cluster 2 | SEAR | Low income | Total DID | 11.5 (8.9, 16.3) | 11.6 (8.9, 16.3) | 11.7 (8.9, 16.4) | 7.7 (5.9, 11.8) |
| **Korea, Dem. People's Rep.** | PRK | Cluster 2 | SEAR | Low income | Access DID | 8.8 (5.9, 13.7) | 8.5 (5.5, 13.3) | 10.4 (7.6, 15.2) | 5 (2.6, 9.2) |
| **Korea, Dem. People's Rep.** | PRK | Cluster 2 | SEAR | Low income | Watch DID | 1.7 (1.5, 2) | 2.1 (1.8, 2.4) | 1.2 (1.1, 1.5) | 1.7 (1.5, 2) |
| **Korea, Dem. People's Rep.** | PRK | Cluster 2 | SEAR | Low income | Reserve DID | 0.9 (0.4, 2.4) | 0.9 (0.4, 2.4) | 0 (0, 0.1) | 0.9 (0.4, 2.4) |
| **Korea, Rep.** | KOR | Cluster 4 | WPR | High income | Total DID | 12.1 (10.4, 14.3) | 12.1 (10.4, 14.3) | 12.1 (10.4, 14.3) | 12.1 (10.4, 14.3) |
| **Korea, Rep.** | KOR | Cluster 4 | WPR | High income | Access DID | 11.4 (9.7, 13.5) | 11.2 (9.6, 13.4) | 11.5 (9.9, 13.7) | 11.4 (9.7, 13.5) |
| **Korea, Rep.** | KOR | Cluster 4 | WPR | High income | Watch DID | 0.6 (0.6, 0.7) | 0.8 (0.7, 0.9) | 0.5 (0.4, 0.5) | 0.6 (0.6, 0.7) |
| **Korea, Rep.** | KOR | Cluster 4 | WPR | High income | Reserve DID | 0.1 (0, 0.2) | 0.1 (0, 0.2) | 0.1 (0, 0.2) | 0.1 (0, 0.2) |
| **Kuwait** | KWT | Cluster 3 | EMR | High income | Total DID | 13.4 (10.2, 19.1) | 13.4 (10.1, 19.1) | 13.5 (10.3, 19.3) | 9 (6.8, 14.1) |
| **Kuwait** | KWT | Cluster 3 | EMR | High income | Access DID | 12.1 (8.9, 17.8) | 11.8 (8.5, 17.4) | 12.4 (9.1, 18.2) | 7.7 (5.4, 12.8) |
| **Kuwait** | KWT | Cluster 3 | EMR | High income | Watch DID | 1.2 (1.1, 1.5) | 1.6 (1.4, 1.8) | 1.1 (0.9, 1.3) | 1.2 (1.1, 1.5) |
| **Kuwait** | KWT | Cluster 3 | EMR | High income | Reserve DID | 0 (0, 0.1) | 0 (0, 0.1) | 0 (0, 0) | 0 (0, 0.1) |
| **Kyrgyz Republic** | KGZ | Cluster 2 | EUR | Lower middle income | Total DID | 9.8 (7.5, 14.1) | 9.8 (7.5, 14.1) | 9.8 (7.5, 14.2) | 6.6 (5, 10.2) |
| **Kyrgyz Republic** | KGZ | Cluster 2 | EUR | Lower middle income | Access DID | 6 (3, 10.4) | 5.7 (2.6, 10.1) | 8.5 (6.1, 12.7) | 2.9 (0.2, 6.6) |
| **Kyrgyz Republic** | KGZ | Cluster 2 | EUR | Lower middle income | Watch DID | 2 (1.7, 2.3) | 2.4 (2.1, 2.7) | 1.3 (1.1, 1.6) | 2 (1.7, 2.3) |
| **Kyrgyz Republic** | KGZ | Cluster 2 | EUR | Lower middle income | Reserve DID | 1.7 (0.8, 3.9) | 1.7 (0.8, 3.9) | 0 (0, 0.1) | 1.7 (0.8, 3.9) |
| **Lao PDR** | LAO | Cluster 2 | WPR | Lower middle income | Total DID | 17.7 (13.3, 25.2) | 17.7 (13.3, 25.3) | 17.5 (13.2, 25.1) | 11.9 (9, 18.7) |

| | | | | | | | | | |
|---|---|---|---|---|---|---|---|---|---|
| **Lao PDR** | LAO | Cluster 2 | WPR | Lower middle income | Access DID | 14.6 (10.4, 22.1) | 12.6 (8.3, 20.3) | 15.6 (11.4, 23.1) | 8.9 (5.8, 15.4) |
| **Lao PDR** | LAO | Cluster 2 | WPR | Lower middle income | Watch DID | 3 (2.7, 3.5) | 5 (4.4, 5.8) | 1.9 (1.6, 2.3) | 3 (2.7, 3.5) |
| **Lao PDR** | LAO | Cluster 2 | WPR | Lower middle income | Reserve DID | 0 (0, 0.2) | 0 (0, 0.2) | 0.1 (0, 0.1) | 0 (0, 0.2) |
| **Latvia** | LVA | Cluster 4 | EUR | High income | Total DID | 10.2 (8.9, 11.7) | 10.2 (8.9, 11.7) | 10.2 (8.9, 11.7) | 10.2 (8.9, 11.7) |
| **Latvia** | LVA | Cluster 4 | EUR | High income | Access DID | 8.6 (7.3, 10.2) | 8.3 (7, 9.9) | 9 (7.7, 10.6) | 8.6 (7.3, 10.2) |
| **Latvia** | LVA | Cluster 4 | EUR | High income | Watch DID | 1.4 (1.3, 1.6) | 1.7 (1.6, 1.9) | 1.1 (1, 1.2) | 1.4 (1.3, 1.6) |
| **Latvia** | LVA | Cluster 4 | EUR | High income | Reserve DID | 0.2 (0.1, 0.4) | 0.2 (0.1, 0.4) | 0.1 (0, 0.1) | 0.2 (0.1, 0.4) |
| **Lebanon** | LBN | Cluster 3 | EMR | Upper middle income | Total DID | 13.2 (9.9, 18.5) | 13.2 (9.9, 18.5) | 13.3 (10, 18.7) | 8.9 (6.6, 13.7) |
| **Lebanon** | LBN | Cluster 3 | EMR | Upper middle income | Access DID | 11.7 (8.5, 17.1) | 11.2 (8, 16.7) | 12.1 (8.8, 17.6) | 7.4 (5.1, 12.3) |
| **Lebanon** | LBN | Cluster 3 | EMR | Upper middle income | Watch DID | 1.4 (1.2, 1.7) | 1.9 (1.6, 2.2) | 1.1 (0.9, 1.5) | 1.4 (1.2, 1.7) |
| **Lebanon** | LBN | Cluster 3 | EMR | Upper middle income | Reserve DID | 0 (0, 0.1) | 0 (0, 0.1) | 0.1 (0, 0.2) | 0 (0, 0.1) |
| **Lesotho** | LSO | Cluster 2 | AFR | Lower middle income | Total DID | 16.4 (12.8, 23.3) | 16.5 (12.8, 23.4) | 16.2 (12.5, 22.9) | 11.1 (8.4, 17.2) |
| **Lesotho** | LSO | Cluster 2 | AFR | Lower middle income | Access DID | 10.6 (6.3, 17.6) | 9.7 (5.5, 16.8) | 12.9 (9.3, 19.8) | 5.3 (1.4, 11.3) |
| **Lesotho** | LSO | Cluster 2 | AFR | Lower middle income | Watch DID | 4.1 (3.5, 5) | 4.9 (4.2, 5.9) | 2.9 (2.4, 3.7) | 4.1 (3.5, 5) |
| **Lesotho** | LSO | Cluster 2 | AFR | Lower middle income | Reserve DID | 1.5 (0.5, 4.3) | 1.5 (0.5, 4.3) | 0.3 (0.1, 0.8) | 1.5 (0.5, 4.3) |
| **Liberia** | LBR | Cluster 1 | AFR | Low income | Total DID | 20.9 (16, 29.6) | 20.9 (16.1, 29.7) | 20.6 (15.8, 29.2) | 14.1 (10.7, 21.7) |
| **Liberia** | LBR | Cluster 1 | AFR | Low income | Access DID | 13.6 (8.6, 21.8) | 11.8 (6.8, 20) | 14.8 (9.9, 22.9) | 6.8 (3, 14.6) |
| **Liberia** | LBR | Cluster 1 | AFR | Low income | Watch DID | 7 (5.5, 9.5) | 8.8 (7.2, 11.3) | 5.6 (4.2, 8.1) | 7 (5.5, 9.5) |
| **Liberia** | LBR | Cluster 1 | AFR | Low income | Reserve DID | 0.2 (0, 0.6) | 0.2 (0, 0.6) | 0.1 (0, 0.3) | 0.2 (0, 0.6) |
| **Libya** | LBY | Cluster 3 | EMR | Upper middle income | Total DID | 14 (10.5, 19.8) | 14 (10.5, 19.8) | 14.1 (10.6, 20) | 9.4 (7.1, 14.5) |
| **Libya** | LBY | Cluster 3 | EMR | Upper middle income | Access DID | 12.5 (9.1, 18.3) | 12.1 (8.7, 17.9) | 12.9 (9.5, 18.8) | 7.9 (5.7, 13) |
| **Libya** | LBY | Cluster 3 | EMR | Upper middle income | Watch DID | 1.4 (1.2, 1.6) | 1.8 (1.6, 2.1) | 1.1 (1, 1.3) | 1.4 (1.2, 1.6) |
| **Libya** | LBY | Cluster 3 | EMR | Upper middle income | Reserve DID | 0 (0, 0.1) | 0 (0, 0.1) | 0 (0, 0.1) | 0 (0, 0.1) |
| **Lithuania** | LTU | Cluster 4 | EUR | High income | Total DID | 10.3 (8.9, 11.7) | 10.3 (8.9, 11.7) | 10.2 (8.9, 11.7) | 10.3 (8.9, 11.7) |
| **Lithuania** | LTU | Cluster 4 | EUR | High income | Access DID | 8.1 (6.7, 9.7) | 7.8 (6.4, 9.4) | 9 (7.6, 10.6) | 8.1 (6.7, 9.7) |

| | | | | | | | | | |
|---|---|---|---|---|---|---|---|---|---|
| **Lithuania** | LTU | Cluster 4 | EUR | High income | Watch DID | 1.5 (1.4, 1.7) | 1.9 (1.7, 2) | 1.1 (1, 1.3) | 1.5 (1.4, 1.7) |
| **Lithuania** | LTU | Cluster 4 | EUR | High income | Reserve DID | 0.6 (0.3, 1.1) | 0.6 (0.3, 1.1) | 0.1 (0, 0.1) | 0.6 (0.3, 1.1) |
| **Luxembourg** | LUX | Cluster 4 | EUR | High income | Total DID | 10.9 (9.4, 12.7) | 10.9 (9.4, 12.7) | 10.9 (9.3, 12.7) | 10.9 (9.4, 12.7) |
| **Luxembourg** | LUX | Cluster 4 | EUR | High income | Access DID | 10.1 (8.5, 11.9) | 9.9 (8.3, 11.6) | 10.3 (8.7, 12) | 10.1 (8.5, 11.9) |
| **Luxembourg** | LUX | Cluster 4 | EUR | High income | Watch DID | 0.8 (0.7, 0.9) | 1.1 (0.9, 1.2) | 0.6 (0.6, 0.7) | 0.8 (0.7, 0.9) |
| **Luxembourg** | LUX | Cluster 4 | EUR | High income | Reserve DID | 0 (0, 0) | 0 (0, 0) | 0 (0, 0) | 0 (0, 0) |
| **Madagascar** | MDG | Cluster 1 | AFR | Low income | Total DID | 19.5 (15.1, 27.9) | 19.6 (15.1, 28) | 19.2 (14.8, 27.6) | 13.2 (10, 20.6) |
| **Madagascar** | MDG | Cluster 1 | AFR | Low income | Access DID | 11.9 (7.5, 20) | 10.1 (5.8, 18.4) | 13.4 (9.1, 21.4) | 5.7 (2.2, 12.6) |
| **Madagascar** | MDG | Cluster 1 | AFR | Low income | Watch DID | 7 (5.7, 9.3) | 8.8 (7.3, 11.2) | 5.5 (4.3, 7.8) | 7 (5.7, 9.3) |
| **Madagascar** | MDG | Cluster 1 | AFR | Low income | Reserve DID | 0.3 (0.1, 1.1) | 0.3 (0.1, 1.1) | 0.1 (0, 0.3) | 0.3 (0.1, 1.1) |
| **Malawi** | MWI | Cluster 1 | AFR | Low income | Total DID | 20.2 (15.6, 29.4) | 20.2 (15.6, 29.4) | 20 (15.4, 29.2) | 13.8 (10.4, 21) |
| **Malawi** | MWI | Cluster 1 | AFR | Low income | Access DID | 14.8 (10.2, 23.8) | 13.7 (9.1, 22.7) | 15.7 (11.2, 24.7) | 8.3 (4.6, 15.6) |
| **Malawi** | MWI | Cluster 1 | AFR | Low income | Watch DID | 5 (4.1, 6.8) | 6.1 (5.1, 7.9) | 4 (3.2, 5.7) | 5 (4.1, 6.8) |
| **Malawi** | MWI | Cluster 1 | AFR | Low income | Reserve DID | 0.3 (0.1, 0.9) | 0.3 (0.1, 0.9) | 0.1 (0, 0.2) | 0.3 (0.1, 0.9) |
| **Malaysia** | MYS | Cluster 2 | WPR | Upper middle income | Total DID | 15.8 (12.2, 22.6) | 15.8 (12.2, 22.6) | 15.9 (12.3, 22.7) | 10.7 (8, 16.3) |
| **Malaysia** | MYS | Cluster 2 | WPR | Upper middle income | Access DID | 13.3 (9.7, 20.1) | 12.1 (8.5, 18.6) | 14.1 (10.5, 21) | 8.2 (5.6, 13.8) |
| **Malaysia** | MYS | Cluster 2 | WPR | Upper middle income | Watch DID | 2.4 (2.1, 2.8) | 3.7 (3.3, 4.2) | 1.7 (1.5, 2.1) | 2.4 (2.1, 2.8) |
| **Malaysia** | MYS | Cluster 2 | WPR | Upper middle income | Reserve DID | 0 (0, 0.1) | 0 (0, 0.1) | 0 (0, 0.1) | 0 (0, 0.1) |
| **Maldives** | MDV | Cluster 2 | SEAR | Upper middle income | Total DID | 16.7 (12.8, 23.6) | 16.7 (12.8, 23.6) | 16.9 (13, 23.9) | 11.3 (8.4, 17.7) |
| **Maldives** | MDV | Cluster 2 | SEAR | Upper middle income | Access DID | 14.6 (10.8, 21.5) | 13.4 (9.6, 20.3) | 15.3 (11.5, 22.3) | 9.2 (6.3, 15.7) |
| **Maldives** | MDV | Cluster 2 | SEAR | Upper middle income | Watch DID | 2.1 (1.8, 2.4) | 3.2 (2.8, 3.8) | 1.6 (1.3, 1.9) | 2.1 (1.8, 2.4) |
| **Maldives** | MDV | Cluster 2 | SEAR | Upper middle income | Reserve DID | 0 (0, 0.1) | 0 (0, 0.1) | 0 (0, 0) | 0 (0, 0.1) |
| **Mali** | MLI | Cluster 1 | AFR | Low income | Total DID | 16.3 (12.5, 23) | 16.4 (12.6, 23.2) | 15.7 (12, 22.3) | 11 (8.4, 16.8) |
| **Mali** | MLI | Cluster 1 | AFR | Low income | Access DID | 8.4 (4.6, 15) | 6.1 (2, 12.7) | 9.7 (6, 16.3) | 3.1 (-0.1, 9.2) |
| **Mali** | MLI | Cluster 1 | AFR | Low income | Watch DID | 7.4 (6.1, 9.3) | 9.9 (8.2, 12.1) | 5.2 (4.2, 7) | 7.4 (6.1, 9.3) |
| **Mali** | MLI | Cluster 1 | AFR | Low income | Reserve DID | 0.3 (0.1, 1) | 0.3 (0.1, 1) | 0.6 (0.2, 1.8) | 0.3 (0.1, 1) |
| **Malta** | MLT | Cluster 4 | EUR | High income | Total DID | 9.2 (8, 10.6) | 9.2 (8, 10.6) | 9.2 (8, 10.6) | 9.2 (8, 10.6) |

| | | | | | | | | | |
|---|---|---|---|---|---|---|---|---|---|
| **Malta** | MLT | Cluster 4 | EUR | High income | Access DID | 8.4 (7.2, 9.8) | 8.2 (7, 9.6) | 8.6 (7.4, 10) | 8.4 (7.2, 9.8) |
| **Malta** | MLT | Cluster 4 | EUR | High income | Watch DID | 0.7 (0.7, 0.8) | 1 (0.9, 1.1) | 0.6 (0.5, 0.6) | 0.7 (0.7, 0.8) |
| **Malta** | MLT | Cluster 4 | EUR | High income | Reserve DID | 0 (0, 0.1) | 0 (0, 0.1) | 0 (0, 0.1) | 0 (0, 0.1) |
| **Mauritania** | MRT | Cluster 1 | AFR | Lower middle income | Total DID | 13.2 (10.1, 19.1) | 13.3 (10.2, 19.1) | 12.9 (9.9, 18.7) | 9 (6.7, 13.7) |
| **Mauritania** | MRT | Cluster 1 | AFR | Lower middle income | Access DID | 7 (3.6, 12.6) | 5.5 (1.9, 10.9) | 7.9 (4.5, 13.3) | 2.7 (-0.1, 7.8) |
| **Mauritania** | MRT | Cluster 1 | AFR | Lower middle income | Watch DID | 6 (4.7, 8.6) | 7.6 (6.3, 10.3) | 4.8 (3.5, 7.4) | 6 (4.7, 8.6) |
| **Mauritania** | MRT | Cluster 1 | AFR | Lower middle income | Reserve DID | 0.1 (0, 0.4) | 0.1 (0, 0.4) | 0.2 (0.1, 0.6) | 0.1 (0, 0.4) |
| **Mauritius** | MUS | Cluster 3 | AFR | Upper middle income | Total DID | 15 (11.7, 20.8) | 15 (11.7, 20.8) | 15.2 (11.8, 21) | 10.1 (7.7, 15.5) |
| **Mauritius** | MUS | Cluster 3 | AFR | Upper middle income | Access DID | 13.5 (10.1, 19.2) | 12.9 (9.5, 18.6) | 13.9 (10.5, 19.7) | 8.5 (6.1, 13.9) |
| **Mauritius** | MUS | Cluster 3 | AFR | Upper middle income | Watch DID | 1.6 (1.4, 1.8) | 2.2 (2, 2.4) | 1.2 (1.1, 1.4) | 1.6 (1.4, 1.8) |
| **Mauritius** | MUS | Cluster 3 | AFR | Upper middle income | Reserve DID | 0 (0, 0) | 0 (0, 0) | 0 (0, 0.1) | 0 (0, 0) |
| **Mexico** | MEX | Cluster 3 | AMR | Upper middle income | Total DID | 17.9 (13.7, 25.5) | 17.9 (13.7, 25.5) | 18.1 (13.9, 25.8) | 12 (9.1, 18.7) |
| **Mexico** | MEX | Cluster 3 | AMR | Upper middle income | Access DID | 16.3 (12.2, 24) | 15.7 (11.6, 23.4) | 16.7 (12.5, 24.6) | 10.4 (7.5, 17.1) |
| **Mexico** | MEX | Cluster 3 | AMR | Upper middle income | Watch DID | 1.5 (1.4, 1.7) | 2.1 (1.9, 2.3) | 1.2 (1.1, 1.4) | 1.5 (1.4, 1.7) |
| **Mexico** | MEX | Cluster 3 | AMR | Upper middle income | Reserve DID | 0.1 (0, 0.1) | 0.1 (0, 0.1) | 0.1 (0, 0.2) | 0.1 (0, 0.1) |
| **Micronesia, Fed. Sts.** | FSM | Cluster 2 | WPR | Lower middle income | Total DID | 18.3 (14.1, 25.9) | 18.3 (14.1, 25.9) | 18.3 (14, 25.9) | 12.3 (9.2, 19.2) |
| **Micronesia, Fed. Sts.** | FSM | Cluster 2 | WPR | Lower middle income | Access DID | 16.3 (12, 24) | 15.4 (11.1, 23.1) | 16.9 (12.7, 24.6) | 10.3 (7.2, 17.3) |
| **Micronesia, Fed. Sts.** | FSM | Cluster 2 | WPR | Lower middle income | Watch DID | 1.9 (1.7, 2.4) | 2.9 (2.5, 3.4) | 1.4 (1.1, 1.8) | 1.9 (1.7, 2.4) |
| **Micronesia, Fed. Sts.** | FSM | Cluster 2 | WPR | Lower middle income | Reserve DID | 0 (0, 0.1) | 0 (0, 0.1) | 0 (0, 0) | 0 (0, 0.1) |
| **Moldova** | MDA | Cluster 4 | EUR | Upper middle income | Total DID | 14.3 (12.4, 16.6) | 14.3 (12.4, 16.6) | 14.2 (12.3, 16.6) | 14.3 (12.4, 16.6) |
| **Moldova** | MDA | Cluster 4 | EUR | Upper middle income | Access DID | 8.6 (5.4, 11.4) | 8.1 (4.9, 10.9) | 12.5 (10.6, 14.8) | 8.6 (5.4, 11.4) |
| **Moldova** | MDA | Cluster 4 | EUR | Upper middle income | Watch DID | 2.7 (2.4, 3) | 3.2 (2.9, 3.5) | 1.6 (1.4, 1.9) | 2.7 (2.4, 3) |
| **Moldova** | MDA | Cluster 4 | EUR | Upper middle income | Reserve DID | 3 (1.5, 5.5) | 3 (1.5, 5.5) | 0.1 (0, 0.2) | 3 (1.5, 5.5) |

| | | | | | | | | | |
|---|---|---|---|---|---|---|---|---|---|
| **Mongolia** | MNG | Cluster 2 | WPR | Lower middle income | Total DID | 10.5 (8, 14.9) | 10.5 (8, 14.9) | 10.6 (8.1, 15) | 7.1 (5.4, 10.9) |
| **Mongolia** | MNG | Cluster 2 | WPR | Lower middle income | Access DID | 8 (5.5, 12.5) | 7.7 (5.1, 12.2) | 9.3 (6.9, 13.8) | 4.6 (2.5, 8.4) |
| **Mongolia** | MNG | Cluster 2 | WPR | Lower middle income | Watch DID | 1.6 (1.4, 2) | 2 (1.7, 2.3) | 1.2 (1, 1.4) | 1.6 (1.4, 2) |
| **Mongolia** | MNG | Cluster 2 | WPR | Lower middle income | Reserve DID | 0.7 (0.3, 2.1) | 0.7 (0.3, 2.1) | 0 (0, 0.1) | 0.7 (0.3, 2.1) |
| **Montenegro** | MNE | Cluster 4 | EUR | Upper middle income | Total DID | 8.1 (6.9, 9.4) | 8.1 (7, 9.4) | 8.1 (6.9, 9.4) | 8.1 (6.9, 9.4) |
| **Montenegro** | MNE | Cluster 4 | EUR | Upper middle income | Access DID | 7.2 (6, 8.5) | 7 (5.9, 8.3) | 7.3 (6.2, 8.6) | 7.2 (6, 8.5) |
| **Montenegro** | MNE | Cluster 4 | EUR | Upper middle income | Watch DID | 0.9 (0.8, 1) | 1.1 (1, 1.2) | 0.8 (0.7, 0.9) | 0.9 (0.8, 1) |
| **Montenegro** | MNE | Cluster 4 | EUR | Upper middle income | Reserve DID | 0 (0, 0) | 0 (0, 0) | 0 (0, 0) | 0 (0, 0) |
| **Morocco** | MAR | Cluster 3 | EMR | Lower middle income | Total DID | 16.6 (14, 22.9) | 16.6 (14, 22.9) | 16.6 (14, 22.9) | 11.4 (8.5, 17.3) |
| **Morocco** | MAR | Cluster 3 | EMR | Lower middle income | Access DID | 14.9 (12.4, 21.2) | 14.4 (11.8, 20.7) | 15.3 (12.8, 21.7) | 9.7 (6.8, 15.7) |
| **Morocco** | MAR | Cluster 3 | EMR | Lower middle income | Watch DID | 1.5 (1.4, 1.8) | 2.1 (1.9, 2.3) | 1.2 (1, 1.4) | 1.5 (1.4, 1.8) |
| **Morocco** | MAR | Cluster 3 | EMR | Lower middle income | Reserve DID | 0.1 (0, 0.3) | 0.1 (0, 0.3) | 0 (0, 0.1) | 0.1 (0, 0.3) |
| **Mozambique** | MOZ | Cluster 1 | AFR | Low income | Total DID | 16 (12.3, 22.8) | 16 (12.3, 22.9) | 15.6 (12, 22.4) | 10.7 (8.1, 17) |
| **Mozambique** | MOZ | Cluster 1 | AFR | Low income | Access DID | 10.3 (6.3, 17.3) | 8.6 (4.5, 15.6) | 12.4 (8.8, 19.2) | 5 (1.9, 10.9) |
| **Mozambique** | MOZ | Cluster 1 | AFR | Low income | Watch DID | 4.5 (3.9, 5.5) | 6.3 (5.5, 7.4) | 3 (2.5, 3.9) | 4.5 (3.9, 5.5) |
| **Mozambique** | MOZ | Cluster 1 | AFR | Low income | Reserve DID | 1 (0.4, 2.6) | 1 (0.4, 2.6) | 0.1 (0.1, 0.3) | 1 (0.4, 2.6) |
| **Myanmar** | MMR | Cluster 2 | SEAR | Lower middle income | Total DID | 14.2 (10.9, 20.2) | 14.2 (10.9, 20.3) | 14.1 (10.7, 20.1) | 9.7 (7.2, 14.8) |
| **Myanmar** | MMR | Cluster 2 | SEAR | Lower middle income | Access DID | 10.4 (6.8, 16.4) | 8.5 (4.9, 14.6) | 12.1 (8.8, 18) | 5.9 (3.1, 11.1) |
| **Myanmar** | MMR | Cluster 2 | SEAR | Lower middle income | Watch DID | 3.1 (2.8, 3.5) | 5 (4.4, 5.7) | 1.9 (1.7, 2.3) | 3.1 (2.8, 3.5) |
| **Myanmar** | MMR | Cluster 2 | SEAR | Lower middle income | Reserve DID | 0.7 (0.3, 1.6) | 0.7 (0.3, 1.6) | 0.1 (0, 0.1) | 0.7 (0.3, 1.6) |
| **Namibia** | NAM | Cluster 2 | AFR | Upper middle income | Total DID | 18 (14, 25.6) | 18 (14, 25.6) | 17.8 (13.8, 25.4) | 12.2 (9.2, 18.9) |
| **Namibia** | NAM | Cluster 2 | AFR | Upper middle income | Access DID | 12.5 (8.1, 20) | 11.8 (7.4, 19.3) | 14.6 (10.5, 22.1) | 6.7 (3.2, 13.3) |
| **Namibia** | NAM | Cluster 2 | AFR | Upper middle income | Watch DID | 4.1 (3.5, 5) | 4.8 (4.1, 5.8) | 3.1 (2.6, 4) | 4.1 (3.5, 5) |

| | | | | | | | | | |
|---|---|---|---|---|---|---|---|---|---|
| **Namibia** | NAM | Cluster 2 | AFR | Upper middle income | Reserve DID | 1.3 (0.5, 3) | 1.3 (0.5, 3) | 0.1 (0, 0.3) | 1.3 (0.5, 3) |
| **Nepal** | NPL | Cluster 2 | SEAR | Lower middle income | Total DID | 17.9 (13.9, 25.4) | 17.9 (13.9, 25.4) | 17.8 (13.8, 25.2) | 12 (9.2, 18.8) |
| **Nepal** | NPL | Cluster 2 | SEAR | Lower middle income | Access DID | 12.8 (8.8, 20.4) | 8.9 (4.7, 16.6) | 15 (11.1, 22.3) | 7 (4, 13.8) |
| **Nepal** | NPL | Cluster 2 | SEAR | Lower middle income | Watch DID | 4.4 (3.9, 5.2) | 8.4 (7.3, 9.7) | 2.5 (2.1, 3.2) | 4.4 (3.9, 5.2) |
| **Nepal** | NPL | Cluster 2 | SEAR | Lower middle income | Reserve DID | 0.5 (0.2, 1.2) | 0.5 (0.2, 1.2) | 0.1 (0, 0.2) | 0.5 (0.2, 1.2) |
| **Netherlands** | NLD | Cluster 4 | EUR | High income | Total DID | 10.7 (9.2, 12.4) | 10.7 (9.2, 12.4) | 10.6 (9.2, 12.4) | 10.7 (9.2, 12.4) |
| **Netherlands** | NLD | Cluster 4 | EUR | High income | Access DID | 9.6 (8.1, 11.3) | 9.3 (7.8, 11.1) | 9.8 (8.3, 11.5) | 9.6 (8.1, 11.3) |
| **Netherlands** | NLD | Cluster 4 | EUR | High income | Watch DID | 1.1 (0.9, 1.2) | 1.4 (1.2, 1.5) | 0.8 (0.8, 1) | 1.1 (0.9, 1.2) |
| **Netherlands** | NLD | Cluster 4 | EUR | High income | Reserve DID | 0 (0, 0) | 0 (0, 0) | 0 (0, 0) | 0 (0, 0) |
| **New Zealand** | NZL | Cluster 4 | WPR | High income | Total DID | 14.3 (12.5, 16.4) | 14.3 (12.5, 16.4) | 14.3 (12.5, 16.4) | 14.3 (12.5, 16.4) |
| **New Zealand** | NZL | Cluster 4 | WPR | High income | Access DID | 13.3 (11.5, 15.4) | 13 (11.2, 15.1) | 13.4 (11.6, 15.5) | 13.3 (11.5, 15.4) |
| **New Zealand** | NZL | Cluster 4 | WPR | High income | Watch DID | 1 (0.9, 1.1) | 1.3 (1.2, 1.4) | 0.8 (0.8, 0.9) | 1 (0.9, 1.1) |
| **New Zealand** | NZL | Cluster 4 | WPR | High income | Reserve DID | 0 (0, 0) | 0 (0, 0) | 0 (0, 0.1) | 0 (0, 0) |
| **Nicaragua** | NIC | Cluster 3 | AMR | Lower middle income | Total DID | 14.9 (11.5, 21) | 14.9 (11.5, 21) | 14.8 (11.4, 21) | 10 (7.6, 15.7) |
| **Nicaragua** | NIC | Cluster 3 | AMR | Lower middle income | Access DID | 13.4 (10, 19.5) | 13 (9.6, 19.1) | 13.7 (10.3, 19.8) | 8.5 (6.1, 14.1) |
| **Nicaragua** | NIC | Cluster 3 | AMR | Lower middle income | Watch DID | 1.4 (1.3, 1.7) | 1.8 (1.6, 2) | 1.1 (1, 1.3) | 1.4 (1.3, 1.7) |
| **Nicaragua** | NIC | Cluster 3 | AMR | Lower middle income | Reserve DID | 0 (0, 0.1) | 0 (0, 0.1) | 0 (0, 0.1) | 0 (0, 0.1) |
| **Niger** | NER | Cluster 1 | AFR | Low income | Total DID | 17.4 (13.2, 25) | 17.5 (13.3, 25.1) | 16.9 (12.7, 24.4) | 11.7 (8.9, 18) |
| **Niger** | NER | Cluster 1 | AFR | Low income | Access DID | 9 (4.6, 16.5) | 6.7 (2.1, 14.2) | 10.4 (6, 17.6) | 3.4 (-0.1, 9.4) |
| **Niger** | NER | Cluster 1 | AFR | Low income | Watch DID | 7.8 (6.2, 10.6) | 10.3 (8.4, 13.1) | 5.7 (4.3, 8.3) | 7.8 (6.2, 10.6) |
| **Niger** | NER | Cluster 1 | AFR | Low income | Reserve DID | 0.3 (0.1, 0.9) | 0.3 (0.1, 0.9) | 0.5 (0.2, 1.6) | 0.3 (0.1, 0.9) |
| **Nigeria** | NGA | Cluster 1 | AFR | Lower middle income | Total DID | 20.4 (15.7, 28.9) | 20.4 (15.8, 29.1) | 19.9 (15.3, 28.4) | 13.7 (10.4, 21.1) |
| **Nigeria** | NGA | Cluster 1 | AFR | Lower middle income | Access DID | 11.5 (6.7, 19.8) | 9.2 (4.2, 17.5) | 13.5 (8.8, 21.6) | 5.1 (0.9, 12.6) |
| **Nigeria** | NGA | Cluster 1 | AFR | Lower middle income | Watch DID | 8.1 (6.2, 11.3) | 10.5 (8.3, 13.8) | 5.8 (4.1, 8.9) | 8.1 (6.2, 11.3) |
| **Nigeria** | NGA | Cluster 1 | AFR | Lower middle income | Reserve DID | 0.5 (0.2, 1.3) | 0.5 (0.2, 1.3) | 0.3 (0.1, 0.9) | 0.5 (0.2, 1.3) |

| | | | | | | | | | |
|---|---|---|---|---|---|---|---|---|---|
| **North Macedonia** | MKD | Cluster 4 | EUR | Upper middle income | Total DID | 9.4 (8.2, 10.9) | 9.4 (8.2, 10.9) | 9.4 (8.1, 10.9) | 9.4 (8.2, 10.9) |
| **North Macedonia** | MKD | Cluster 4 | EUR | Upper middle income | Access DID | 8.3 (7.1, 9.8) | 8.1 (6.9, 9.6) | 8.4 (7.2, 10) | 8.3 (7.1, 9.8) |
| **North Macedonia** | MKD | Cluster 4 | EUR | Upper middle income | Watch DID | 1.1 (0.9, 1.2) | 1.3 (1.1, 1.4) | 0.9 (0.8, 1.1) | 1.1 (0.9, 1.2) |
| **North Macedonia** | MKD | Cluster 4 | EUR | Upper middle income | Reserve DID | 0 (0, 0.1) | 0 (0, 0.1) | 0 (0, 0.1) | 0 (0, 0.1) |
| **Norway** | NOR | Cluster 4 | EUR | High income | Total DID | 11.7 (10.2, 13.5) | 11.7 (10.2, 13.5) | 11.7 (10.2, 13.5) | 11.7 (10.2, 13.5) |
| **Norway** | NOR | Cluster 4 | EUR | High income | Access DID | 10.6 (9, 12.4) | 10.3 (8.7, 12.1) | 10.8 (9.2, 12.6) | 10.6 (9, 12.4) |
| **Norway** | NOR | Cluster 4 | EUR | High income | Watch DID | 1.1 (1, 1.2) | 1.4 (1.3, 1.6) | 0.9 (0.8, 1) | 1.1 (1, 1.2) |
| **Norway** | NOR | Cluster 4 | EUR | High income | Reserve DID | 0 (0, 0.1) | 0 (0, 0.1) | 0 (0, 0.1) | 0 (0, 0.1) |
| **Oman** | OMN | Cluster 3 | EMR | High income | Total DID | 13.1 (10, 18.7) | 13.1 (10, 18.7) | 13.3 (10.1, 19) | 8.9 (6.7, 13.7) |
| **Oman** | OMN | Cluster 3 | EMR | High income | Access DID | 11.9 (8.7, 17.4) | 11.5 (8.4, 17.1) | 12.1 (9, 17.8) | 7.6 (5.4, 12.6) |
| **Oman** | OMN | Cluster 3 | EMR | High income | Watch DID | 1.2 (1, 1.5) | 1.6 (1.4, 1.9) | 1.1 (0.9, 1.3) | 1.2 (1, 1.5) |
| **Oman** | OMN | Cluster 3 | EMR | High income | Reserve DID | 0 (0, 0) | 0 (0, 0) | 0 (0, 0.1) | 0 (0, 0) |
| **Pakistan** | PAK | Cluster 2 | EMR | Lower middle income | Total DID | 19.9 (15.2, 28) | 19.9 (15.2, 28) | 19.8 (15.1, 28) | 13.5 (10.1, 20.6) |
| **Pakistan** | PAK | Cluster 2 | EMR | Lower middle income | Access DID | 13.2 (8.5, 21.6) | 9.6 (4.7, 18) | 15.6 (10.9, 24) | 6.8 (3.2, 14.4) |
| **Pakistan** | PAK | Cluster 2 | EMR | Lower middle income | Watch DID | 5.9 (5.2, 7) | 9.5 (8.4, 11.1) | 4 (3.4, 5) | 5.9 (5.2, 7) |
| **Pakistan** | PAK | Cluster 2 | EMR | Lower middle income | Reserve DID | 0.6 (0.3, 1.7) | 0.6 (0.3, 1.7) | 0.1 (0, 0.3) | 0.6 (0.3, 1.7) |
| **Panama** | PAN | Cluster 3 | AMR | High income | Total DID | 16.4 (12.6, 23.3) | 16.4 (12.6, 23.3) | 16.6 (12.7, 23.5) | 11.2 (8.4, 17.3) |
| **Panama** | PAN | Cluster 3 | AMR | High income | Access DID | 14.5 (10.7, 21.4) | 14.1 (10.3, 21) | 14.9 (11.1, 21.9) | 9.3 (6.5, 15.4) |
| **Panama** | PAN | Cluster 3 | AMR | High income | Watch DID | 1.9 (1.6, 2.3) | 2.2 (2, 2.6) | 1.5 (1.3, 1.9) | 1.9 (1.6, 2.3) |
| **Panama** | PAN | Cluster 3 | AMR | High income | Reserve DID | 0.1 (0, 0.2) | 0.1 (0, 0.2) | 0 (0, 0.1) | 0.1 (0, 0.2) |
| **Papua New Guinea** | PNG | Cluster 2 | WPR | Lower middle income | Total DID | 24.3 (18.6, 33.9) | 24.3 (18.7, 34) | 24.2 (18.5, 33.7) | 16.4 (12.2, 25.1) |
| **Papua New Guinea** | PNG | Cluster 2 | WPR | Lower middle income | Access DID | 18.1 (12.6, 27.9) | 14 (8, 24) | 20.3 (15, 30.1) | 10.3 (6, 19) |
| **Papua New Guinea** | PNG | Cluster 2 | WPR | Lower middle income | Watch DID | 5.7 (4.9, 7) | 9.8 (8.3, 12) | 3.6 (2.9, 4.8) | 5.7 (4.9, 7) |
| **Papua New Guinea** | PNG | Cluster 2 | WPR | Lower middle income | Reserve DID | 0.3 (0.1, 0.8) | 0.3 (0.1, 0.8) | 0 (0, 0.1) | 0.3 (0.1, 0.8) |
| **Paraguay** | PRY | Cluster 3 | AMR | Upper middle income | Total DID | 22.8 (17.8, 32.4) | 22.8 (17.8, 32.4) | 23 (17.9, 32.7) | 15.4 (11.6, 24.1) |

| | | | | | | | | | |
|---|---|---|---|---|---|---|---|---|---|
| **Paraguay** | PRY | Cluster 3 | AMR | Upper middle income | Access DID | 20.9 (15.9, 30.6) | 20.4 (15.4, 30) | 21.4 (16.2, 31.1) | 13.5 (9.7, 22.2) |
| **Paraguay** | PRY | Cluster 3 | AMR | Upper middle income | Watch DID | 1.8 (1.6, 2) | 2.3 (2.1, 2.6) | 1.4 (1.3, 1.7) | 1.8 (1.6, 2) |
| **Paraguay** | PRY | Cluster 3 | AMR | Upper middle income | Reserve DID | 0.1 (0, 0.3) | 0.1 (0, 0.3) | 0.2 (0.1, 0.4) | 0.1 (0, 0.3) |
| **Peru** | PER | Cluster 2 | AMR | Upper middle income | Total DID | 20.5 (16, 28.7) | 20.5 (16, 28.7) | 20.6 (16.1, 28.9) | 13.9 (10.3, 21.7) |
| **Peru** | PER | Cluster 2 | AMR | Upper middle income | Access DID | 17.7 (13.3, 25.9) | 17 (12.5, 25.2) | 18.7 (14.2, 27) | 11.1 (7.5, 19) |
| **Peru** | PER | Cluster 2 | AMR | Upper middle income | Watch DID | 2.3 (2.1, 2.6) | 2.9 (2.7, 3.3) | 1.8 (1.6, 2) | 2.3 (2.1, 2.6) |
| **Peru** | PER | Cluster 2 | AMR | Upper middle income | Reserve DID | 0.4 (0.2, 0.9) | 0.4 (0.2, 0.9) | 0.1 (0, 0.3) | 0.4 (0.2, 0.9) |
| **Philippines** | PHL | Cluster 2 | WPR | Lower middle income | Total DID | 20 (15.5, 27.9) | 20 (15.5, 28) | 20 (15.4, 27.9) | 13.4 (10.1, 21.1) |
| **Philippines** | PHL | Cluster 2 | WPR | Lower middle income | Access DID | 16.5 (11.9, 24.8) | 14.8 (10.2, 23.2) | 18.2 (13.5, 26.2) | 9.9 (6.5, 17.5) |
| **Philippines** | PHL | Cluster 2 | WPR | Lower middle income | Watch DID | 2.9 (2.5, 3.3) | 4.6 (4, 5.1) | 1.8 (1.5, 2.1) | 2.9 (2.5, 3.3) |
| **Philippines** | PHL | Cluster 2 | WPR | Lower middle income | Reserve DID | 0.5 (0.2, 1.5) | 0.5 (0.2, 1.5) | 0.1 (0, 0.1) | 0.5 (0.2, 1.5) |
| **Poland** | POL | Cluster 4 | EUR | High income | Total DID | 8.4 (7.3, 9.7) | 8.4 (7.3, 9.7) | 8.4 (7.3, 9.7) | 8.4 (7.3, 9.7) |
| **Poland** | POL | Cluster 4 | EUR | High income | Access DID | 7.4 (6.3, 8.7) | 7.2 (6.1, 8.5) | 7.6 (6.4, 8.8) | 7.4 (6.3, 8.7) |
| **Poland** | POL | Cluster 4 | EUR | High income | Watch DID | 1 (0.9, 1.1) | 1.2 (1, 1.3) | 0.8 (0.7, 0.9) | 1 (0.9, 1.1) |
| **Poland** | POL | Cluster 4 | EUR | High income | Reserve DID | 0 (0, 0) | 0 (0, 0) | 0 (0, 0.1) | 0 (0, 0) |
| **Portugal** | PRT | Cluster 4 | EUR | High income | Total DID | 10.6 (9.3, 12.4) | 10.6 (9.3, 12.4) | 10.6 (9.3, 12.3) | 10.6 (9.3, 12.4) |
| **Portugal** | PRT | Cluster 4 | EUR | High income | Access DID | 9.7 (8.3, 11.4) | 9.4 (8, 11.1) | 9.9 (8.5, 11.6) | 9.7 (8.3, 11.4) |
| **Portugal** | PRT | Cluster 4 | EUR | High income | Watch DID | 0.9 (0.8, 1) | 1.2 (1.1, 1.4) | 0.6 (0.6, 0.7) | 0.9 (0.8, 1) |
| **Portugal** | PRT | Cluster 4 | EUR | High income | Reserve DID | 0 (0, 0.1) | 0 (0, 0.1) | 0.1 (0, 0.1) | 0 (0, 0.1) |
| **Puerto Rico** | PRI | Cluster 3 | AMR | High income | Total DID | 15.6 (12.3, 22) | 15.6 (12.4, 22) | 15.7 (12.4, 22.1) | 10.6 (8.1, 16.3) |
| **Puerto Rico** | PRI | Cluster 3 | AMR | High income | Access DID | 13.8 (10.5, 20.2) | 13.3 (9.9, 19.7) | 14.2 (10.9, 20.6) | 8.8 (6.3, 14.5) |
| **Puerto Rico** | PRI | Cluster 3 | AMR | High income | Watch DID | 1.8 (1.6, 2) | 2.3 (2.1, 2.6) | 1.4 (1.2, 1.6) | 1.8 (1.6, 2) |
| **Puerto Rico** | PRI | Cluster 3 | AMR | High income | Reserve DID | 0 (0, 0.1) | 0 (0, 0.1) | 0.1 (0, 0.2) | 0 (0, 0.1) |
| **Qatar** | QAT | Cluster 3 | EMR | High income | Total DID | 13.6 (10.3, 19.2) | 13.6 (10.3, 19.2) | 13.7 (10.4, 19.5) | 9.1 (6.9, 14.2) |
| **Qatar** | QAT | Cluster 3 | EMR | High income | Access DID | 12.4 (9.1, 18.1) | 12 (8.8, 17.7) | 12.6 (9.3, 18.4) | 7.9 (5.7, 13.1) |
| **Qatar** | QAT | Cluster 3 | EMR | High income | Watch DID | 1.1 (0.9, 1.4) | 1.4 (1.2, 1.7) | 1 (0.9, 1.3) | 1.1 (0.9, 1.4) |

| | | | | | | | | | |
|---|---|---|---|---|---|---|---|---|---|
| **Qatar** | QAT | Cluster 3 | EMR | High income | Reserve DID | 0 (0, 0.1) | 0 (0, 0.1) | 0 (0, 0.1) | 0 (0, 0.1) |
| **Romania** | ROU | Cluster 4 | EUR | High income | Total DID | 7.8 (6.8, 9.1) | 7.8 (6.8, 9.1) | 7.8 (6.8, 9) | 7.8 (6.8, 9.1) |
| **Romania** | ROU | Cluster 4 | EUR | High income | Access DID | 6.7 (5.6, 7.8) | 6.4 (5.3, 7.6) | 7 (5.9, 8.1) | 6.7 (5.6, 7.8) |
| **Romania** | ROU | Cluster 4 | EUR | High income | Watch DID | 1.1 (0.9, 1.2) | 1.3 (1.2, 1.4) | 0.8 (0.7, 0.9) | 1.1 (0.9, 1.2) |
| **Romania** | ROU | Cluster 4 | EUR | High income | Reserve DID | 0.1 (0.1, 0.3) | 0.1 (0.1, 0.3) | 0.1 (0, 0.2) | 0.1 (0.1, 0.3) |
| **Russian Federation** | RUS | Cluster 4 | EUR | Upper middle income | Total DID | 11.2 (9.7, 13) | 11.2 (9.7, 13) | 11.2 (9.7, 13) | 11.2 (9.7, 13) |
| **Russian Federation** | RUS | Cluster 4 | EUR | Upper middle income | Access DID | 7.8 (5.8, 9.8) | 7.5 (5.4, 9.4) | 9.9 (8.5, 11.7) | 7.8 (5.8, 9.8) |
| **Russian Federation** | RUS | Cluster 4 | EUR | Upper middle income | Watch DID | 1.8 (1.6, 2) | 2.2 (2, 2.4) | 1.2 (1.1, 1.4) | 1.8 (1.6, 2) |
| **Russian Federation** | RUS | Cluster 4 | EUR | Upper middle income | Reserve DID | 1.5 (0.7, 2.9) | 1.5 (0.7, 2.9) | 0 (0, 0.1) | 1.5 (0.7, 2.9) |
| **Rwanda** | RWA | Cluster 1 | AFR | Low income | Total DID | 19.7 (15.2, 28) | 19.7 (15.2, 28) | 19.4 (15, 27.8) | 13.3 (10.1, 20.4) |
| **Rwanda** | RWA | Cluster 1 | AFR | Low income | Access DID | 14.6 (10.1, 23.1) | 13.1 (8.6, 21.6) | 15.8 (11.4, 24.1) | 8.3 (4.9, 15.2) |
| **Rwanda** | RWA | Cluster 1 | AFR | Low income | Watch DID | 4.7 (3.9, 6) | 6.2 (5.4, 7.6) | 3.5 (2.8, 4.7) | 4.7 (3.9, 6) |
| **Rwanda** | RWA | Cluster 1 | AFR | Low income | Reserve DID | 0.3 (0.1, 0.8) | 0.3 (0.1, 0.8) | 0.1 (0, 0.2) | 0.3 (0.1, 0.8) |
| **Samoa** | WSM | Cluster 2 | WPR | Upper middle income | Total DID | 21.1 (15.9, 29.8) | 21.1 (15.9, 29.8) | 21.2 (15.9, 29.9) | 14.1 (10.7, 21.8) |
| **Samoa** | WSM | Cluster 2 | WPR | Upper middle income | Access DID | 18.6 (13.2, 27.2) | 17.7 (12.4, 26.4) | 19.2 (13.8, 27.8) | 11.6 (8.2, 19.2) |
| **Samoa** | WSM | Cluster 2 | WPR | Upper middle income | Watch DID | 2.5 (2, 3.3) | 3.3 (2.8, 4.1) | 1.9 (1.5, 2.7) | 2.5 (2, 3.3) |
| **Samoa** | WSM | Cluster 2 | WPR | Upper middle income | Reserve DID | 0 (0, 0) | 0 (0, 0) | 0 (0, 0) | 0 (0, 0) |
| **Sao Tome and Principe** | STP | Cluster 2 | AFR | Lower middle income | Total DID | 14.7 (11.4, 20.9) | 14.8 (11.4, 20.9) | 14.6 (11.2, 20.7) | 9.9 (7.5, 15.5) |
| **Sao Tome and Principe** | STP | Cluster 2 | AFR | Lower middle income | Access DID | 11.4 (7.8, 17.4) | 10.1 (6.6, 16.1) | 12.2 (8.7, 18.2) | 6.5 (4.1, 12.3) |
| **Sao Tome and Principe** | STP | Cluster 2 | AFR | Lower middle income | Watch DID | 3.2 (2.7, 4.1) | 4.5 (3.8, 5.4) | 2.3 (1.8, 3.2) | 3.2 (2.7, 4.1) |
| **Sao Tome and Principe** | STP | Cluster 2 | AFR | Lower middle income | Reserve DID | 0.1 (0, 0.5) | 0.1 (0, 0.5) | 0 (0, 0) | 0.1 (0, 0.5) |
| **Saudi Arabia** | SAU | Cluster 3 | EMR | High income | Total DID | 14.1 (10.6, 19.8) | 14.1 (10.6, 19.8) | 14.2 (10.7, 20) | 9.5 (7.2, 14.7) |
| **Saudi Arabia** | SAU | Cluster 3 | EMR | High income | Access DID | 12.4 (8.9, 18) | 12 (8.5, 17.6) | 12.8 (9.2, 18.5) | 7.9 (5.5, 13.1) |
| **Saudi Arabia** | SAU | Cluster 3 | EMR | High income | Watch DID | 1.5 (1.3, 1.9) | 2 (1.7, 2.3) | 1.3 (1.1, 1.7) | 1.5 (1.3, 1.9) |
| **Saudi Arabia** | SAU | Cluster 3 | EMR | High income | Reserve DID | 0.1 (0, 0.3) | 0.1 (0, 0.3) | 0.1 (0, 0.1) | 0.1 (0, 0.3) |

| | | | | | | | | | |
|---|---|---|---|---|---|---|---|---|---|
| **Senegal** | SEN | Cluster 1 | AFR | Lower middle income | Total DID | 15.7 (11.9, 22) | 15.7 (11.9, 22.1) | 15.4 (11.6, 21.8) | 10.6 (7.8, 16.2) |
| **Senegal** | SEN | Cluster 1 | AFR | Lower middle income | Access DID | 8.5 (4.8, 14.8) | 7.1 (3.3, 13.2) | 9.4 (5.7, 15.6) | 3.4 (0.6, 9.4) |
| **Senegal** | SEN | Cluster 1 | AFR | Lower middle income | Watch DID | 6.7 (5.1, 9.7) | 8.2 (6.6, 11.2) | 5.6 (4, 8.5) | 6.7 (5.1, 9.7) |
| **Senegal** | SEN | Cluster 1 | AFR | Lower middle income | Reserve DID | 0.2 (0.1, 0.4) | 0.2 (0.1, 0.4) | 0.2 (0.1, 0.5) | 0.2 (0.1, 0.4) |
| **Serbia** | SRB | Cluster 4 | EUR | Upper middle income | Total DID | 10.2 (8.8, 11.8) | 10.2 (8.8, 11.8) | 10.2 (8.8, 11.8) | 10.2 (8.8, 11.8) |
| **Serbia** | SRB | Cluster 4 | EUR | Upper middle income | Access DID | 9 (7.6, 10.6) | 8.7 (7.3, 10.3) | 9.1 (7.8, 10.8) | 9 (7.6, 10.6) |
| **Serbia** | SRB | Cluster 4 | EUR | Upper middle income | Watch DID | 1.2 (1.1, 1.4) | 1.5 (1.3, 1.7) | 1 (0.8, 1.2) | 1.2 (1.1, 1.4) |
| **Serbia** | SRB | Cluster 4 | EUR | Upper middle income | Reserve DID | 0 (0, 0.1) | 0 (0, 0.1) | 0 (0, 0.1) | 0 (0, 0.1) |
| **Seychelles** | SYC | Cluster 2 | AFR | High income | Total DID | 16.8 (13.2, 23.5) | 16.8 (13.2, 23.5) | 16.9 (13.2, 23.7) | 11.3 (8.6, 17.8) |
| **Seychelles** | SYC | Cluster 2 | AFR | High income | Access DID | 14.4 (10.7, 21.3) | 13 (9.3, 19.8) | 15.3 (11.6, 22.2) | 9 (6.3, 15.4) |
| **Seychelles** | SYC | Cluster 2 | AFR | High income | Watch DID | 2.3 (2.1, 2.6) | 3.8 (3.4, 4.2) | 1.5 (1.3, 1.8) | 2.3 (2.1, 2.6) |
| **Seychelles** | SYC | Cluster 2 | AFR | High income | Reserve DID | 0 (0, 0) | 0 (0, 0) | 0 (0, 0) | 0 (0, 0) |
| **Sierra Leone** | SLE | Cluster 1 | AFR | Low income | Total DID | 16.8 (12.9, 24.1) | 16.9 (12.9, 24.2) | 16.3 (12.5, 23.4) | 11.3 (8.7, 17.6) |
| **Sierra Leone** | SLE | Cluster 1 | AFR | Low income | Access DID | 9.3 (5.3, 16.4) | 7.1 (3, 14.2) | 10.9 (7.1, 17.8) | 3.9 (0.8, 9.9) |
| **Sierra Leone** | SLE | Cluster 1 | AFR | Low income | Watch DID | 7.1 (5.8, 9) | 9.4 (7.8, 11.5) | 5.1 (4, 6.8) | 7.1 (5.8, 9) |
| **Sierra Leone** | SLE | Cluster 1 | AFR | Low income | Reserve DID | 0.3 (0.1, 1.2) | 0.3 (0.1, 1.2) | 0.2 (0.1, 0.6) | 0.3 (0.1, 1.2) |
| **Singapore** | SGP | Cluster 4 | WPR | High income | Total DID | 12.6 (10.8, 14.6) | 12.6 (10.8, 14.6) | 12.6 (10.8, 14.6) | 12.6 (10.8, 14.6) |
| **Singapore** | SGP | Cluster 4 | WPR | High income | Access DID | 11.8 (10, 13.9) | 11.6 (9.8, 13.7) | 12 (10.2, 14.1) | 11.8 (10, 13.9) |
| **Singapore** | SGP | Cluster 4 | WPR | High income | Watch DID | 0.7 (0.6, 0.8) | 0.9 (0.8, 1.1) | 0.5 (0.5, 0.6) | 0.7 (0.6, 0.8) |
| **Singapore** | SGP | Cluster 4 | WPR | High income | Reserve DID | 0 (0, 0.1) | 0 (0, 0.1) | 0 (0, 0.1) | 0 (0, 0.1) |
| **Slovak Republic** | SVK | Cluster 4 | EUR | High income | Total DID | 8 (6.9, 9.3) | 8 (6.9, 9.3) | 7.9 (6.8, 9.2) | 8 (6.9, 9.3) |
| **Slovak Republic** | SVK | Cluster 4 | EUR | High income | Access DID | 6.9 (5.8, 8.2) | 6.7 (5.6, 8) | 7.1 (5.9, 8.3) | 6.9 (5.8, 8.2) |
| **Slovak Republic** | SVK | Cluster 4 | EUR | High income | Watch DID | 1 (0.9, 1.2) | 1.2 (1.1, 1.4) | 0.8 (0.7, 0.9) | 1 (0.9, 1.2) |
| **Slovak Republic** | SVK | Cluster 4 | EUR | High income | Reserve DID | 0 (0, 0) | 0 (0, 0) | 0 (0, 0.1) | 0 (0, 0) |
| **Slovenia** | SVN | Cluster 4 | EUR | High income | Total DID | 7.7 (6.7, 8.9) | 7.7 (6.7, 8.9) | 7.7 (6.7, 8.8) | 7.7 (6.7, 8.9) |
| **Slovenia** | SVN | Cluster 4 | EUR | High income | Access DID | 6.8 (5.8, 7.9) | 6.6 (5.6, 7.7) | 6.9 (5.9, 8.1) | 6.8 (5.8, 7.9) |
| **Slovenia** | SVN | Cluster 4 | EUR | High income | Watch DID | 0.9 (0.8, 1) | 1.1 (1, 1.2) | 0.7 (0.6, 0.8) | 0.9 (0.8, 1) |

| | | | | | | | | | |
|---|---|---|---|---|---|---|---|---|---|
| **Slovenia** | SVN | Cluster 4 | EUR | High income | Reserve DID | 0 (0, 0) | 0 (0, 0) | 0 (0, 0) | 0 (0, 0) |
| **Solomon Islands** | SLB | Cluster 2 | WPR | Lower middle income | Total DID | 21.3 (16.3, 30) | 21.3 (16.3, 30) | 21.2 (16.1, 29.9) | 14.3 (10.7, 22.4) |
| **Solomon Islands** | SLB | Cluster 2 | WPR | Lower middle income | Access DID | 18.5 (13.5, 27.3) | 17.2 (12.1, 25.9) | 19.3 (14.3, 28) | 11.6 (8, 19.7) |
| **Solomon Islands** | SLB | Cluster 2 | WPR | Lower middle income | Watch DID | 2.6 (2.3, 3.2) | 4 (3.5, 4.8) | 1.7 (1.4, 2.3) | 2.6 (2.3, 3.2) |
| **Solomon Islands** | SLB | Cluster 2 | WPR | Lower middle income | Reserve DID | 0 (0, 0.1) | 0 (0, 0.1) | 0 (0, 0) | 0 (0, 0.1) |
| **Somalia** | SOM | Cluster 1 | EMR | Low income | Total DID | 25.8 (19.9, 37.1) | 25.9 (19.9, 37.1) | 25.6 (19.6, 36.7) | 17.4 (13.1, 27.4) |
| **Somalia** | SOM | Cluster 1 | EMR | Low income | Access DID | 16.5 (8.5, 27.6) | 14.2 (6.2, 25.3) | 20.3 (14.2, 30.8) | 8 (1.2, 17.9) |
| **Somalia** | SOM | Cluster 1 | EMR | Low income | Watch DID | 7.1 (6, 9) | 9.4 (8.1, 11.3) | 5.1 (4.1, 6.9) | 7.1 (6, 9) |
| **Somalia** | SOM | Cluster 1 | EMR | Low income | Reserve DID | 2 (0.6, 7) | 2 (0.6, 7) | 0.2 (0.1, 0.4) | 2 (0.6, 7) |
| **South Africa** | ZAF | Cluster 2 | AFR | Upper middle income | Total DID | 23.3 (18.1, 32.9) | 23.4 (18.1, 32.9) | 23.3 (18.1, 32.9) | 15.8 (12, 24.2) |
| **South Africa** | ZAF | Cluster 2 | AFR | Upper middle income | Access DID | 17.6 (12.2, 27.5) | 16.8 (11.4, 26.7) | 19.4 (14.1, 28.9) | 10.1 (5.9, 18.7) |
| **South Africa** | ZAF | Cluster 2 | AFR | Upper middle income | Watch DID | 4.8 (4.1, 5.8) | 5.6 (4.9, 6.6) | 3.8 (3.2, 4.9) | 4.8 (4.1, 5.8) |
| **South Africa** | ZAF | Cluster 2 | AFR | Upper middle income | Reserve DID | 0.8 (0.3, 2.2) | 0.8 (0.3, 2.2) | 0.1 (0, 0.2) | 0.8 (0.3, 2.2) |
| **South Sudan** | SSD | Cluster 1 | AFR | Low income | Total DID | 19 (14.6, 27.1) | 19.1 (14.7, 27.2) | 18.6 (14.2, 26.5) | 12.9 (9.9, 19.9) |
| **South Sudan** | SSD | Cluster 1 | AFR | Low income | Access DID | 10.4 (5.5, 18.6) | 8.4 (3.5, 16.8) | 12.5 (8, 20.5) | 4.2 (0.2, 11.6) |
| **South Sudan** | SSD | Cluster 1 | AFR | Low income | Watch DID | 7.7 (6.3, 10) | 9.7 (8.1, 12.1) | 5.8 (4.5, 7.9) | 7.7 (6.3, 10) |
| **South Sudan** | SSD | Cluster 1 | AFR | Low income | Reserve DID | 0.7 (0.2, 2.6) | 0.7 (0.2, 2.6) | 0.2 (0.1, 0.6) | 0.7 (0.2, 2.6) |
| **Spain** | ESP | Cluster 4 | EUR | High income | Total DID | 10.5 (8.9, 12) | 10.5 (8.9, 12) | 10.4 (8.9, 12) | 10.5 (8.9, 12) |
| **Spain** | ESP | Cluster 4 | EUR | High income | Access DID | 9.5 (8, 11.1) | 9.3 (7.7, 10.8) | 9.7 (8.2, 11.3) | 9.5 (8, 11.1) |
| **Spain** | ESP | Cluster 4 | EUR | High income | Watch DID | 0.9 (0.8, 1) | 1.2 (1.1, 1.3) | 0.7 (0.6, 0.7) | 0.9 (0.8, 1) |
| **Spain** | ESP | Cluster 4 | EUR | High income | Reserve DID | 0 (0, 0) | 0 (0, 0) | 0 (0, 0) | 0 (0, 0) |
| **Sri Lanka** | LKA | Cluster 2 | SEAR | Upper middle income | Total DID | 15.9 (12.2, 22.7) | 15.9 (12.2, 22.7) | 16 (12.2, 22.8) | 10.8 (8, 16.6) |
| **Sri Lanka** | LKA | Cluster 2 | SEAR | Upper middle income | Access DID | 13.8 (10.2, 20.6) | 12.6 (8.9, 19.5) | 14.5 (10.8, 21.3) | 8.7 (5.9, 14.6) |
| **Sri Lanka** | LKA | Cluster 2 | SEAR | Upper middle income | Watch DID | 2.1 (1.8, 2.4) | 3.3 (2.9, 3.8) | 1.4 (1.2, 1.7) | 2.1 (1.8, 2.4) |
| **Sri Lanka** | LKA | Cluster 2 | SEAR | Upper middle income | Reserve DID | 0 (0, 0.1) | 0 (0, 0.1) | 0 (0, 0.1) | 0 (0, 0.1) |
| **St. Lucia** | LCA | Cluster 3 | AMR | Upper middle income | Total DID | 15.5 (12, 21.8) | 15.5 (12, 21.8) | 15.5 (12, 21.8) | 10.5 (8, 16.2) |

| | | | | | | | | | |
|---|---|---|---|---|---|---|---|---|---|
| **St. Lucia** | LCA | Cluster 3 | AMR | Upper middle income | Access DID | 13.8 (10.3, 20.1) | 13.3 (9.9, 19.7) | 14.1 (10.7, 20.5) | 8.7 (6.3, 14.6) |
| **St. Lucia** | LCA | Cluster 3 | AMR | Upper middle income | Watch DID | 1.7 (1.6, 1.9) | 2.2 (2, 2.4) | 1.4 (1.2, 1.6) | 1.7 (1.6, 1.9) |
| **St. Lucia** | LCA | Cluster 3 | AMR | Upper middle income | Reserve DID | 0 (0, 0) | 0 (0, 0) | 0 (0, 0) | 0 (0, 0) |
| **St. Vincent and the Grenadines** | VCT | Cluster 3 | AMR | Upper middle income | Total DID | 17.6 (13.5, 24.6) | 17.6 (13.5, 24.6) | 17.6 (13.5, 24.7) | 11.9 (9, 18.3) |
| **St. Vincent and the Grenadines** | VCT | Cluster 3 | AMR | Upper middle income | Access DID | 15.7 (11.6, 22.7) | 15.2 (11.1, 22.2) | 16.2 (12, 23.3) | 10 (7.2, 16.4) |
| **St. Vincent and the Grenadines** | VCT | Cluster 3 | AMR | Upper middle income | Watch DID | 1.8 (1.7, 2.1) | 2.4 (2.1, 2.6) | 1.4 (1.2, 1.6) | 1.8 (1.7, 2.1) |
| **St. Vincent and the Grenadines** | VCT | Cluster 3 | AMR | Upper middle income | Reserve DID | 0 (0, 0) | 0 (0, 0) | 0 (0, 0) | 0 (0, 0) |
| **Sudan** | SDN | Cluster 1 | EMR | Low income | Total DID | 15.1 (11.9, 21.6) | 15.2 (11.9, 21.6) | 14.9 (11.6, 21.3) | 10.3 (7.8, 16) |
| **Sudan** | SDN | Cluster 1 | EMR | Low income | Access DID | 12.3 (8.9, 18.7) | 11.5 (8, 17.9) | 12.7 (9.3, 19) | 7.5 (4.7, 13) |
| **Sudan** | SDN | Cluster 1 | EMR | Low income | Watch DID | 2.7 (2.2, 3.6) | 3.6 (2.9, 4.5) | 2 (1.6, 2.7) | 2.7 (2.2, 3.6) |
| **Sudan** | SDN | Cluster 1 | EMR | Low income | Reserve DID | 0.1 (0, 0.3) | 0.1 (0, 0.3) | 0.2 (0.1, 0.8) | 0.1 (0, 0.3) |
| **Suriname** | SUR | Cluster 3 | AMR | Upper middle income | Total DID | 15.9 (12.3, 22.6) | 15.9 (12.3, 22.6) | 15.9 (12.3, 22.6) | 10.7 (8.2, 16.8) |
| **Suriname** | SUR | Cluster 3 | AMR | Upper middle income | Access DID | 14.1 (10.5, 20.8) | 13.7 (10, 20.4) | 14.5 (10.9, 21.2) | 9 (6.4, 15) |
| **Suriname** | SUR | Cluster 3 | AMR | Upper middle income | Watch DID | 1.7 (1.5, 1.9) | 2.2 (1.9, 2.5) | 1.2 (1.1, 1.5) | 1.7 (1.5, 1.9) |
| **Suriname** | SUR | Cluster 3 | AMR | Upper middle income | Reserve DID | 0 (0, 0.1) | 0 (0, 0.1) | 0.1 (0, 0.3) | 0 (0, 0.1) |
| **Sweden** | SWE | Cluster 4 | EUR | High income | Total DID | 11.7 (10.1, 13.5) | 11.7 (10.1, 13.5) | 11.7 (10.1, 13.5) | 11.7 (10.1, 13.5) |
| **Sweden** | SWE | Cluster 4 | EUR | High income | Access DID | 10.3 (8.8, 12.1) | 10 (8.5, 11.8) | 10.5 (8.9, 12.3) | 10.3 (8.8, 12.1) |
| **Sweden** | SWE | Cluster 4 | EUR | High income | Watch DID | 1.4 (1.2, 1.5) | 1.6 (1.5, 1.8) | 1.2 (1, 1.3) | 1.4 (1.2, 1.5) |
| **Sweden** | SWE | Cluster 4 | EUR | High income | Reserve DID | 0 (0, 0) | 0 (0, 0) | 0 (0, 0) | 0 (0, 0) |
| **Switzerland** | CHE | Cluster 4 | EUR | High income | Total DID | 11 (11, 11) | 11 (11, 11) | 11 (11, 11) | 11 (11, 11) |
| **Switzerland** | CHE | Cluster 4 | EUR | High income | Access DID | 10 (9.8, 10.1) | 9.7 (9.6, 9.9) | 10.1 (10, 10.2) | 10 (9.8, 10.1) |
| **Switzerland** | CHE | Cluster 4 | EUR | High income | Watch DID | 1 (0.9, 1.1) | 1.3 (1.1, 1.4) | 0.9 (0.8, 1) | 1 (0.9, 1.1) |
| **Switzerland** | CHE | Cluster 4 | EUR | High income | Reserve DID | 0 (0, 0) | 0 (0, 0) | 0 (0, 0) | 0 (0, 0) |
| **Syrian Arab Republic** | SYR | Cluster 2 | EMR | Low income | Total DID | 9.8 (7.5, 13.8) | 9.8 (7.5, 13.9) | 9.6 (7.3, 13.7) | 6.6 (5, 10.1) |
| **Syrian Arab Republic** | SYR | Cluster 2 | EMR | Low income | Access DID | 8.1 (5.8, 12.3) | 7.5 (5.1, 11.8) | 8.5 (6.2, 12.5) | 4.9 (3.2, 8.6) |
| **Syrian Arab Republic** | SYR | Cluster 2 | EMR | Low income | Watch DID | 1.6 (1.1, 2.3) | 2.1 (1.5, 3.2) | 1 (0.8, 1.3) | 1.6 (1.1, 2.3) |

| | | | | | | | | | |
|---|---|---|---|---|---|---|---|---|---|
| **Syrian Arab Republic** | SYR | Cluster 2 | EMR | Low income | Reserve DID | 0 (0, 0.1) | 0 (0, 0.1) | 0.1 (0, 0.4) | 0 (0, 0.1) |
| **Taiwan** | TWN | Cluster 3 | WPR | High income | Total DID | 11.1 (8.6, 15.7) | 11.1 (8.6, 15.7) | 11 (8.5, 15.6) | 7.4 (5.7, 11.4) |
| **Taiwan** | TWN | Cluster 3 | WPR | High income | Access DID | 9.2 (6.7, 13.9) | 8.7 (6.2, 13.4) | 9.6 (7.1, 14.2) | 5.6 (3.8, 9.6) |
| **Taiwan** | TWN | Cluster 3 | WPR | High income | Watch DID | 1.8 (1.6, 2) | 2.2 (2, 2.5) | 1.3 (1.1, 1.5) | 1.8 (1.6, 2) |
| **Taiwan** | TWN | Cluster 3 | WPR | High income | Reserve DID | 0.1 (0, 0.3) | 0.1 (0, 0.3) | 0.1 (0, 0.1) | 0.1 (0, 0.3) |
| **Tajikistan** | TJK | Cluster 2 | EUR | Lower middle income | Total DID | 9.6 (7.5, 13.6) | 9.7 (7.5, 13.7) | 9.6 (7.4, 13.5) | 6.5 (4.9, 9.9) |
| **Tajikistan** | TJK | Cluster 2 | EUR | Lower middle income | Access DID | 6.4 (4, 10.4) | 5.8 (3.4, 9.9) | 7.7 (5.4, 11.6) | 3.2 (1.3, 6.7) |
| **Tajikistan** | TJK | Cluster 2 | EUR | Lower middle income | Watch DID | 2.6 (2.1, 3.1) | 3.1 (2.6, 3.8) | 1.8 (1.5, 2.2) | 2.6 (2.1, 3.1) |
| **Tajikistan** | TJK | Cluster 2 | EUR | Lower middle income | Reserve DID | 0.7 (0.3, 1.4) | 0.7 (0.3, 1.4) | 0.1 (0, 0.2) | 0.7 (0.3, 1.4) |
| **Tanzania** | TZA | Cluster 1 | AFR | Lower middle income | Total DID | 19.1 (14.6, 26.8) | 19.1 (14.7, 26.8) | 18.9 (14.4, 26.5) | 12.8 (9.7, 19.8) |
| **Tanzania** | TZA | Cluster 1 | AFR | Lower middle income | Access DID | 13 (8.5, 20.6) | 11 (6.4, 18.6) | 14.3 (9.9, 21.8) | 6.8 (3.6, 14) |
| **Tanzania** | TZA | Cluster 1 | AFR | Lower middle income | Watch DID | 5.6 (4.7, 7.2) | 7.7 (6.7, 9.4) | 4.3 (3.4, 5.8) | 5.6 (4.7, 7.2) |
| **Tanzania** | TZA | Cluster 1 | AFR | Lower middle income | Reserve DID | 0.2 (0.1, 0.9) | 0.2 (0.1, 0.9) | 0.1 (0, 0.2) | 0.2 (0.1, 0.9) |
| **Thailand** | THA | Cluster 2 | SEAR | Upper middle income | Total DID | 20.1 (15.6, 28.2) | 20.1 (15.6, 28.2) | 20.1 (15.6, 28.3) | 13.6 (10.2, 20.6) |
| **Thailand** | THA | Cluster 2 | SEAR | Upper middle income | Access DID | 17.5 (12.9, 25.5) | 16.3 (11.7, 24.5) | 18.5 (13.8, 26.6) | 11 (7.6, 18) |
| **Thailand** | THA | Cluster 2 | SEAR | Upper middle income | Watch DID | 2.3 (2.1, 2.6) | 3.5 (3.1, 3.8) | 1.6 (1.4, 1.8) | 2.3 (2.1, 2.6) |
| **Thailand** | THA | Cluster 2 | SEAR | Upper middle income | Reserve DID | 0.2 (0.1, 0.7) | 0.2 (0.1, 0.7) | 0.1 (0, 0.4) | 0.2 (0.1, 0.7) |
| **Timor-Leste** | TLS | Cluster 2 | SEAR | Lower middle income | Total DID | 18 (13.7, 25.6) | 18 (13.7, 25.6) | 18 (13.5, 25.6) | 12.1 (9.1, 18.6) |
| **Timor-Leste** | TLS | Cluster 2 | SEAR | Lower middle income | Access DID | 14.5 (10.2, 22.2) | 12.2 (7.6, 19.8) | 15.8 (11.5, 23.4) | 8.7 (5.6, 15.2) |
| **Timor-Leste** | TLS | Cluster 2 | SEAR | Lower middle income | Watch DID | 3.3 (2.9, 3.9) | 5.7 (4.9, 6.6) | 2 (1.7, 2.5) | 3.3 (2.9, 3.9) |
| **Timor-Leste** | TLS | Cluster 2 | SEAR | Lower middle income | Reserve DID | 0.1 (0, 0.4) | 0.1 (0, 0.4) | 0 (0, 0.2) | 0.1 (0, 0.4) |
| **Togo** | TGO | Cluster 1 | AFR | Low income | Total DID | 18 (13.9, 25.5) | 18.1 (13.9, 25.5) | 17.7 (13.6, 25) | 12.3 (9.2, 19.1) |
| **Togo** | TGO | Cluster 1 | AFR | Low income | Access DID | 11.9 (7.8, 19.2) | 10.2 (6, 17.5) | 13 (8.8, 20.1) | 6.1 (2.7, 13) |
| **Togo** | TGO | Cluster 1 | AFR | Low income | Watch DID | 5.7 (4.6, 7.7) | 7.4 (6.2, 9.5) | 4.3 (3.3, 6.3) | 5.7 (4.6, 7.7) |
| **Togo** | TGO | Cluster 1 | AFR | Low income | Reserve DID | 0.3 (0.1, 0.8) | 0.3 (0.1, 0.8) | 0.3 (0.1, 0.7) | 0.3 (0.1, 0.8) |

| | | | | | | | | | |
|---|---|---|---|---|---|---|---|---|---|
| **Tonga** | TON | Cluster 2 | WPR | Upper middle income | Total DID | 21.6 (16.5, 30.5) | 21.6 (16.5, 30.5) | 21.7 (16.6, 30.7) | 14.5 (10.8, 22.7) |
| **Tonga** | TON | Cluster 2 | WPR | Upper middle income | Access DID | 18.9 (13.8, 27.9) | 17.9 (12.9, 26.9) | 19.6 (14.5, 28.7) | 11.9 (8.1, 20.3) |
| **Tonga** | TON | Cluster 2 | WPR | Upper middle income | Watch DID | 2.6 (2.2, 3.4) | 3.6 (3.1, 4.4) | 2 (1.6, 2.8) | 2.6 (2.2, 3.4) |
| **Tonga** | TON | Cluster 2 | WPR | Upper middle income | Reserve DID | 0 (0, 0) | 0 (0, 0) | 0 (0, 0) | 0 (0, 0) |
| **Trinidad and Tobago** | TTO | Cluster 3 | AMR | High income | Total DID | 14.3 (11.1, 20.5) | 14.3 (11.1, 20.5) | 14.5 (11.3, 20.8) | 9.7 (7.3, 14.9) |
| **Trinidad and Tobago** | TTO | Cluster 3 | AMR | High income | Access DID | 13 (9.8, 19) | 12.6 (9.5, 18.7) | 13.3 (10.1, 19.6) | 8.3 (6, 13.6) |
| **Trinidad and Tobago** | TTO | Cluster 3 | AMR | High income | Watch DID | 1.3 (1.2, 1.5) | 1.7 (1.5, 1.9) | 1.1 (1, 1.3) | 1.3 (1.2, 1.5) |
| **Trinidad and Tobago** | TTO | Cluster 3 | AMR | High income | Reserve DID | 0 (0, 0.1) | 0 (0, 0.1) | 0.1 (0, 0.2) | 0 (0, 0.1) |
| **Tunisia** | TUN | Cluster 3 | EMR | Lower middle income | Total DID | 13.4 (10.3, 18.9) | 13.4 (10.3, 18.9) | 13.5 (10.3, 19) | 9 (6.8, 13.9) |
| **Tunisia** | TUN | Cluster 3 | EMR | Lower middle income | Access DID | 12 (8.9, 17.5) | 11.6 (8.4, 17) | 12.3 (9.1, 17.8) | 7.5 (5.4, 12.4) |
| **Tunisia** | TUN | Cluster 3 | EMR | Lower middle income | Watch DID | 1.4 (1.2, 1.6) | 1.8 (1.6, 2.1) | 1.2 (1, 1.4) | 1.4 (1.2, 1.6) |
| **Tunisia** | TUN | Cluster 3 | EMR | Lower middle income | Reserve DID | 0 (0, 0.1) | 0 (0, 0.1) | 0 (0, 0.1) | 0 (0, 0.1) |
| **Turkiye** | TUR | Cluster 3 | EUR | Upper middle income | Total DID | 18.4 (13.9, 25.8) | 18.4 (13.9, 25.8) | 18.6 (14, 26.2) | 12.4 (9.4, 19.2) |
| **Turkiye** | TUR | Cluster 3 | EUR | Upper middle income | Access DID | 17 (12.5, 24.4) | 16.6 (12, 24) | 17.4 (12.8, 25) | 11 (8, 17.8) |
| **Turkiye** | TUR | Cluster 3 | EUR | Upper middle income | Watch DID | 1.3 (1.2, 1.5) | 1.8 (1.6, 2) | 1.1 (1, 1.3) | 1.3 (1.2, 1.5) |
| **Turkiye** | TUR | Cluster 3 | EUR | Upper middle income | Reserve DID | 0 (0, 0.1) | 0 (0, 0.1) | 0 (0, 0.1) | 0 (0, 0.1) |
| **Turkmenistan** | TKM | Cluster 2 | EUR | Upper middle income | Total DID | 8.7 (6.7, 12.5) | 8.7 (6.7, 12.5) | 8.7 (6.7, 12.5) | 5.9 (4.5, 9.1) |
| **Turkmenistan** | TKM | Cluster 2 | EUR | Upper middle income | Access DID | 6.7 (4.6, 10.4) | 6.4 (4.3, 10.1) | 7.6 (5.6, 11.3) | 3.9 (2.2, 7.1) |
| **Turkmenistan** | TKM | Cluster 2 | EUR | Upper middle income | Watch DID | 1.4 (1.3, 1.7) | 1.8 (1.5, 2) | 1.1 (0.9, 1.2) | 1.4 (1.3, 1.7) |
| **Turkmenistan** | TKM | Cluster 2 | EUR | Upper middle income | Reserve DID | 0.5 (0.2, 1.2) | 0.5 (0.2, 1.2) | 0.1 (0, 0.1) | 0.5 (0.2, 1.2) |
| **Uganda** | UGA | Cluster 1 | AFR | Low income | Total DID | 23.3 (17.8, 33.1) | 23.3 (17.8, 33.1) | 23.2 (17.7, 33.1) | 15.8 (11.9, 24.1) |
| **Uganda** | UGA | Cluster 1 | AFR | Low income | Access DID | 18.2 (12.6, 27.8) | 16.7 (11, 26.3) | 19.6 (14.1, 29.1) | 10.7 (6.3, 19) |
| **Uganda** | UGA | Cluster 1 | AFR | Low income | Watch DID | 4.7 (3.9, 6.1) | 6.2 (5.3, 7.7) | 3.5 (2.7, 4.9) | 4.7 (3.9, 6.1) |

| | | | | | | | | | |
|---|---|---|---|---|---|---|---|---|---|
| **Uganda** | UGA | Cluster 1 | AFR | Low income | Reserve DID | 0.4 (0.1, 1.1) | 0.4 (0.1, 1.1) | 0.1 (0, 0.2) | 0.4 (0.1, 1.1) |
| **Ukraine** | UKR | Cluster 4 | EUR | Lower middle income | Total DID | 10.8 (9.3, 12.4) | 10.8 (9.3, 12.4) | 10.8 (9.2, 12.3) | 10.8 (9.3, 12.4) |
| **Ukraine** | UKR | Cluster 4 | EUR | Lower middle income | Access DID | 7.1 (4.8, 9) | 6.8 (4.5, 8.7) | 9.5 (8, 11.1) | 7.1 (4.8, 9) |
| **Ukraine** | UKR | Cluster 4 | EUR | Lower middle income | Watch DID | 1.8 (1.6, 2.1) | 2.2 (2, 2.4) | 1.1 (1, 1.3) | 1.8 (1.6, 2.1) |
| **Ukraine** | UKR | Cluster 4 | EUR | Lower middle income | Reserve DID | 1.8 (0.9, 3.5) | 1.8 (0.9, 3.5) | 0.1 (0, 0.2) | 1.8 (0.9, 3.5) |
| **United Arab Emirates** | ARE | Cluster 3 | EMR | High income | Total DID | 13.5 (10.2, 19.1) | 13.5 (10.2, 19.1) | 13.7 (10.3, 19.4) | 9.2 (6.8, 14.6) |
| **United Arab Emirates** | ARE | Cluster 3 | EMR | High income | Access DID | 12.4 (9.1, 18) | 12 (8.8, 17.7) | 12.6 (9.3, 18.4) | 8 (5.6, 13.4) |
| **United Arab Emirates** | ARE | Cluster 3 | EMR | High income | Watch DID | 1.1 (1, 1.4) | 1.4 (1.3, 1.7) | 1 (0.8, 1.2) | 1.1 (1, 1.4) |
| **United Arab Emirates** | ARE | Cluster 3 | EMR | High income | Reserve DID | 0 (0, 0.1) | 0 (0, 0.1) | 0.1 (0, 0.2) | 0 (0, 0.1) |
| **United Kingdom** | GBR | Cluster 4 | EUR | High income | Total DID | 11.6 (10.1, 13.4) | 11.6 (10.1, 13.4) | 11.5 (10.1, 13.4) | 11.6 (10.1, 13.4) |
| **United Kingdom** | GBR | Cluster 4 | EUR | High income | Access DID | 10.6 (9.1, 12.5) | 10.3 (8.9, 12.2) | 10.8 (9.3, 12.6) | 10.6 (9.1, 12.5) |
| **United Kingdom** | GBR | Cluster 4 | EUR | High income | Watch DID | 0.9 (0.8, 1.1) | 1.2 (1.1, 1.4) | 0.7 (0.6, 0.7) | 0.9 (0.8, 1.1) |
| **United Kingdom** | GBR | Cluster 4 | EUR | High income | Reserve DID | 0 (0, 0) | 0 (0, 0) | 0.1 (0, 0.2) | 0 (0, 0) |
| **United States** | USA | Cluster 4 | AMR | High income | Total DID | 16.2 (14.1, 18.7) | 16.2 (14.1, 18.7) | 16.2 (14.1, 18.7) | 16.2 (14.1, 18.7) |
| **United States** | USA | Cluster 4 | AMR | High income | Access DID | 15.2 (13.2, 17.8) | 14.9 (12.9, 17.5) | 15.3 (13.2, 17.9) | 15.2 (13.2, 17.8) |
| **United States** | USA | Cluster 4 | AMR | High income | Watch DID | 0.9 (0.8, 0.9) | 1.2 (1.1, 1.3) | 0.6 (0.6, 0.7) | 0.9 (0.8, 0.9) |
| **United States** | USA | Cluster 4 | AMR | High income | Reserve DID | 0 (0, 0.1) | 0 (0, 0.1) | 0.2 (0.1, 0.4) | 0 (0, 0.1) |
| **Uruguay** | URY | Cluster 3 | AMR | High income | Total DID | 18.7 (14.4, 26.6) | 18.7 (14.4, 26.6) | 18.9 (14.6, 26.9) | 12.6 (9.5, 19.5) |
| **Uruguay** | URY | Cluster 3 | AMR | High income | Access DID | 17.6 (13.3, 25.5) | 17.2 (13, 25.1) | 18 (13.7, 26) | 11.5 (8.3, 18.4) |
| **Uruguay** | URY | Cluster 3 | AMR | High income | Watch DID | 1.1 (1, 1.2) | 1.5 (1.3, 1.6) | 0.7 (0.7, 0.8) | 1.1 (1, 1.2) |
| **Uruguay** | URY | Cluster 3 | AMR | High income | Reserve DID | 0 (0, 0.1) | 0 (0, 0.1) | 0.1 (0.1, 0.3) | 0 (0, 0.1) |
| **Uzbekistan** | UZB | Cluster 2 | EUR | Lower middle income | Total DID | 9.7 (7.5, 13.9) | 9.7 (7.5, 13.9) | 9.8 (7.5, 14) | 6.5 (4.9, 10) |
| **Uzbekistan** | UZB | Cluster 2 | EUR | Lower middle income | Access DID | 6.8 (4.4, 11.2) | 6.5 (4, 10.8) | 8.4 (6.2, 12.7) | 3.7 (1.8, 7.3) |
| **Uzbekistan** | UZB | Cluster 2 | EUR | Lower middle income | Watch DID | 1.8 (1.5, 2.1) | 2.2 (1.9, 2.5) | 1.3 (1.1, 1.5) | 1.8 (1.5, 2.1) |
| **Uzbekistan** | UZB | Cluster 2 | EUR | Lower middle income | Reserve DID | 1 (0.4, 2.1) | 1 (0.4, 2.1) | 0 (0, 0.1) | 1 (0.4, 2.1) |

| | | | | | | | | | |
|---|---|---|---|---|---|---|---|---|---|
| **Vanuatu** | VUT | Cluster 2 | WPR | Lower middle income | Total DID | 21.3 (16.3, 30) | 21.3 (16.3, 30) | 21.3 (16.2, 29.9) | 14.4 (10.8, 22.2) |
| **Vanuatu** | VUT | Cluster 2 | WPR | Lower middle income | Access DID | 18.5 (13.4, 27.1) | 17.3 (12.2, 25.8) | 19.2 (14.1, 27.8) | 11.5 (8, 19.2) |
| **Vanuatu** | VUT | Cluster 2 | WPR | Lower middle income | Watch DID | 2.8 (2.3, 3.5) | 4 (3.5, 4.7) | 2 (1.6, 2.7) | 2.8 (2.3, 3.5) |
| **Vanuatu** | VUT | Cluster 2 | WPR | Lower middle income | Reserve DID | 0 (0, 0) | 0 (0, 0) | 0 (0, 0) | 0 (0, 0) |
| **Venezuela, RB** | VEN | Cluster 3 | AMR | Lower middle income | Total DID | 16.4 (12.6, 23.3) | 16.4 (12.6, 23.3) | 16.5 (12.7, 23.5) | 11.1 (8.4, 17.2) |
| **Venezuela, RB** | VEN | Cluster 3 | AMR | Lower middle income | Access DID | 14.6 (10.7, 21.4) | 14.2 (10.3, 21) | 15.1 (11.1, 22) | 9.3 (6.6, 15.4) |
| **Venezuela, RB** | VEN | Cluster 3 | AMR | Lower middle income | Watch DID | 1.8 (1.5, 2.1) | 2.1 (1.9, 2.5) | 1.4 (1.2, 1.7) | 1.8 (1.5, 2.1) |
| **Venezuela, RB** | VEN | Cluster 3 | AMR | Lower middle income | Reserve DID | 0 (0, 0.1) | 0 (0, 0.1) | 0.1 (0, 0.1) | 0 (0, 0.1) |
| **Viet Nam** | VNM | Cluster 2 | WPR | Lower middle income | Total DID | 14.2 (10.8, 19.9) | 14.2 (10.8, 19.9) | 14.2 (10.7, 19.9) | 9.6 (7.3, 14.7) |
| **Viet Nam** | VNM | Cluster 2 | WPR | Lower middle income | Access DID | 11.7 (8.3, 17.5) | 10.5 (7.1, 16.4) | 12.8 (9.4, 18.6) | 7 (4.6, 12.2) |
| **Viet Nam** | VNM | Cluster 2 | WPR | Lower middle income | Watch DID | 2.1 (1.8, 2.4) | 3.2 (2.8, 3.7) | 1.4 (1.2, 1.7) | 2.1 (1.8, 2.4) |
| **Viet Nam** | VNM | Cluster 2 | WPR | Lower middle income | Reserve DID | 0.4 (0.2, 1.2) | 0.4 (0.2, 1.2) | 0 (0, 0.1) | 0.4 (0.2, 1.2) |
| **West Bank and Gaza** | PSE | Cluster 3 | EMR | Upper middle income | Total DID | 15.7 (12.2, 22.6) | 15.7 (12.2, 22.6) | 15.9 (12.3, 22.9) | 10.6 (7.8, 16.6) |
| **West Bank and Gaza** | PSE | Cluster 3 | EMR | Upper middle income | Access DID | 14.4 (10.9, 21.3) | 13.9 (10.4, 20.8) | 14.7 (11.1, 21.8) | 9.2 (6.5, 15.3) |
| **West Bank and Gaza** | PSE | Cluster 3 | EMR | Upper middle income | Watch DID | 1.3 (1.1, 1.6) | 1.8 (1.6, 2.1) | 1.1 (0.9, 1.4) | 1.3 (1.1, 1.6) |
| **West Bank and Gaza** | PSE | Cluster 3 | EMR | Upper middle income | Reserve DID | 0 (0, 0.1) | 0 (0, 0.1) | 0.1 (0, 0.1) | 0 (0, 0.1) |
| **Yemen, Rep.** | YEM | Cluster 1 | EMR | Low income | Total DID | 18 (13.8, 25.7) | 18 (13.8, 25.7) | 17.8 (13.6, 25.5) | 12.1 (9.1, 18.3) |
| **Yemen, Rep.** | YEM | Cluster 1 | EMR | Low income | Access DID | 15.1 (10.7, 22.8) | 14.1 (9.8, 21.8) | 15.6 (11.3, 23.2) | 9.1 (6, 15.6) |
| **Yemen, Rep.** | YEM | Cluster 1 | EMR | Low income | Watch DID | 2.8 (2.3, 3.5) | 3.7 (3.1, 4.5) | 2 (1.7, 2.5) | 2.8 (2.3, 3.5) |
| **Yemen, Rep.** | YEM | Cluster 1 | EMR | Low income | Reserve DID | 0.1 (0, 0.2) | 0.1 (0, 0.2) | 0.1 (0.1, 0.4) | 0.1 (0, 0.2) |
| **Zambia** | ZMB | Cluster 1 | AFR | Lower middle income | Total DID | 23 (18.1, 32.7) | 23.1 (18.1, 32.7) | 22.9 (18, 32.6) | 15.5 (11.8, 23.9) |
| **Zambia** | ZMB | Cluster 1 | AFR | Lower middle income | Access DID | 16.3 (11.1, 25.9) | 14.9 (9.7, 24.4) | 17.7 (12.6, 27.3) | 8.8 (4.7, 17.3) |
| **Zambia** | ZMB | Cluster 1 | AFR | Lower middle income | Watch DID | 6.1 (5, 8) | 7.6 (6.3, 9.5) | 5 (3.8, 6.8) | 6.1 (5, 8) |

| | | | | | | | | | |
|---|---|---|---|---|---|---|---|---|---|
| **Zambia** | ZMB | Cluster 1 | AFR | Lower middle income | Reserve DID | 0.5 (0.1, 1.7) | 0.5 (0.1, 1.7) | 0.1 (0, 0.3) | 0.5 (0.1, 1.7) |
| **Zimbabwe** | ZWE | Cluster 1 | AFR | Lower middle income | Total DID | 19.7 (15.1, 28.1) | 19.8 (15.1, 28.1) | 19.6 (14.9, 27.9) | 13.3 (10.1, 20.7) |
| **Zimbabwe** | ZWE | Cluster 1 | AFR | Lower middle income | Access DID | 15.6 (10.3, 23.8) | 14.9 (9.5, 23.1) | 17.3 (12.7, 25.7) | 9.2 (4.9, 16.8) |
| **Zimbabwe** | ZWE | Cluster 1 | AFR | Lower middle income | Watch DID | 3.1 (2.7, 3.7) | 3.9 (3.4, 4.5) | 2.1 (1.7, 2.5) | 3.1 (2.7, 3.7) |
| **Zimbabwe** | ZWE | Cluster 1 | AFR | Lower middle income | Reserve DID | 0.9 (0.2, 3.5) | 0.9 (0.2, 3.5) | 0.1 (0, 0.3) | 0.9 (0.2, 3.5) |

*Table 13. Estimated optimal percentage Access for each analysis scenario. Scenario 1 = primary analysis, Scenario 2 = high watch scenario, Scenario 3 = unadjusted cases & excl. TB, Scenario 4 = alternate benchmark CTA*

| CTA Name | ISO3 Code | Cluster | % Access Scenario 1 | % Access Scenario 2 | % Access Scenario 3 | % Access Scenario 4 |
|---|---|---|---|---|---|---|
| **Afghanistan** | AFG | Cluster 1 | 76 (63.2, 84) | 67.4 (53, 77.8) | 85.5 (79.3, 90) | 64.4 (46.2, 77.8) |
| **Albania** | ALB | Cluster 4 | 87.8 (85.1, 89.8) | 85.4 (82.3, 87.7) | 89.8 (87.4, 91.7) | 87.8 (85.1, 89.8) |
| **Algeria** | DZA | Cluster 3 | 90 (86.1, 93.2) | 86.7 (81.9, 90.8) | 92.1 (88.8, 94.6) | 85.3 (78.9, 90.6) |
| **Angola** | AGO | Cluster 1 | 73.5 (60.6, 81.8) | 65.9 (51.8, 76.5) | 82.7 (75.2, 88) | 60.9 (39.7, 75.9) |
| **Antigua and Barbuda** | ATG | Cluster 3 | 89.6 (86.3, 92.8) | 87 (82.9, 91) | 91.6 (88.8, 94.2) | 84.6 (79.4, 90.2) |
| **Argentina** | ARG | Cluster 3 | 94.1 (92, 95.9) | 91.9 (89.2, 94.4) | 95.4 (93.7, 96.8) | 91.2 (88, 94.3) |
| **Armenia** | ARM | Cluster 4 | 79.3 (72.6, 83.4) | 75.9 (69.1, 80.5) | 85.8 (82.6, 88.4) | 79.3 (72.6, 83.4) |
| **Australia** | AUS | Cluster 4 | 94 (92.8, 94.9) | 91.9 (90.4, 93.2) | 94.8 (93.8, 95.7) | 94 (92.8, 94.9) |
| **Austria** | AUT | Cluster 4 | 91.9 (90.4, 93.1) | 89.3 (87.3, 91) | 93.1 (91.8, 94.2) | 91.9 (90.4, 93.1) |
| **Azerbaijan** | AZE | Cluster 2 | 70.4 (53.2, 81.4) | 66.1 (48.5, 78.5) | 85.1 (79.7, 89.8) | 56.7 (28.5, 73) |
| **Bahamas, The** | BHS | Cluster 3 | 90.1 (87, 93.1) | 87.5 (83.8, 91.4) | 91.9 (88.9, 94.5) | 85.4 (80, 90.8) |
| **Bahrain** | BHR | Cluster 3 | 91.1 (87.9, 93.8) | 88.3 (84.4, 91.8) | 92.4 (89.5, 94.7) | 86.6 (81.7, 91.3) |
| **Bangladesh** | BGD | Cluster 2 | 75.9 (67.6, 83.3) | 58.6 (45.1, 71.8) | 85.7 (80.3, 90.1) | 64.2 (51.8, 77.2) |
| **Barbados** | BRB | Cluster 3 | 86.9 (82.7, 90.9) | 83.4 (78.3, 88.5) | 89.7 (86, 92.8) | 80.5 (73.8, 87.4) |
| **Belarus** | BLR | Cluster 4 | 77.5 (69.7, 82.3) | 74.6 (66.8, 79.8) | 89.1 (86.9, 90.8) | 77.5 (69.7, 82.3) |
| **Belgium** | BEL | Cluster 4 | 90.2 (88.3, 91.7) | 87.5 (85.1, 89.4) | 92.4 (91, 93.6) | 90.2 (88.3, 91.7) |
| **Belize** | BLZ | Cluster 2 | 90.4 (87.3, 93.4) | 88 (84.2, 91.7) | 92.4 (89.8, 94.8) | 85.8 (80.7, 90.8) |
| **Benin** | BEN | Cluster 1 | 65.7 (52.2, 77.1) | 53.9 (37.2, 68.9) | 73 (60.8, 82.3) | 48.7 (27.1, 67.7) |
| **Bhutan** | BTN | Cluster 2 | 73.6 (64, 81.9) | 52.1 (34.9, 66.7) | 85.3 (80, 89.8) | 60.7 (45.2, 74.9) |
| **Bolivia** | BOL | Cluster 2 | 88 (82.8, 91.9) | 84.5 (78.3, 89.4) | 92.1 (89.1, 94.5) | 82.3 (73.9, 88.8) |
| **Bosnia and Herzegovina** | BIH | Cluster 4 | 87.9 (85.5, 90) | 85.4 (82.7, 87.8) | 90 (87.8, 91.7) | 87.9 (85.5, 90) |
| **Botswana** | BWA | Cluster 2 | 75.3 (56.6, 83.9) | 71.2 (51.6, 81) | 84.3 (78, 89.4) | 63.4 (35.1, 77.2) |
| **Brazil** | BRA | Cluster 3 | 90.3 (87, 93.3) | 87.5 (83.4, 91.3) | 91.5 (88.6, 94.2) | 85.7 (80.8, 90.8) |
| **Brunei Darussalam** | BRN | Cluster 3 | 96 (94.6, 97.2) | 94.7 (92.9, 96.4) | 96.8 (95.6, 97.8) | 94 (91.8, 96.1) |
| **Bulgaria** | BGR | Cluster 4 | 86.3 (83.3, 88.6) | 83.5 (80.3, 86.1) | 89.3 (86.9, 91.1) | 86.3 (83.3, 88.6) |
| **Burkina Faso** | BFA | Cluster 1 | 61.4 (46.3, 73.2) | 34.5 (11.8, 54.6) | 75.6 (63.7, 83.4) | 42.8 (20.1, 63.1) |
| **Burundi** | BDI | Cluster 1 | 67.7 (53.7, 78.6) | 58.9 (42.5, 72) | 76.6 (66.3, 84.5) | 52.3 (28.9, 70.5) |
| **Cabo Verde** | CPV | Cluster 2 | 75.9 (67, 83.9) | 68 (57.7, 78.3) | 81.9 (73.9, 88.2) | 64.4 (48.5, 77.5) |
| **Cambodia** | KHM | Cluster 2 | 80.9 (73.6, 86.7) | 68.5 (57.3, 78) | 88 (83.1, 91.8) | 71.7 (60.4, 81.6) |
| **Cameroon** | CMR | Cluster 1 | 72.4 (60.6, 81.6) | 63.5 (49.7, 75.1) | 79.6 (69.4, 86.9) | 58.7 (41, 74.7) |
| **Canada** | CAN | Cluster 4 | 95.2 (94.3, 96) | 93.5 (92.4, 94.6) | 96.2 (95.3, 96.8) | 95.2 (94.3, 96) |
| **Central African Republic** | CAF | Cluster 1 | 75.7 (64.1, 84.3) | 69.9 (56.8, 80.2) | 82.5 (72.9, 88.5) | 64.1 (46.5, 77.7) |
| **Chad** | TCD | Cluster 1 | 53.9 (34.7, 68.7) | 40.8 (18.1, 58.9) | 64.4 (46, 76) | 31.6 (0.5, 56.3) |
| **Chile** | CHL | Cluster 3 | 95.2 (93.6, 96.6) | 93.5 (91.5, 95.4) | 95.8 (94.2, 97.1) | 92.9 (90.3, 95.5) |
| **China** | CHN | Cluster 3 | 89.1 (84.4, 92.7) | 86.6 (81.4, 90.8) | 92.6 (90, 95) | 84 (75.9, 90) |
| **Colombia** | COL | Cluster 3 | 92.2 (89.4, 94.8) | 90.5 (87.3, 93.6) | 93.7 (91.2, 95.7) | 88.5 (84.8, 92.6) |
| **Comoros** | COM | Cluster 1 | 64.6 (39.8, 77.6) | 56.5 (29.5, 71.4) | 78 (68.2, 85.7) | 47.8 (9.2, 67.5) |
| **Congo, Dem. Rep.** | COD | Cluster 1 | 78.4 (68, 86) | 72.3 (60.6, 81.5) | 86.2 (79.5, 91) | 68 (51.4, 80.3) |
| **Congo, Rep.** | COG | Cluster 1 | 80.5 (70.7, 86.7) | 75.4 (64.6, 83) | 86.3 (79.1, 90.7) | 71.1 (56.6, 81.7) |
| **Costa Rica** | CRI | Cluster 3 | 89.7 (85.8, 93) | 87.8 (83.4, 91.7) | 90.8 (87.3, 93.7) | 84.5 (78.3, 90.3) |
| **Cote d'Ivoire** | CIV | Cluster 1 | 70 (58.8, 79.8) | 60.3 (46.1, 72.3) | 78.6 (69, 86) | 55.5 (36.1, 72.2) |
| **Croatia** | HRV | Cluster 4 | 87 (84.4, 89.3) | 84.1 (81.1, 86.8) | 89 (86.6, 91.1) | 87 (84.4, 89.3) |
| **Cuba** | CUB | Cluster 3 | 89.4 (86, 92.6) | 86.4 (82.3, 90.4) | 91.3 (88.3, 93.9) | 84.2 (79, 89.9) |
| **Cyprus** | CYP | Cluster 4 | 92.4 (90.9, 93.6) | 90.5 (88.8, 92) | 93.3 (91.7, 94.5) | 92.4 (90.9, 93.6) |
| **Czechia** | CZE | Cluster 4 | 86.8 (84.2, 88.9) | 83.7 (80.7, 86.3) | 89.2 (87, 91) | 86.8 (84.2, 88.9) |
| **Denmark** | DNK | Cluster 4 | 87.9 (85.6, 89.9) | 85.2 (82.5, 87.6) | 89.7 (87.8, 91.4) | 87.9 (85.6, 89.9) |

| | | | | | | |
|---|---|---|---|---|---|---|
| **Djibouti** | DJI | Cluster 2 | 70.3 (56.7, 80.6) | 62.9 (47.4, 75.5) | 79.9 (72, 87) | 56.2 (35, 72) |
| **Dominican Republic** | DOM | Cluster 3 | 88 (83.6, 91.7) | 84.7 (78.9, 89.4) | 90.7 (87.2, 93.6) | 82.1 (75, 88.6) |
| **Ecuador** | ECU | Cluster 3 | 89.6 (85.2, 92.7) | 86.7 (81.5, 90.6) | 91.7 (89, 94.3) | 84.5 (78.4, 89.7) |
| **Egypt, Arab Rep.** | EGY | Cluster 2 | 86.3 (81.5, 90.6) | 82.2 (76.2, 87.7) | 89.2 (85.3, 92.6) | 79.9 (72.5, 87) |
| **El Salvador** | SLV | Cluster 3 | 90.1 (86.3, 93.1) | 87.8 (83.3, 91.5) | 92.1 (89, 94.5) | 85.2 (79.6, 90.5) |
| **Equatorial Guinea** | GNQ | Cluster 1 | 81.6 (72.5, 88) | 76.9 (65.7, 84.6) | 87.1 (79.9, 91.7) | 72.6 (58.9, 83.2) |
| **Eritrea** | ERI | Cluster 1 | 62.6 (43.7, 75.6) | 54.1 (33.7, 69.3) | 73.7 (61.7, 82.5) | 44.9 (14.5, 66.1) |
| **Estonia** | EST | Cluster 4 | 84.7 (81.3, 87.2) | 81.9 (78, 84.8) | 89.3 (87, 91) | 84.7 (81.3, 87.2) |
| **Eswatini** | SWZ | Cluster 2 | 57.3 (17.4, 74.1) | 52.8 (12.4, 70.9) | 79.5 (69.7, 86.4) | 37.1 (-28, 64.3) |
| **Ethiopia** | ETH | Cluster 1 | 58.3 (38, 72.8) | 46.3 (24, 64.5) | 68.3 (50.7, 79.9) | 38.3 (7.9, 61.2) |
| **Fiji** | FJI | Cluster 2 | 88 (83.5, 91.8) | 83.6 (77.5, 88.6) | 91.1 (87.3, 94.1) | 82.2 (75.4, 88.8) |
| **Finland** | FIN | Cluster 4 | 92.1 (90.7, 93.3) | 89.5 (87.8, 91.1) | 93.5 (92.4, 94.6) | 92.1 (90.7, 93.3) |
| **France** | FRA | Cluster 4 | 90.1 (87.9, 91.8) | 87.8 (85.2, 89.8) | 91.8 (89.9, 93.3) | 90.1 (87.9, 91.8) |
| **Gabon** | GAB | Cluster 2 | 83.9 (76.7, 89.5) | 79.7 (71.3, 86.6) | 88.6 (83.4, 92.3) | 76.2 (64.1, 85.2) |
| **Gambia, The** | GMB | Cluster 1 | 65.9 (52.6, 76.1) | 55.1 (38.8, 68.4) | 74.2 (61.9, 82.7) | 49.5 (28.6, 68) |
| **Georgia** | GEO | Cluster 4 | 62.5 (47.3, 70.9) | 58.1 (42.8, 66.8) | 81.4 (77.5, 84.4) | 62.5 (47.3, 70.9) |
| **Germany** | DEU | Cluster 4 | 91.1 (89.6, 92.4) | 88.2 (86.2, 90) | 93 (91.8, 94.1) | 91.1 (89.6, 92.4) |
| **Ghana** | GHA | Cluster 1 | 64.8 (51, 76.1) | 50.6 (33.8, 66.2) | 74.7 (62.3, 82.9) | 47.7 (25.8, 67) |
| **Greece** | GRC | Cluster 4 | 91.4 (89.6, 92.8) | 88.9 (86.7, 90.6) | 92.7 (91, 94) | 91.4 (89.6, 92.8) |
| **Grenada** | GRD | Cluster 3 | 88.3 (84.3, 91.8) | 85.2 (80.5, 89.6) | 90.9 (87.7, 93.7) | 82.6 (76.3, 88.8) |
| **Guatemala** | GTM | Cluster 2 | 85.7 (80.4, 90.2) | 80.5 (73.7, 86.5) | 88.8 (84.5, 92.2) | 78.9 (70.4, 86.3) |
| **Guinea** | GIN | Cluster 1 | 62.1 (46.5, 73.8) | 49.5 (30.9, 64.8) | 72.1 (58, 81.7) | 43.6 (19.4, 64.9) |
| **Guinea-Bissau** | GNB | Cluster 1 | 70.3 (57.5, 80.1) | 64 (50.2, 75.4) | 75.9 (64.8, 84.2) | 55.9 (35.8, 72.9) |
| **Guyana** | GUY | Cluster 3 | 89.8 (86.1, 92.8) | 85.4 (80.5, 89.7) | 91.9 (88.8, 94.3) | 84.8 (79.2, 90.3) |
| **Haiti** | HTI | Cluster 2 | 89.6 (86, 92.8) | 85.3 (80.2, 89.9) | 92.8 (90.3, 95) | 84.7 (78.8, 90.3) |
| **Honduras** | HND | Cluster 2 | 88.5 (84.1, 92.2) | 86.4 (81.5, 90.8) | 90.8 (87.1, 93.8) | 82.8 (76.2, 89.4) |
| **Hungary** | HUN | Cluster 4 | 87.4 (84.9, 89.4) | 84.8 (82.1, 87.1) | 89.6 (87.4, 91.3) | 87.4 (84.9, 89.4) |
| **Iceland** | ISL | Cluster 4 | 91.1 (89.2, 92.6) | 89 (86.8, 90.8) | 92.3 (90.5, 93.7) | 91.1 (89.2, 92.6) |
| **India** | IND | Cluster 2 | 52.2 (31.7, 66.9) | 23.2 (-6.4, 46.1) | 72.2 (60.8, 80.3) | 28.8 (-1.3, 54.6) |
| **Indonesia** | IDN | Cluster 2 | 84.2 (78.6, 89.1) | 74.4 (65.7, 82.5) | 89.7 (85.6, 93.1) | 76.6 (67.7, 85) |
| **Iran, Islamic Rep.** | IRN | Cluster 3 | 88.5 (83.2, 92.1) | 85.4 (79.5, 89.9) | 89.9 (85, 93.1) | 82.8 (75.5, 89.2) |
| **Iraq** | IRQ | Cluster 2 | 87.5 (83.1, 91.6) | 83.5 (77.8, 88.6) | 90.4 (86.6, 93.5) | 81.5 (74.2, 88.3) |
| **Ireland** | IRL | Cluster 4 | 91.7 (90, 93.2) | 89.7 (87.6, 91.4) | 92.6 (90.9, 94) | 91.7 (90, 93.2) |
| **Israel** | ISR | Cluster 3 | 95.1 (93.5, 96.6) | 93.6 (91.4, 95.5) | 96 (94.5, 97.1) | 92.7 (90.2, 95.3) |
| **Italy** | ITA | Cluster 4 | 93.7 (92.5, 94.6) | 91.8 (90.2, 93) | 95.1 (94.1, 95.8) | 93.7 (92.5, 94.6) |
| **Jamaica** | JAM | Cluster 3 | 89.7 (86.3, 93) | 87 (82.7, 91.1) | 91.8 (88.9, 94.5) | 84.9 (79.4, 90.5) |
| **Japan** | JPN | Cluster 4 | 91.9 (90.2, 93.3) | 89.7 (87.6, 91.5) | 93.9 (92.7, 94.9) | 91.9 (90.2, 93.3) |
| **Jordan** | JOR | Cluster 3 | 90.9 (87.5, 93.7) | 87.3 (82.6, 91.1) | 92.6 (89.6, 94.8) | 86.6 (81.7, 91.3) |
| **Kazakhstan** | KAZ | Cluster 2 | 78.2 (67.3, 85.6) | 75.4 (64, 83.5) | 89.7 (86, 92.9) | 67.9 (50.1, 80.3) |
| **Kenya** | KEN | Cluster 1 | 69 (57.4, 78.6) | 57.1 (41.8, 70.7) | 76.6 (67.3, 84.2) | 53.7 (36.3, 71.1) |
| **Kiribati** | KIR | Cluster 2 | 86.1 (80.5, 90.7) | 80.7 (73.7, 87.1) | 90.6 (86.2, 93.8) | 79.6 (70.9, 86.9) |
| **Korea, Dem. People's Rep.** | PRK | Cluster 2 | 76.9 (61.5, 85.4) | 74 (58.1, 83.3) | 89.3 (85.1, 92.6) | 65.6 (41.6, 80.3) |
| **Korea, Rep.** | KOR | Cluster 4 | 94.2 (92.6, 95.3) | 92.6 (90.8, 93.9) | 95.4 (94.3, 96.2) | 94.2 (92.6, 95.3) |
| **Kuwait** | KWT | Cluster 3 | 90.7 (87.1, 93.5) | 87.9 (83.6, 91.4) | 91.8 (88.4, 94.3) | 86.1 (80.5, 91.2) |
| **Kyrgyz Republic** | KGZ | Cluster 2 | 62.5 (35.3, 76.7) | 58.9 (31.1, 74.1) | 86.2 (81.5, 90.2) | 44.5 (3.9, 67.5) |
| **Lao PDR** | LAO | Cluster 2 | 82.7 (76.1, 88.1) | 71.6 (61, 80.2) | 89 (84.6, 92.5) | 74.4 (65.3, 83.7) |
| **Latvia** | LVA | Cluster 4 | 84.2 (80.7, 86.8) | 81.1 (77.3, 84.1) | 88.7 (86.5, 90.6) | 84.2 (80.7, 86.8) |
| **Lebanon** | LBN | Cluster 3 | 89 (84.4, 92.5) | 85.4 (80, 89.9) | 90.9 (87, 94) | 83.6 (76.4, 89.6) |
| **Lesotho** | LSO | Cluster 2 | 65.3 (44.4, 77.9) | 60.5 (39, 74.2) | 80 (72.3, 86.9) | 49.1 (15, 68.8) |

| | | | | | | |
|---|---|---|---|---|---|---|
| **Liberia** | LBR | Cluster 1 | 65.7 (51.1, 76.5) | 57.1 (40.6, 70) | 72.3 (59.3, 81.7) | 48.7 (26.1, 68.3) |
| **Libya** | LBY | Cluster 3 | 89.7 (85.9, 92.8) | 86.4 (81.5, 90.5) | 91.7 (88.5, 94.3) | 84.8 (78.7, 90.3) |
| **Lithuania** | LTU | Cluster 4 | 79.3 (72.9, 83.7) | 76.1 (69.6, 80.9) | 88.2 (85.8, 90) | 79.3 (72.9, 83.7) |
| **Luxembourg** | LUX | Cluster 4 | 92.5 (91.1, 93.7) | 90.3 (88.3, 91.8) | 94.1 (92.9, 95) | 92.5 (91.1, 93.7) |
| **Madagascar** | MDG | Cluster 1 | 61.7 (46.5, 74.6) | 52.5 (35.4, 67.7) | 70.2 (57.6, 80.9) | 43.8 (19.3, 63.8) |
| **Malawi** | MWI | Cluster 1 | 73.5 (62, 82.2) | 68.2 (55.7, 78.2) | 79.3 (69, 86.2) | 60.8 (42.8, 75) |
| **Malaysia** | MYS | Cluster 2 | 84.4 (78.8, 89.4) | 76.4 (68.2, 83.9) | 88.9 (84.6, 92.5) | 76.8 (68, 85.2) |
| **Maldives** | MDV | Cluster 2 | 87.5 (83.2, 91.3) | 80.6 (73.5, 86.5) | 90.8 (87.3, 93.7) | 81.6 (74.4, 88.5) |
| **Mali** | MLI | Cluster 1 | 52.2 (34.2, 66.9) | 37.6 (14.5, 56.5) | 62.5 (46.3, 74.9) | 28.6 (-1.4, 54.2) |
| **Malta** | MLT | Cluster 4 | 91.7 (90.1, 93.1) | 89.4 (87.4, 91.2) | 93.8 (92.3, 94.9) | 91.7 (90.1, 93.1) |
| **Mauritania** | MRT | Cluster 1 | 53.6 (33.7, 68) | 41.5 (17.9, 59.3) | 61.1 (42, 74) | 31 (-0.7, 57.9) |
| **Mauritius** | MUS | Cluster 3 | 89.6 (86, 92.6) | 85.6 (81, 89.8) | 91.6 (88.6, 94.1) | 84.5 (78.6, 89.9) |
| **Mexico** | MEX | Cluster 3 | 91.1 (88.3, 93.8) | 87.9 (84.1, 91.5) | 92.6 (90, 94.8) | 87 (82.4, 91.6) |
| **Micronesia, Fed. Sts.** | FSM | Cluster 2 | 89.3 (85, 92.6) | 84.2 (78.3, 89.1) | 92.6 (89.2, 95) | 84.2 (77.8, 89.9) |
| **Moldova** | MDA | Cluster 4 | 59.8 (40, 71.7) | 56.7 (36.2, 68.6) | 87.9 (85.4, 90) | 59.8 (40, 71.7) |
| **Mongolia** | MNG | Cluster 2 | 77.2 (62.3, 85.3) | 74 (58.7, 82.9) | 88.5 (84.5, 92.1) | 65.8 (42.7, 79.9) |
| **Montenegro** | MNE | Cluster 4 | 88.6 (86, 90.5) | 86.3 (83.5, 88.5) | 90.6 (88.3, 92.2) | 88.6 (86, 90.5) |
| **Morocco** | MAR | Cluster 3 | 90.1 (87.4, 93) | 86.9 (83.7, 90.7) | 92.7 (90.7, 94.9) | 85.4 (79.5, 90.7) |
| **Mozambique** | MOZ | Cluster 1 | 64.8 (48.5, 77) | 54 (35, 69.2) | 79.6 (71.1, 86.2) | 47.6 (22.9, 67.5) |
| **Myanmar** | MMR | Cluster 2 | 73.1 (61.5, 81.8) | 59.7 (44.4, 72.7) | 85.9 (80.9, 90.2) | 60.4 (41.4, 75.3) |
| **Namibia** | NAM | Cluster 2 | 70.1 (54.9, 80.4) | 66.3 (50.5, 77.4) | 82 (74.1, 87.5) | 55.5 (32.3, 72.4) |
| **Nepal** | NPL | Cluster 2 | 72.3 (62, 80.8) | 50.2 (32, 65.7) | 85.1 (79.6, 90) | 58.8 (42.7, 73.8) |
| **Netherlands** | NLD | Cluster 4 | 89.9 (87.7, 91.7) | 87.2 (84.6, 89.4) | 91.9 (90.1, 93.4) | 89.9 (87.7, 91.7) |
| **New Zealand** | NZL | Cluster 4 | 93 (91.8, 94) | 91 (89.4, 92.2) | 94 (92.9, 94.8) | 93 (91.8, 94) |
| **Nicaragua** | NIC | Cluster 3 | 89.9 (86.4, 93) | 87.6 (83.3, 91.3) | 92.2 (89.4, 94.6) | 85.1 (79.5, 90.4) |
| **Niger** | NER | Cluster 1 | 52.8 (31.7, 68.1) | 39 (13.8, 58.8) | 62.9 (42.2, 75.2) | 29.9 (-0.8, 54.6) |
| **Nigeria** | NGA | Cluster 1 | 57.6 (40.1, 72.3) | 45.6 (24.2, 64.2) | 68.6 (52.7, 80.5) | 37.2 (8.4, 61.1) |
| **North Macedonia** | MKD | Cluster 4 | 88.1 (85.6, 90.2) | 86 (83.2, 88.5) | 90 (87.8, 91.9) | 88.1 (85.6, 90.2) |
| **Norway** | NOR | Cluster 4 | 90.2 (88.4, 91.7) | 87.6 (85.3, 89.5) | 91.9 (90.3, 93.1) | 90.2 (88.4, 91.7) |
| **Oman** | OMN | Cluster 3 | 90.6 (87.2, 93.7) | 87.7 (83.5, 91.7) | 91.7 (88.6, 94.5) | 86 (80.6, 91) |
| **Pakistan** | PAK | Cluster 2 | 66.6 (54.4, 77.4) | 48.4 (30.6, 65.1) | 79.1 (70.9, 85.8) | 51.1 (30, 68.9) |
| **Panama** | PAN | Cluster 3 | 88.3 (84.2, 91.9) | 86.1 (81.4, 90.3) | 90.4 (86.8, 93.3) | 82.7 (75.8, 88.9) |
| **Papua New Guinea** | PNG | Cluster 2 | 74.9 (65.2, 83.1) | 58.2 (41, 71.1) | 84.6 (78.1, 89.8) | 63 (48.2, 76.4) |
| **Paraguay** | PRY | Cluster 3 | 91.7 (89.1, 94.4) | 89.3 (86, 92.7) | 93 (90.4, 95.2) | 87.7 (83.3, 92.3) |
| **Peru** | PER | Cluster 2 | 86.6 (81.6, 90.8) | 83.5 (77.6, 88.6) | 90.9 (87.7, 93.7) | 80.3 (71.9, 87.4) |
| **Philippines** | PHL | Cluster 2 | 82.7 (75.5, 88.1) | 74.3 (65.1, 81.8) | 90.9 (87.6, 93.7) | 74.4 (62.7, 84.3) |
| **Poland** | POL | Cluster 4 | 88.2 (85.7, 90.1) | 85.9 (83.1, 88) | 90.3 (88, 92) | 88.2 (85.7, 90.1) |
| **Portugal** | PRT | Cluster 4 | 91 (89.2, 92.5) | 88.3 (85.9, 90.2) | 93.3 (92, 94.4) | 91 (89.2, 92.5) |
| **Puerto Rico** | PRI | Cluster 3 | 88.3 (84.6, 91.8) | 84.9 (80.2, 89.4) | 90.5 (87.4, 93.5) | 82.7 (77, 88.8) |
| **Qatar** | QAT | Cluster 3 | 91.3 (87.9, 94.2) | 89 (85.1, 92.4) | 92.2 (89, 94.7) | 87.1 (81.7, 91.9) |
| **Romania** | ROU | Cluster 4 | 84.6 (81, 87.4) | 81.7 (77.6, 84.8) | 89 (86.5, 90.9) | 84.6 (81, 87.4) |
| **Russian Federation** | RUS | Cluster 4 | 70 (55.7, 78.3) | 66.9 (52.4, 75.6) | 88.9 (86.5, 90.6) | 70 (55.7, 78.3) |
| **Rwanda** | RWA | Cluster 1 | 74.6 (64.2, 83.1) | 66.8 (54.9, 77.6) | 81.4 (73.2, 88) | 62.4 (46, 75.6) |
| **Samoa** | WSM | Cluster 2 | 88.2 (82.3, 92.3) | 84.1 (77.1, 89.4) | 90.8 (85.5, 94.3) | 82.3 (74.7, 89.1) |
| **Sao Tome and Principe** | STP | Cluster 2 | 77.5 (67.8, 84.5) | 68.8 (57, 78) | 84.1 (75.7, 89.4) | 66.4 (51.9, 79.1) |
| **Saudi Arabia** | SAU | Cluster 3 | 88.4 (83.9, 92.1) | 85.4 (80.1, 90) | 90.1 (86.1, 93.3) | 82.6 (75.2, 89) |
| **Senegal** | SEN | Cluster 1 | 55.2 (37, 69.2) | 45.7 (25.5, 61.9) | 62.1 (44, 74.3) | 33.3 (5.8, 58) |
| **Serbia** | SRB | Cluster 4 | 87.8 (85.1, 89.9) | 85.5 (82.3, 87.9) | 90.1 (87.6, 91.8) | 87.8 (85.1, 89.9) |
| **Seychelles** | SYC | Cluster 2 | 86.1 (81.7, 90.4) | 77.4 (69.9, 84.5) | 90.9 (87.7, 93.7) | 79.4 (72.7, 86.9) |

| | | | | | | |
|---|---|---|---|---|---|---|
| **Sierra Leone** | SLE | Cluster 1 | 55.5 (39.4, 69.4) | 42.2 (22.9, 60) | 67.5 (54.3, 77.6) | 34.5 (8.3, 57.9) |
| **Singapore** | SGP | Cluster 4 | 94 (92.6, 95) | 92.3 (90.5, 93.6) | 95.6 (94.5, 96.3) | 94 (92.6, 95) |
| **Slovak Republic** | SVK | Cluster 4 | 87.1 (84.3, 89.3) | 84.4 (81.1, 87) | 89.2 (86.8, 91.2) | 87.1 (84.3, 89.3) |
| **Slovenia** | SVN | Cluster 4 | 88.3 (86.1, 90.2) | 85.4 (82.7, 87.8) | 90.4 (88.6, 92.1) | 88.3 (86.1, 90.2) |
| **Solomon Islands** | SLB | Cluster 2 | 87.4 (82.8, 91.3) | 81.1 (74.1, 86.7) | 91.6 (88, 94.4) | 81.4 (73.8, 88) |
| **Somalia** | SOM | Cluster 1 | 64.2 (38.5, 77) | 55.2 (28.8, 70.8) | 79.3 (70.9, 86) | 46.4 (8.1, 68.6) |
| **South Africa** | ZAF | Cluster 2 | 75.8 (65.7, 83.9) | 72.3 (61.4, 81.4) | 83.2 (77, 88.4) | 64.1 (48.2, 77.6) |
| **South Sudan** | SSD | Cluster 1 | 55 (35.4, 70.4) | 44.7 (23.6, 62.9) | 67.5 (53.4, 78.1) | 33.2 (1.9, 59.2) |
| **Spain** | ESP | Cluster 4 | 91.3 (89.6, 92.7) | 88.7 (86.5, 90.5) | 93.2 (91.9, 94.2) | 91.3 (89.6, 92.7) |
| **Sri Lanka** | LKA | Cluster 2 | 87 (82.8, 90.9) | 79.3 (72.2, 85.7) | 91 (87.9, 93.7) | 80.6 (72.6, 87.8) |
| **St. Lucia** | LCA | Cluster 3 | 88.7 (85.3, 92.2) | 85.8 (81.6, 90.1) | 91.2 (88.5, 93.9) | 83.3 (77.3, 89.4) |
| **St. Vincent and the Grenadines** | VCT | Cluster 3 | 89.4 (85.8, 92.5) | 86.5 (82, 90.4) | 92 (89.2, 94.4) | 84.2 (79.1, 90) |
| **Sudan** | SDN | Cluster 1 | 81.5 (73.1, 87.7) | 75.5 (65.5, 83.7) | 85.1 (77.2, 90.3) | 72.3 (59.4, 82.5) |
| **Suriname** | SUR | Cluster 3 | 89.1 (85.5, 92.5) | 86.1 (81.6, 90.4) | 91.5 (88.4, 94.2) | 83.8 (77.6, 89.6) |
| **Sweden** | SWE | Cluster 4 | 88.2 (85.7, 90.1) | 85.8 (82.9, 88) | 89.8 (87.7, 91.5) | 88.2 (85.7, 90.1) |
| **Switzerland** | CHE | Cluster 4 | 90.6 (89.4, 91.5) | 88.4 (87.2, 89.5) | 92.1 (90.9, 92.9) | 90.6 (89.4, 91.5) |
| **Syrian Arab Republic** | SYR | Cluster 2 | 83.3 (74.3, 90) | 77.5 (65, 86.5) | 88.4 (82.5, 92.7) | 75.5 (60, 85.9) |
| **Tajikistan** | TJK | Cluster 2 | 66.3 (51.2, 77.3) | 60.5 (43.5, 73.5) | 80.1 (72.8, 86.6) | 50.4 (25.2, 68.4) |
| **Tanzania** | TZA | Cluster 1 | 68.4 (55.8, 78.8) | 57.6 (41.9, 70.6) | 76.3 (66.1, 84.3) | 53.2 (35.5, 71) |
| **Thailand** | THA | Cluster 2 | 87 (82.4, 91.1) | 81.3 (75.1, 87.1) | 91.4 (88.3, 94) | 80.7 (72.8, 87.8) |
| **Timor-Leste** | TLS | Cluster 2 | 81 (73.6, 86.9) | 67.8 (55.3, 77.7) | 88.5 (83.5, 92.1) | 71.8 (60.8, 82.3) |
| **Togo** | TGO | Cluster 1 | 66.7 (53.4, 77.3) | 57.2 (41.7, 70.2) | 73.9 (61.5, 82.5) | 50.3 (27.7, 69.4) |
| **Tonga** | TON | Cluster 2 | 87.6 (82.5, 91.7) | 83.1 (77.1, 88.6) | 90.7 (86.1, 93.9) | 81.6 (73.3, 88.9) |
| **Trinidad and Tobago** | TTO | Cluster 3 | 90.4 (87.3, 93.4) | 88 (84.1, 91.6) | 91.9 (89.2, 94.4) | 85.9 (80.8, 90.9) |
| **Tunisia** | TUN | Cluster 3 | 89.4 (85.9, 92.6) | 86.1 (81.5, 90.2) | 91.2 (88.1, 93.8) | 84.3 (78.6, 89.8) |
| **Türkiye** | TUR | Cluster 3 | 92.5 (89.9, 94.8) | 90.2 (86.7, 93.1) | 93.9 (91.8, 95.8) | 88.9 (85, 92.8) |
| **Turkmenistan** | TKM | Cluster 2 | 77.3 (65.2, 85.3) | 73.7 (61.1, 82.9) | 87.2 (82.5, 91.3) | 66.3 (47.2, 79.2) |
| **Uganda** | UGA | Cluster 1 | 78 (69.3, 85.3) | 71.4 (60.9, 80.4) | 84.5 (77.3, 89.5) | 67.4 (51.6, 79.7) |
| **Ukraine** | UKR | Cluster 4 | 66.3 (48, 76.8) | 63.2 (44.7, 73.7) | 88.5 (85.9, 90.5) | 66.3 (48, 76.8) |
| **United Arab Emirates** | ARE | Cluster 3 | 91.5 (88.4, 94.2) | 89.2 (85.4, 92.5) | 92.3 (89.1, 94.8) | 87.5 (82.6, 92) |
| **United Kingdom** | GBR | Cluster 4 | 91.7 (90, 93.2) | 89.4 (87.2, 91.3) | 93.6 (92.1, 94.7) | 91.7 (90, 93.2) |
| **United States** | USA | Cluster 4 | 94.3 (93.3, 95.3) | 92.4 (91.1, 93.7) | 94.9 (93.3, 95.9) | 94.3 (93.3, 95.3) |
| **Uruguay** | URY | Cluster 3 | 94.1 (92.2, 95.9) | 92 (89.4, 94.4) | 95.3 (93.6, 96.7) | 91.2 (88, 94.5) |
| **Uzbekistan** | UZB | Cluster 2 | 71.5 (55, 81.6) | 67.5 (50.3, 78.6) | 86.6 (81.7, 90.8) | 57.8 (33.2, 74.1) |
| **Vanuatu** | VUT | Cluster 2 | 87 (81.7, 90.9) | 81.2 (74.4, 86.6) | 90.7 (86.2, 93.7) | 80.6 (72.7, 87.7) |
| **Venezuela, RB** | VEN | Cluster 3 | 88.9 (84.7, 92.3) | 86.6 (81.7, 90.7) | 91 (87.4, 93.7) | 83.4 (77.1, 89.4) |
| **Viet Nam** | VNM | Cluster 2 | 82.2 (75, 88.1) | 74.1 (65, 82.5) | 90.1 (86.4, 93.1) | 73.7 (60.7, 83.6) |
| **West Bank and Gaza** | PSE | Cluster 3 | 91.5 (88.4, 94.2) | 88.3 (84.5, 92.1) | 92.7 (89.8, 95) | 87.5 (82.2, 92.2) |
| **Yemen, Rep.** | YEM | Cluster 1 | 83.9 (77.4, 89.1) | 78.7 (70.6, 85.7) | 87.7 (82.8, 91.8) | 76 (65.5, 84.7) |
| **Zambia** | ZMB | Cluster 1 | 70.9 (59.1, 80.6) | 64.5 (51.4, 76) | 77.6 (67.7, 85.5) | 56.9 (36.5, 73.5) |
| **Zimbabwe** | ZWE | Cluster 1 | 79.8 (62.2, 87) | 75.7 (57.5, 84.2) | 88.8 (84.6, 92.5) | 69.9 (43.3, 82.3) |
| **Taiwan** | TWN | Cluster 3 | 83.3 (77.7, 88.5) | 79 (72.2, 85.5) | 87.8 (83.6, 91.5) | 75.5 (66.3, 84.5) |

##### 2.3.3 Actual vs. estimated optimal scenario comparison

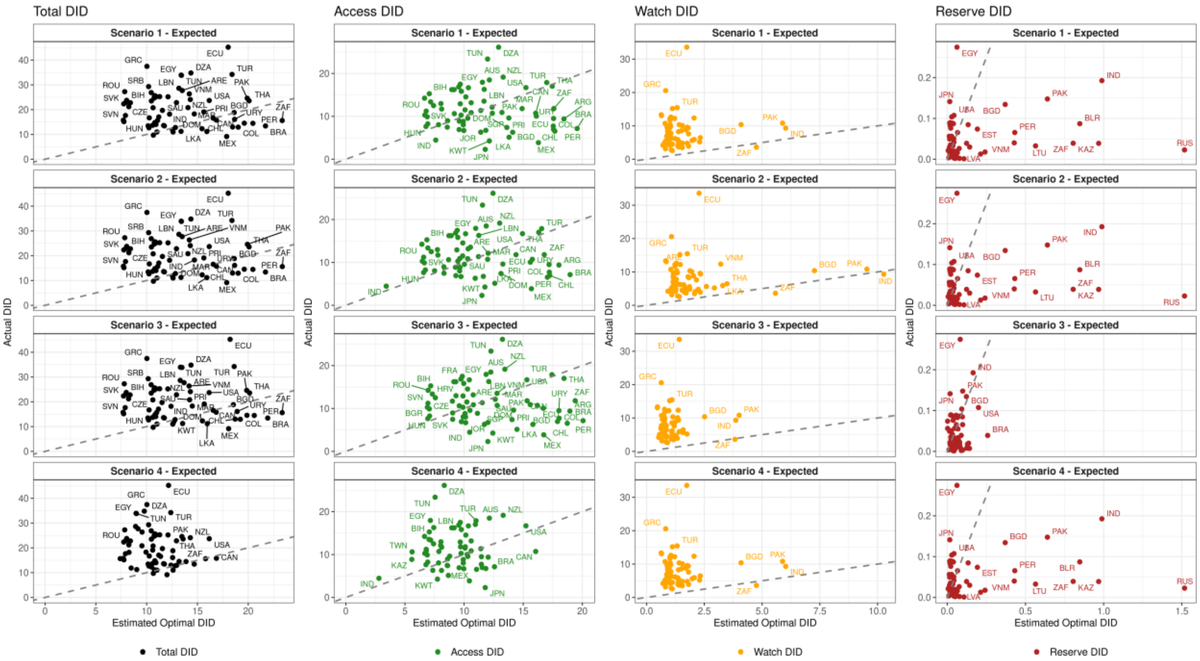

*Figure 28. Comparison of actual (IQVIA MIDAS) vs. median estimated optimal by analysis scenario (rows) and AWaRe category (columns) for CTAs in IQVIA MIDAS® with observed antibiotic use data available Dashed line indicates where observed = expected. Scenario 1 = primary analysis, Scenario 2 = high watch scenario, Scenario 3= unadjusted case counts & excl. TB, Scenario 4 = alternate benchmark CTA. Source: Actual DID based on IQVIA MIDAS® data for 2019, reflecting estimates of real-world activity. Copyright IQVIA. All Rights Reserved*

*Table 14. Estimated optimal total defined daily doses/1000 inhabitants/year (DID) and Access, Watch and Reserve DID for all four analysis scenarios compared to Actual DID in IQVIA MIDAS for 67 CTAs with available actual antibiotic use in IQVIA MIDAS. Scenario 1 = primary analysis, Scenario 2 = high watch scenario, Scenario 3 = unadjusted cases & excl. TB, Scenario 4 = alternate benchmark CTA*

| ISO3 Code | CTA Name | Cluster | AWaRe Category | Actual DID in IQVIA MIDAS* | Scenario 1 | Scenario 2 | Scenario 3 | Scenario 4 |
|---|---|---|---|---|---|---|---|---|
| **DZA** | Algeria | Cluster 3 | Total DID | 34.8 (33.7, 36.4) | 14.4 (11, 20.5) | 14.4 (11, 20.5) | 14.4 (11, 20.6) | 9.8 (7.3, 15) |
| **DZA** | Algeria | Cluster 3 | Access DID | 26.1 (25.6, 28.2) | 12.9 (9.6, 19.2) | 12.4 (9.1, 18.7) | 13.2 (9.9, 19.5) | 8.3 (5.9, 13.6) |
| **DZA** | Algeria | Cluster 3 | Watch DID | 5.6 (5.3, 5.7) | 1.4 (1.2, 1.7) | 1.9 (1.6, 2.2) | 1.1 (0.9, 1.4) | 1.4 (1.2, 1.7) |
| **DZA** | Algeria | Cluster 3 | Reserve DID | 0 (0, 0) | 0.1 (0, 0.2) | 0.1 (0, 0.2) | 0 (0, 0.1) | 0.1 (0, 0.2) |
| **ARG** | Argentina | Cluster 3 | Total DID | 14.5 (13.9, 16.1) | 19.6 (15.3, 27.6) | 19.6 (15.3, 27.6) | 19.9 (15.5, 28) | 13.2 (9.9, 20.1) |
| **ARG** | Argentina | Cluster 3 | Access DID | 9.4 (8.9, 10.7) | 18.4 (14.1, 26.5) | 18 (13.7, 26.1) | 18.9 (14.5, 27.1) | 12 (8.8, 18.9) |
| **ARG** | Argentina | Cluster 3 | Watch DID | 4.5 (4.2, 4.6) | 1.1 (1, 1.2) | 1.5 (1.4, 1.7) | 0.8 (0.7, 0.9) | 1.1 (1, 1.2) |
| **ARG** | Argentina | Cluster 3 | Reserve DID | 0 (0, 0) | 0 (0, 0.1) | 0 (0, 0.1) | 0.1 (0.1, 0.3) | 0 (0, 0.1) |
| **AUS** | Australia | Cluster 4 | Total DID | 22.8 (22.8, 22.8) | 12.9 (11.2, 14.9) | 12.9 (11.2, 14.9) | 12.9 (11.2, 14.9) | 12.9 (11.2, 14.9) |
| **AUS** | Australia | Cluster 4 | Access DID | 18.5 (18.5, 18.5) | 12.1 (10.4, 14.1) | 11.9 (10.2, 13.8) | 12.3 (10.6, 14.2) | 12.1 (10.4, 14.1) |
| **AUS** | Australia | Cluster 4 | Watch DID | 4.3 (4.3, 4.3) | 0.8 (0.7, 0.8) | 1 (0.9, 1.1) | 0.6 (0.5, 0.7) | 0.8 (0.7, 0.8) |
| **AUS** | Australia | Cluster 4 | Reserve DID | 0 (0, 0) | 0 (0, 0) | 0 (0, 0) | 0.1 (0, 0.1) | 0 (0, 0) |
| **AUT** | Austria | Cluster 4 | Total DID | 14.8 (14.8, 14.8) | 10.8 (9.3, 12.4) | 10.8 (9.3, 12.4) | 10.8 (9.3, 12.5) | 10.8 (9.3, 12.4) |
| **AUT** | Austria | Cluster 4 | Access DID | 8.8 (8.8, 8.8) | 9.9 (8.5, 11.6) | 9.7 (8.2, 11.3) | 10.1 (8.6, 11.7) | 9.9 (8.5, 11.6) |
| **AUT** | Austria | Cluster 4 | Watch DID | 5.7 (5.7, 5.7) | 0.9 (0.8, 1) | 1.1 (1, 1.3) | 0.7 (0.6, 0.8) | 0.9 (0.8, 1) |
| **AUT** | Austria | Cluster 4 | Reserve DID | 0.1 (0.1, 0.1) | 0 (0, 0) | 0 (0, 0) | 0 (0, 0) | 0 (0, 0) |
| **BGD** | Bangladesh | Cluster 2 | Total DID | 18.8 (16.8, 19.9) | 18.6 (14.4, 26.7) | 18.6 (14.4, 26.7) | 18.5 (14.2, 26.5) | 12.6 (9.6, 19.4) |
| **BGD** | Bangladesh | Cluster 2 | Access DID | 6.3 (6.1, 6.7) | 14.1 (9.9, 21.9) | 10.9 (6.6, 18.9) | 15.8 (11.6, 23.8) | 8.1 (5.1, 15) |
| **BGD** | Bangladesh | Cluster 2 | Watch DID | 10.4 (8.7, 11.4) | 4.1 (3.6, 4.8) | 7.3 (6.4, 8.3) | 2.5 (2.2, 3.1) | 4.1 (3.6, 4.8) |
| **BGD** | Bangladesh | Cluster 2 | Reserve DID | 0.1 (0.1, 0.1) | 0.4 (0.1, 1) | 0.4 (0.1, 1) | 0.1 (0, 0.4) | 0.4 (0.1, 1) |
| **BLR** | Belarus | Cluster 4 | Total DID | 23.5 (23.5, 23.5) | 10.5 (9, 12.1) | 10.5 (9, 12.1) | 10.5 (9, 12) | 10.5 (9, 12.1) |
| **BLR** | Belarus | Cluster 4 | Access DID | 13 (13, 13) | 8 (6.5, 9.8) | 7.7 (6.2, 9.5) | 9.3 (7.9, 10.9) | 8 (6.5, 9.8) |
| **BLR** | Belarus | Cluster 4 | Watch DID | 8.5 (8.5, 8.5) | 1.5 (1.4, 1.7) | 1.8 (1.6, 2) | 1.1 (1, 1.2) | 1.5 (1.4, 1.7) |
| **BLR** | Belarus | Cluster 4 | Reserve DID | 0.1 (0.1, 0.1) | 0.8 (0.5, 1.6) | 0.8 (0.5, 1.6) | 0 (0, 0.1) | 0.8 (0.5, 1.6) |
| **BEL** | Belgium | Cluster 4 | Total DID | 25.6 (25.6, 25.6) | 10.8 (9.4, 12.5) | 10.8 (9.4, 12.5) | 10.8 (9.4, 12.5) | 10.8 (9.4, 12.5) |
| **BEL** | Belgium | Cluster 4 | Access DID | 15.8 (15.8, 15.8) | 9.8 (8.4, 11.5) | 9.5 (8.1, 11.2) | 10 (8.6, 11.7) | 9.8 (8.4, 11.5) |
| **BEL** | Belgium | Cluster 4 | Watch DID | 9.8 (9.8, 9.8) | 1 (0.9, 1.2) | 1.3 (1.2, 1.5) | 0.8 (0.7, 0.9) | 1 (0.9, 1.2) |
| **BEL** | Belgium | Cluster 4 | Reserve DID | 0 (0, 0) | 0 (0, 0) | 0 (0, 0) | 0 (0, 0.1) | 0 (0, 0) |
| **BIH** | Bosnia and Herzegovina | Cluster 4 | Total DID | 23.8 (22.1, 26.2) | 8 (7, 9.2) | 8 (7, 9.2) | 8 (6.9, 9.1) | 8 (7, 9.2) |
| **BIH** | Bosnia and Herzegovina | Cluster 4 | Access DID | 15.3 (14.1, 17.6) | 7.1 (6, 8.2) | 6.9 (5.8, 8) | 7.2 (6.1, 8.4) | 7.1 (6, 8.2) |
| **BIH** | Bosnia and Herzegovina | Cluster 4 | Watch DID | 7 (6.6, 7.8) | 1 (0.8, 1.1) | 1.1 (1, 1.3) | 0.8 (0.7, 0.9) | 1 (0.8, 1.1) |
| **BIH** | Bosnia and Herzegovina | Cluster 4 | Reserve DID | 0 (0, 0.1) | 0 (0, 0) | 0 (0, 0) | 0 (0, 0.1) | 0 (0, 0) |

| ISO3 Code | CTA Name | Cluster | AWaRe Category | Actual DID in IQVIA MIDAS* | Scenario 1 | Scenario 2 | Scenario 3 | Scenario 4 |
|---|---|---|---|---|---|---|---|---|
| **BRA** | Brazil | Cluster 3 | Total DID | 13.4 (11.8, 14.5) | 21.7 (16.6, 30.7) | 21.7 (16.6, 30.8) | 21.9 (16.8, 31.1) | 14.7 (11.2, 22.6) |
| **BRA** | Brazil | Cluster 3 | Access DID | 7.2 (7, 7.6) | 19.6 (14.5, 28.6) | 19 (13.8, 28.1) | 20.1 (14.9, 29.1) | 12.6 (9.1, 20.6) |
| **BRA** | Brazil | Cluster 3 | Watch DID | 5.6 (4.5, 7) | 2 (1.8, 2.2) | 2.6 (2.4, 2.8) | 1.6 (1.4, 1.8) | 2 (1.8, 2.2) |
| **BRA** | Brazil | Cluster 3 | Reserve DID | 0 (0, 0.1) | 0.1 (0.1, 0.3) | 0.1 (0.1, 0.3) | 0.3 (0.1, 0.5) | 0.1 (0.1, 0.3) |
| **BGR** | Bulgaria | Cluster 4 | Total DID | 22.3 (22.3, 22.3) | 7.7 (6.7, 8.9) | 7.8 (6.7, 8.9) | 7.7 (6.6, 8.9) | 7.7 (6.7, 8.9) |
| **BGR** | Bulgaria | Cluster 4 | Access DID | 10.3 (10.3, 10.3) | 6.7 (5.6, 7.8) | 6.5 (5.4, 7.6) | 6.9 (5.8, 8) | 6.7 (5.6, 7.8) |
| **BGR** | Bulgaria | Cluster 4 | Watch DID | 11.7 (11.7, 11.7) | 1 (0.9, 1.2) | 1.2 (1.1, 1.4) | 0.8 (0.7, 0.9) | 1 (0.9, 1.2) |
| **BGR** | Bulgaria | Cluster 4 | Reserve DID | 0 (0, 0) | 0 (0, 0.1) | 0 (0, 0.1) | 0 (0, 0.1) | 0 (0, 0.1) |
| **CAN** | Canada | Cluster 4 | Total DID | 15.8 (15.8, 15.8) | 16.9 (14.5, 19.4) | 16.9 (14.5, 19.4) | 16.8 (14.5, 19.4) | 16.9 (14.5, 19.4) |
| **CAN** | Canada | Cluster 4 | Access DID | 10.7 (10.7, 10.7) | 16.1 (13.7, 18.6) | 15.8 (13.4, 18.3) | 16.2 (13.8, 18.7) | 16.1 (13.7, 18.6) |
| **CAN** | Canada | Cluster 4 | Watch DID | 5.1 (5.1, 5.1) | 0.8 (0.7, 0.9) | 1.1 (1, 1.2) | 0.6 (0.5, 0.6) | 0.8 (0.7, 0.9) |
| **CAN** | Canada | Cluster 4 | Reserve DID | 0 (0, 0) | 0 (0, 0) | 0 (0, 0) | 0.1 (0, 0.2) | 0 (0, 0) |
| **CHL** | Chile | Cluster 3 | Total DID | 13 (11.8, 14.5) | 18.5 (14.3, 26.1) | 18.5 (14.3, 26.1) | 18.7 (14.5, 26.4) | 12.4 (9.3, 19.4) |
| **CHL** | Chile | Cluster 3 | Access DID | 7 (6.6, 7.9) | 17.6 (13.5, 25.1) | 17.2 (13.2, 24.8) | 17.9 (13.7, 25.6) | 11.5 (8.4, 18.5) |
| **CHL** | Chile | Cluster 3 | Watch DID | 5.2 (4.7, 6.8) | 0.8 (0.8, 0.9) | 1.2 (1.1, 1.3) | 0.6 (0.6, 0.7) | 0.8 (0.8, 0.9) |
| **CHL** | Chile | Cluster 3 | Reserve DID | 0 (0, 0) | 0 (0, 0.1) | 0 (0, 0.1) | 0.1 (0.1, 0.3) | 0 (0, 0.1) |
| **COL** | Colombia | Cluster 3 | Total DID | 12.9 (11.4, 14.5) | 19 (14.5, 27.3) | 19 (14.5, 27.3) | 19.2 (14.6, 27.6) | 12.9 (10.4, 19.3) |
| **COL** | Colombia | Cluster 3 | Access DID | 7.7 (7.3, 9.1) | 17.5 (13.1, 25.8) | 17.2 (12.7, 25.4) | 18 (13.5, 26.4) | 11.5 (8.9, 17.9) |
| **COL** | Colombia | Cluster 3 | Watch DID | 4.8 (3.7, 5.9) | 1.4 (1.2, 1.6) | 1.7 (1.5, 2) | 1.1 (1, 1.4) | 1.4 (1.2, 1.6) |
| **COL** | Colombia | Cluster 3 | Reserve DID | 0 (0, 0) | 0.1 (0, 0.2) | 0.1 (0, 0.2) | 0.1 (0, 0.2) | 0.1 (0, 0.2) |
| **HRV** | Croatia | Cluster 4 | Total DID | 21.2 (21.2, 21.2) | 8.2 (7.1, 9.6) | 8.2 (7.1, 9.6) | 8.2 (7.1, 9.6) | 8.2 (7.1, 9.6) |
| **HRV** | Croatia | Cluster 4 | Access DID | 13.4 (13.4, 13.4) | 7.2 (6.1, 8.5) | 6.9 (5.8, 8.3) | 7.3 (6.2, 8.7) | 7.2 (6.1, 8.5) |
| **HRV** | Croatia | Cluster 4 | Watch DID | 7.8 (7.8, 7.8) | 1.1 (0.9, 1.2) | 1.3 (1.2, 1.5) | 0.9 (0.7, 1) | 1.1 (0.9, 1.2) |
| **HRV** | Croatia | Cluster 4 | Reserve DID | 0 (0, 0) | 0 (0, 0) | 0 (0, 0) | 0 (0, 0.1) | 0 (0, 0) |
| **CZE** | Czechia | Cluster 4 | Total DID | 17.6 (17.6, 17.6) | 7.9 (6.9, 9.1) | 7.9 (6.9, 9.1) | 7.9 (6.8, 9.1) | 7.9 (6.9, 9.1) |
| **CZE** | Czechia | Cluster 4 | Access DID | 9.9 (9.9, 9.9) | 6.9 (5.8, 8) | 6.6 (5.6, 7.8) | 7.1 (6, 8.2) | 6.9 (5.8, 8) |
| **CZE** | Czechia | Cluster 4 | Watch DID | 6.6 (6.6, 6.6) | 1 (0.9, 1.2) | 1.3 (1.1, 1.4) | 0.8 (0.7, 0.9) | 1 (0.9, 1.2) |
| **CZE** | Czechia | Cluster 4 | Reserve DID | 0 (0, 0) | 0 (0, 0) | 0 (0, 0) | 0 (0, 0.1) | 0 (0, 0) |
| **DOM** | Dominican Republic | Cluster 3 | Total DID | 12.3 (10.3, 14.7) | 15.5 (11.9, 21.5) | 15.5 (11.9, 21.5) | 15.5 (11.9, 21.6) | 10.5 (7.8, 16.3) |
| **DOM** | Dominican Republic | Cluster 3 | Access DID | 7.4 (5.4, 9.7) | 13.6 (10, 19.8) | 13.1 (9.5, 19.4) | 14.1 (10.5, 20.3) | 8.6 (5.9, 14.5) |
| **DOM** | Dominican Republic | Cluster 3 | Watch DID | 3.9 (3.6, 4.4) | 1.8 (1.5, 2.1) | 2.3 (2, 2.7) | 1.3 (1.2, 1.6) | 1.8 (1.5, 2.1) |
| **DOM** | Dominican Republic | Cluster 3 | Reserve DID | 0 (0, 0) | 0 (0, 0.1) | 0 (0, 0.1) | 0.1 (0, 0.3) | 0 (0, 0.1) |
| **ECU** | Ecuador | Cluster 3 | Total DID | 45.2 (43.4, 45.9) | 18 (14.1, 25.7) | 18 (14.1, 25.7) | 18.2 (14.2, 26) | 12.2 (9.2, 18.4) |

| ISO3 Code | CTA Name | Cluster | AWaRe Category | Actual DID in IQVIA MIDAS* | Scenario 1 | Scenario 2 | Scenario 3 | Scenario 4 |
|---|---|---|---|---|---|---|---|---|
| **ECU** | Ecuador | Cluster 3 | Access DID | 10 (9.9, 10.1) | 16.1 (12, 23.8) | 15.6 (11.4, 23.3) | 16.7 (12.6, 24.4) | 10.3 (7.2, 16.5) |
| **ECU** | Ecuador | Cluster 3 | Watch DID | 33.6 (31.6, 34.1) | 1.7 (1.5, 1.9) | 2.3 (2, 2.5) | 1.4 (1.2, 1.6) | 1.7 (1.5, 1.9) |
| **ECU** | Ecuador | Cluster 3 | Reserve DID | 0.1 (0.1, 0.1) | 0.1 (0.1, 0.5) | 0.1 (0.1, 0.5) | 0.1 (0, 0.2) | 0.1 (0.1, 0.5) |
| **EGY** | Egypt, Arab Rep. | Cluster 2 | Total DID | 33.9 (33.9, 33.9) | 13.4 (10.3, 18.9) | 13.4 (10.3, 18.9) | 13.3 (10.2, 18.8) | 9 (6.8, 14.1) |
| **EGY** | Egypt, Arab Rep. | Cluster 2 | Access DID | 17.9 (17.9, 17.9) | 11.5 (8.4, 17.2) | 11 (7.8, 16.6) | 11.9 (8.7, 17.5) | 7.2 (5, 12.2) |
| **EGY** | Egypt, Arab Rep. | Cluster 2 | Watch DID | 9.6 (9.6, 9.6) | 1.8 (1.6, 2) | 2.3 (2.1, 2.6) | 1.4 (1.2, 1.5) | 1.8 (1.6, 2) |
| **EGY** | Egypt, Arab Rep. | Cluster 2 | Reserve DID | 0.3 (0.3, 0.3) | 0.1 (0, 0.1) | 0.1 (0, 0.1) | 0.1 (0, 0.2) | 0.1 (0, 0.1) |
| **EST** | Estonia | Cluster 4 | Total DID | 13.5 (12.1, 14.9) | 10.1 (8.7, 11.8) | 10.1 (8.8, 11.8) | 10.1 (8.7, 11.8) | 10.1 (8.7, 11.8) |
| **EST** | Estonia | Cluster 4 | Access DID | 7.5 (6.9, 8.6) | 8.6 (7.2, 10.3) | 8.3 (6.9, 10) | 9 (7.6, 10.7) | 8.6 (7.2, 10.3) |
| **EST** | Estonia | Cluster 4 | Watch DID | 5.9 (4.8, 6.5) | 1.3 (1.2, 1.5) | 1.6 (1.5, 1.8) | 1.1 (1, 1.2) | 1.3 (1.2, 1.5) |
| **EST** | Estonia | Cluster 4 | Reserve DID | 0.1 (0, 0.1) | 0.2 (0.1, 0.4) | 0.2 (0.1, 0.4) | 0 (0, 0) | 0.2 (0.1, 0.4) |
| **FIN** | Finland | Cluster 4 | Total DID | 13.7 (13.7, 13.7) | 10.6 (9.3, 12.3) | 10.6 (9.3, 12.2) | 10.6 (9.3, 12.3) | 10.6 (9.3, 12.3) |
| **FIN** | Finland | Cluster 4 | Access DID | 10.3 (10.3, 10.3) | 9.7 (8.4, 11.5) | 9.5 (8.2, 11.2) | 9.9 (8.6, 11.6) | 9.7 (8.4, 11.5) |
| **FIN** | Finland | Cluster 4 | Watch DID | 3.1 (3.1, 3.1) | 0.8 (0.8, 0.9) | 1.1 (1, 1.2) | 0.7 (0.6, 0.7) | 0.8 (0.8, 0.9) |
| **FIN** | Finland | Cluster 4 | Reserve DID | 0 (0, 0) | 0 (0, 0) | 0 (0, 0) | 0 (0, 0) | 0 (0, 0) |
| **FRA** | France | Cluster 4 | Total DID | 25.1 (25.1, 25.1) | 10.7 (9.3, 12.4) | 10.7 (9.3, 12.4) | 10.7 (9.3, 12.3) | 10.7 (9.3, 12.4) |
| **FRA** | France | Cluster 4 | Access DID | 17.5 (17.5, 17.5) | 9.7 (8.2, 11.3) | 9.4 (8, 11.1) | 9.8 (8.4, 11.5) | 9.7 (8.2, 11.3) |
| **FRA** | France | Cluster 4 | Watch DID | 6.5 (6.5, 6.5) | 1 (0.9, 1.2) | 1.3 (1.2, 1.5) | 0.9 (0.8, 1) | 1 (0.9, 1.2) |
| **FRA** | France | Cluster 4 | Reserve DID | 0 (0, 0) | 0 (0, 0) | 0 (0, 0) | 0 (0, 0) | 0 (0, 0) |
| **DEU** | Germany | Cluster 4 | Total DID | 14 (14, 14) | 10.9 (9.5, 12.6) | 10.9 (9.5, 12.6) | 10.9 (9.5, 12.6) | 10.9 (9.5, 12.6) |
| **DEU** | Germany | Cluster 4 | Access DID | 8 (8, 8) | 9.9 (8.5, 11.6) | 9.6 (8.2, 11.3) | 10.1 (8.7, 11.8) | 9.9 (8.5, 11.6) |
| **DEU** | Germany | Cluster 4 | Watch DID | 5.8 (5.8, 5.8) | 0.9 (0.9, 1) | 1.3 (1.1, 1.4) | 0.7 (0.7, 0.8) | 0.9 (0.9, 1) |
| **DEU** | Germany | Cluster 4 | Reserve DID | 0.1 (0.1, 0.1) | 0 (0, 0.1) | 0 (0, 0.1) | 0 (0, 0.1) | 0 (0, 0.1) |
| **GRC** | Greece | Cluster 4 | Total DID | 37.5 (36.7, 38.8) | 10 (8.7, 11.7) | 10 (8.7, 11.7) | 10 (8.7, 11.7) | 10 (8.7, 11.7) |
| **GRC** | Greece | Cluster 4 | Access DID | 16.2 (16.1, 16.7) | 9.2 (7.9, 10.9) | 8.9 (7.6, 10.7) | 9.3 (8, 11) | 9.2 (7.9, 10.9) |
| **GRC** | Greece | Cluster 4 | Watch DID | 20.6 (20.1, 21.8) | 0.8 (0.8, 0.9) | 1.1 (1, 1.2) | 0.6 (0.6, 0.7) | 0.8 (0.8, 0.9) |
| **GRC** | Greece | Cluster 4 | Reserve DID | 0.1 (0.1, 0.1) | 0 (0, 0.1) | 0 (0, 0.1) | 0.1 (0, 0.2) | 0 (0, 0.1) |
| **HUN** | Hungary | Cluster 4 | Total DID | 15.2 (15.2, 15.2) | 7.8 (6.8, 9) | 7.8 (6.8, 9) | 7.7 (6.7, 9) | 7.8 (6.8, 9) |
| **HUN** | Hungary | Cluster 4 | Access DID | 7.6 (7.6, 7.6) | 6.8 (5.8, 8) | 6.6 (5.6, 7.8) | 6.9 (6, 8.1) | 6.8 (5.8, 8) |
| **HUN** | Hungary | Cluster 4 | Watch DID | 7.5 (7.5, 7.5) | 1 (0.8, 1.1) | 1.2 (1, 1.3) | 0.8 (0.7, 0.9) | 1 (0.8, 1.1) |
| **HUN** | Hungary | Cluster 4 | Reserve DID | 0 (0, 0) | 0 (0, 0) | 0 (0, 0) | 0 (0, 0.1) | 0 (0, 0) |
| **IND** | India | Cluster 2 | Total DID | 18.3 (16, 21) | 14.7 (11.4, 20.7) | 14.7 (11.4, 20.8) | 14.5 (11.2, 20.5) | 9.9 (7.5, 15.5) |
| **IND** | India | Cluster 2 | Access DID | 4.5 (4, 7.8) | 7.6 (3.8, 13.7) | 3.4 (-0.8, 9.5) | 10.5 (7, 16.3) | 2.8 (-0.1, 8.2) |
| **IND** | India | Cluster 2 | Watch DID | 9.3 (7.8, 10.5) | 6 (5.3, 7) | 10.3 (8.9, 11.7) | 3.9 (3.3, 4.9) | 6 (5.3, 7) |

| ISO3 Code | CTA Name | Cluster | AWaRe Category | Actual DID in IQVIA MIDAS* | Scenario 1 | Scenario 2 | Scenario 3 | Scenario 4 |
|---|---|---|---|---|---|---|---|---|
| **IND** | India | Cluster 2 | Reserve DID | 0.2 (0.2, 0.2) | 1 (0.4, 2.8) | 1 (0.4, 2.8) | 0.2 (0.1, 0.4) | 1 (0.4, 2.8) |
| **IRL** | Ireland | Cluster 4 | Total DID | 25.3 (25.3, 25.3) | 11.3 (9.9, 13.1) | 11.3 (9.9, 13.1) | 11.3 (9.8, 13.1) | 11.3 (9.9, 13.1) |
| **IRL** | Ireland | Cluster 4 | Access DID | 16.7 (16.7, 16.7) | 10.4 (8.9, 12.2) | 10.1 (8.7, 12) | 10.4 (9, 12.3) | 10.4 (8.9, 12.2) |
| **IRL** | Ireland | Cluster 4 | Watch DID | 8.5 (8.5, 8.5) | 0.9 (0.8, 1) | 1.1 (1, 1.3) | 0.8 (0.7, 0.9) | 0.9 (0.8, 1) |
| **IRL** | Ireland | Cluster 4 | Reserve DID | 0.1 (0.1, 0.1) | 0 (0, 0) | 0 (0, 0) | 0.1 (0, 0.1) | 0 (0, 0) |
| **ITA** | Italy | Cluster 4 | Total DID | 24.7 (24.7, 24.7) | 10.7 (9.3, 12.3) | 10.7 (9.3, 12.3) | 10.7 (9.3, 12.3) | 10.7 (9.3, 12.3) |
| **ITA** | Italy | Cluster 4 | Access DID | 11.5 (11.5, 11.5) | 10.1 (8.7, 11.6) | 9.9 (8.4, 11.4) | 10.2 (8.8, 11.8) | 10.1 (8.7, 11.6) |
| **ITA** | Italy | Cluster 4 | Watch DID | 13.1 (13.1, 13.1) | 0.6 (0.6, 0.7) | 0.9 (0.8, 0.9) | 0.5 (0.4, 0.5) | 0.6 (0.6, 0.7) |
| **ITA** | Italy | Cluster 4 | Reserve DID | 0.1 (0.1, 0.1) | 0 (0, 0.1) | 0 (0, 0.1) | 0.1 (0, 0.1) | 0 (0, 0.1) |
| **JPN** | Japan | Cluster 4 | Total DID | 12.8 (12.8, 12.8) | 12.8 (11.2, 14.8) | 12.8 (11.2, 14.9) | 12.8 (11.1, 14.8) | 12.8 (11.2, 14.8) |
| **JPN** | Japan | Cluster 4 | Access DID | 2.3 (2.3, 2.3) | 11.8 (10.1, 13.8) | 11.5 (9.8, 13.6) | 12 (10.3, 14.1) | 11.8 (10.1, 13.8) |
| **JPN** | Japan | Cluster 4 | Watch DID | 10.4 (10.4, 10.4) | 1 (0.9, 1.2) | 1.3 (1.2, 1.5) | 0.8 (0.7, 0.9) | 1 (0.9, 1.2) |
| **JPN** | Japan | Cluster 4 | Reserve DID | 0.1 (0.1, 0.1) | 0 (0, 0) | 0 (0, 0) | 0 (0, 0) | 0 (0, 0) |
| **JOR** | Jordan | Cluster 3 | Total DID | 11.9 (10.6, 13.7) | 12.5 (9.5, 18.1) | 12.5 (9.5, 18.1) | 12.7 (9.6, 18.3) | 8.5 (6.3, 13.1) |
| **JOR** | Jordan | Cluster 3 | Access DID | 6.3 (5.8, 8) | 11.4 (8.4, 16.9) | 10.9 (8, 16.4) | 11.7 (8.7, 17.3) | 7.4 (5.1, 11.9) |
| **JOR** | Jordan | Cluster 3 | Watch DID | 4.5 (4.1, 5.6) | 1.1 (1, 1.3) | 1.6 (1.4, 1.8) | 0.9 (0.8, 1.1) | 1.1 (1, 1.3) |
| **JOR** | Jordan | Cluster 3 | Reserve DID | 0 (0, 0) | 0 (0, 0.1) | 0 (0, 0.1) | 0.1 (0, 0.1) | 0 (0, 0.1) |
| **KAZ** | Kazakhstan | Cluster 2 | Total DID | 18.2 (18.2, 18.2) | 12.3 (9.4, 17.3) | 12.3 (9.4, 17.3) | 12.4 (9.5, 17.6) | 8.3 (6.2, 12.7) |
| **KAZ** | Kazakhstan | Cluster 2 | Access DID | 8.9 (8.9, 8.9) | 9.6 (6.7, 14.7) | 9.3 (6.3, 14.3) | 11.1 (8.2, 16.3) | 5.6 (3.2, 10.2) |
| **KAZ** | Kazakhstan | Cluster 2 | Watch DID | 8.8 (8.8, 8.8) | 1.7 (1.5, 1.9) | 2 (1.8, 2.3) | 1.2 (1.1, 1.5) | 1.7 (1.5, 1.9) |
| **KAZ** | Kazakhstan | Cluster 2 | Reserve DID | 0 (0, 0) | 1 (0.5, 1.9) | 1 (0.5, 1.9) | 0 (0, 0.1) | 1 (0.5, 1.9) |
| **KOR** | Korea, Rep. | Cluster 4 | Total DID | 25.2 (25.2, 25.2) | 12.1 (10.4, 14.3) | 12.1 (10.4, 14.3) | 12.1 (10.4, 14.3) | 12.1 (10.4, 14.3) |
| **KOR** | Korea, Rep. | Cluster 4 | Access DID | 11.4 (11.4, 11.4) | 11.4 (9.7, 13.5) | 11.2 (9.6, 13.4) | 11.5 (9.9, 13.7) | 11.4 (9.7, 13.5) |
| **KOR** | Korea, Rep. | Cluster 4 | Watch DID | 13.3 (13.3, 13.3) | 0.6 (0.6, 0.7) | 0.8 (0.7, 0.9) | 0.5 (0.4, 0.5) | 0.6 (0.6, 0.7) |
| **KOR** | Korea, Rep. | Cluster 4 | Reserve DID | 0 (0, 0) | 0.1 (0, 0.2) | 0.1 (0, 0.2) | 0.1 (0, 0.2) | 0.1 (0, 0.2) |
| **KWT** | Kuwait | Cluster 3 | Total DID | 11.2 (10.1, 12.3) | 13.4 (10.2, 19.1) | 13.4 (10.1, 19.1) | 13.5 (10.3, 19.3) | 9 (6.8, 14.1) |
| **KWT** | Kuwait | Cluster 3 | Access DID | 4.3 (4, 4.7) | 12.1 (8.9, 17.8) | 11.8 (8.5, 17.4) | 12.4 (9.1, 18.2) | 7.7 (5.4, 12.8) |
| **KWT** | Kuwait | Cluster 3 | Watch DID | 6.2 (5.4, 7.3) | 1.2 (1.1, 1.5) | 1.6 (1.4, 1.8) | 1.1 (0.9, 1.3) | 1.2 (1.1, 1.5) |
| **KWT** | Kuwait | Cluster 3 | Reserve DID | 0 (0, 0) | 0 (0, 0.1) | 0 (0, 0.1) | 0 (0, 0) | 0 (0, 0.1) |
| **LVA** | Latvia | Cluster 4 | Total DID | 12.7 (12.7, 12.7) | 10.2 (8.9, 11.7) | 10.2 (8.9, 11.7) | 10.2 (8.9, 11.7) | 10.2 (8.9, 11.7) |
| **LVA** | Latvia | Cluster 4 | Access DID | 9 (9, 9) | 8.6 (7.3, 10.2) | 8.3 (7, 9.9) | 9 (7.7, 10.6) | 8.6 (7.3, 10.2) |
| **LVA** | Latvia | Cluster 4 | Watch DID | 3.7 (3.7, 3.7) | 1.4 (1.3, 1.6) | 1.7 (1.6, 1.9) | 1.1 (1, 1.2) | 1.4 (1.3, 1.6) |
| **LVA** | Latvia | Cluster 4 | Reserve DID | 0 (0, 0) | 0.2 (0.1, 0.4) | 0.2 (0.1, 0.4) | 0.1 (0, 0.1) | 0.2 (0.1, 0.4) |
| **LBN** | Lebanon | Cluster 3 | Total DID | 28.6 (27.3, 30.4) | 13.2 (9.9, 18.5) | 13.2 (9.9, 18.5) | 13.3 (10, 18.7) | 8.9 (6.6, 13.7) |

| ISO3 Code | CTA Name | Cluster | AWaRe Category | Actual DID in IQVIA MIDAS* | Scenario 1 | Scenario 2 | Scenario 3 | Scenario 4 |
|---|---|---|---|---|---|---|---|---|
| **LBN** | Lebanon | Cluster 3 | Access DID | 16.3 (15.8, 17.7) | 11.7 (8.5, 17.1) | 11.2 (8, 16.7) | 12.1 (8.8, 17.6) | 7.4 (5.1, 12.3) |
| **LBN** | Lebanon | Cluster 3 | Watch DID | 10.3 (10, 12.1) | 1.4 (1.2, 1.7) | 1.9 (1.6, 2.2) | 1.1 (0.9, 1.5) | 1.4 (1.2, 1.7) |
| **LBN** | Lebanon | Cluster 3 | Reserve DID | 0 (0, 0) | 0 (0, 0.1) | 0 (0, 0.1) | 0.1 (0, 0.2) | 0 (0, 0.1) |
| **LTU** | Lithuania | Cluster 4 | Total DID | 16.7 (16.7, 16.7) | 10.3 (8.9, 11.7) | 10.3 (8.9, 11.7) | 10.2 (8.9, 11.7) | 10.3 (8.9, 11.7) |
| **LTU** | Lithuania | Cluster 4 | Access DID | 11.7 (11.7, 11.7) | 8.1 (6.7, 9.7) | 7.8 (6.4, 9.4) | 9 (7.6, 10.6) | 8.1 (6.7, 9.7) |
| **LTU** | Lithuania | Cluster 4 | Watch DID | 4.8 (4.8, 4.8) | 1.5 (1.4, 1.7) | 1.9 (1.7, 2) | 1.1 (1, 1.3) | 1.5 (1.4, 1.7) |
| **LTU** | Lithuania | Cluster 4 | Reserve DID | 0 (0, 0) | 0.6 (0.3, 1.1) | 0.6 (0.3, 1.1) | 0.1 (0, 0.1) | 0.6 (0.3, 1.1) |
| **LUX** | Luxembourg | Cluster 4 | Total DID | 25.2 (23.8, 26.9) | 10.9 (9.4, 12.7) | 10.9 (9.4, 12.7) | 10.9 (9.3, 12.7) | 10.9 (9.4, 12.7) |
| **LUX** | Luxembourg | Cluster 4 | Access DID | 14.5 (13.3, 15.1) | 10.1 (8.5, 11.9) | 9.9 (8.3, 11.6) | 10.3 (8.7, 12) | 10.1 (8.5, 11.9) |
| **LUX** | Luxembourg | Cluster 4 | Watch DID | 10.7 (9.6, 11.4) | 0.8 (0.7, 0.9) | 1.1 (0.9, 1.2) | 0.6 (0.6, 0.7) | 0.8 (0.7, 0.9) |
| **LUX** | Luxembourg | Cluster 4 | Reserve DID | 0.1 (0, 0.1) | 0 (0, 0) | 0 (0, 0) | 0 (0, 0) | 0 (0, 0) |
| **MEX** | Mexico | Cluster 3 | Total DID | 9.2 (7.9, 10.5) | 17.9 (13.7, 25.5) | 17.9 (13.7, 25.5) | 18.1 (13.9, 25.8) | 12 (9.1, 18.7) |
| **MEX** | Mexico | Cluster 3 | Access DID | 3.9 (3.6, 4.4) | 16.3 (12.2, 24) | 15.7 (11.6, 23.4) | 16.7 (12.5, 24.6) | 10.4 (7.5, 17.1) |
| **MEX** | Mexico | Cluster 3 | Watch DID | 4.1 (3.3, 5.6) | 1.5 (1.4, 1.7) | 2.1 (1.9, 2.3) | 1.2 (1.1, 1.4) | 1.5 (1.4, 1.7) |
| **MEX** | Mexico | Cluster 3 | Reserve DID | 0 (0, 0) | 0.1 (0, 0.1) | 0.1 (0, 0.1) | 0.1 (0, 0.2) | 0.1 (0, 0.1) |
| **MAR** | Morocco | Cluster 3 | Total DID | 16.6 (15.2, 18.4) | 16.6 (14, 22.9) | 16.6 (14, 22.9) | 16.6 (14, 22.9) | 11.4 (8.5, 17.3) |
| **MAR** | Morocco | Cluster 3 | Access DID | 11.8 (10.5, 13.2) | 14.9 (12.4, 21.2) | 14.4 (11.8, 20.7) | 15.3 (12.8, 21.7) | 9.7 (6.8, 15.7) |
| **MAR** | Morocco | Cluster 3 | Watch DID | 3.5 (3.2, 4) | 1.5 (1.4, 1.8) | 2.1 (1.9, 2.3) | 1.2 (1, 1.4) | 1.5 (1.4, 1.8) |
| **MAR** | Morocco | Cluster 3 | Reserve DID | 0 (0, 0) | 0.1 (0, 0.3) | 0.1 (0, 0.3) | 0 (0, 0.1) | 0.1 (0, 0.3) |
| **NLD** | Netherlands | Cluster 4 | Total DID | 9.7 (9.7, 9.7) | 10.7 (9.2, 12.4) | 10.7 (9.2, 12.4) | 10.6 (9.2, 12.4) | 10.7 (9.2, 12.4) |
| **NLD** | Netherlands | Cluster 4 | Access DID | 6.9 (6.9, 6.9) | 9.6 (8.1, 11.3) | 9.3 (7.8, 11.1) | 9.8 (8.3, 11.5) | 9.6 (8.1, 11.3) |
| **NLD** | Netherlands | Cluster 4 | Watch DID | 2.8 (2.8, 2.8) | 1.1 (0.9, 1.2) | 1.4 (1.2, 1.5) | 0.8 (0.8, 1) | 1.1 (0.9, 1.2) |
| **NLD** | Netherlands | Cluster 4 | Reserve DID | 0 (0, 0) | 0 (0, 0) | 0 (0, 0) | 0 (0, 0) | 0 (0, 0) |
| **NZL** | New Zealand | Cluster 4 | Total DID | 24.1 (24.1, 24.1) | 14.3 (12.5, 16.4) | 14.3 (12.5, 16.4) | 14.3 (12.5, 16.4) | 14.3 (12.5, 16.4) |
| **NZL** | New Zealand | Cluster 4 | Access DID | 19.1 (19.1, 19.1) | 13.3 (11.5, 15.4) | 13 (11.2, 15.1) | 13.4 (11.6, 15.5) | 13.3 (11.5, 15.4) |
| **NZL** | New Zealand | Cluster 4 | Watch DID | 4.9 (4.9, 4.9) | 1 (0.9, 1.1) | 1.3 (1.2, 1.4) | 0.8 (0.8, 0.9) | 1 (0.9, 1.1) |
| **NZL** | New Zealand | Cluster 4 | Reserve DID | 0 (0, 0) | 0 (0, 0) | 0 (0, 0) | 0 (0, 0.1) | 0 (0, 0) |
| **NOR** | Norway | Cluster 4 | Total DID | 13.5 (13.5, 13.5) | 11.7 (10.2, 13.5) | 11.7 (10.2, 13.5) | 11.7 (10.2, 13.5) | 11.7 (10.2, 13.5) |
| **NOR** | Norway | Cluster 4 | Access DID | 10.9 (10.9, 10.9) | 10.6 (9, 12.4) | 10.3 (8.7, 12.1) | 10.8 (9.2, 12.6) | 10.6 (9, 12.4) |
| **NOR** | Norway | Cluster 4 | Watch DID | 2.6 (2.6, 2.6) | 1.1 (1, 1.2) | 1.4 (1.3, 1.6) | 0.9 (0.8, 1) | 1.1 (1, 1.2) |
| **NOR** | Norway | Cluster 4 | Reserve DID | 0 (0, 0) | 0 (0, 0.1) | 0 (0, 0.1) | 0 (0, 0.1) | 0 (0, 0.1) |
| **PAK** | Pakistan | Cluster 2 | Total DID | 24.6 (22, 27.4) | 19.9 (15.2, 28) | 19.9 (15.2, 28) | 19.8 (15.1, 28) | 13.5 (10.1, 20.6) |
| **PAK** | Pakistan | Cluster 2 | Access DID | 10.9 (10.4, 14.3) | 13.2 (8.5, 21.6) | 9.6 (4.7, 18) | 15.6 (10.9, 24) | 6.8 (3.2, 14.4) |
| **PAK** | Pakistan | Cluster 2 | Watch DID | 10.8 (10.1, 12.6) | 5.9 (5.2, 7) | 9.5 (8.4, 11.1) | 4 (3.4, 5) | 5.9 (5.2, 7) |

| ISO3 Code | CTA Name | Cluster | AWaRe Category | Actual DID in IQVIA MIDAS* | Scenario 1 | Scenario 2 | Scenario 3 | Scenario 4 |
|---|---|---|---|---|---|---|---|---|
| **PAK** | Pakistan | Cluster 2 | Reserve DID | 0.1 (0.1, 0.2) | 0.6 (0.3, 1.7) | 0.6 (0.3, 1.7) | 0.1 (0, 0.3) | 0.6 (0.3, 1.7) |
| **PER** | Peru | Cluster 2 | Total DID | 14.5 (13.2, 16) | 20.5 (16, 28.7) | 20.5 (16, 28.7) | 20.6 (16.1, 28.9) | 13.9 (10.3, 21.7) |
| **PER** | Peru | Cluster 2 | Access DID | 6.5 (6.1, 7.9) | 17.7 (13.3, 25.9) | 17 (12.5, 25.2) | 18.7 (14.2, 27) | 11.1 (7.5, 19) |
| **PER** | Peru | Cluster 2 | Watch DID | 5.2 (4.7, 6.8) | 2.3 (2.1, 2.6) | 2.9 (2.7, 3.3) | 1.8 (1.6, 2) | 2.3 (2.1, 2.6) |
| **PER** | Peru | Cluster 2 | Reserve DID | 0.1 (0, 0.1) | 0.4 (0.2, 0.9) | 0.4 (0.2, 0.9) | 0.1 (0, 0.3) | 0.4 (0.2, 0.9) |
| **POL** | Poland | Cluster 4 | Total DID | 22.8 (22.8, 22.8) | 8.4 (7.3, 9.7) | 8.4 (7.3, 9.7) | 8.4 (7.3, 9.7) | 8.4 (7.3, 9.7) |
| **POL** | Poland | Cluster 4 | Access DID | 12.5 (12.5, 12.5) | 7.4 (6.3, 8.7) | 7.2 (6.1, 8.5) | 7.6 (6.4, 8.8) | 7.4 (6.3, 8.7) |
| **POL** | Poland | Cluster 4 | Watch DID | 8.9 (8.9, 8.9) | 1 (0.9, 1.1) | 1.2 (1, 1.3) | 0.8 (0.7, 0.9) | 1 (0.9, 1.1) |
| **POL** | Poland | Cluster 4 | Reserve DID | 0 (0, 0) | 0 (0, 0) | 0 (0, 0) | 0 (0, 0.1) | 0 (0, 0) |
| **PRT** | Portugal | Cluster 4 | Total DID | 20.8 (20.8, 20.8) | 10.6 (9.3, 12.4) | 10.6 (9.3, 12.4) | 10.6 (9.3, 12.3) | 10.6 (9.3, 12.4) |
| **PRT** | Portugal | Cluster 4 | Access DID | 12.9 (12.9, 12.9) | 9.7 (8.3, 11.4) | 9.4 (8, 11.1) | 9.9 (8.5, 11.6) | 9.7 (8.3, 11.4) |
| **PRT** | Portugal | Cluster 4 | Watch DID | 7.7 (7.7, 7.7) | 0.9 (0.8, 1) | 1.2 (1.1, 1.4) | 0.6 (0.6, 0.7) | 0.9 (0.8, 1) |
| **PRT** | Portugal | Cluster 4 | Reserve DID | 0 (0, 0) | 0 (0, 0.1) | 0 (0, 0.1) | 0.1 (0, 0.1) | 0 (0, 0.1) |
| **PRI** | Puerto Rico | Cluster 3 | Total DID | 19.1 (19.1, 19.1) | 15.6 (12.3, 22) | 15.6 (12.4, 22) | 15.7 (12.4, 22.1) | 10.6 (8.1, 16.3) |
| **PRI** | Puerto Rico | Cluster 3 | Access DID | 9.6 (9.6, 9.6) | 13.8 (10.5, 20.2) | 13.3 (9.9, 19.7) | 14.2 (10.9, 20.6) | 8.8 (6.3, 14.5) |
| **PRI** | Puerto Rico | Cluster 3 | Watch DID | 9.4 (9.4, 9.4) | 1.8 (1.6, 2) | 2.3 (2.1, 2.6) | 1.4 (1.2, 1.6) | 1.8 (1.6, 2) |
| **PRI** | Puerto Rico | Cluster 3 | Reserve DID | 0.1 (0.1, 0.1) | 0 (0, 0.1) | 0 (0, 0.1) | 0.1 (0, 0.2) | 0 (0, 0.1) |
| **ROU** | Romania | Cluster 4 | Total DID | 27.3 (27.3, 27.3) | 7.8 (6.8, 9.1) | 7.8 (6.8, 9.1) | 7.8 (6.8, 9) | 7.8 (6.8, 9.1) |
| **ROU** | Romania | Cluster 4 | Access DID | 14.2 (14.2, 14.2) | 6.7 (5.6, 7.8) | 6.4 (5.3, 7.6) | 7 (5.9, 8.1) | 6.7 (5.6, 7.8) |
| **ROU** | Romania | Cluster 4 | Watch DID | 12.6 (12.6, 12.6) | 1.1 (0.9, 1.2) | 1.3 (1.2, 1.4) | 0.8 (0.7, 0.9) | 1.1 (0.9, 1.2) |
| **ROU** | Romania | Cluster 4 | Reserve DID | 0 (0, 0) | 0.1 (0.1, 0.3) | 0.1 (0.1, 0.3) | 0.1 (0, 0.2) | 0.1 (0.1, 0.3) |
| **RUS** | Russian Federation | Cluster 4 | Total DID | 17.1 (17.1, 17.1) | 11.2 (9.7, 13) | 11.2 (9.7, 13) | 11.2 (9.7, 13) | 11.2 (9.7, 13) |
| **RUS** | Russian Federation | Cluster 4 | Access DID | 8 (8, 8) | 7.8 (5.8, 9.8) | 7.5 (5.4, 9.4) | 9.9 (8.5, 11.7) | 7.8 (5.8, 9.8) |
| **RUS** | Russian Federation | Cluster 4 | Watch DID | 7.6 (7.6, 7.6) | 1.8 (1.6, 2) | 2.2 (2, 2.4) | 1.2 (1.1, 1.4) | 1.8 (1.6, 2) |
| **RUS** | Russian Federation | Cluster 4 | Reserve DID | 0 (0, 0) | 1.5 (0.7, 2.9) | 1.5 (0.7, 2.9) | 0 (0, 0.1) | 1.5 (0.7, 2.9) |
| **SAU** | Saudi Arabia | Cluster 3 | Total DID | 20.8 (20.8, 20.8) | 14.1 (10.6, 19.8) | 14.1 (10.6, 19.8) | 14.2 (10.7, 20) | 9.5 (7.2, 14.7) |
| **SAU** | Saudi Arabia | Cluster 3 | Access DID | 10.7 (10.7, 10.7) | 12.4 (8.9, 18) | 12 (8.5, 17.6) | 12.8 (9.2, 18.5) | 7.9 (5.5, 13.1) |
| **SAU** | Saudi Arabia | Cluster 3 | Watch DID | 10.1 (10.1, 10.1) | 1.5 (1.3, 1.9) | 2 (1.7, 2.3) | 1.3 (1.1, 1.7) | 1.5 (1.3, 1.9) |
| **SAU** | Saudi Arabia | Cluster 3 | Reserve DID | 0 (0, 0) | 0.1 (0, 0.3) | 0.1 (0, 0.3) | 0.1 (0, 0.1) | 0.1 (0, 0.3) |
| **SRB** | Serbia | Cluster 4 | Total DID | 29.3 (28.9, 30) | 10.2 (8.8, 11.8) | 10.2 (8.8, 11.8) | 10.2 (8.8, 11.8) | 10.2 (8.8, 11.8) |
| **SRB** | Serbia | Cluster 4 | Access DID | 16.2 (16, 16.7) | 9 (7.6, 10.6) | 8.7 (7.3, 10.3) | 9.1 (7.8, 10.8) | 9 (7.6, 10.6) |
| **SRB** | Serbia | Cluster 4 | Watch DID | 12.2 (12.1, 12.7) | 1.2 (1.1, 1.4) | 1.5 (1.3, 1.7) | 1 (0.8, 1.2) | 1.2 (1.1, 1.4) |
| **SRB** | Serbia | Cluster 4 | Reserve DID | 0 (0, 0) | 0 (0, 0.1) | 0 (0, 0.1) | 0 (0, 0.1) | 0 (0, 0.1) |

| ISO3 Code | CTA Name | Cluster | AWaRe Category | Actual DID in IQVIA MIDAS* | Scenario 1 | Scenario 2 | Scenario 3 | Scenario 4 |
|---|---|---|---|---|---|---|---|---|
| **SGP** | Singapore | Cluster 4 | Total DID | 11 (11, 11) | 12.6 (10.8, 14.6) | 12.6 (10.8, 14.6) | 12.6 (10.8, 14.6) | 12.6 (10.8, 14.6) |
| **SGP** | Singapore | Cluster 4 | Access DID | 6.7 (6.7, 6.7) | 11.8 (10, 13.9) | 11.6 (9.8, 13.7) | 12 (10.2, 14.1) | 11.8 (10, 13.9) |
| **SGP** | Singapore | Cluster 4 | Watch DID | 4.2 (4.2, 4.2) | 0.7 (0.6, 0.8) | 0.9 (0.8, 1.1) | 0.5 (0.5, 0.6) | 0.7 (0.6, 0.8) |
| **SGP** | Singapore | Cluster 4 | Reserve DID | 0 (0, 0) | 0 (0, 0.1) | 0 (0, 0.1) | 0 (0, 0.1) | 0 (0, 0.1) |
| **SVK** | Slovak Republic | Cluster 4 | Total DID | 23 (23, 23) | 8 (6.9, 9.3) | 8 (6.9, 9.3) | 7.9 (6.8, 9.2) | 8 (6.9, 9.3) |
| **SVK** | Slovak Republic | Cluster 4 | Access DID | 8.2 (8.2, 8.2) | 6.9 (5.8, 8.2) | 6.7 (5.6, 8) | 7.1 (5.9, 8.3) | 6.9 (5.8, 8.2) |
| **SVK** | Slovak Republic | Cluster 4 | Watch DID | 12.2 (12.2, 12.2) | 1 (0.9, 1.2) | 1.2 (1.1, 1.4) | 0.8 (0.7, 0.9) | 1 (0.9, 1.2) |
| **SVK** | Slovak Republic | Cluster 4 | Reserve DID | 0 (0, 0) | 0 (0, 0) | 0 (0, 0) | 0 (0, 0.1) | 0 (0, 0) |
| **SVN** | Slovenia | Cluster 4 | Total DID | 15.7 (15.2, 16.3) | 7.7 (6.7, 8.9) | 7.7 (6.7, 8.9) | 7.7 (6.7, 8.8) | 7.7 (6.7, 8.9) |
| **SVN** | Slovenia | Cluster 4 | Access DID | 11.2 (10.9, 11.4) | 6.8 (5.8, 7.9) | 6.6 (5.6, 7.7) | 6.9 (5.9, 8.1) | 6.8 (5.8, 7.9) |
| **SVN** | Slovenia | Cluster 4 | Watch DID | 4.5 (4.2, 4.9) | 0.9 (0.8, 1) | 1.1 (1, 1.2) | 0.7 (0.6, 0.8) | 0.9 (0.8, 1) |
| **SVN** | Slovenia | Cluster 4 | Reserve DID | 0 (0, 0) | 0 (0, 0) | 0 (0, 0) | 0 (0, 0) | 0 (0, 0) |
| **ZAF** | South Africa | Cluster 2 | Total DID | 15.6 (15.6, 15.6) | 23.3 (18.1, 32.9) | 23.4 (18.1, 32.9) | 23.3 (18.1, 32.9) | 15.8 (12, 24.2) |
| **ZAF** | South Africa | Cluster 2 | Access DID | 11.8 (11.8, 11.8) | 17.6 (12.2, 27.5) | 16.8 (11.4, 26.7) | 19.4 (14.1, 28.9) | 10.1 (5.9, 18.7) |
| **ZAF** | South Africa | Cluster 2 | Watch DID | 3.6 (3.6, 3.6) | 4.8 (4.1, 5.8) | 5.6 (4.9, 6.6) | 3.8 (3.2, 4.9) | 4.8 (4.1, 5.8) |
| **ZAF** | South Africa | Cluster 2 | Reserve DID | 0 (0, 0) | 0.8 (0.3, 2.2) | 0.8 (0.3, 2.2) | 0.1 (0, 0.2) | 0.8 (0.3, 2.2) |
| **ESP** | Spain | Cluster 4 | Total DID | 26.9 (26.9, 26.9) | 10.5 (8.9, 12) | 10.5 (8.9, 12) | 10.4 (8.9, 12) | 10.5 (8.9, 12) |
| **ESP** | Spain | Cluster 4 | Access DID | 16.9 (16.9, 16.9) | 9.5 (8, 11.1) | 9.3 (7.7, 10.8) | 9.7 (8.2, 11.3) | 9.5 (8, 11.1) |
| **ESP** | Spain | Cluster 4 | Watch DID | 9.6 (9.6, 9.6) | 0.9 (0.8, 1) | 1.2 (1.1, 1.3) | 0.7 (0.6, 0.7) | 0.9 (0.8, 1) |
| **ESP** | Spain | Cluster 4 | Reserve DID | 0.1 (0.1, 0.1) | 0 (0, 0) | 0 (0, 0) | 0 (0, 0) | 0 (0, 0) |
| **LKA** | Sri Lanka | Cluster 2 | Total DID | 11.2 (10.1, 12.1) | 15.9 (12.2, 22.7) | 15.9 (12.2, 22.7) | 16 (12.2, 22.8) | 10.8 (8, 16.6) |
| **LKA** | Sri Lanka | Cluster 2 | Access DID | 5.1 (4.9, 5.6) | 13.8 (10.2, 20.6) | 12.6 (8.9, 19.5) | 14.5 (10.8, 21.3) | 8.7 (5.9, 14.6) |
| **LKA** | Sri Lanka | Cluster 2 | Watch DID | 5.9 (4.8, 6.7) | 2.1 (1.8, 2.4) | 3.3 (2.9, 3.8) | 1.4 (1.2, 1.7) | 2.1 (1.8, 2.4) |
| **LKA** | Sri Lanka | Cluster 2 | Reserve DID | 0.1 (0, 0.1) | 0 (0, 0.1) | 0 (0, 0.1) | 0 (0, 0.1) | 0 (0, 0.1) |
| **SWE** | Sweden | Cluster 4 | Total DID | 13.6 (13.1, 14.5) | 11.7 (10.1, 13.5) | 11.7 (10.1, 13.5) | 11.7 (10.1, 13.5) | 11.7 (10.1, 13.5) |
| **SWE** | Sweden | Cluster 4 | Access DID | 9.6 (9.3, 9.8) | 10.3 (8.8, 12.1) | 10 (8.5, 11.8) | 10.5 (8.9, 12.3) | 10.3 (8.8, 12.1) |
| **SWE** | Sweden | Cluster 4 | Watch DID | 3.9 (3.6, 4.5) | 1.4 (1.2, 1.5) | 1.6 (1.5, 1.8) | 1.2 (1, 1.3) | 1.4 (1.2, 1.5) |
| **SWE** | Sweden | Cluster 4 | Reserve DID | 0 (0, 0.1) | 0 (0, 0) | 0 (0, 0) | 0 (0, 0) | 0 (0, 0) |
| **CHE** | Switzerland | Cluster 4 | Total DID | 11 (11, 11) | 11 (11, 11) | 11 (11, 11) | 11 (11, 11) | 11 (11, 11) |
| **CHE** | Switzerland | Cluster 4 | Access DID | 6.7 (6.7, 6.7) | 10 (9.8, 10.1) | 9.7 (9.6, 9.9) | 10.1 (10, 10.2) | 10 (9.8, 10.1) |
| **CHE** | Switzerland | Cluster 4 | Watch DID | 4.2 (4.2, 4.2) | 1 (0.9, 1.1) | 1.3 (1.1, 1.4) | 0.9 (0.8, 1) | 1 (0.9, 1.1) |
| **CHE** | Switzerland | Cluster 4 | Reserve DID | 0 (0, 0) | 0 (0, 0) | 0 (0, 0) | 0 (0, 0) | 0 (0, 0) |
| **TWN** | Taiwan | Cluster 3 | Total DID | 15.7 (15.7, 15.7) | 11.1 (8.6, 15.7) | 11.1 (8.6, 15.7) | 11 (8.5, 15.6) | 7.4 (5.7, 11.4) |
| **TWN** | Taiwan | Cluster 3 | Access DID | 10.6 (10.6, 10.6) | 9.2 (6.7, 13.9) | 8.7 (6.2, 13.4) | 9.6 (7.1, 14.2) | 5.6 (3.8, 9.6) |

| ISO3 Code | CTA Name | Cluster | AWaRe Category | Actual DID in IQVIA MIDAS* | Scenario 1 | Scenario 2 | Scenario 3 | Scenario 4 |
|---|---|---|---|---|---|---|---|---|
| **TWN** | Taiwan | Cluster 3 | Watch DID | 4.8 (4.8, 4.8) | 1.8 (1.6, 2) | 2.2 (2, 2.5) | 1.3 (1.1, 1.5) | 1.8 (1.6, 2) |
| **TWN** | Taiwan | Cluster 3 | Reserve DID | 0.1 (0.1, 0.1) | 0.1 (0, 0.3) | 0.1 (0, 0.3) | 0.1 (0, 0.1) | 0.1 (0, 0.3) |
| **THA** | Thailand | Cluster 2 | Total DID | 23.6 (23.6, 23.6) | 20.1 (15.6, 28.2) | 20.1 (15.6, 28.2) | 20.1 (15.6, 28.3) | 13.6 (10.2, 20.6) |
| **THA** | Thailand | Cluster 2 | Access DID | 17 (17, 17) | 17.5 (12.9, 25.5) | 16.3 (11.7, 24.5) | 18.5 (13.8, 26.6) | 11 (7.6, 18) |
| **THA** | Thailand | Cluster 2 | Watch DID | 6.5 (6.5, 6.5) | 2.3 (2.1, 2.6) | 3.5 (3.1, 3.8) | 1.6 (1.4, 1.8) | 2.3 (2.1, 2.6) |
| **THA** | Thailand | Cluster 2 | Reserve DID | 0 (0, 0) | 0.2 (0.1, 0.7) | 0.2 (0.1, 0.7) | 0.1 (0, 0.4) | 0.2 (0.1, 0.7) |
| **TUN** | Tunisia | Cluster 3 | Total DID | 33.8 (33.8, 33.8) | 13.4 (10.3, 18.9) | 13.4 (10.3, 18.9) | 13.5 (10.3, 19) | 9 (6.8, 13.9) |
| **TUN** | Tunisia | Cluster 3 | Access DID | 23.4 (23.4, 23.4) | 12 (8.9, 17.5) | 11.6 (8.4, 17) | 12.3 (9.1, 17.8) | 7.5 (5.4, 12.4) |
| **TUN** | Tunisia | Cluster 3 | Watch DID | 8.3 (8.3, 8.3) | 1.4 (1.2, 1.6) | 1.8 (1.6, 2.1) | 1.2 (1, 1.4) | 1.4 (1.2, 1.6) |
| **TUN** | Tunisia | Cluster 3 | Reserve DID | 0 (0, 0) | 0 (0, 0.1) | 0 (0, 0.1) | 0 (0, 0.1) | 0 (0, 0.1) |
| **TUR** | Turkiye | Cluster 3 | Total DID | 34.2 (34.2, 34.2) | 18.4 (13.9, 25.8) | 18.4 (13.9, 25.8) | 18.6 (14, 26.2) | 12.4 (9.4, 19.2) |
| **TUR** | Turkiye | Cluster 3 | Access DID | 17.9 (17.9, 17.9) | 17 (12.5, 24.4) | 16.6 (12, 24) | 17.4 (12.8, 25) | 11 (8, 17.8) |
| **TUR** | Turkiye | Cluster 3 | Watch DID | 15.4 (15.4, 15.4) | 1.3 (1.2, 1.5) | 1.8 (1.6, 2) | 1.1 (1, 1.3) | 1.3 (1.2, 1.5) |
| **TUR** | Turkiye | Cluster 3 | Reserve DID | 0.1 (0.1, 0.1) | 0 (0, 0.1) | 0 (0, 0.1) | 0 (0, 0.1) | 0 (0, 0.1) |
| **ARE** | United Arab Emirates | Cluster 3 | Total DID | 27.7 (26.7, 28.6) | 13.5 (10.2, 19.1) | 13.5 (10.2, 19.1) | 13.7 (10.3, 19.4) | 9.2 (6.8, 14.6) |
| **ARE** | United Arab Emirates | Cluster 3 | Access DID | 12.1 (11.8, 12.7) | 12.4 (9.1, 18) | 12 (8.8, 17.7) | 12.6 (9.3, 18.4) | 8 (5.6, 13.4) |
| **ARE** | United Arab Emirates | Cluster 3 | Watch DID | 15.2 (14.2, 15.9) | 1.1 (1, 1.4) | 1.4 (1.3, 1.7) | 1 (0.8, 1.2) | 1.1 (1, 1.4) |
| **ARE** | United Arab Emirates | Cluster 3 | Reserve DID | 0 (0, 0) | 0 (0, 0.1) | 0 (0, 0.1) | 0.1 (0, 0.2) | 0 (0, 0.1) |
| **GBR** | United Kingdom | Cluster 4 | Total DID | 19.7 (19.7, 19.7) | 11.6 (10.1, 13.4) | 11.6 (10.1, 13.4) | 11.5 (10.1, 13.4) | 11.6 (10.1, 13.4) |
| **GBR** | United Kingdom | Cluster 4 | Access DID | 13.2 (13.2, 13.2) | 10.6 (9.1, 12.5) | 10.3 (8.9, 12.2) | 10.8 (9.3, 12.6) | 10.6 (9.1, 12.5) |
| **GBR** | United Kingdom | Cluster 4 | Watch DID | 6.5 (6.5, 6.5) | 0.9 (0.8, 1.1) | 1.2 (1.1, 1.4) | 0.7 (0.6, 0.7) | 0.9 (0.8, 1.1) |
| **GBR** | United Kingdom | Cluster 4 | Reserve DID | 0.1 (0.1, 0.1) | 0 (0, 0) | 0 (0, 0) | 0.1 (0, 0.2) | 0 (0, 0) |
| **USA** | United States | Cluster 4 | Total DID | 23.7 (23.7, 23.7) | 16.2 (14.1, 18.7) | 16.2 (14.1, 18.7) | 16.2 (14.1, 18.7) | 16.2 (14.1, 18.7) |
| **USA** | United States | Cluster 4 | Access DID | 16.7 (16.7, 16.7) | 15.2 (13.2, 17.8) | 14.9 (12.9, 17.5) | 15.3 (13.2, 17.9) | 15.2 (13.2, 17.8) |
| **USA** | United States | Cluster 4 | Watch DID | 6.9 (6.9, 6.9) | 0.9 (0.8, 0.9) | 1.2 (1.1, 1.3) | 0.6 (0.6, 0.7) | 0.9 (0.8, 0.9) |
| **USA** | United States | Cluster 4 | Reserve DID | 0.1 (0.1, 0.1) | 0 (0, 0.1) | 0 (0, 0.1) | 0.2 (0.1, 0.4) | 0 (0, 0.1) |
| **URY** | Uruguay | Cluster 3 | Total DID | 16.1 (15.7, 16.7) | 18.7 (14.4, 26.6) | 18.7 (14.4, 26.6) | 18.9 (14.6, 26.9) | 12.6 (9.5, 19.5) |
| **URY** | Uruguay | Cluster 3 | Access DID | 9.5 (9.3, 9.9) | 17.6 (13.3, 25.5) | 17.2 (13, 25.1) | 18 (13.7, 26) | 11.5 (8.3, 18.4) |
| **URY** | Uruguay | Cluster 3 | Watch DID | 6.2 (5.9, 6.3) | 1.1 (1, 1.2) | 1.5 (1.3, 1.6) | 0.7 (0.7, 0.8) | 1.1 (1, 1.2) |
| **URY** | Uruguay | Cluster 3 | Reserve DID | 0 (0, 0.1) | 0 (0, 0.1) | 0 (0, 0.1) | 0.1 (0.1, 0.3) | 0 (0, 0.1) |
| **VNM** | Viet Nam | Cluster 2 | Total DID | 26.4 (26.4, 26.4) | 14.2 (10.8, 19.9) | 14.2 (10.8, 19.9) | 14.2 (10.7, 19.9) | 9.6 (7.3, 14.7) |
| **VNM** | Viet Nam | Cluster 2 | Access DID | 13.5 (13.5, 13.5) | 11.7 (8.3, 17.5) | 10.5 (7.1, 16.4) | 12.8 (9.4, 18.6) | 7 (4.6, 12.2) |
| **VNM** | Viet Nam | Cluster 2 | Watch DID | 12.3 (12.3, 12.3) | 2.1 (1.8, 2.4) | 3.2 (2.8, 3.7) | 1.4 (1.2, 1.7) | 2.1 (1.8, 2.4) |

| ISO3 Code | CTA Name | Cluster | AWaRe Category | Actual DID in IQVIA MIDAS* | Scenario 1 | Scenario 2 | Scenario 3 | Scenario 4 |
|---|---|---|---|---|---|---|---|---|
| **VNM** | Viet Nam | Cluster 2 | Reserve DID | 0 (0, 0) | 0.4 (0.2, 1.2) | 0.4 (0.2, 1.2) | 0 (0, 0.1) | 0.4 (0.2, 1.2) |

** Source: Actual DID based on IQVIA MIDAS® data for 2019, reflecting estimates of real-world activity. Copyright IQVIA. All Rights Reserved*
